# Estimates of underlying health biases in SARS-CoV-2 vaccination recipients: a nationwide study in previously-infected adults

**DOI:** 10.1101/2025.02.19.25322515

**Authors:** Uwe Riedmann, Alena Chalupka, Lukas Richter, Dirk Werber, Martin Sprenger, Peter Willeit, Marc Rijksen, Julia Lodron, Tracy Beth Høeg, John PA Ioannidis, Stefan Pilz

## Abstract

**Background:** Observational studies may over- or under-estimate SARS-CoV-2 vaccine effectiveness (VE) depending on whether healthier (i.e. healthy vaccine effect (HVE)) or more ill individuals are preferentially vaccinated. To evaluate this issue, we compared non-COVID-19, all-cause, cancer and COVID-19 mortality in vaccinated versus unvaccinated individuals.

**Methods:** This is a nationwide retrospective observational study in the entire adult population in Austria with previously documented SARS-CoV-2 infection with a follow-up from 2021 to 2023. Cox regression analyses were used to calculate hazard ratios (HRs) according to the number of SARS-CoV-2 vaccinations. We also performed matched analyses, where on each day, newly vaccinated individuals were matched with unvaccinated individuals based on age, sex and nursing home residency.

**Results:** Overall, 4,324,485 individuals (median age (IQR): 46 (33-59) years; 52.56% female) were eligible and 2.23 non-COVID-19 deaths occurred per 100,000 person days. Group differences in non-COVID-19 mortality risk were most prominent in the early periods (e.g., in Q4 2021, adjusted HRs (95% CI) in vaccinated versus unvaccinated were 0.69 (0.59 - 0.81), 0.65 (0.58 - 0.74), and 0.56 (0.48 - 0.66) for 1-, 2-, and 3-vaccinations, respectively) and decreased thereafter. Matched analyses for the first two weeks after vaccination showed HRs below 0.5 for vaccinated versus unvaccinated individuals irrespective of vaccination numbers. Similar findings were retrieved for non-COVID-19, all-cause, and cancer deaths. Overall, COVID-19 deaths were significantly reduced in vaccinated individuals.

**Conclusions:** HVE for SARS-CoV-2 vaccines was strong early after vaccination and diminished over time. HVE should be considered when estimating VE.

## INTRODUCTION

Randomized controlled trials (RCTs) documented a high efficacy of SARS-CoV-2 vaccines against symptomatic and critical or severe COVID-19 in individuals without a previous SARS-CoV-2 infection or vaccination.^1^ Evidence on booster doses and the vaccine effectiveness (VE) against COVID-19 mortality is almost exclusively based on observational studies that are prone to different sources of bias and confounding that may cause over- or under-estimation of VE.^2–6^

The “healthy vaccinee effect” (HVE), also termed healthy vaccinee bias, occurs when individuals with generally better health outcomes such as lower all-cause mortality are more likely to receive a vaccination versus those who do not, thus leading to an overestimation of VE.^7–12^ Causes of this HVE are challenging to capture by administrative data on co-morbidities, as they are probably related to unmeasured factors such as a healthier lifestyle, timing of vaccinations during good health conditions, or fewer functional status limitations.^7,10,13^ The HVE is documented for different vaccines, but only insufficiently explored for SARS-CoV-2 vaccines, as no previous study on this issue included all previously infected individuals of a nation or covered the whole COVID-19 pandemic.^4,7,14–18^ Conversely, sometimes individuals with a poorer health status may be more likely to be vaccinated; such “confounding by indication”^7^ creates a healthy non-vaccinee bias. Terminal illness may also decrease the likelihood of vaccination. All these biases may be present simultaneously with opposing effects on VE and can thus be challenging to disentangle.^7^

Assessing mortality unrelated to the target disease in vaccinated versus unvaccinated individuals during times of low infection rates has been proposed to evaluate the HVE.^17^ This estimates the net effect of the HVE, confounding by indication and other related biases. Part of these biases may be explained by differences in age and long-term care residence status (a proxy for comorbidities) between vaccinated and unvaccinated individuals. Stratifying for these factors would thus provide insight into whether there are biases independent of age and long-term care residency.^4,7,9^

In this nationwide cohort study in previously infected adults from Austria, we compare non-COVID-19 and all-cause mortality (primary outcome measures) in vaccinated versus unvaccinated individuals during 2021-2023. Additional analyses focus on the months with low and high COVID-19 mortality rates. Furthermore, we match vaccinated with unvaccinated individuals to assess differences in mortality for the first two weeks after vaccination, i.e. before reasonable onset of vaccination effects. Cancer and transport accident deaths are evaluated as control outcomes that appear unrelated to COVID-19 and vaccines. Finally, we evaluate VE regarding COVID-19 mortality to put into perspective potential effect sizes of the HVE and VE.

## METHODS

### Study design, procedures, and participants

We conducted a nationwide retrospective observational study in previously SARS-CoV-2 infected adults (aged ≥ 18 years) from Austria from the start of mass vaccinations in January 2021 till the end of 2023 (Figure 1). We used national health data from the Austrian epidemiological reporting system (German: *Epidemiologisches Meldesystem*; EMS) provided by the Austrian Agency for Health and Food Safety (German: *Österreichische Agentur für Gesundheit und Ernährungssicherheit*; AGES), which includes data on age, gender, SARS-CoV-2 infections (date of infection) vaccinations against SARS-CoV-2 (date of vaccination and vaccine product), administrative district and vital status regarding COVID-19 deaths.^19–21^ Detection of SARS-CoV-2 infections was based on polymerase chain reaction (PCR) tests or, restricted to the end of 2020 until May 2021, antigen tests from accredited diagnostic facilities. Presence of repeated previous documented infection was classified if there were >=2 positive test results more than 90 days apart. Postal address was used to classify nursing home residency. All-cause and cause-specific mortality data (date and cause of death) were derived from Statistics Austria.^19^

**Figure 1:**
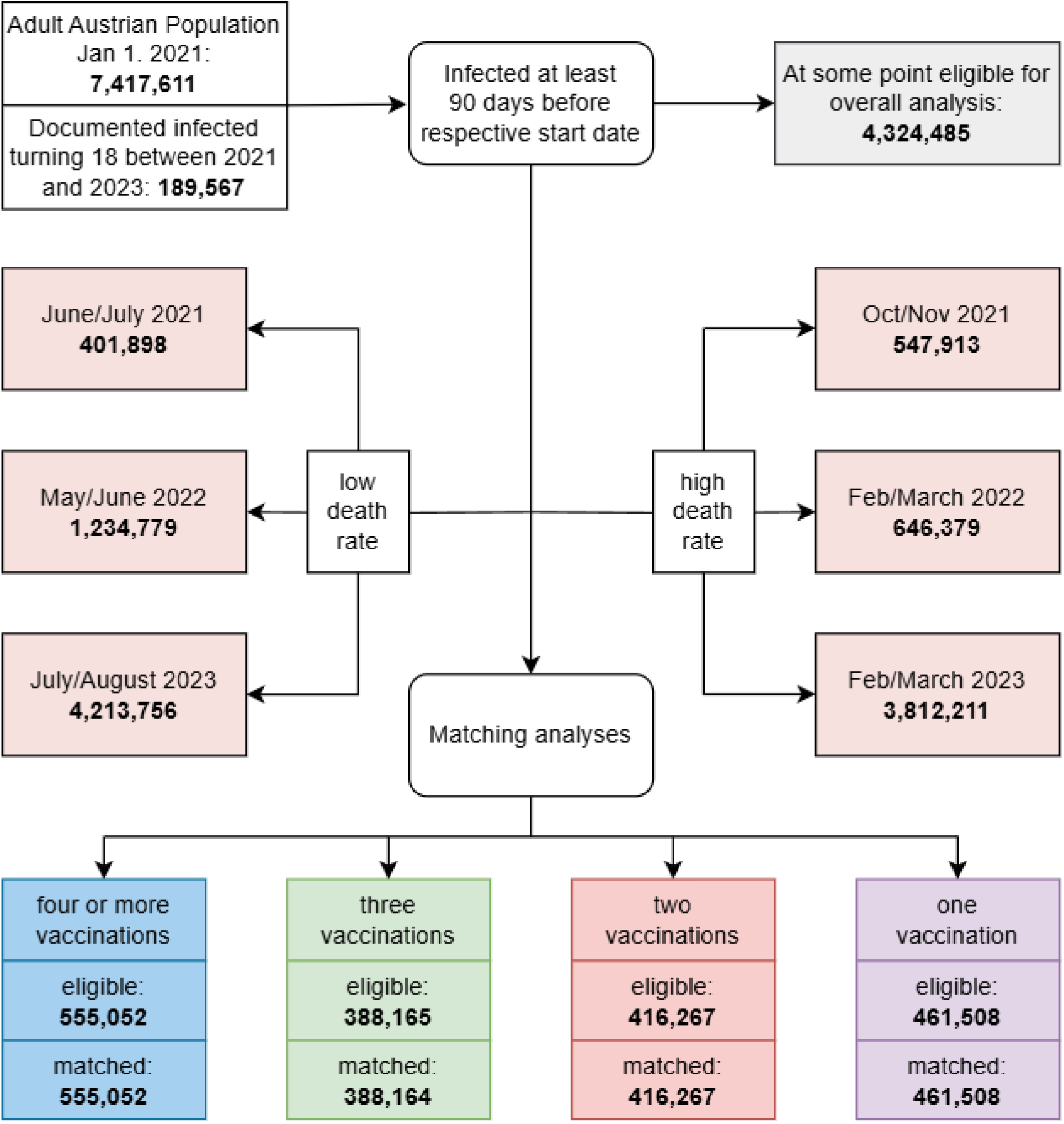
Participant selection chart. Three vaccinations had one eligible case that was unmatched, all other vaccinations were successfully matched with controls.

We followed the Strengthening the Reporting of Observational Studies in Epidemiology (STROBE) checklist (Table S1). No sample size calculation was performed prior to initiation of the study but outcome event numbers were considered sufficient for analysis (Table S2). Ethical approval was obtained from the ethics committee at the Medical University of Graz, Austria (no. 33-144 ex 20/21). The Severe Acute Respiratory Syndrome Coronavirus 2 Re-Infection Risk and Vaccine Efficacy Study in Austria (SARIVA) is registered at clinicaltrials.gov (NCT06162533) and was funded by the Austrian Science Fund (FWF) KLI 1188.

### Statistical analysis

Categorical data are presented as percentages, and continuous data are shown as medians (with 25^th^ to 75^th^ percentile). We used Cox regression analyses to calculate hazard ratios (HRs) with 95% confidence intervals (CI) for non-COVID-19 and all-cause mortality in groups according to the number of vaccinations against SARS-CoV-2. Analyses were restricted to individuals with a SARS-CoV-2 infection at least 90 days before the start of the observation period. Censoring occurred at the end of the respective observation period, or the date of death. Individuals changed groups on the day of vaccination.

Proportional hazards assumptions were checked by graphically examining Schoenfeld residuals. In the case of violation, we planned to split the observation period. Cox proportional HR were only calculated for analyses with at least 10 outcome events. Statistical analyses were performed in R (version 4.4.1).^22^

#### Mortality analyses in different time periods

For the entire observation period, we calculated HRs for vaccinated individuals versus unvaccinated individuals. Individuals were tracked starting 90 days after their first infection. We adjusted for age, gender and nursing home residency, which allows to study for HVE and confounding by indication that goes beyond these three factors that may readily affect the chances of getting vaccinated.

We specifically analysed the two consecutive months of the years 2021 to 2023 with the lowest COVID-19 mortality rates. Here we excluded individuals with a SARS-CoV-2 infection within the previous 90 days to reduce the potential impact of recent infections on outcome measures (higher mortality in recently infected patients) and group allocation (e.g., vaccination is unlikely in recently infected individuals). This approach allows to evaluate “off-season” VE estimates in times in which the virus is hardly circulating and thus no or only minimal VE should be measurable.^7^ Additional analyses were performed in the respective two months with the highest COVID-19 mortality rates.^23^ If SARS-CoV-2 vaccines protect against non-COVID mortality (e.g., due to post SARS-CoV-2 infection sequelae or death misclassifications), mortality reductions in vaccinated versus unvaccinated individuals should be significantly more pronounced in time periods with high versus low COVID-19 mortality rates. These analyses were additionally adjusted for year of last infection to account for potential effects of COVID-19 post-acute sequelae.

For sensitivity analyses we stratified by gender, age groups (18-39, 40-59, 60-74, 75-84 and 85+ years old), nursing home residency, and year of previous SARS-CoV-2 infections. We also stratified for individuals with and without an incident SARS-CoV-2 infection and according to mRNA and non-mRNA vaccines.

To investigate the time period close to vaccination, we matched (1 to 1) individuals who received a vaccination for each calendar day to unvaccinated controls, based on sex (binary variable), age group (18 to 24 years, then 5 years intervals up to 95+) and nursing home residency (binary variable). The unvaccinated control group had no documented vaccinations up to 14 days after the relevant matched vaccination day.

#### Control outcomes and COVID-19 mortality analyses

Analyses were performed for cancer mortality (ICD10: C00-C99) and transport accident mortality (ICD10: V00-V99) as control outcomes. These are proxies for worse physical health and risk behaviour, respectively, that should not be affected by vaccination. Therefore, group differences for these outcomes may indicate bias. Moreover, we calculated group differences for COVID-19 mortality to estimate VE (1-HR).

## RESULTS

### Study population

On January 1^st^, 2021, the adult population in Austria comprised 7,417,611 individuals (51.18% females) with a median age (25 to 75^th^ percentile) of 50 years (34-64).^24^ In addition, all younger individuals with a documented infection who turned 18 years before 2024 (n= 189,567), were considered for study inclusion. Participant flow-chart is shown in Figure 1.

### Analyses of overall observation period

Characteristics of all eligible individuals between 2021 and 2023 are shown in Table S3. Deaths per 100,000 person days were 2.23 and 2.28 for non-COVID-19 and all-cause mortality respectively (Table S2). Due to violations of the proportional hazard assumption, we split the observation period between into three-month blocks. We present overall and split estimates (Figure 2, Table 1, S4 and S5). Adjusted HRs (aHR) indicate modestly lower all-cause mortality and non-COVID-19 mortality in vaccinated versus unvaccinated individuals irrespective of the number of vaccinations (Table 1 and Figure 2). Mortality was much lower in the vaccinated than unvaccinated groups in 2021, but this difference diminished later and even reversed for some analyses (Figure 2 and Table S5). E.g., HR for non-COVD-19 deaths was 0.69 (0.59 - 0.81), 0.65 (0.58 - 0.74), and 0.56 (0.48 - 0.66) for 1-, 2-, and 3-vaccinations, respectively in Q4 2021, but became 1.28 (1.08 - 1.51), 1.25 (1.14 - 1.37), and 1.08 (1.00 - 1.17) in Q4 2022. This reversal happened by Q2 2022 for 1- and 2-dose groups, and by Q4 2022 for the 3-dose group, while initial mortality differences for those with 4+ vaccinations only diminished in magnitude (Figure 2).

**Table 1.**
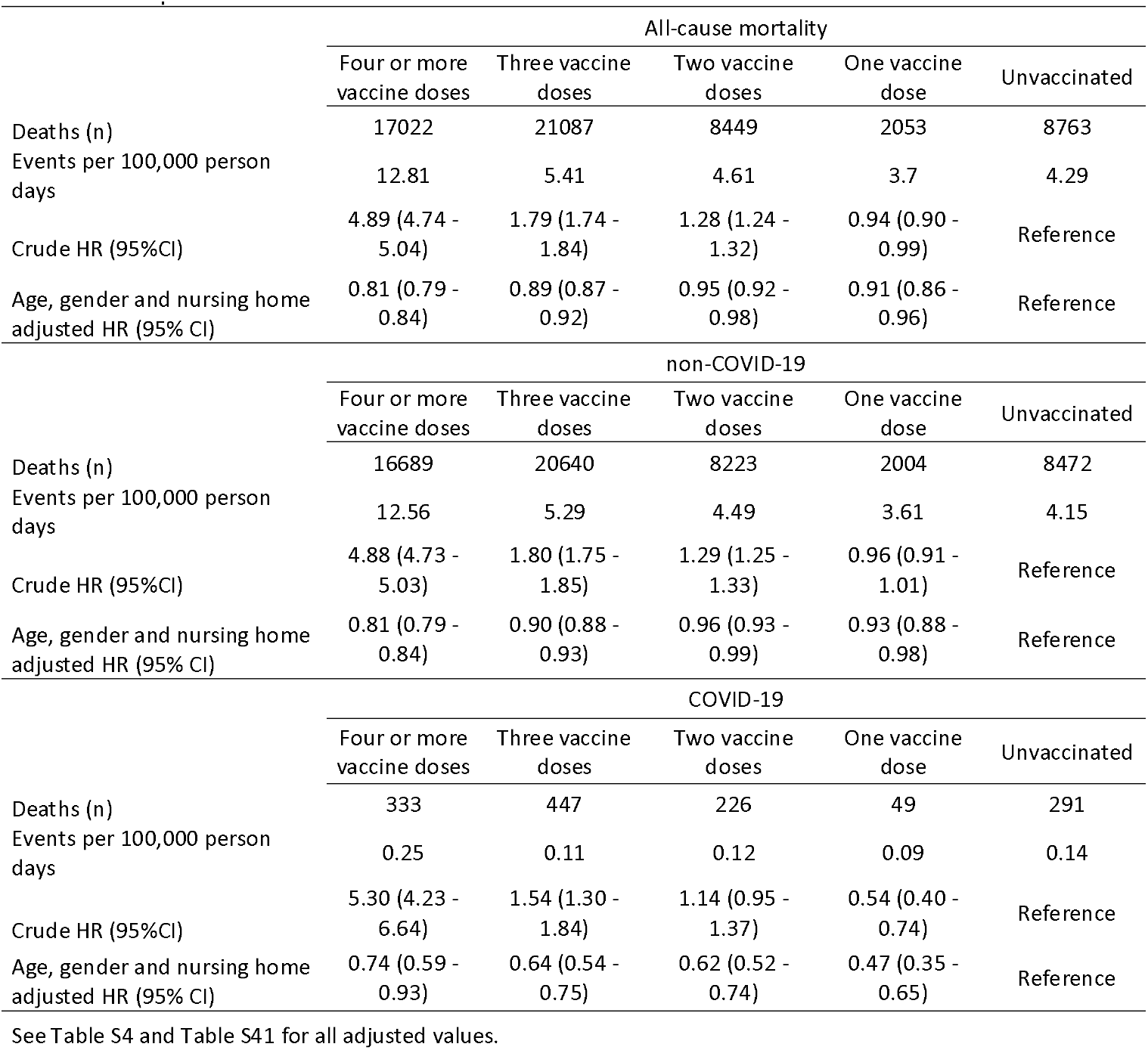
Hazard ratios (HR) with 95% confidence intervals (95% CI) for all-cause, non-COVID-19 and COVID-19 mortality according to number of SARS-CoV-2 vaccine doses over the entire observation period.

**Figure 2:**
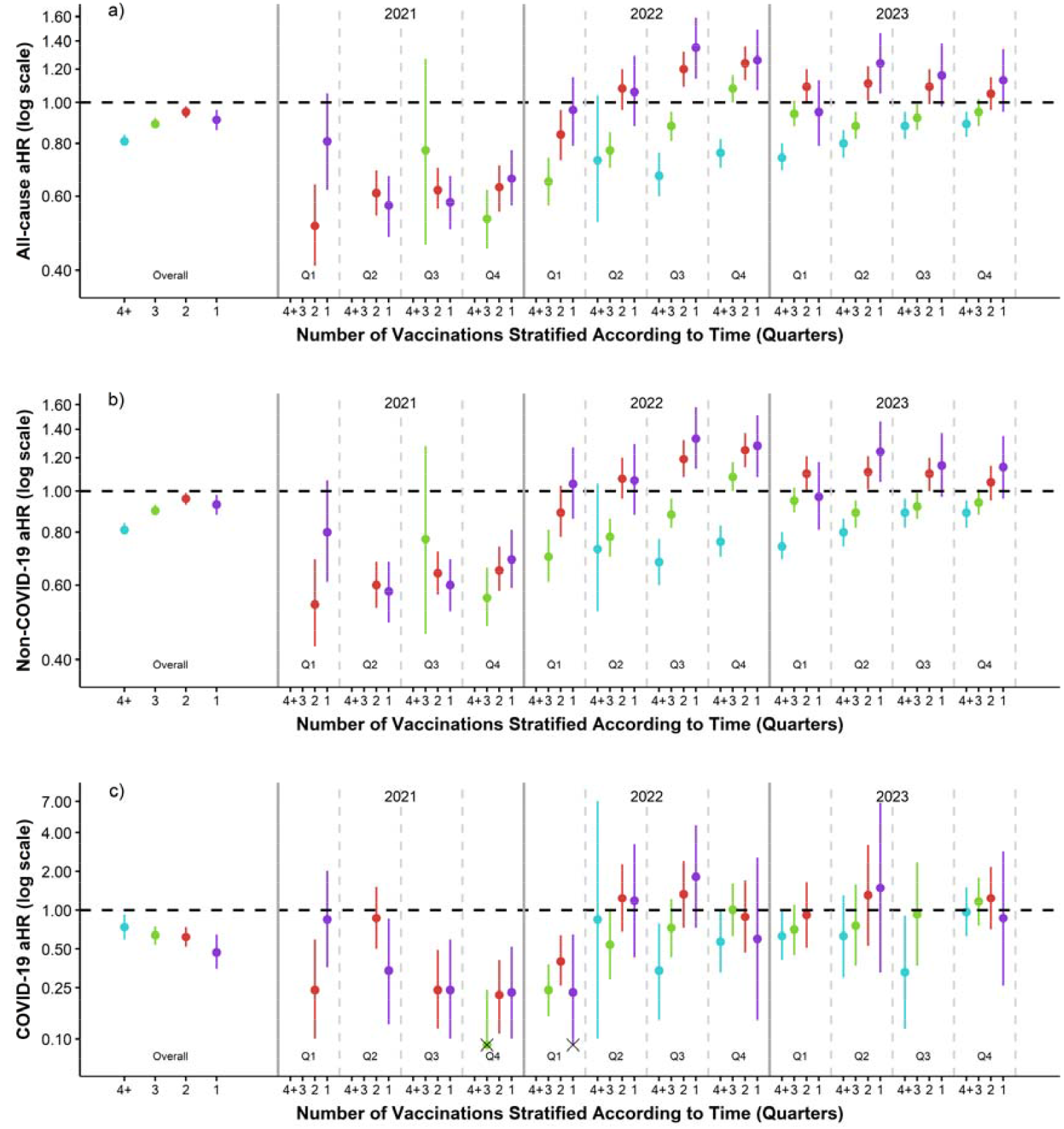
Age, gender and nursing home adjusted HRs and their CIs for all-cause mortality (a), non-COVID-19 mortality (b) and COVID-19 mortality (c) over the whole observation period, as well as split into three-month intervals by number of vaccinations. Unvaccinated individuals as the reference group. Note differences in the y-axis scaling. See Table 1, S4, S5, S41 and S42 for details and crude estimates. Confidence values with lower bounds below 0.09 are indicated by a black x.

### Analyses during periods with high and low COVID-19 mortality rates

Baseline characteristics and COVID-19 mortality rates for these analyses are shown in Tables S2 and S6-S11. Respective results of Cox regression analyses are shown in Table 2 (Table S12 for a detailed version). Vaccinated individuals had significantly decreased all-cause and non-COVID-19 mortality risk in 2021, for both low and high COVID-19 mortality time periods. In 2022, this trend was also tentatively present in both periods. In both periods of 2023, there was no difference between vaccinated and unvaccinated groups other than the 4+ vaccinations group which had lower all-cause and non-COVID-19 mortality risk in both periods.

**Table 2.**
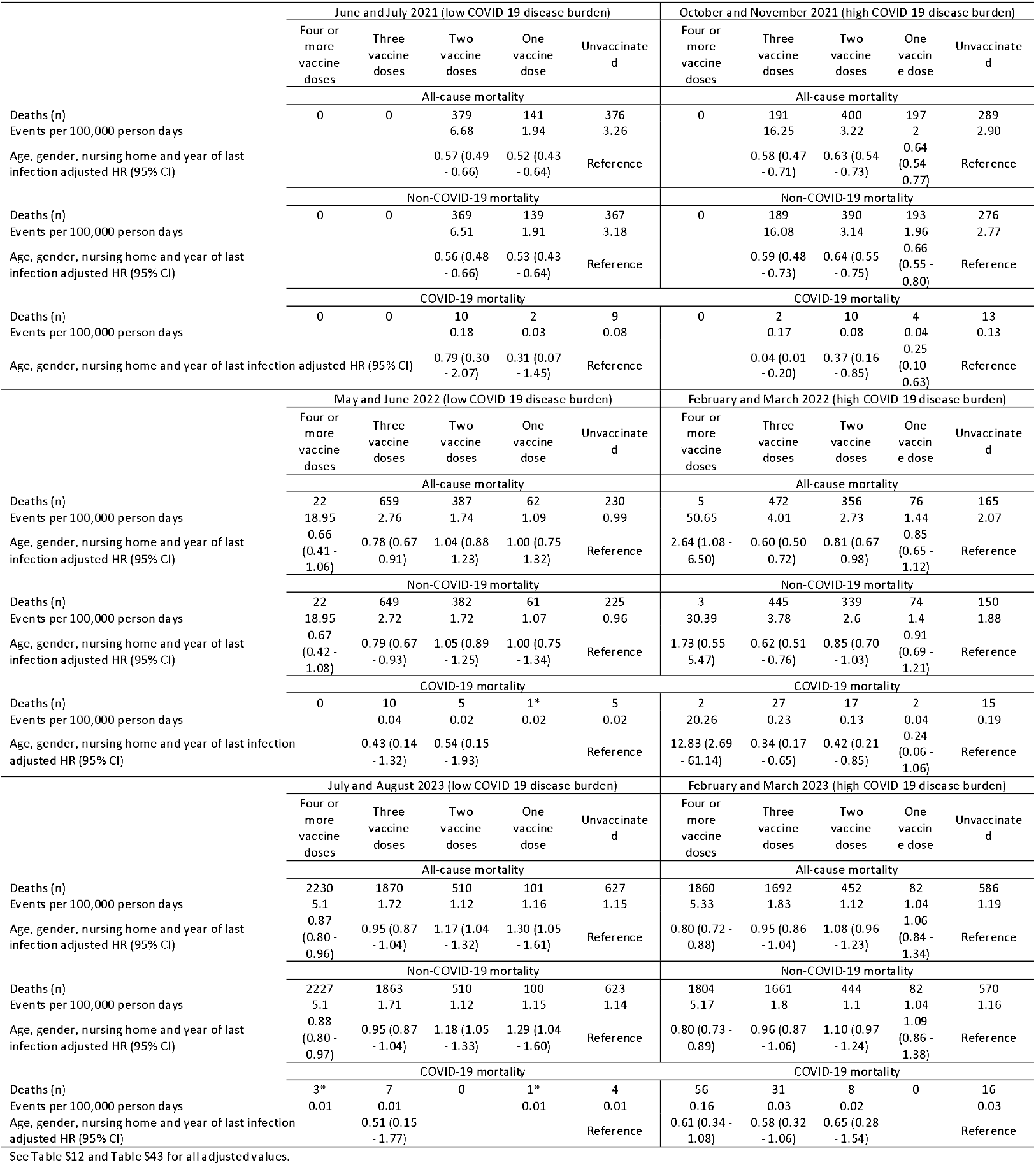
Hazard ratios (HR) with 95% confidence intervals (95% CI) for non-COVID-19 and all-cause mortality according to number of SARS-CoV-2 vaccine doses during different time periods of high and low COVID-19 disease burden.

### Sensitivity analyses

Stratified results of calendar years split quarterly and periods of low and high COVID-19 mortality can be found in Tables S13-S21 and Tables S22-S37, respectively. These generally mirror the main results showing lower mortality risk for vaccinated versus unvaccinated individuals in earlier periods (especially 2021) which wanes in later periods for all groups other than the 4+ vaccination group.

### Analyses on control outcomes

Analyses are shown in Table S38-S40. Cancer mortality reflected the main results, showing a tentatively lower risk (significant for 1-dose and 4+ doses) over the whole observation period. Split analysis and periods of low and high COVID-19 mortality both showed significant lower cancer mortality risk in earlier parts of the pandemic which gradually vanished. Transport accident deaths showed a similar tendency but for several analyses but statistical power was low.

### COVID-19 mortality

COVID-19 mortality analyses are shown in Tables S41-S43. Adjusted HRs show a significant lower risk in the vaccinated irrespective of number of vaccinations (Table S41). While quarterly and low/high COVID-19 disease burden period analyses indicate similar results to the main analyses, the number of outcomes were very low.

### Matched analyses

Matching analysis shows significantly reduced mortality risk in vaccinated compared to unvaccinated individuals within two weeks after vaccination. The HRs are all below 0.5 irrespective of vaccination numbers (Table 3). Cancer mortality (Table S45) and COVID-19 mortality (Table 3) mirrored these results.

**Table 3.**
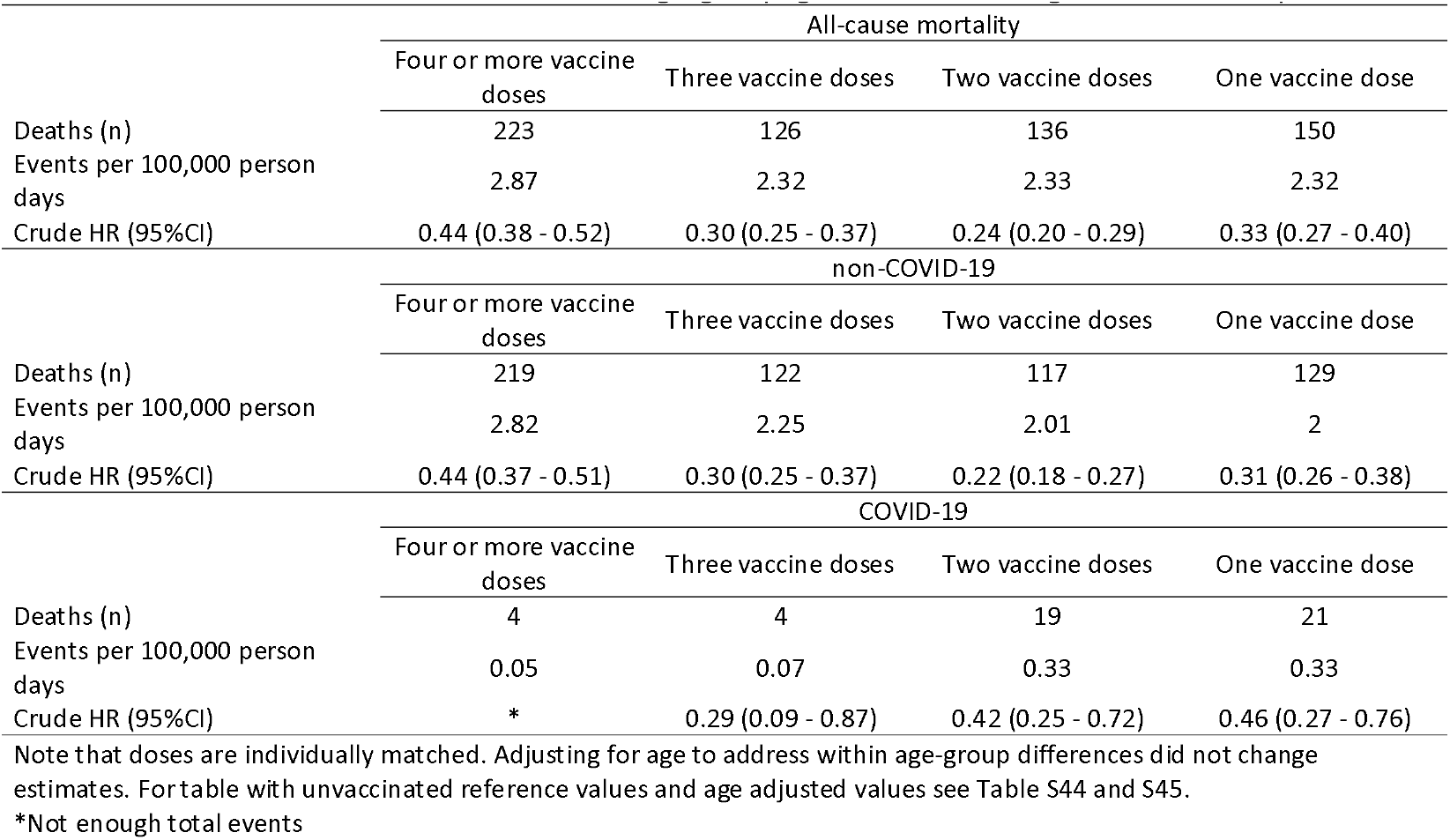
Hazard ratios (HR) with 95% confidence intervals (95% CI) for all-cause, non-COVID-19 and COVID-19 mortality according to number of SARS-CoV-2 vaccine doses in the two weeks after vaccination, with controls matched based on age-group, gender and nursing home residency.

| All-cause mortality |  |  |  |  |
| --- | --- | --- | --- | --- |
|  | Four or more vaccine doses | Three vaccine doses | Two vaccine doses | One vaccine dose |
| Deaths (n) | 223 | 126 | 136 | 150 |
| Events per 100,000 person days | 2.87 | 2.32 | 2.33 | 2.32 |
| Crude HR (95%CI) | 0.44 (0.38 - 0.52) | 0.30 (0.25 - 0.37) | 0.24 (0.20 - 0.29) | 0.33 (0.27 - 0.40) |
| non-COVID-19 |  |  |  |  |
|  | Four or more vaccine doses | Three vaccine doses | Two vaccine doses | One vaccine dose |
| Deaths (n) | 219 | 122 | 117 | 129 |
| Events per 100,000 person days | 2.82 | 2.25 | 2.01 | 2 |
| Crude HR (95%CI) | 0.44 (0.37 - 0.51) | 0.30 (0.25 - 0.37) | 0.22 (0.18 - 0.27) | 0.31 (0.26 - 0.38) |
| COVID-19 |  |  |  |  |
|  | Four or more vaccine doses | Three vaccine doses | Two vaccine doses | One vaccine dose |
| Deaths (n) | 4 | 4 | 19 | 21 |
| Events per 100,000 person days | 0.05 | 0.07 | 0.33 | 0.33 |
| Crude HR (95%CI) | * | 0.29 (0.09 - 0.87) | 0.42 (0.25 - 0.72) | 0.46 (0.27 - 0.76) |

Note that doses are individually matched. Adjusting for age to address within age-group differences did not change estimates. For table with unvaccinated reference values and age adjusted values see Table S44 and S45.
\*Not enough total events

## DISCUSSION

In previously SARS-CoV-2 infected adults in Austria, we observed significantly lower non-COVID-19 and all-cause mortality in the hybrid immune population versus the natural immune population suggesting the presence of a healthy user bias where individuals with lower mortality risk are more likely to get vaccinated. This bias was especially pronounced close to vaccination. Moreover, this HVE was more prominent in 2021, but it progressively waned thereafter until it vanished or even got reversed in 2022 and 2023 for some vaccination groups.

Our results add to the growing number of studies showing different non-COVID-19 or all-cause mortality in vaccinated versus unvaccinated populations.^4,14–17,25,26^ Reductions in non-COVID-19 mortality in vaccinated versus unvaccinated individuals were particularly pronounced during and shortly after times of high vaccine rollout. These rollouts occurred during summer 2021 for the first and second vaccination, at the end of 2021 for the third vaccination, and in autumn 2022 for the fourth vaccination (Figure S1). The pattern of reduced non-COVID-19 mortality shortly after vaccinations indicates that good health status is associated with vaccination. This reflects an HVE that outweighs any confounding by indication. Over time, this HVE waned or even reversed as indicated by progressively increasing aHRs in non-COVID mortality in vaccinated versus unvaccinated individuals.

One may argue whether group differences in non-COVID-19 mortality may be explained by COVID-19 deaths being misclassified as non-COVID-19 deaths or patients who succumbed to deaths that were classified as non-COVID-19 but were prompted by COVID-19 sequelae. In this context, it should be noted that in 2021, the difference in COVID-19 mortality rates between periods with high versus low SARS-CoV-2 infection rates was particularly significant providing a good opportunity to evaluate whether mortality differences indicative of the HVE are potentially altered depending on COVID-19 disease burden (Table S2). As we observed similar reductions of non-COVID-19 mortality risk in vaccinated versus unvaccinated individuals during both time periods in 2021, this argues for bias consistent with HVE. Therefore, misclassifications of COVID-19 deaths as non-COVID-19 deaths or COVID-19 post-acute sequela are probably not significantly impacting these results. These findings are consistent with previous reports.^17,27^ Importantly, analyses within two weeks after vaccination indicate a significantly reduced mortality in vaccinated versus unvaccinated individuals; in these two weeks, any VE against COVID-19 is unlikely to have been substantial so that these analyses strongly support the presence of a HVE.

Group differences in mortality were similar regardless of whether non-COVID-19, all-cause or cancer mortality were analysed, thus further supporting early HVE. While older people and those in nursing home facilities were more likely to get vaccinated earlier and more often compared to younger people, the adjusted analyses indicate that HVE outweighs confounding by indications other than age and nursing home residency. Confounding by indication, if present, may have a greater effect in older population that more frequently suffer from various co-morbidities. However, our age stratified analyses suggest that even in older populations, such potential confounding by indication was not strong enough to reverse the reduced non-COVID-19 mortality in vaccinated individuals.

Analyses on COVID-19 mortality confirm established knowledge on VE. While it is beyond the scope of this work to evaluate approaches on how to correct VE estimates for biases consistent with an HVE, the magnitude of VE for the first three vaccinations were numerically greater than for the HVE. Therefore, simply correcting VE for HVE (e.g., by dividing the two HR estimates) would still suggest that vaccines were substantially effective for prevention of COVID-19 deaths, although VE might have been overestimated. However, such simple correction may not necessarily be accurate.

As current and future vaccination policies are based on estimates of VE, and almost everyone has been previously infected, it is useful to further investigate the role of HVE on hybrid immunity estimates.^28^ Interestingly, some previous study results suggest faster waning immunity in individuals with hybrid compared to natural immunity.^6,28^ Apart from waning VE, this might hypothetically be explained by differences in general health between the groups at the time of vaccination (i.e. HVE), which diminish over time.

Our study is limited as we had no access to data on co-morbidities or other potential confounders such as measures of functional or socioeconomic status and could therefore not adjust for these parameters. Consideration of nursing home residency works in part as a proxy for comorbidities. Nevertheless, previous investigations on HVE indicate only a minor impact of adjustments for other co-morbidities or even an increase in HVE estimates by such additional adjustments.^7,29^ The main strengths of our study are the nationwide population, the follow-up period throughout the whole COVID-19 pandemic, and the confirmation of our main findings during different time frames, in various sensitivity and subgroup analyses.

In conclusion, we show evidence for a significant HVE in previously infected adults in Austria, reflected by lower non-COVID-19 and all-cause mortality in the vaccinated versus unvaccinated groups. This HVE was particularly pronounced during and shortly after times of high vaccination rollout but diminished or even got reversed with further follow-up. Future epidemiological studies should consider HVE in interpreting SARS-CoV-2 VE.

## Data Availability

The data that support the findings of this study are available upon request with approval needed from the Austrian Agency for Health and Food Safety (AGES), Vienna, Austria and Statistics Austria respectively.

## Contributors

UR and SP conceptualized the study with contributions of JPAI, TBH and DW. UR and SP wrote the original draft. UR performed the formal analyses and visualization. AC and LR were involved in data curation. All authors contributed to supervision, writing, reviewing, and editing the manuscript, and approved the final version before submission.

## Data sharing statement

The data that support the findings of this study are available upon request with approval needed from the Austrian Agency for Health and Food Safety (AGES), Vienna, Austria and Statistics Austria respectively. The data are not publicly available due to restrictions pertaining to contained information that could compromise the privacy of patients.

## Funding

This study was funded by the Austrian Science Fund (FWF) KLI 1188.

## Declaration of interests

The authors declare no conflict of interests.

## Acknowledgements

The authors thank all persons and organizations involved in data collection.

## Notes

### Competing Interest Statement

The authors have declared no competing interest.

### Author Declarations

Ethics committee at the Medical University of Graz approved this work (no. 33- 144 ex 20/21)

### Summary of Updates

Corrected an error in the analysis which led to controls in matching potentially having a previous vaccination. Also adjusted the matched cancer mortality results, as previously the table showed results for non-cancer outcomes.

## REFERENCES

1. Graña C, Ghosn L, Evrenoglou T, et al. Efficacy and safety of COVID-19 vaccines. Cochrane Emergency and Critical Care Group, ed. Cochrane Database of Systematic Reviews. 2022;2023(3). doi:10.1002/14651858.CD015477

2. Ioannidis JPA. Factors influencing estimated effectiveness of COVID-19 vaccines in non-randomised studies. BMJ Evid Based Med. 2022;27(6):324–329. doi:10.1136/bmjebm-2021-111901

3. Fung K, Jones M, Doshi P. Sources of bias in observational studies of covid-19 vaccine effectiveness. J Eval Clin Pract. 2024;30(1):30–36. doi:10.1111/jep.13839

4. Høeg TB, Duriseti R, Prasad V. Potential “Healthy Vaccinee Bias” in a Study of BNT162b2 Vaccine against Covid-19. N Engl J Med. 2023;389(3):284–285. doi:10.1056/NEJMc2306683

5. Berrino F, Donzelli A, Bellavite P, Malatesta G. COVID-19 vaccination and all-cause and non-COVID-19 mortality. A revaluation of a study carried out in an Italian Province. Epidemiol Prev. 2023;47(6):374–378. doi:10.19191/EP23.6.A643.075

6. Bobrovitz N, Ware H, Ma X, et al. Protective effectiveness of previous SARS-CoV-2 infection and hybrid immunity against the omicron variant and severe disease: a systematic review and meta-regression. The Lancet Infectious Diseases. 2023;23(5):556–567. doi:10.1016/S1473-3099(22)00801-5

7. Remschmidt C, Wichmann O, Harder T. Frequency and impact of confounding by indication and healthy vaccinee bias in observational studies assessing influenza vaccine effectiveness: a systematic review. BMC Infectious Diseases. 2015;15(1):429. doi:10.1186/s12879-015-1154-y

8. Hosseini-Moghaddam SM, He S, Calzavara A, Campitelli MA, Kwong JC. Association of Influenza Vaccination With SARS-CoV-2 Infection and Associated Hospitalization and Mortality Among Patients Aged 66 Years or Older. JAMA Network Open. 2022;5(9):e2233730. doi:10.1001/jamanetworkopen.2022.33730

9. McCarthy NL, Weintraub E, Vellozzi C, et al. Mortality rates and cause-of-death patterns in a vaccinated population. Am J Prev Med. 2013;45(1):91–97. doi:10.1016/j.amepre.2013.02.020

10. Jackson LA, Nelson JC, Benson P, et al. Functional status is a confounder of the association of influenza vaccine and risk of all cause mortality in seniors. International Journal of Epidemiology. 2006;35(2):345–352. doi:10.1093/ije/dyi275

11. Tielemans SMAJ, de Melker HE, Hahné SJM, et al. Non-specific effects of measles, mumps, and rubella (MMR) vaccination in high income setting: population based cohort study in the Netherlands. BMJ. 2017;358:j3862. doi:10.1136/bmj.j3862

12. Donzelli A. Influenza Vaccination of Pregnant Women and Serious Adverse Events in the Offspring. Int J Environ Res Public Health. 2019;16(22):4347. doi:10.3390/ijerph16224347

13. Donzelli A. Influenza Vaccinations for All Pregnant Women? Better Evidence Is Needed. Int J Environ Res Public Health. 2018;15(9):2034. doi:10.3390/ijerph15092034

14. Pálinkás A, Sándor J. Effectiveness of COVID-19 Vaccination in Preventing All-Cause Mortality among Adults during the Third Wave of the Epidemic in Hungary: Nationwide Retrospective Cohort Study. Vaccines (Basel). 2022;10(7):1009. doi:10.3390/vaccines10071009

15. Fürst T, Straka R, Janošek J. Healthy vaccinee effect: a bias not to be forgotten in observational studies on COVID-19 vaccine effectiveness. Pol Arch Intern Med. 2024;134(2). doi:10.20452/pamw.16634

16. Xu S, Huang R, Sy LS, et al. COVID-19 Vaccination and Non-COVID-19 Mortality Risk - Seven Integrated Health Care Organizations, United States, December 14, 2020-July 31, 2021. MMWR Morb Mortal Wkly Rep. 2021;70(43):1520–1524. doi:10.15585/mmwr.mm7043e2

17. Fürst T, Bazalová A, Fryčák T, Janošek J. Does the healthy vaccinee bias rule them all? Association of COVID-19 vaccination status and all-cause mortality from an analysis of data from 2.2 million individual health records. International Journal of Infectious Diseases. 2024;142:106976. doi:10.1016/j.ijid.2024.02.019

18. Arbel R, Hammerman A, Sergienko R, et al. BNT162b2 Vaccine Booster and Mortality Due to Covid-19. New England Journal of Medicine. 2021;385(26):2413–2420. doi:10.1056/NEJMoa2115624

19. Chalupka A, Riedmann U, Richter L, et al. Effectiveness of the First and Second Severe Acute Respiratory Syndrome Coronavirus 2 Vaccine Dose: A Nationwide Cohort Study From Austria on Hybrid Versus Natural Immunity. Open Forum Infectious Diseases. 2024;11(10):ofae547. doi:10.1093/ofid/ofae547

20. Pilz S, Chakeri A, Ioannidis JP, et al. SARS-CoV-2 re-infection risk in Austria. Eur J Clin Invest. 2021;51(4):e13520. doi:10.1111/eci.13520

21. Chalupka A, Richter L, Chakeri A, et al. Effectiveness of a fourth SARS-CoV-2 vaccine dose in previously infected individuals from Austria. European Journal of Clinical Investigation. 2024;54(3):e14136. doi:10.1111/eci.14136

22. R Core Team: A Language and Environment for Statistical Computing. Accessed September 2, 2024. https://www.R-project.org/.

23. Riedmann U, Chalupka A, Richter L, et al. COVID-19 case fatality rate and infection fatality rate from 2020 to 2023: Nationwide analysis in Austria. Journal of Infection and Public Health. 2025;18(4):102698. doi:10.1016/j.jiph.2025.102698

24. Bevölkerung nach Alter/Geschlecht. STATISTIK AUSTRIA. Accessed December 26, 2024. https://www.statistik.at/statistiken/bevoelkerung-und-soziales/bevoelkerung/bevoelkerungsstand/bevoelkerung-nach-alter/geschlecht

25. Arbel R, Sergienko R, Netzer D, et al. Potential “Healthy Vaccinee Bias” in a Study of BNT162b2 Vaccine against Covid-19. Reply. N Engl J Med. 2023;389(3):285–286. doi:10.1056/NEJMc2306683

26. Bardenheier BH, Gravenstein S, Blackman C, et al. Adverse events following mRNA SARS-CoV-2 vaccination among U.S. nursing home residents. Vaccine. 2021;39(29):3844–3851. doi:10.1016/j.vaccine.2021.05.088

27. Barda N, Dagan N, Cohen C, et al. Effectiveness of a third dose of the BNT162b2 mRNA COVID-19 vaccine for preventing severe outcomes in Israel: an observational study. The Lancet. 2021;398(10316):2093–2100. doi:10.1016/S0140-6736(21)02249-2

28. Riedmann U, Chalupka A, Richter L, et al. Estimates of SARS-CoV-2 Infections and Population Immunity After the COVID-19 Pandemic in Austria: Analysis of National Wastewater Data. The Journal of Infectious Diseases. Published online February 18, 2025:jiaf054. doi:10.1093/infdis/jiaf054

29. Jackson LA, Jackson ML, Nelson JC, Neuzil KM, Weiss NS. Evidence of bias in estimates of influenza vaccine effectiveness in seniors. International Journal of Epidemiology. 2006;35(2):337–344. doi:10.1093/ije/dyi274

