## Supplements for "Estimates of underlying health biases in SARS-CoV-2 vaccination recipients: a nationwide study in previously-infected adults"

**Supplements:** **Estimates of biases on SARS-CoV-2 vaccine effectiveness: a nationwide study in previously infected adults from Austria**

Uwe Riedmann; Alena Chalupka; Lukas Richter; Dirk Werber; Martin Sprenger; Peter Willeit; Marc Rijksen; Julia Lodron; Tracy Beth Høeg; John PA Ioannidis; Stefan Pilz

#### **Table S1:** STROBE Statement—Checklist of items for *cohort studies*

|  | **Item No** | **Recommendation** | **Main text page** |
| --- | --- | --- | --- |
| **Title and abstract** | 1 | (*a*) Indicate the study’s design with a commonly used term in the title or the abstract | 2 |
|  |  | (*b*) Provide in the abstract an informative and balanced summary of what was done and what was found | 2 & 3 |
| **Introduction** | | |  |
| Background/rationale | 2 | Explain the scientific background and rationale for the investigation being reported | 4 and 5 |
| Objectives | 3 | State specific objectives, including any prespecified hypotheses | 5 |
| **Methods** | | |  |
| Study design | 4 | Present key elements of study design early in the paper | 5 and 6 |
| Setting | 5 | Describe the setting, locations, and relevant dates, including periods of recruitment, exposure, follow-up, and data collection | 5 and 6 |
| Participants | 6 | (*a*) Give the eligibility criteria, and the sources and methods of selection of participants. Describe methods of follow-up | 6, 7 and 8 |
|  |  | (*b*) For matched studies, give matching criteria and number of exposed and unexposed | 7 and 8 |
| Variables | 7 | Clearly define all outcomes, exposures, predictors, potential confounders, and effect modifiers. Give diagnostic criteria, if applicable | 6, 7 and 8 |
| Data sources/ measurement | 8* | For each variable of interest, give sources of data and details of methods of assessment (measurement). Describe comparability of assessment methods if there is more than one group | 5 and 6 |
| Bias | 9 | Describe any efforts to address potential sources of bias | 7 and 8 |
| Study size | 10 | Explain how the study size was arrived at | 6 |
| Quantitative variables | 11 | Explain how quantitative variables were handled in the analyses. If applicable, describe which groupings were chosen and why | 6,7 and 8 |
| Statistical methods | 12 | (*a*) Describe all statistical methods, including those used to control for confounding | 6,7 and 8 |
|  |  | (*b*) Describe any methods used to examine subgroups and interactions | 6,7 and 8 |
|  |  | (*c*) Explain how missing data were addressed | not applicable |
|  |  | (*d*) If applicable, explain how loss to follow-up was addressed | not applicable |
|  |  | (*e*) Describe any sensitivity analyses | 7 and 8 |
| **Results** | | |  |
| Participants | 13* | (a) Report numbers of individuals at each stage of study—eg numbers potentially eligible, examined for eligibility, confirmed eligible, included in the study, completing follow-up, and analysed | 8, 9 and 10; Figure 1 |
|  |  | (b) Give reasons for non-participation at each stage | not applicable |
|  |  | (c) Consider use of a flow diagram | Figure 1 |
| Descriptive data | 14* | (a) Give characteristics of study participants (eg demographic, clinical, social) and information on exposures and potential confounders | 8, 9 and 10; Supplements (Table S3 and Tables S6-S11) |
|  |  | (b) Indicate number of participants with missing data for each variable of interest | not applicable |
|  |  | (c) Summarise follow-up time (eg, average and total amount) | not applicable |
| Outcome data | 15* | Report numbers of outcome events or summary measures over time | Table S2 |
| Main results | 16 | (*a*) Give unadjusted estimates and, if applicable, confounder-adjusted estimates and their precision (eg, 95% confidence interval). Make clear which confounders were adjusted for and why they were included | Table 1, 2 and 3; Supplements |
|  |  | (*b*) Report category boundaries when continuous variables were categorized | Table 1, 2 and 3; Supplements |
|  |  | (*c*) If relevant, consider translating estimates of relative risk into absolute risk for a meaningful time period | not applicable |
| Other analyses | 17 | Report other analyses done—eg analyses of subgroups and interactions, and sensitivity analyses | 9 and 10; Supplements |
| **Discussion** | | |  |
| Key results | 18 | Summarise key results with reference to study objectives | 10 and 11 |
| Limitations | 19 | Discuss limitations of the study, taking into account sources of potential bias or imprecision. Discuss both direction and magnitude of any potential bias | 12 and 13 |
| Interpretation | 20 | Give a cautious overall interpretation of results considering objectives, limitations, multiplicity of analyses, results from similar studies, and other relevant evidence | 10 and 11 |
| Generalisability | 21 | Discuss the generalisability (external validity) of the study results | 10 and 11 |
| **Other information** | | |  |
| Funding | 22 | Give the source of funding and the role of the funders for the present study and, if applicable, for the original study on which the present article is based | 13 |

*Give information separately for exposed and unexposed groups.

**Note:** An Explanation and Elaboration article discusses each checklist item and gives methodological background and published examples of transparent reporting. The STROBE checklist is best used in conjunction with this article (freely available on the Web sites of PLoS Medicine at http://www.plosmedicine.org/, Annals of Internal Medicine at http://www.annals.org/, and Epidemiology at http://www.epidem.com/). Information on the STROBE Initiative is available at <http://www.strobe-statement.org>.

| **Table S2:** Number of documented deaths for different time periods and outcome measures in the studied population and the COVID-19 deaths for the whole population | | | | | | |
| --- | --- | --- | --- | --- | --- | --- |
|  | **cause of death** | | | | | **whole population** |
|  | **All-Cause** | **Non-COVDI-19** | **Cancer** | **Transport Accidents** | **COVID-19** | **COVID-19** |
| **2021-2023 deaths** | 57,374 | 56,028 | 13,789 | 303 | 1,346 | 15,238 |
| **Deaths per 100,000 person days** | 2.28 | 2.23 | 0.55 | 0.01 | 0.05 | N/A |
| **low periods** |  |  |  |  |  |  |
| June/July 2021 | 896 | 875 | 183 | 7 | 21 | 149 |
| May/June 2022 | 4,611 | 4,534 | 1,049 | 14 | 77 | 453 |
| July/Aug 2023 | 4,834 | 4,790 | 1,147 | 26 | 44 | 22 |
| **high periods** |  |  |  |  |  |  |
| Oct/Nov 2021 | 1,102 | 1,068 | 225 | 1 | 34 | 1,702 |
| Feb/March 2022 | 1,074 | 1,011 | 226 | 3 | 63 | 2,071 |
| Feb/March 2023 | 4,672 | 4,561 | 1,153 | 24 | 111 | 637 |

### **Overall analysis**

| **Table S3:** Characteristics of the population used in the overall analysis | | | | | | | | |
| --- | --- | --- | --- | --- | --- | --- | --- | --- |
|  | All | Male | Female | Unvaccinated | one dose | two doses | three doses | four or more doses |
| No. of participants | 4324485 | 2051494 | 2272991 | 896242 | 144264 | 754912 | 1798501 | 730566 |
| Age, year (median, IQR)* | 46 (33-59) | 45 (31.5-58.5) | 46 (32.5-59.5) | 42 (30.5-53.5) | 38 (26.5-49.5) | 38 (26.5-49.5) | 46 (33.5-58.5) | 61 (48-74) |
| Nursing home residency (%) | 1.21 | 0.71 | 1.66 | 0.47 | 0.65 | 0.63 | 0.86 | 3.69 |
| Repeated previous infections (%) | 16.34 | 14.58 | 17.92 | 23.11 | 33.59 | 19.1 | 13.23 | 9.41 |
| Last infection in 2020 (%) | 4.29 | 4.63 | 3.98 | 2.28 | 9.8 | 7.05 | 3.86 | 3.86 |
| Last infection in 2021 (%) | 11.11 | 12.1 | 10.22 | 19.25 | 31.53 | 14.96 | 6.18 | 5.24 |
| Last infection in 2022 (%) | 76.84 | 76.31 | 77.33 | 75.04 | 54.32 | 72.44 | 81.11 | 77.55 |
| *Age is calculated for the 01.01.2024 and as if everyone was alive. | | | | | | | | |

| **Table S4:** Hazard ratios (HR) with 95% confidence intervals (95% CI) for non-COVID 19 and all-cause mortality according to number of SARS-CoV-2 vaccine doses over the entire period. | | | | | |
| --- | --- | --- | --- | --- | --- |
|  | All-cause mortality | | | | |
|  | Four or more vaccine doses | Three vaccine doses | Two vaccine doses | One vaccine dose | Unvaccinated |
| Deaths (n) | 17022 | 21087 | 8449 | 2053 | 8763 |
| Events per 100,000 person days | 12.81 | 5.41 | 4.61 | 3.7 | 4.29 |
| Crude HR (95%CI) | 4.89 (4.74 - 5.04) | 1.79 (1.74 - 1.84) | 1.28 (1.24 - 1.32) | 0.94 (0.90 - 0.99) | Reference |
| Age adjusted HR (95% CI) | 0.91 (0.89 - 0.94) | 0.97 (0.94 - 0.99) | 1.03 (1.00 - 1.07) | 0.91 (0.87 - 0.96) | Reference |
| Age and gender adjusted HR (95% CI) | 0.89 (0.86 - 0.92) | 0.95 (0.92 - 0.97) | 1.03 (0.99 - 1.06) | 0.90 (0.86 - 0.95) | Reference |
| Age, gender and nursing home adjusted HR (95% CI) | 0.81 (0.79 - 0.84) | 0.89 (0.87 - 0.92) | 0.95 (0.92 - 0.98) | 0.91 (0.86 - 0.96) | Reference |
|  | non-COVID-19 | | | | |
|  | Four or more vaccine doses | Three vaccine doses | Two vaccine doses | One vaccine dose | Unvaccinated |
| Deaths (n) | 16689 | 20640 | 8223 | 2004 | 8472 |
| Events per 100,000 person days | 12.56 | 5.29 | 4.49 | 3.61 | 4.15 |
| Crude HR (95%CI) | 4.88 (4.73 - 5.03) | 1.80 (1.75 - 1.85) | 1.29 (1.25 - 1.33) | 0.96 (0.91 - 1.01) | Reference |
| Age adjusted HR (95% CI) | 0.92 (0.89 - 0.94) | 0.97 (0.95 - 1.00) | 1.05 (1.01 - 1.08) | 0.93 (0.88 - 0.98) | Reference |
| Age and gender adjusted HR (95% CI) | 0.89 (0.86 - 0.92) | 0.95 (0.93 - 0.98) | 1.04 (1.01 - 1.07) | 0.92 (0.88 - 0.97) | Reference |
| Age, gender and nursing home adjusted HR (95% CI) | 0.81 (0.79 - 0.84) | 0.90 (0.88 - 0.93) | 0.96 (0.93 - 0.99) | 0.93 (0.88 - 0.98) | Reference |

| **Table S5:** Hazard ratios (HR) with 95% confidence intervals (95% CI) for **non-COVID 19 and all-cause mortality** according to number of SARS-CoV-2 vaccine doses for the period from 2021 to 2023 split into 3-month intervals. | | | | | | | | | | | | | | | | | | | | |
| --- | --- | --- | --- | --- | --- | --- | --- | --- | --- | --- | --- | --- | --- | --- | --- | --- | --- | --- | --- | --- |
|  | 2021 | | | | | | | | | | | | | | | | | | | |
|  | first quarter | | | | | second quarter | | | | | third quarter | | | | | fourth quarter | | | | |
|  | Four or more vaccine doses | Three vaccine doses | Two vaccine doses | One vaccine dose | Unvaccinated | Four or more vaccine doses | Three vaccine doses | Two vaccine doses | One vaccine dose | Unvaccinated | Four or more vaccine doses | Three vaccine doses | Two vaccine doses | One vaccine dose | Unvaccinated | Four or more vaccine doses | Three vaccine doses | Two vaccine doses | One vaccine dose | Unvaccinated |
|  | All-cause mortality | | | | | All-cause mortality | | | | | All-cause mortality | | | | | All-cause mortality | | | | |
| Deaths (n) |  |  | 146 | 70 | 399 |  |  | 555 | 187 | 704 |  | 19 | 717 | 270 | 587 |  | 415 | 594 | 283 | 423 |
| Events per 100,000 person days |  |  | 20.07 | 9.23 | 2.92 |  |  | 13.66 | 2.75 | 3.33 |  | 39.24 | 5.35 | 2.35 | 3.14 |  | 9.99 | 3.36 | 2.13 | 3.19 |
| Crude HR (95%CI) |  |  | 6.90 (5.68 - 8.39) | 3.42 (2.63 - 4.43) | Reference |  |  | 4.35 (3.88 - 4.87) | 0.87 (0.74 - 1.03) | Reference |  | 13.45 (8.24 - 21.93) | 1.73 (1.55 - 1.93) | 0.76 (0.66 - 0.88) | Reference |  | 4.19 (3.58 - 4.90) | 1.07 (0.94 - 1.21) | 0.68 (0.59 - 0.79) | Reference |
| Age adjusted HR (95% CI) |  |  | 0.88 (0.71 - 1.09) | 0.99 (0.76 - 1.28) | Reference |  |  | 0.77 (0.69 - 0.87) | 0.57 (0.48 - 0.67) | Reference |  | 1.05 (0.64 - 1.72) | 0.73 (0.65 - 0.81) | 0.57 (0.50 - 0.66) | Reference |  | 0.69 (0.60 - 0.80) | 0.67 (0.59 - 0.75) | 0.64 (0.55 - 0.74) | Reference |
| Age and gender adjusted HR (95% CI) |  |  | 0.91 (0.74 - 1.13) | 0.99 (0.76 - 1.28) | Reference |  |  | 0.78 (0.69 - 0.87) | 0.56 (0.47 - 0.66) | Reference |  | 1.08 (0.66 - 1.77) | 0.72 (0.64 - 0.80) | 0.56 (0.49 - 0.65) | Reference |  | 0.69 (0.60 - 0.80) | 0.66 (0.58 - 0.75) | 0.62 (0.53 - 0.72) | Reference |
| Age, gender and nursing home adjusted HR (95% CI) |  |  | 0.51 (0.41 - 0.64) | 0.81 (0.62 - 1.05) | Reference |  |  | 0.61 (0.54 - 0.69) | 0.57 (0.48 - 0.67) | Reference |  | 0.77 (0.46 - 1.27) | 0.62 (0.56 - 0.70) | 0.58 (0.50 - 0.67) | Reference |  | 0.53 (0.45 - 0.62) | 0.63 (0.55 - 0.71) | 0.66 (0.57 - 0.77) | Reference |
|  | Non-COVID-19 mortality | | | | | Non-COVID-19 mortality | | | | | Non-COVID-19 mortality | | | | | Non-COVID-19 mortality | | | | |
| Deaths (n) |  |  | 139 | 63 | 367 |  |  | 526 | 182 | 669 |  | 19 | 706 | 264 | 558 |  | 410 | 580 | 276 | 395 |
| Events per 100,000 person days |  |  | 19.11 | 8.31 | 2.68 |  |  | 12.94 | 2.68 | 3.16 |  | 39.24 | 5.27 | 2.3 | 2.99 |  | 9.87 | 3.28 | 2.08 | 2.98 |
| Crude HR (95%CI) |  |  | 7.09 (5.80 - 8.67) | 3.35 (2.55 - 4.40) | Reference |  |  | 4.30 (3.82 - 4.83) | 0.89 (0.75 - 1.05) | Reference |  | 14.02 (8.58 - 22.91) | 1.79 (1.60 - 2.00) | 0.78 (0.68 - 0.91) | Reference |  | 4.57 (3.89 - 5.37) | 1.12 (0.98 - 1.27) | 0.72 (0.61 - 0.83) | Reference |
| Age adjusted HR (95% CI) |  |  | 0.91 (0.73 - 1.13) | 0.97 (0.74 - 1.28) | Reference |  |  | 0.76 (0.68 - 0.86) | 0.58 (0.49 - 0.68) | Reference |  | 1.09 (0.66 - 1.79) | 0.75 (0.67 - 0.84) | 0.59 (0.51 - 0.68) | Reference |  | 0.74 (0.64 - 0.86) | 0.70 (0.61 - 0.79) | 0.66 (0.57 - 0.78) | Reference |
| Age and gender adjusted HR (95% CI) |  |  | 0.95 (0.76 - 1.18) | 0.97 (0.74 - 1.28) | Reference |  |  | 0.77 (0.68 - 0.87) | 0.57 (0.48 - 0.67) | Reference |  | 1.12 (0.68 - 1.84) | 0.74 (0.67 - 0.83) | 0.58 (0.50 - 0.67) | Reference |  | 0.74 (0.64 - 0.86) | 0.69 (0.61 - 0.79) | 0.65 (0.56 - 0.76) | Reference |
| Age, gender and nursing home adjusted HR (95% CI) |  |  | 0.54 (0.43 - 0.69) | 0.80 (0.61 - 1.06) | Reference |  |  | 0.60 (0.53 - 0.68) | 0.58 (0.49 - 0.68) | Reference |  | 0.77 (0.46 - 1.28) | 0.64 (0.57 - 0.72) | 0.60 (0.52 - 0.69) | Reference |  | 0.56 (0.48 - 0.66) | 0.65 (0.58 - 0.74) | 0.69 (0.59 - 0.81) | Reference |
|  | 2022 | | | | | | | | | | | | | | | | | | | |
|  | first quarter | | | | | second quarter | | | | | third quarter | | | | | fourth quarter | | | | |
|  | Four or more vaccine doses | Three vaccine doses | Two vaccine doses | One vaccine dose | Unvaccinated | Four or more vaccine doses | Three vaccine doses | Two vaccine doses | One vaccine dose | Unvaccinated | Four or more vaccine doses | Three vaccine doses | Two vaccine doses | One vaccine dose | Unvaccinated | Four or more vaccine doses | Three vaccine doses | Two vaccine doses | One vaccine dose | Unvaccinated |
|  | All-cause mortality | | | | | All-cause mortality | | | | | All-cause mortality | | | | | All-cause mortality | | | | |
| Deaths (n) | 6 | 815 | 655 | 164 | 337 | 38 | 1700 | 809 | 135 | 533 | 649 | 3182 | 920 | 176 | 779 | 2333 | 3212 | 880 | 167 | 914 |
| Events per 100,000 person days | 47.04 | 4.99 | 2.91 | 1.78 | 1.85 | 18.16 | 3.12 | 2.01 | 1.35 | 1.21 | 11.54 | 2.73 | 1.7 | 1.63 | 1.25 | 7.42 | 2.57 | 1.58 | 1.51 | 1.37 |
| Crude HR (95%CI) | 27.97 (12.47 - 62.74) | 2.64 (2.33 - 3.00) | 1.56 (1.37 - 1.78) | 0.90 (0.75 - 1.09) | Reference | 16.08 (11.50 - 22.48) | 2.69 (2.44 - 2.97) | 1.67 (1.50 - 1.86) | 1.11 (0.92 - 1.34) | Reference | 9.72 (8.73 - 10.84) | 2.20 (2.03 - 2.38) | 1.36 (1.24 - 1.50) | 1.31 (1.11 - 1.54) | Reference | 5.46 (5.05 - 5.90) | 1.89 (1.76 - 2.03) | 1.16 (1.05 - 1.27) | 1.10 (0.94 - 1.30) | Reference |
| Age adjusted HR (95% CI) | 3.47 (1.55 - 7.81) | 0.78 (0.68 - 0.89) | 0.84 (0.74 - 0.96) | 0.92 (0.76 - 1.11) | Reference | 1.03 (0.73 - 1.45) | 0.91 (0.82 - 1.00) | 1.15 (1.03 - 1.28) | 1.08 (0.90 - 1.31) | Reference | 0.89 (0.80 - 1.00) | 0.98 (0.91 - 1.06) | 1.28 (1.16 - 1.41) | 1.40 (1.19 - 1.65) | Reference | 0.89 (0.82 - 0.97) | 1.15 (1.07 - 1.24) | 1.29 (1.18 - 1.42) | 1.29 (1.09 - 1.52) | Reference |
| Age and gender adjusted HR (95% CI) | 3.38 (1.50 - 7.61) | 0.77 (0.67 - 0.87) | 0.83 (0.73 - 0.94) | 0.90 (0.75 - 1.09) | Reference | 1.01 (0.72 - 1.42) | 0.89 (0.80 - 0.98) | 1.12 (1.01 - 1.25) | 1.08 (0.89 - 1.30) | Reference | 0.88 (0.78 - 0.98) | 0.96 (0.89 - 1.04) | 1.26 (1.15 - 1.39) | 1.39 (1.18 - 1.64) | Reference | 0.88 (0.81 - 0.95) | 1.13 (1.05 - 1.22) | 1.28 (1.17 - 1.40) | 1.28 (1.09 - 1.51) | Reference |
| Age, gender and nursing home adjusted HR (95% CI) | 2.54 (1.12 - 5.73) | 0.65 (0.57 - 0.74) | 0.84 (0.73 - 0.96) | 0.96 (0.79 - 1.15) | Reference | 0.73 (0.52 - 1.04) | 0.77 (0.70 - 0.85) | 1.08 (0.96 - 1.20) | 1.06 (0.88 - 1.29) | Reference | 0.67 (0.60 - 0.76) | 0.88 (0.81 - 0.95) | 1.20 (1.09 - 1.32) | 1.35 (1.14 - 1.59) | Reference | 0.76 (0.70 - 0.82) | 1.08 (1.00 - 1.16) | 1.24 (1.13 - 1.36) | 1.26 (1.07 - 1.49) | Reference |

| **Table S5 continued:** | | | | | | | | | | | | | | | | | | | | |
| --- | --- | --- | --- | --- | --- | --- | --- | --- | --- | --- | --- | --- | --- | --- | --- | --- | --- | --- | --- | --- |
|  | 2022 | | | | | | | | | | | | | | | | | | | |
|  | first quarter | | | | | second quarter | | | | | third quarter | | | | | fourth quarter | | | | |
|  | Four or more vaccine doses | Three vaccine doses | Two vaccine doses | One vaccine dose | Unvaccinated | Four or more vaccine doses | Three vaccine doses | Two vaccine doses | One vaccine dose | Unvaccinated | Four or more vaccine doses | Three vaccine doses | Two vaccine doses | One vaccine dose | Unvaccinated | Four or more vaccine doses | Three vaccine doses | Two vaccine doses | One vaccine dose | Unvaccinated |
|  | Non-COVID-19 mortality | | | | | Non-COVID-19 mortality | | | | | Non-COVID-19 mortality | | | | | Non-COVID-19 mortality | | | | |
| Deaths (n) | 4 | 776 | 618 | 160 | 298 | 37 | 1660 | 777 | 130 | 517 | 639 | 3111 | 894 | 170 | 760 | 2283 | 3131 | 864 | 165 | 892 |
| Events per 100,000 person days | 31.36 | 4.75 | 2.75 | 1.74 | 1.64 | 17.68 | 3.04 | 1.93 | 1.3 | 1.17 | 11.36 | 2.67 | 1.65 | 1.58 | 1.22 | 7.26 | 2.51 | 1.55 | 1.5 | 1.34 |
| Crude HR (95%CI) | 21.34 (7.95 - 57.26) | 2.84 (2.48 - 3.24) | 1.66 (1.45 - 1.91) | 0.98 (0.81 - 1.20) | Reference | 16.02 (11.41 - 22.49) | 2.70 (2.44 - 2.98) | 1.66 (1.48 - 1.85) | 1.11 (0.91 - 1.34) | Reference | 9.81 (8.79 - 10.94) | 2.20 (2.04 - 2.39) | 1.36 (1.23 - 1.49) | 1.29 (1.10 - 1.53) | Reference | 5.47 (5.05 - 5.91) | 1.89 (1.75 - 2.03) | 1.16 (1.06 - 1.28) | 1.12 (0.95 - 1.32) | Reference |
| Age adjusted HR (95% CI) | 2.67 (0.99 - 7.18) | 0.84 (0.73 - 0.96) | 0.90 (0.78 - 1.03) | 1.00 (0.82 - 1.21) | Reference | 1.03 (0.73 - 1.46) | 0.92 (0.83 - 1.01) | 1.14 (1.02 - 1.27) | 1.08 (0.89 - 1.31) | Reference | 0.90 (0.81 - 1.01) | 0.99 (0.91 - 1.07) | 1.27 (1.16 - 1.40) | 1.39 (1.17 - 1.64) | Reference | 0.89 (0.83 - 0.97) | 1.15 (1.07 - 1.24) | 1.30 (1.19 - 1.43) | 1.31 (1.11 - 1.54) | Reference |
| Age and gender adjusted HR (95% CI) | 2.60 (0.97 - 7.00) | 0.83 (0.72 - 0.95) | 0.88 (0.77 - 1.02) | 0.99 (0.81 - 1.20) | Reference | 1.02 (0.72 - 1.44) | 0.90 (0.81 - 0.99) | 1.12 (1.00 - 1.25) | 1.07 (0.89 - 1.30) | Reference | 0.89 (0.79 - 1.00) | 0.96 (0.89 - 1.04) | 1.26 (1.14 - 1.39) | 1.38 (1.17 - 1.63) | Reference | 0.88 (0.81 - 0.95) | 1.13 (1.05 - 1.22) | 1.29 (1.17 - 1.41) | 1.30 (1.10 - 1.53) | Reference |
| Age, gender and nursing home adjusted HR (95% CI) | 1.98 (0.74 - 5.36) | 0.70 (0.61 - 0.81) | 0.89 (0.78 - 1.03) | 1.04 (0.86 - 1.27) | Reference | 0.73 (0.52 - 1.04) | 0.78 (0.70 - 0.86) | 1.07 (0.96 - 1.20) | 1.06 (0.88 - 1.29) | Reference | 0.68 (0.60 - 0.77) | 0.88 (0.82 - 0.96) | 1.19 (1.08 - 1.32) | 1.33 (1.13 - 1.58) | Reference | 0.76 (0.70 - 0.83) | 1.08 (1.00 - 1.17) | 1.25 (1.14 - 1.37) | 1.28 (1.08 - 1.51) | Reference |
|  | 2023 | | | | | | | | | | | | | | | | | | | |
|  | first quarter | | | | | second quarter | | | | | third quarter | | | | | fourth quarter | | | | |
|  | Four or more vaccine doses | Three vaccine doses | Two vaccine doses | One vaccine dose | Unvaccinated | Four or more vaccine doses | Three vaccine doses | Two vaccine doses | One vaccine dose | Unvaccinated | Four or more vaccine doses | Three vaccine doses | Two vaccine doses | One vaccine dose | Unvaccinated | Four or more vaccine doses | Three vaccine doses | Two vaccine doses | One vaccine dose | Unvaccinated |
|  | All-cause mortality | | | | | All-cause mortality | | | | | All-cause mortality | | | | | All-cause mortality | | | | |
| Deaths (n) | 3185 | 3062 | 832 | 135 | 1049 | 3367 | 2772 | 799 | 165 | 1011 | 3493 | 2826 | 770 | 149 | 986 | 3951 | 3084 | 772 | 152 | 1041 |
| Events per 100,000 person days | 6.66 | 2.36 | 1.48 | 1.24 | 1.56 | 6.18 | 2 | 1.37 | 1.47 | 1.45 | 6.05 | 1.98 | 1.29 | 1.31 | 1.39 | 6.86 | 2.16 | 1.28 | 1.33 | 1.46 |
| Crude HR (95%CI) | 4.28 (3.99 - 4.59) | 1.52 (1.42 - 1.63) | 0.96 (0.87 - 1.05) | 0.80 (0.67 - 0.95) | Reference | 4.27 (3.98 - 4.58) | 1.38 (1.29 - 1.49) | 0.95 (0.86 - 1.04) | 1.02 (0.86 - 1.20) | Reference | 4.37 (4.07 - 4.69) | 1.43 (1.33 - 1.53) | 0.93 (0.85 - 1.02) | 0.94 (0.80 - 1.12) | Reference | 4.70 (4.39 - 5.03) | 1.48 (1.38 - 1.58) | 0.88 (0.80 - 0.97) | 0.91 (0.77 - 1.08) | Reference |
| Age adjusted HR (95% CI) | 0.85 (0.79 - 0.91) | 0.99 (0.92 - 1.06) | 1.14 (1.04 - 1.25) | 0.97 (0.81 - 1.16) | Reference | 0.89 (0.83 - 0.95) | 0.92 (0.85 - 0.99) | 1.15 (1.04 - 1.26) | 1.27 (1.08 - 1.50) | Reference | 0.95 (0.89 - 1.02) | 0.96 (0.89 - 1.03) | 1.13 (1.03 - 1.25) | 1.19 (1.00 - 1.41) | Reference | 0.97 (0.90 - 1.04) | 0.98 (0.91 - 1.05) | 1.09 (0.99 - 1.19) | 1.15 (0.97 - 1.37) | Reference |
| Age and gender adjusted HR (95% CI) | 0.82 (0.77 - 0.88) | 0.97 (0.90 - 1.04) | 1.12 (1.03 - 1.23) | 0.96 (0.81 - 1.15) | Reference | 0.86 (0.80 - 0.92) | 0.90 (0.84 - 0.97) | 1.13 (1.03 - 1.24) | 1.26 (1.07 - 1.48) | Reference | 0.93 (0.86 - 1.00) | 0.94 (0.87 - 1.01) | 1.12 (1.02 - 1.23) | 1.18 (1.00 - 1.40) | Reference | 0.94 (0.88 - 1.01) | 0.96 (0.89 - 1.03) | 1.07 (0.98 - 1.18) | 1.15 (0.97 - 1.36) | Reference |
| Age, gender and nursing home adjusted HR (95% CI) | 0.74 (0.69 - 0.80) | 0.94 (0.88 - 1.01) | 1.09 (1.00 - 1.20) | 0.95 (0.79 - 1.13) | Reference | 0.80 (0.74 - 0.86) | 0.88 (0.82 - 0.95) | 1.11 (1.01 - 1.22) | 1.24 (1.05 - 1.46) | Reference | 0.88 (0.82 - 0.95) | 0.92 (0.86 - 0.99) | 1.09 (0.99 - 1.20) | 1.16 (0.98 - 1.38) | Reference | 0.89 (0.83 - 0.95) | 0.95 (0.88 - 1.02) | 1.05 (0.96 - 1.15) | 1.13 (0.95 - 1.34) | Reference |
|  | Non-COVID-19 mortality | | | | | Non-COVID-19 mortality | | | | | Non-COVID-19 mortality | | | | | Non-COVID-19 mortality | | | | |
| Deaths (n) | 3087 | 2997 | 813 | 135 | 1021 | 3334 | 2747 | 790 | 163 | 1001 | 3482 | 2808 | 767 | 147 | 980 | 3823 | 2981 | 749 | 149 | 1014 |
| Events per 100,000 person days | 6.46 | 2.31 | 1.45 | 1.24 | 1.52 | 6.12 | 1.98 | 1.36 | 1.45 | 1.44 | 6.03 | 1.97 | 1.29 | 1.29 | 1.38 | 6.64 | 2.08 | 1.24 | 1.3 | 1.42 |
| Crude HR (95%CI) | 4.26 (3.97 - 4.58) | 1.53 (1.42 - 1.64) | 0.96 (0.88 - 1.05) | 0.82 (0.68 - 0.98) | Reference | 4.27 (3.97 - 4.58) | 1.38 (1.29 - 1.49) | 0.95 (0.86 - 1.04) | 1.02 (0.86 - 1.20) | Reference | 4.38 (4.08 - 4.70) | 1.43 (1.33 - 1.53) | 0.93 (0.85 - 1.03) | 0.94 (0.79 - 1.12) | Reference | 4.67 (4.36 - 5.01) | 1.47 (1.37 - 1.57) | 0.88 (0.80 - 0.97) | 0.92 (0.77 - 1.09) | Reference |
| Age adjusted HR (95% CI) | 0.85 (0.79 - 0.91) | 1.00 (0.93 - 1.07) | 1.14 (1.04 - 1.25) | 1.00 (0.83 - 1.19) | Reference | 0.89 (0.83 - 0.96) | 0.92 (0.85 - 0.99) | 1.14 (1.04 - 1.26) | 1.27 (1.07 - 1.49) | Reference | 0.96 (0.89 - 1.03) | 0.96 (0.89 - 1.03) | 1.14 (1.03 - 1.25) | 1.18 (0.99 - 1.40) | Reference | 0.97 (0.90 - 1.04) | 0.97 (0.91 - 1.04) | 1.08 (0.98 - 1.19) | 1.16 (0.98 - 1.38) | Reference |
| Age and gender adjusted HR (95% CI) | 0.82 (0.77 - 0.88) | 0.98 (0.91 - 1.05) | 1.13 (1.03 - 1.24) | 0.99 (0.83 - 1.19) | Reference | 0.86 (0.80 - 0.92) | 0.90 (0.84 - 0.97) | 1.13 (1.03 - 1.24) | 1.26 (1.06 - 1.48) | Reference | 0.93 (0.87 - 1.01) | 0.94 (0.87 - 1.01) | 1.12 (1.02 - 1.24) | 1.17 (0.99 - 1.40) | Reference | 0.94 (0.87 - 1.01) | 0.95 (0.89 - 1.02) | 1.07 (0.97 - 1.17) | 1.15 (0.97 - 1.37) | Reference |
| Age, gender and nursing home adjusted HR (95% CI) | 0.74 (0.69 - 0.80) | 0.95 (0.89 - 1.02) | 1.10 (1.00 - 1.21) | 0.97 (0.81 - 1.17) | Reference | 0.80 (0.74 - 0.86) | 0.89 (0.82 - 0.95) | 1.11 (1.01 - 1.21) | 1.24 (1.05 - 1.46) | Reference | 0.89 (0.82 - 0.96) | 0.92 (0.86 - 0.99) | 1.10 (1.00 - 1.20) | 1.15 (0.97 - 1.37) | Reference | 0.89 (0.82 - 0.95) | 0.94 (0.88 - 1.01) | 1.05 (0.95 - 1.15) | 1.14 (0.96 - 1.35) | Reference |

### **Analyses of periods with low and high COVID-19 mortality rates**

| **Table S6**: Characteristics of population for 2021 low COVID-19 disease burden on June 1. 2021 | | | | | | | | |
| --- | --- | --- | --- | --- | --- | --- | --- | --- |
|  | All | Male | Female | Unvaccinated | 1 dose | 2 doses | 3 doses | 4 or more doses |
| No. of participants | 401898 | 193552 | 208346 | 240541 | 105713 | 55643 | 1 | 0 |
| Age, y (median, IQR) | 45 (31.5-58.5) | 45 (31.5-58.5) | 46 (33-59) | 40 (27.5-52.5) | 51 (39.5-62.5) | 57 (40-74) | 52 (52-52) |  |
| Nursing home residency (%) | 3.3 | 1.67 | 4.81 | 1.19 | 1.34 | 16.14 | 0 |  |
| Repeated previous infections (%) | 0.04 | 0.03 | 0.04 | 0.03 | 0.03 | 0.08 | 0 |  |
| Last infection in 2020 (%) | 78.96 | 78.99 | 78.93 | 73.97 | 86.51 | 86.17 | 100 |  |
| Last infection in 2021 (%) | 21.04 | 21.01 | 21.07 | 26.03 | 13.49 | 13.83 | 0 |  |
| Last infection in 2022 (%) | 0 | 0 | 0 | 0 | 0 | 0 | 0 |  |
| mRNA (%) | 32.19 | 31.78 | 32.58 | 0 | 77.04 | 86.17 | 100 |  |
| Infected in the observation period (%) | 0.04 | 0.04 | 0.04 | 0.05 | 0.02 | 0.03 | 0 |  |

| **Table S7**: Characteristics of population for 2022 low COVID-19 disease burden on May 1. 2022 | | | | | | | | |
| --- | --- | --- | --- | --- | --- | --- | --- | --- |
|  | All | Male | Female | Unvaccinated | 1 dose | 2 doses | 3 doses | 4 or more doses |
| No. of participants | 1234779 | 611421 | 623358 | 383730 | 97561 | 380078 | 372752 | 658 |
| Age, y (median, IQR) | 41 (28.5-53.5) | 41 (28.5-53.5) | 41 (29-53) | 38 (27-49) | 37 (25-49) | 40 (27-53) | 47 (33.5-60.5) | 69 (56-82) |
| Nursing home residency (%) | 1.01 | 0.51 | 1.5 | 0.27 | 0.34 | 0.64 | 2.31 | 14.74 |
| Repeated previous infections (%) | 3.88 | 3.71 | 4.04 | 6.49 | 7.04 | 2.73 | 1.53 | 1.67 |
| Last infection in 2020 (%) | 18.57 | 18.64 | 18.5 | 5.79 | 20.03 | 21.06 | 28.76 | 41.95 |
| Last infection in 2021 (%) | 46.42 | 47.05 | 45.8 | 52.08 | 62.74 | 47.34 | 35.4 | 38.3 |
| Last infection in 2022 (%) | 35.01 | 34.3 | 35.71 | 42.13 | 17.23 | 31.61 | 35.84 | 19.76 |
| mRNA (%) | 66.17 | 66.25 | 66.1 | 0 | 86.04 | 94.74 | 99.91 | 98.78 |
| Infected in the observation period (%) | 2.34 | 2 | 2.68 | 2.63 | 2.75 | 2.08 | 2.2 | 1.98 |

| **Table S8**: Characteristics of population for 2023 low COVID-19 disease burden on July 1. 2023 | | | | | | | | |
| --- | --- | --- | --- | --- | --- | --- | --- | --- |
|  | All | Male | Female | Unvaccinated | 1 dose | 2 doses | 3 doses | 4 or more doses |
| No. of participants | 4213756 | 2001207 | 2212549 | 877984 | 140202 | 732517 | 1755912 | 707141 |
| Age, y (median, IQR) | 43 (30-56) | 42 (28.5-55.5) | 43 (30-56) | 39 (27.5-50.5) | 35 (23.5-46.5) | 36 (24.5-47.5) | 43 (30.5-55.5) | 58 (45-71) |
| Nursing home residency (%) | 0.87 | 0.49 | 1.22 | 0.28 | 0.28 | 0.29 | 0.56 | 3.08 |
| Repeated previous infections (%) | 16.02 | 14.3 | 17.58 | 23.01 | 33.64 | 18.95 | 12.81 | 8.79 |
| Last infection in 2020 (%) | 4.19 | 4.56 | 3.86 | 2.11 | 9.48 | 6.89 | 3.82 | 3.85 |
| Last infection in 2021 (%) | 11.15 | 12.14 | 10.26 | 19.24 | 31.61 | 15.05 | 6.2 | 5.31 |
| Last infection in 2022 (%) | 77.73 | 77.07 | 78.33 | 75.57 | 54.97 | 73.02 | 82.02 | 79.17 |
| mRNA (%) | 77.95 | 77.87 | 78.03 | 0 | 85.81 | 96.71 | 99.84 | 99.39 |
| Infected in the observation period (%) | 0 | 0 | 0 | 0 | 0 | 0 | 0 | 0 |

| **Table S9**: Characteristics of population for 2021 high COVID-19 disease burden on October 1. 2021 | | | | | | | | |
| --- | --- | --- | --- | --- | --- | --- | --- | --- |
|  | All | Male | Female | Unvaccinated | 1 dose | 2 doses | 3 doses | 4 or more doses |
| No. of participants | 547913 | 267625 | 280288 | 193095 | 153297 | 197032 | 4489 | 0 |
| Age, y (median, IQR) | 44 (30.5-57.5) | 44 (31-57) | 45 (32-58) | 39 (27-51) | 45 (32-58) | 48 (34.5-61.5) | 80 (68-92) |  |
| Nursing home residency (%) | 2.39 | 1.2 | 3.52 | 1.02 | 1.07 | 3.46 | 59.32 |  |
| Repeated previous infections (%) | 0.16 | 0.15 | 0.17 | 0.16 | 0.16 | 0.15 | 0.45 |  |
| Last infection in 2020 (%) | 57.77 | 57.03 | 58.46 | 47.7 | 53.82 | 70.26 | 77.41 |  |
| Last infection in 2021 (%) | 42.23 | 42.97 | 41.54 | 52.3 | 46.18 | 29.74 | 22.59 |  |
| Last infection in 2022 (%) | 0 | 0 | 0 | 0 | 0 | 0 | 0 |  |
| mRNA (%) | 53.22 | 53.35 | 53.1 | 0 | 74.88 | 87.47 | 99.67 |  |
| Infected in the observation period (%) | 1.03 | 0.96 | 1.1 | 2.2 | 0.33 | 0.46 | 0.2 |  |

| **Table S10**: Characteristics of population for 2022 high COVID-19 disease burden on February 1. 2022 | | | | | | | | |
| --- | --- | --- | --- | --- | --- | --- | --- | --- |
|  | All | Male | Female | Unvaccinated | 1 dose | 2 doses | 3 doses | 4 or more doses |
| No. of participants | 646379 | 317556 | 328823 | 138595 | 99631 | 227605 | 180447 | 101 |
| Age, y (median, IQR) | 43 (30-56) | 43 (29.5-56.5) | 44 (30.5-57.5) | 38 (26.5-49.5) | 38 (26-50) | 44 (30.5-57.5) | 50 (36.5-63.5) | 67 (54-80) |
| Nursing home residency (%) | 1.92 | 0.96 | 2.85 | 0.73 | 0.47 | 1.22 | 4.51 | 13.86 |
| Repeated previous infections (%) | 0.45 | 0.41 | 0.48 | 0.91 | 0.52 | 0.31 | 0.23 | 0 |
| Last infection in 2020 (%) | 44.78 | 44.15 | 45.38 | 26.25 | 33.1 | 45.11 | 65.02 | 68.32 |
| Last infection in 2021 (%) | 55.22 | 55.85 | 54.62 | 73.75 | 66.9 | 54.89 | 34.98 | 31.68 |
| Last infection in 2022 (%) | 0 | 0 | 0 | 0 | 0 | 0 | 0 | 0 |
| mRNA (%) | 76.21 | 76.46 | 75.97 | 0 | 94.17 | 95.9 | 99.97 | 98.02 |
| Infected in the observation period (%) | 17.8 | 16.25 | 19.3 | 29.21 | 18.02 | 13.65 | 14.17 | 13.86 |

| **Table S11**: Characteristics of population for 2023 high COVID-19 disease burden on February 1. 2023 | | | | | | | | |
| --- | --- | --- | --- | --- | --- | --- | --- | --- |
|  | All | Male | Female | Unvaccinated | 1 dose | 2 doses | 3 doses | 4 or more doses |
| No. of participants | 3812211 | 1822960 | 1989251 | 833460 | 133393 | 684205 | 1577022 | 584131 |
| Age, y (median, IQR) | 42 (29-55) | 42 (29-55) | 43 (30-56) | 39 (27.5-50.5) | 35 (23.5-46.5) | 36 (25-47) | 43 (30.5-55.5) | 58 (45-71) |
| Nursing home residency (%) | 0.85 | 0.48 | 1.19 | 0.27 | 0.28 | 0.29 | 0.57 | 3.24 |
| Repeated previous infections (%) | 12.17 | 11.02 | 13.23 | 20.72 | 30.36 | 14.68 | 7.88 | 4.47 |
| Last infection in 2020 (%) | 4.84 | 5.19 | 4.51 | 2.26 | 10.33 | 7.68 | 4.47 | 4.92 |
| Last infection in 2021 (%) | 12.65 | 13.63 | 11.77 | 20.4 | 33.82 | 16.63 | 7.28 | 6.63 |
| Last infection in 2022 (%) | 82.51 | 81.18 | 83.72 | 77.34 | 55.85 | 75.69 | 88.25 | 88.45 |
| mRNA (%) | 76.94 | 76.91 | 76.97 | 0 | 85.76 | 96.71 | 99.85 | 99.69 |
| Infected in the observation period (%) | 2.92 | 2.45 | 3.34 | 1.4 | 2.45 | 2.82 | 3.53 | 3.66 |

| **Table S12:** Hazard ratios (HR) with 95% confidence intervals (95% CI) for non-COVID 19 and all-cause mortality according to number of SARS-CoV-2 vaccine doses during different time periods of high and low COVID-19 disease burden. | | | | | | | | | | |
| --- | --- | --- | --- | --- | --- | --- | --- | --- | --- | --- |
|  | June and July 2021 (low COVID-19 disease burden) | | | | | October and November 2021 (high COVID-19 disease burden) | | | | |
|  | Four or more vaccine doses | Three vaccine doses | Two vaccine doses | One vaccine dose | Unvaccinated | Four or more vaccine doses | Three vaccine doses | Two vaccine doses | One vaccine dose | Unvaccinated |
|  | All-cause mortality | | | | | All-cause mortality | | | | |
| Deaths (n) | 0 | 0 | 379 | 141 | 376 | 0 | 191 | 400 | 197 | 289 |
| Events per 100,000 person days |  |  | 6.68 | 1.94 | 3.26 |  | 16.25 | 3.22 | 2 | 2.90 |
| Crude HR (95%CI) |  |  | 2.13 (1.84 - 2.47) | 0.59 (0.48 - 0.71) | Reference |  | 6.16 (5.05 - 7.51) | 1.10 (0.95 - 1.29) | 0.70 (0.58 - 0.83) | Reference |
| Age adjusted HR (95% CI) |  |  | 0.66 (0.57 - 0.77) | 0.52 (0.43 - 0.63) | Reference |  | 0.73 (0.60 - 0.89) | 0.67 (0.58 - 0.78) | 0.63 (0.53 - 0.76) | Reference |
| Age and gender adjusted HR (95% CI) |  |  | 0.66 (0.57 - 0.77) | 0.51 (0.42 - 0.62) | Reference |  | 0.73 (0.60 - 0.89) | 0.66 (0.57 - 0.77) | 0.62 (0.51 - 0.74) | Reference |
| Age, gender and nursing home adjusted HR (95% CI) |  |  | 0.55 (0.47 - 0.64) | 0.52 (0.43 - 0.63) | Reference |  | 0.56 (0.46 - 0.70) | 0.62 (0.53 - 0.72) | 0.65 (0.54 - 0.78) | Reference |
| Age, gender, nursing home and year of last infection adjusted HR (95% CI) |  |  | 0.57 (0.49 - 0.66) | 0.52 (0.43 - 0.64) | Reference |  | 0.58 (0.47 - 0.71) | 0.63 (0.54 - 0.73) | 0.64 (0.54 - 0.77) | Reference |
|  | Non-COVID-19 mortality | | | | | Non-COVID-19 mortality | | | | |
| Deaths (n) | 0 | 0 | 369 | 139 | 367 | 0 | 189 | 390 | 193 | 276 |
| Events per 100.000 person days |  |  | 6.51 | 1.91 | 3.18 |  | 16.08 | 3.14 | 1.96 | 2.77 |
| Crude HR (95%CI) |  |  | 2.13 (1.84 - 2.47) | 0.59 (0.49 - 0.72) | Reference |  | 6.48 (5.30 - 7.93) | 1.13 (0.97 - 1.32) | 0.72 (0.60 - 0.86) | Reference |
| Age adjusted HR (95% CI) |  |  | 0.66 (0.57 - 0.77) | 0.53 (0.43 - 0.64) | Reference |  | 0.76 (0.62 - 0.93) | 0.69 (0.59 - 0.80) | 0.65 (0.54 - 0.78) | Reference |
| Age and gender adjusted HR (95% CI) |  |  | 0.66 (0.57 - 0.77) | 0.51 (0.42 - 0.62) | Reference |  | 0.76 (0.62 - 0.93) | 0.68 (0.58 - 0.79) | 0.63 (0.53 - 0.76) | Reference |
| Age, gender and nursing home adjusted HR (95% CI) |  |  | 0.55 (0.47 - 0.64) | 0.52 (0.43 - 0.64) | Reference |  | 0.58 (0.47 - 0.72) | 0.63 (0.54 - 0.74) | 0.67 (0.55 - 0.80) | Reference |
| Age, gender, nursing home and year of last infection adjusted HR (95% CI) |  |  | 0.56 (0.48 - 0.66) | 0.53 (0.43 - 0.64) | Reference |  | 0.59 (0.48 - 0.73) | 0.64 (0.55 - 0.75) | 0.66 (0.55 - 0.80) | Reference |
|  | May and June 2022 (low COVID-19 disease burden) | | | | | February and March 2022 (high COVID-19 disease burden) | | | | |
|  | Four or more vaccine doses | Three vaccine doses | Two vaccine doses | One vaccine dose | Unvaccinated | Four or more vaccine doses | Three vaccine doses | Two vaccine doses | One vaccine dose | Unvaccinated |
|  | All-cause mortality | | | | | All-cause mortality | | | | |
| Deaths (n) | 22 | 659 | 387 | 62 | 230 | 5 | 472 | 356 | 76 | 165 |
| Events per 100,000 person days | 18.95 | 2.76 | 1.74 | 1.09 | 0.99 | 50.65 | 4.01 | 2.73 | 1.44 | 2.07 |
| Crude HR (95%CI) | 18.47 (11.87 - 28.74) | 2.80 (2.41 - 3.25) | 1.77 (1.50 - 2.08) | 1.11 (0.84 - 1.47) | Reference | 23.99 (9.85 - 58.45) | 1.93 (1.62 - 2.30) | 1.32 (1.10 - 1.59) | 0.70 (0.53 - 0.91) | Reference |
| Age adjusted HR (95% CI) | 0.98 (0.62 - 1.54) | 0.90 (0.78 - 1.05) | 1.11 (0.94 - 1.30) | 1.01 (0.76 - 1.34) | Reference | 3.63 (1.49 - 8.86) | 0.71 (0.60 - 0.85) | 0.82 (0.68 - 0.98) | 0.80 (0.61 - 1.06) | Reference |
| Age and gender adjusted HR (95% CI) | 0.98 (0.62 - 1.54) | 0.89 (0.76 - 1.03) | 1.09 (0.92 - 1.28) | 1.01 (0.76 - 1.34) | Reference | 3.57 (1.46 - 8.70) | 0.70 (0.59 - 0.84) | 0.80 (0.67 - 0.96) | 0.79 (0.60 - 1.03) | Reference |
| Age, gender and nursing home adjusted HR (95% CI) | 0.67 (0.42 - 1.08) | 0.77 (0.66 - 0.91) | 1.06 (0.90 - 1.25) | 1.00 (0.76 - 1.33) | Reference | 2.65 (1.08 - 6.51) | 0.60 (0.50 - 0.71) | 0.81 (0.67 - 0.98) | 0.85 (0.64 - 1.12) | Reference |
| Age, gender, nursing home and year of last infection adjusted HR (95% CI) | 0.66 (0.41 - 1.06) | 0.78 (0.67 - 0.91) | 1.04 (0.88 - 1.23) | 1.00 (0.75 - 1.32) | Reference | 2.64 (1.08 - 6.50) | 0.60 (0.50 - 0.72) | 0.81 (0.67 - 0.98) | 0.85 (0.65 - 1.12) | Reference |
|  | Non-COVID-19 mortality | | | | | Non-COVID-19 mortality | | | | |
| Deaths (n) | 22 | 649 | 382 | 61 | 225 | 3 | 445 | 339 | 74 | 150 |
| Events per 100.000 person days | 18.95 | 2.72 | 1.72 | 1.07 | 0.96 | 30.39 | 3.78 | 2.6 | 1.4 | 1.88 |
| Crude HR (95%CI) | 18.90 (12.14 - 29.43) | 2.82 (2.42 - 3.28) | 1.78 (1.51 - 2.10) | 1.11 (0.84 - 1.48) | Reference | 15.93 (5.08 - 49.98) | 2.00 (1.67 - 2.41) | 1.38 (1.14 - 1.68) | 0.74 (0.56 - 0.98) | Reference |
| Age adjusted HR (95% CI) | 1.01 (0.64 - 1.60) | 0.91 (0.78 - 1.06) | 1.12 (0.95 - 1.32) | 1.02 (0.77 - 1.35) | Reference | 2.41 (0.77 - 7.56) | 0.74 (0.62 - 0.90) | 0.85 (0.71 - 1.04) | 0.86 (0.65 - 1.14) | Reference |
| Age and gender adjusted HR (95% CI) | 1.01 (0.64 - 1.59) | 0.90 (0.77 - 1.05) | 1.10 (0.93 - 1.30) | 1.02 (0.77 - 1.35) | Reference | 2.37 (0.75 - 7.43) | 0.73 (0.61 - 0.88) | 0.84 (0.69 - 1.02) | 0.84 (0.64 - 1.11) | Reference |
| Age, gender and nursing home adjusted HR (95% CI) | 0.69 (0.43 - 1.10) | 0.78 (0.67 - 0.92) | 1.07 (0.91 - 1.26) | 1.01 (0.76 - 1.34) | Reference | 1.74 (0.55 - 5.48) | 0.62 (0.52 - 0.75) | 0.85 (0.70 - 1.03) | 0.91 (0.69 - 1.21) | Reference |
| Age, gender, nursing home and year of last infection adjusted HR (95% CI) | 0.67 (0.42 - 1.08) | 0.79 (0.67 - 0.93) | 1.05 (0.89 - 1.25) | 1.00 (0.75 - 1.34) | Reference | 1.73 (0.55 - 5.47) | 0.62 (0.51 - 0.76) | 0.85 (0.70 - 1.03) | 0.91 (0.69 - 1.21) | Reference |

| **Table S12 continued:** | | | | | | | | | | |
| --- | --- | --- | --- | --- | --- | --- | --- | --- | --- | --- |
|  | July and August 2023 (low COVID-19 disease burden) | | | | | February and March 2023 (high COVID-19 disease burden) | | | | |
|  | Four or more vaccine doses | Three vaccine doses | Two vaccine doses | One vaccine dose | Unvaccinated | Four or more vaccine doses | Three vaccine doses | Two vaccine doses | One vaccine dose | Unvaccinated |
|  | All-cause mortality | | | | | All-cause mortality | | | | |
| Deaths (n) | 2230 | 1870 | 510 | 101 | 627 | 1860 | 1692 | 452 | 82 | 586 |
| Events per 100,000 person days | 5.1 | 1.72 | 1.12 | 1.16 | 1.15 | 5.33 | 1.83 | 1.12 | 1.04 | 1.19 |
| Crude HR (95%CI) | 4.43 (4.05 - 4.84) | 1.49 (1.36 - 1.63) | 0.97 (0.87 - 1.10) | 1.01 (0.82 - 1.24) | Reference | 4.47 (4.07 - 4.90) | 1.53 (1.40 - 1.69) | 0.94 (0.83 - 1.06) | 0.87 (0.69 - 1.10) | Reference |
| Age adjusted HR (95% CI) | 0.96 (0.88 - 1.05) | 1.00 (0.91 - 1.09) | 1.18 (1.05 - 1.33) | 1.26 (1.03 - 1.56) | Reference | 0.90 (0.82 - 0.99) | 0.99 (0.91 - 1.09) | 1.11 (0.99 - 1.26) | 1.07 (0.85 - 1.34) | Reference |
| Age and gender adjusted HR (95% CI) | 0.94 (0.86 - 1.03) | 0.98 (0.89 - 1.07) | 1.17 (1.04 - 1.31) | 1.26 (1.02 - 1.55) | Reference | 0.87 (0.80 - 0.96) | 0.97 (0.89 - 1.07) | 1.10 (0.97 - 1.25) | 1.06 (0.84 - 1.33) | Reference |
| Age, gender and nursing home adjusted HR (95% CI) | 0.89 (0.81 - 0.97) | 0.96 (0.88 - 1.05) | 1.14 (1.01 - 1.28) | 1.23 (1.00 - 1.52) | Reference | 0.80 (0.72 - 0.88) | 0.95 (0.86 - 1.04) | 1.07 (0.95 - 1.21) | 1.05 (0.83 - 1.32) | Reference |
| Age, gender, nursing home and year of last infection adjusted HR (95% CI) | 0.87 (0.80 - 0.96) | 0.95 (0.87 - 1.04) | 1.17 (1.04 - 1.32) | 1.30 (1.05 - 1.61) | Reference | 0.80 (0.72 - 0.88) | 0.95 (0.86 - 1.04) | 1.08 (0.96 - 1.23) | 1.06 (0.84 - 1.34) | Reference |
|  | Non-COVID-19 mortality | | | | | Non-COVID-19 mortality | | | | |
| Deaths (n) | 2227 | 1863 | 510 | 100 | 623 | 1804 | 1661 | 444 | 82 | 570 |
| Events per 100.000 person days | 5.1 | 1.71 | 1.12 | 1.15 | 1.14 | 5.17 | 1.8 | 1.1 | 1.04 | 1.16 |
| Crude HR (95%CI) | 4.45 (4.07 - 4.87) | 1.50 (1.37 - 1.64) | 0.98 (0.87 - 1.10) | 1.01 (0.81 - 1.24) | Reference | 4.46 (4.05 - 4.90) | 1.55 (1.41 - 1.70) | 0.95 (0.84 - 1.07) | 0.90 (0.71 - 1.13) | Reference |
| Age adjusted HR (95% CI) | 0.97 (0.88 - 1.06) | 1.00 (0.91 - 1.09) | 1.19 (1.06 - 1.34) | 1.26 (1.02 - 1.56) | Reference | 0.90 (0.82 - 0.99) | 1.00 (0.91 - 1.11) | 1.13 (0.99 - 1.27) | 1.10 (0.87 - 1.38) | Reference |
| Age and gender adjusted HR (95% CI) | 0.94 (0.86 - 1.03) | 0.98 (0.89 - 1.07) | 1.18 (1.05 - 1.32) | 1.25 (1.02 - 1.55) | Reference | 0.88 (0.80 - 0.97) | 0.98 (0.89 - 1.08) | 1.11 (0.98 - 1.26) | 1.09 (0.86 - 1.37) | Reference |
| Age, gender and nursing home adjusted HR (95% CI) | 0.89 (0.81 - 0.98) | 0.96 (0.88 - 1.06) | 1.14 (1.02 - 1.29) | 1.23 (1.00 - 1.52) | Reference | 0.80 (0.73 - 0.89) | 0.96 (0.87 - 1.06) | 1.08 (0.96 - 1.23) | 1.07 (0.85 - 1.35) | Reference |
| Age, gender, nursing home and year of last infection adjusted HR (95% CI) | 0.88 (0.80 - 0.97) | 0.95 (0.87 - 1.04) | 1.18 (1.05 - 1.33) | 1.29 (1.04 - 1.60) | Reference | 0.80 (0.73 - 0.89) | 0.96 (0.87 - 1.06) | 1.10 (0.97 - 1.24) | 1.09 (0.86 - 1.38) | Reference |

### **Sensitivity analyses**

| **Table S13:** Hazard ratios (HR) with 95% confidence intervals (95% CI) for non-COVID 19 and all-cause mortality according to number of SARS-CoV-2 vaccine doses for the period from 2021 to 2023 split into 3-month intervals for **males** only. | | | | | | | | | | | | | | | | | | | | |
| --- | --- | --- | --- | --- | --- | --- | --- | --- | --- | --- | --- | --- | --- | --- | --- | --- | --- | --- | --- | --- |
|  | 2021 | | | | | | | | | | | | | | | | | | | |
|  | first quarter | | | | | second quarter | | | | | third quarter | | | | | fourth quarter | | | | |
|  | Four or more vaccine doses | Three vaccine doses | Two vaccine doses | One vaccine dose | Unvaccinated | Four or more vaccine doses | Three vaccine doses | Two vaccine doses | One vaccine dose | Unvaccinated | Four or more vaccine doses | Three vaccine doses | Two vaccine doses | One vaccine dose | Unvaccinated | Four or more vaccine doses | Three vaccine doses | Two vaccine doses | One vaccine dose | Unvaccinated |
|  | All-cause mortality | | | | | All-cause mortality | | | | | All-cause mortality | | | | | All-cause mortality | | | | |
| Deaths (n) |  |  | 45 | 26 | 184 |  |  | 193 | 91 | 301 |  | 9 | 275 | 120 | 257 |  | 143 | 219 | 141 | 171 |
| Events per 100,000 person days | |  | 20.21 | 8.75 | 2.66 |  |  | 11.79 | 2.69 | 2.88 |  | 57.97 | 4.2 | 2.09 | 2.86 |  | 7.62 | 2.47 | 2.13 | 2.71 |
| Crude HR (95%CI) |  |  | 7.56 (5.42 - 10.54) | 3.51 (2.30 - 5.34) | Reference |  |  | 4.35 (3.60 - 5.24) | 0.95 (0.75 - 1.21) | Reference |  | 22.63 (11.02 - 46.45) | 1.51 (1.27 - 1.79) | 0.75 (0.60 - 0.93) | Reference |  | 3.93 (3.02 - 5.11) | 0.92 (0.75 - 1.13) | 0.79 (0.63 - 0.99) | Reference |
| Age adjusted HR (95% CI) |  |  | 1.10 (0.77 - 1.56) | 0.93 (0.60 - 1.42) | Reference |  |  | 0.78 (0.65 - 0.95) | 0.54 (0.42 - 0.68) | Reference |  | 2.17 (1.04 - 4.53) | 0.67 (0.56 - 0.79) | 0.51 (0.41 - 0.63) | Reference |  | 0.67 (0.52 - 0.86) | 0.51 (0.42 - 0.62) | 0.65 (0.52 - 0.81) | Reference |
| Age and nursing home adjusted HR (95% CI) |  |  | 0.56 (0.37 - 0.84) | 0.79 (0.51 - 1.21) | Reference |  |  | 0.60 (0.49 - 0.73) | 0.55 (0.43 - 0.70) | Reference |  | 1.33 (0.62 - 2.87) | 0.60 (0.50 - 0.71) | 0.52 (0.42 - 0.65) | Reference |  | 0.51 (0.39 - 0.66) | 0.50 (0.41 - 0.61) | 0.69 (0.55 - 0.86) | Reference |
|  | Non-COVID-19 mortality | | | | | Non-COVID-19 mortality | | | | | Non-COVID-19 mortality | | | | | Non-COVID-19 mortality | | | | |
| Deaths (n) |  |  | 45 | 22 | 168 |  |  | 180 | 88 | 287 |  | 9 | 269 | 117 | 241 |  | 139 | 214 | 135 | 156 |
| Events per 100,000 person days | |  | 20.21 | 7.41 | 2.43 |  |  | 11 | 2.6 | 2.75 |  | 57.97 | 4.11 | 2.04 | 2.68 |  | 7.4 | 2.42 | 2.04 | 2.47 |
| Crude HR (95%CI) |  |  | 8.26 (5.90 - 11.55) | 3.23 (2.05 - 5.10) | Reference |  |  | 4.18 (3.44 - 5.07) | 0.95 (0.74 - 1.22) | Reference |  | 24.77 (11.98 - 51.19) | 1.58 (1.32 - 1.88) | 0.78 (0.62 - 0.97) | Reference |  | 4.33 (3.30 - 5.69) | 0.99 (0.80 - 1.22) | 0.84 (0.66 - 1.06) | Reference |
| Age adjusted HR (95% CI) |  |  | 1.20 (0.84 - 1.73) | 0.85 (0.54 - 1.35) | Reference |  |  | 0.76 (0.63 - 0.93) | 0.54 (0.42 - 0.69) | Reference |  | 2.37 (1.13 - 4.98) | 0.70 (0.58 - 0.83) | 0.53 (0.43 - 0.66) | Reference |  | 0.72 (0.56 - 0.93) | 0.55 (0.44 - 0.67) | 0.68 (0.54 - 0.86) | Reference |
| Age and nursing home adjusted HR (95% CI) |  |  | 0.64 (0.42 - 0.97) | 0.73 (0.46 - 1.16) | Reference |  |  | 0.58 (0.47 - 0.71) | 0.55 (0.43 - 0.70) | Reference |  | 1.42 (0.66 - 3.08) | 0.62 (0.52 - 0.75) | 0.55 (0.44 - 0.68) | Reference |  | 0.53 (0.41 - 0.70) | 0.53 (0.43 - 0.66) | 0.73 (0.58 - 0.91) | Reference |
|  | 2022 | | | | | | | | | | | | | | | | | | | |
|  | first quarter | | | | | second quarter | | | | | third quarter | | | | | fourth quarter | | | | |
|  | Four or more vaccine doses | Three vaccine doses | Two vaccine doses | One vaccine dose | Unvaccinated | Four or more vaccine doses | Three vaccine doses | Two vaccine doses | One vaccine dose | Unvaccinated | Four or more vaccine doses | Three vaccine doses | Two vaccine doses | One vaccine dose | Unvaccinated | Four or more vaccine doses | Three vaccine doses | Two vaccine doses | One vaccine dose | Unvaccinated |
|  | All-cause mortality | | | | | All-cause mortality | | | | | All-cause mortality | | | | | All-cause mortality | | | | |
| Deaths (n) | 1 | 328 | 298 | 77 | 144 | 15 | 787 | 418 | 66 | 223 | 265 | 1546 | 445 | 80 | 332 | 1020 | 1567 | 438 | 71 | 404 |
| Events per 100,000 person days | 14.24 | 4.07 | 2.69 | 1.69 | 1.64 | 14.47 | 3 | 2.11 | 1.32 | 1.05 | 9.63 | 2.81 | 1.67 | 1.49 | 1.12 | 6.76 | 2.68 | 1.59 | 1.29 | 1.28 |
| Crude HR (95%CI) | 9.17 (1.28 - 65.55) | 2.46 (2.02 - 2.99) | 1.65 (1.35 - 2.02) | 1.01 (0.76 - 1.33) | Reference | 14.54 (8.55 - 24.73) | 2.91 (2.51 - 3.38) | 2.01 (1.71 - 2.37) | 1.25 (0.95 - 1.65) | Reference | 8.87 (7.50 - 10.49) | 2.53 (2.24 - 2.84) | 1.50 (1.30 - 1.73) | 1.33 (1.04 - 1.70) | Reference | 5.35 (4.76 - 6.01) | 2.11 (1.89 - 2.36) | 1.25 (1.09 - 1.43) | 1.01 (0.79 - 1.30) | Reference |
| Age adjusted HR (95% CI) | 1.15 (0.16 - 8.24) | 0.71 (0.58 - 0.86) | 0.79 (0.65 - 0.97) | 0.94 (0.71 - 1.24) | Reference | 1.07 (0.62 - 1.85) | 0.93 (0.80 - 1.08) | 1.32 (1.12 - 1.56) | 1.24 (0.94 - 1.63) | Reference | 0.84 (0.69 - 1.00) | 1.00 (0.89 - 1.13) | 1.38 (1.19 - 1.59) | 1.47 (1.15 - 1.88) | Reference | 0.83 (0.73 - 0.94) | 1.12 (1.01 - 1.25) | 1.36 (1.19 - 1.56) | 1.19 (0.93 - 1.53) | Reference |
| Age and nursing home adjusted HR (95% CI) | 0.84 (0.12 - 6.02) | 0.63 (0.51 - 0.77) | 0.80 (0.65 - 0.97) | 0.98 (0.74 - 1.30) | Reference | 0.82 (0.47 - 1.44) | 0.85 (0.73 - 1.00) | 1.26 (1.07 - 1.49) | 1.22 (0.92 - 1.61) | Reference | 0.67 (0.55 - 0.82) | 0.96 (0.85 - 1.08) | 1.30 (1.12 - 1.49) | 1.42 (1.11 - 1.82) | Reference | 0.77 (0.68 - 0.88) | 1.09 (0.98 - 1.22) | 1.32 (1.15 - 1.51) | 1.18 (0.92 - 1.52) | Reference |

| **Table S13 continued:** | | | | | | | | | | | | | | | | | | | | |
| --- | --- | --- | --- | --- | --- | --- | --- | --- | --- | --- | --- | --- | --- | --- | --- | --- | --- | --- | --- | --- |
|  | 2022 | | | | | | | | | | | | | | | | | | | |
|  | first quarter | | | | | second quarter | | | | | third quarter | | | | | fourth quarter | | | | |
|  | Four or more vaccine doses | Three vaccine doses | Two vaccine doses | One vaccine dose | Unvacc | Four or more vaccine doses | Three vaccine doses | Two vaccine doses | One vaccine dose | Unvacc | Four or more vaccine doses | Three vaccine doses | Two vaccine doses | One vaccine dose | Unvacc | Four or more vaccine doses | Three vaccine doses | Two vaccine doses | One vaccine dose | Unvacc |
|  | Non-COVID-19 mortality | | | | | Non-COVID-19 mortality | | | | | Non-COVID-19 mortality | | | | | Non-COVID-19 mortality | | | | |
| Deaths (n) | 1 | 315 | 280 | 75 | 126 | 14 | 770 | 401 | 61 | 211 | 258 | 1512 | 437 | 78 | 322 | 1001 | 1525 | 429 | 70 | 395 |
| Events per 100,000 person days | 14.24 | 3.91 | 2.53 | 1.65 | 1.43 | 13.5 | 2.93 | 2.03 | 1.22 | 0.99 | 9.37 | 2.75 | 1.64 | 1.45 | 1.09 | 6.63 | 2.61 | 1.56 | 1.27 | 1.25 |
| Crude HR (95%CI) | 10.54 (1.47 - 75.45) | 2.69 (2.19 - 3.31) | 1.77 (1.44 - 2.19) | 1.11 (0.83 - 1.49) | Reference | 14.21 (8.21 - 24.62) | 3.00 (2.58 - 3.50) | 2.04 (1.73 - 2.42) | 1.23 (0.92 - 1.64) | Reference | 8.93 (7.54 - 10.59) | 2.55 (2.26 - 2.87) | 1.52 (1.31 - 1.75) | 1.34 (1.05 - 1.72) | Reference | 5.37 (4.77 - 6.04) | 2.10 (1.88 - 2.35) | 1.26 (1.09 - 1.44) | 1.02 (0.79 - 1.32) | Reference |
| Age adjusted HR (95% CI) | 1.32 (0.18 - 9.48) | 0.78 (0.63 - 0.96) | 0.85 (0.69 - 1.05) | 1.04 (0.78 - 1.39) | Reference | 1.08 (0.61 - 1.91) | 0.97 (0.83 - 1.13) | 1.35 (1.14 - 1.60) | 1.22 (0.92 - 1.62) | Reference | 0.84 (0.70 - 1.02) | 1.01 (0.90 - 1.14) | 1.39 (1.21 - 1.61) | 1.48 (1.15 - 1.89) | Reference | 0.84 (0.74 - 0.95) | 1.12 (1.00 - 1.25) | 1.36 (1.19 - 1.56) | 1.20 (0.93 - 1.55) | Reference |
| Age and nursing home adjusted HR (95% CI) | 0.96 (0.13 - 6.92) | 0.70 (0.56 - 0.87) | 0.85 (0.69 - 1.06) | 1.09 (0.81 - 1.45) | Reference | 0.83 (0.46 - 1.47) | 0.89 (0.76 - 1.04) | 1.29 (1.09 - 1.53) | 1.20 (0.90 - 1.60) | Reference | 0.68 (0.56 - 0.83) | 0.97 (0.86 - 1.10) | 1.31 (1.14 - 1.52) | 1.43 (1.12 - 1.83) | Reference | 0.78 (0.69 - 0.89) | 1.09 (0.98 - 1.22) | 1.32 (1.15 - 1.51) | 1.19 (0.92 - 1.53) | Reference |
|  | 2023 | | | | | | | | | | | | | | | | | | | |
|  | first quarter | | | | | second quarter | | | | | third quarter | | | | | fourth quarter | | | | |
|  | Four or more vaccine doses | Three vaccine doses | Two vaccine doses | One vaccine dose | Unvaccinated | Four or more vaccine doses | Three vaccine doses | Two vaccine doses | One vaccine dose | Unvaccinated | Four or more vaccine doses | Three vaccine doses | Two vaccine doses | One vaccine dose | Unvaccinated | Four or more vaccine doses | Three vaccine doses | Two vaccine doses | One vaccine dose | Unvaccinated |
|  | All-cause mortality | | | | | All-cause mortality | | | | | All-cause mortality | | | | | All-cause mortality | | | | |
| Deaths (n) | 1507 | 1536 | 401 | 58 | 469 | 1666 | 1376 | 399 | 86 | 467 | 1678 | 1465 | 399 | 64 | 450 | 1915 | 1550 | 387 | 82 | 478 |
| Events per 100,000 person days | 6.61 | 2.54 | 1.45 | 1.07 | 1.47 | 6.46 | 2.13 | 1.39 | 1.54 | 1.41 | 6.16 | 2.2 | 1.35 | 1.13 | 1.34 | 7.04 | 2.33 | 1.3 | 1.43 | 1.41 |
| Crude HR (95%CI) | 4.50 (4.06 - 4.99) | 1.73 (1.56 - 1.92) | 0.99 (0.87 - 1.13) | 0.73 (0.55 - 0.96) | Reference | 4.57 (4.12 - 5.06) | 1.51 (1.36 - 1.68) | 0.98 (0.86 - 1.12) | 1.09 (0.87 - 1.37) | Reference | 4.61 (4.15 - 5.11) | 1.65 (1.48 - 1.83) | 1.02 (0.89 - 1.16) | 0.84 (0.65 - 1.10) | Reference | 4.98 (4.51 - 5.51) | 1.65 (1.48 - 1.82) | 0.92 (0.81 - 1.06) | 1.02 (0.81 - 1.29) | Reference |
| Age adjusted HR (95% CI) | 0.80 (0.72 - 0.89) | 1.00 (0.90 - 1.11) | 1.14 (0.99 - 1.30) | 0.89 (0.68 - 1.17) | Reference | 0.83 (0.75 - 0.93) | 0.89 (0.80 - 0.99) | 1.15 (1.00 - 1.31) | 1.36 (1.08 - 1.71) | Reference | 0.91 (0.81 - 1.01) | 1.00 (0.90 - 1.11) | 1.19 (1.04 - 1.37) | 1.06 (0.82 - 1.38) | Reference | 0.93 (0.83 - 1.03) | 0.98 (0.89 - 1.09) | 1.10 (0.96 - 1.26) | 1.29 (1.02 - 1.63) | Reference |
| Age and nursing home adjusted HR (95% CI) | 0.77 (0.69 - 0.87) | 0.99 (0.89 - 1.09) | 1.10 (0.96 - 1.26) | 0.88 (0.67 - 1.16) | Reference | 0.81 (0.73 - 0.91) | 0.88 (0.79 - 0.97) | 1.12 (0.98 - 1.28) | 1.36 (1.08 - 1.71) | Reference | 0.90 (0.80 - 1.00) | 0.99 (0.89 - 1.10) | 1.17 (1.02 - 1.34) | 1.06 (0.82 - 1.38) | Reference | 0.91 (0.82 - 1.02) | 0.97 (0.88 - 1.08) | 1.08 (0.94 - 1.23) | 1.28 (1.02 - 1.62) | Reference |
|  | Non-COVID-19 mortality | | | | | Non-COVID-19 mortality | | | | | Non-COVID-19 mortality | | | | | Non-COVID-19 mortality | | | | |
| Deaths (n) | 1469 | 1499 | 393 | 58 | 458 | 1651 | 1364 | 396 | 85 | 463 | 1672 | 1455 | 398 | 64 | 446 | 1850 | 1488 | 378 | 80 | 465 |
| Events per 100,000 person days | 6.44 | 2.48 | 1.42 | 1.07 | 1.43 | 6.4 | 2.11 | 1.38 | 1.52 | 1.40 | 6.13 | 2.19 | 1.35 | 1.13 | 1.33 | 6.8 | 2.23 | 1.27 | 1.4 | 1.37 |
| Crude HR (95%CI) | 4.49 (4.04 - 4.99) | 1.73 (1.56 - 1.92) | 0.99 (0.87 - 1.14) | 0.75 (0.57 - 0.98) | Reference | 4.56 (4.12 - 5.06) | 1.51 (1.36 - 1.68) | 0.98 (0.86 - 1.13) | 1.09 (0.86 - 1.37) | Reference | 4.63 (4.17 - 5.14) | 1.65 (1.48 - 1.84) | 1.02 (0.89 - 1.17) | 0.85 (0.66 - 1.11) | Reference | 4.95 (4.47 - 5.47) | 1.62 (1.46 - 1.80) | 0.93 (0.81 - 1.06) | 1.02 (0.80 - 1.29) | Reference |
| Age adjusted HR (95% CI) | 0.81 (0.72 - 0.90) | 1.00 (0.90 - 1.12) | 1.14 (1.00 - 1.30) | 0.91 (0.70 - 1.20) | Reference | 0.84 (0.75 - 0.93) | 0.89 (0.80 - 0.99) | 1.15 (1.00 - 1.31) | 1.36 (1.08 - 1.71) | Reference | 0.91 (0.82 - 1.02) | 1.00 (0.90 - 1.11) | 1.20 (1.05 - 1.38) | 1.07 (0.83 - 1.39) | Reference | 0.92 (0.83 - 1.03) | 0.97 (0.88 - 1.08) | 1.10 (0.96 - 1.26) | 1.29 (1.02 - 1.64) | Reference |
| Age and nursing home adjusted HR (95% CI) | 0.78 (0.70 - 0.88) | 0.99 (0.89 - 1.10) | 1.11 (0.97 - 1.27) | 0.90 (0.69 - 1.19) | Reference | 0.82 (0.73 - 0.91) | 0.88 (0.79 - 0.98) | 1.12 (0.98 - 1.28) | 1.35 (1.07 - 1.70) | Reference | 0.90 (0.81 - 1.01) | 0.99 (0.89 - 1.10) | 1.18 (1.03 - 1.35) | 1.07 (0.82 - 1.39) | Reference | 0.91 (0.82 - 1.02) | 0.96 (0.87 - 1.07) | 1.08 (0.95 - 1.24) | 1.29 (1.02 - 1.63) | Reference |

| **Table S14:** Hazard ratios (HR) with 95% confidence intervals (95% CI) for non-COVID 19 and all-cause mortality according to number of SARS-CoV-2 vaccine doses for the period from 2021 to 2023 split into 3-month intervals for **females** only. | | | | | | | | | | | | | | | | | | | | |
| --- | --- | --- | --- | --- | --- | --- | --- | --- | --- | --- | --- | --- | --- | --- | --- | --- | --- | --- | --- | --- |
|  | 2021 | | | | | | | | | | | | | | | | | | | |
|  | first quarter | | | | | second quarter | | | | | third quarter | | | | | fourth quarter | | | | |
|  | Four or more vaccine doses | Three vaccine doses | Two vaccine doses | One vaccine dose | Unvaccinated | Four or more vaccine doses | Three vaccine doses | Two vaccine doses | One vaccine dose | Unvaccinated | Four or more vaccine doses | Three vaccine doses | Two vaccine doses | One vaccine dose | Unvaccinated | Four or more vaccine doses | Three vaccine doses | Two vaccine doses | One vaccine dose | Unvaccinated |
|  | All-cause mortality | | | | | All-cause mortality | | | | | All-cause mortality | | | | | All-cause mortality | | | | |
| Deaths (n) |  |  | 101 | 44 | 215 |  |  | 362 | 96 | 403 |  | 10 | 442 | 150 | 330 |  | 272 | 375 | 142 | 252 |
| Events per 100,000 person days | |  | 20.01 | 9.53 | 3.18 |  |  | 14.91 | 2.81 | 3.76 |  | 30.4 | 6.45 | 2.61 | 3.40 |  | 11.94 | 4.24 | 2.14 | 3.63 |
| Crude HR (95%CI) |  |  | 6.37 (4.99 - 8.14) | 3.24 (2.33 - 4.52) | Reference |  |  | 4.18 (3.61 - 4.82) | 0.81 (0.64 - 1.01) | Reference |  | 9.37 (4.80 - 18.28) | 1.91 (1.66 - 2.21) | 0.78 (0.64 - 0.95) | Reference |  | 4.23 (3.48 - 5.14) | 1.19 (1.01 - 1.40) | 0.61 (0.49 - 0.74) | Reference |
| Age adjusted HR (95% CI) |  |  | 0.84 (0.65 - 1.10) | 1.00 (0.72 - 1.40) | Reference |  |  | 0.77 (0.66 - 0.89) | 0.57 (0.46 - 0.72) | Reference |  | 0.72 (0.36 - 1.40) | 0.76 (0.66 - 0.88) | 0.62 (0.51 - 0.75) | Reference |  | 0.71 (0.59 - 0.85) | 0.78 (0.66 - 0.91) | 0.60 (0.49 - 0.74) | Reference |
| Age and nursing home adjusted HR (95% CI) |  |  | 0.49 (0.37 - 0.64) | 0.79 (0.56 - 1.12) | Reference |  |  | 0.62 (0.53 - 0.72) | 0.58 (0.46 - 0.72) | Reference |  | 0.55 (0.28 - 1.09) | 0.65 (0.56 - 0.75) | 0.64 (0.52 - 0.77) | Reference |  | 0.55 (0.45 - 0.66) | 0.72 (0.61 - 0.84) | 0.64 (0.52 - 0.79) | Reference |
|  | Non-COVID-19 mortality | | | | | Non-COVID-19 mortality | | | | | Non-COVID-19 mortality | | | | | Non-COVID-19 mortality | | | | |
| Deaths (n) |  |  | 94 | 41 | 199 |  |  | 346 | 94 | 382 |  | 10 | 437 | 147 | 317 |  | 271 | 366 | 141 | 239 |
| Events per 100,000 person days | |  | 18.62 | 8.88 | 2.94 |  |  | 14.25 | 2.75 | 3.56 |  | 30.4 | 6.38 | 2.56 | 3.27 |  | 11.89 | 4.14 | 2.13 | 3.44 |
| Crude HR (95%CI) |  |  | 6.35 (4.92 - 8.18) | 3.29 (2.33 - 4.64) | Reference |  |  | 4.19 (3.62 - 4.86) | 0.83 (0.66 - 1.04) | Reference |  | 9.44 (4.84 - 18.43) | 1.97 (1.70 - 2.27) | 0.80 (0.65 - 0.97) | Reference |  | 4.57 (3.75 - 5.58) | 1.23 (1.04 - 1.44) | 0.64 (0.52 - 0.78) | Reference |
| Age adjusted HR (95% CI) |  |  | 0.85 (0.65 - 1.11) | 1.03 (0.73 - 1.46) | Reference |  |  | 0.77 (0.66 - 0.89) | 0.59 (0.47 - 0.74) | Reference |  | 0.72 (0.36 - 1.40) | 0.78 (0.68 - 0.91) | 0.63 (0.52 - 0.77) | Reference |  | 0.75 (0.63 - 0.91) | 0.80 (0.68 - 0.94) | 0.63 (0.51 - 0.78) | Reference |
| Age and nursing home adjusted HR (95% CI) |  |  | 0.50 (0.38 - 0.67) | 0.83 (0.58 - 1.17) | Reference |  |  | 0.61 (0.52 - 0.72) | 0.60 (0.47 - 0.75) | Reference |  | 0.54 (0.27 - 1.06) | 0.66 (0.57 - 0.77) | 0.65 (0.53 - 0.79) | Reference |  | 0.58 (0.48 - 0.70) | 0.74 (0.63 - 0.87) | 0.67 (0.54 - 0.83) | Reference |
|  | 2022 | | | | | | | | | | | | | | | | | | | |
|  | first quarter | | | | | second quarter | | | | | third quarter | | | | | fourth quarter | | | | |
|  | Four or more vaccine doses | Three vaccine doses | Two vaccine doses | One vaccine dose | Unvaccinated | Four or more vaccine doses | Three vaccine doses | Two vaccine doses | One vaccine dose | Unvaccinated | Four or more vaccine doses | Three vaccine doses | Two vaccine doses | One vaccine dose | Unvaccinated | Four or more vaccine doses | Three vaccine doses | Two vaccine doses | One vaccine dose | Unvaccinated |
|  | All-cause mortality | | | | | All-cause mortality | | | | | All-cause mortality | | | | | All-cause mortality | | | | |
| Deaths (n) | 5 | 487 | 357 | 87 | 193 | 23 | 913 | 391 | 69 | 310 | 384 | 1636 | 475 | 96 | 447 | 1313 | 1645 | 442 | 96 | 510 |
| Events per 100,000 person days | 87.26 | 5.88 | 3.13 | 1.87 | 2.05 | 21.78 | 3.23 | 1.92 | 1.38 | 1.35 | 13.36 | 2.67 | 1.73 | 1.78 | 1.37 | 8.02 | 2.48 | 1.57 | 1.74 | 1.46 |
| Crude HR (95%CI) | 47.68 (19.60 - 115.96) | 2.80 (2.37 - 3.31) | 1.50 (1.26 - 1.79) | 0.83 (0.64 - 1.07) | Reference | 17.44 (11.32 - 26.87) | 2.53 (2.22 - 2.88) | 1.42 (1.23 - 1.65) | 1.01 (0.78 - 1.31) | Reference | 10.47 (9.08 - 12.06) | 1.96 (1.76 - 2.17) | 1.26 (1.11 - 1.44) | 1.30 (1.04 - 1.62) | Reference | 5.56 (5.01 - 6.17) | 1.72 (1.55 - 1.89) | 1.08 (0.95 - 1.22) | 1.19 (0.96 - 1.48) | Reference |
| Age adjusted HR (95% CI) | 5.43 (2.23 - 13.27) | 0.81 (0.69 - 0.96) | 0.86 (0.72 - 1.03) | 0.88 (0.68 - 1.14) | Reference | 1.00 (0.65 - 1.55) | 0.87 (0.76 - 0.99) | 0.99 (0.85 - 1.15) | 0.96 (0.74 - 1.25) | Reference | 0.93 (0.81 - 1.08) | 0.94 (0.85 - 1.05) | 1.18 (1.04 - 1.34) | 1.33 (1.07 - 1.66) | Reference | 0.94 (0.85 - 1.04) | 1.15 (1.04 - 1.27) | 1.21 (1.07 - 1.38) | 1.36 (1.09 - 1.69) | Reference |
| Age and nursing home adjusted HR (95% CI) | 4.08 (1.66 - 10.03) | 0.66 (0.56 - 0.79) | 0.87 (0.73 - 1.03) | 0.93 (0.72 - 1.21) | Reference | 0.70 (0.45 - 1.10) | 0.73 (0.64 - 0.83) | 0.94 (0.81 - 1.09) | 0.95 (0.73 - 1.24) | Reference | 0.69 (0.59 - 0.81) | 0.83 (0.75 - 0.92) | 1.12 (0.98 - 1.28) | 1.29 (1.03 - 1.60) | Reference | 0.75 (0.67 - 0.84) | 1.07 (0.97 - 1.18) | 1.17 (1.03 - 1.33) | 1.33 (1.07 - 1.66) | Reference |

| **Table S14 continued:** | | | | | | | | | | | | | | | | | | | | |
| --- | --- | --- | --- | --- | --- | --- | --- | --- | --- | --- | --- | --- | --- | --- | --- | --- | --- | --- | --- | --- |
|  | 2022 | | | | | | | | | | | | | | | | | | | |
|  | first quarter | | | | | second quarter | | | | | third quarter | | | | | fourth quarter | | | | |
|  | Four or more vaccine doses | Three vaccine doses | Two vaccine doses | One vaccine dose | Unvacc | Four or more vaccine doses | Three vaccine doses | Two vaccine doses | One vaccine dose | Unvacc | Four or more vaccine doses | Three vaccine doses | Two vaccine doses | One vaccine dose | Unvacc | Four or more vaccine doses | Three vaccine doses | Two vaccine doses | One vaccine dose | Unvacc |
|  | Non-COVID-19 mortality | | | | | Non-COVID-19 mortality | | | | | Non-COVID-19 mortality | | | | | Non-COVID-19 mortality | | | | |
| Deaths (n) | 3 | 461 | 338 | 85 | 172 | 23 | 890 | 376 | 69 | 306 | 381 | 1599 | 457 | 92 | 438 | 1282 | 1606 | 435 | 95 | 497 |
| Events per 100,000 person days | 52.36 | 5.57 | 2.96 | 1.83 | 1.83 | 21.78 | 3.14 | 1.84 | 1.38 | 1.33 | 13.26 | 2.61 | 1.66 | 1.7 | 1.35 | 7.83 | 2.42 | 1.54 | 1.72 | 1.42 |
| Crude HR (95%CI) | 32.55 (10.39 - 102.03) | 2.96 (2.48 - 3.53) | 1.59 (1.32 - 1.91) | 0.89 (0.69 - 1.16) | Reference | 17.54 (11.38 - 27.02) | 2.49 (2.19 - 2.84) | 1.39 (1.19 - 1.61) | 1.02 (0.79 - 1.33) | Reference | 10.56 (9.15 - 12.18) | 1.95 (1.76 - 2.17) | 1.24 (1.09 - 1.41) | 1.27 (1.01 - 1.59) | Reference | 5.55 (5.00 - 6.17) | 1.72 (1.55 - 1.90) | 1.09 (0.96 - 1.24) | 1.21 (0.97 - 1.51) | Reference |
| Age adjusted HR (95% CI) | 3.76 (1.20 - 11.82) | 0.86 (0.72 - 1.03) | 0.92 (0.76 - 1.10) | 0.95 (0.73 - 1.24) | Reference | 1.01 (0.65 - 1.56) | 0.86 (0.75 - 0.98) | 0.96 (0.83 - 1.12) | 0.98 (0.75 - 1.27) | Reference | 0.95 (0.82 - 1.10) | 0.94 (0.85 - 1.04) | 1.16 (1.02 - 1.32) | 1.30 (1.04 - 1.63) | Reference | 0.94 (0.85 - 1.04) | 1.15 (1.04 - 1.27) | 1.23 (1.08 - 1.39) | 1.38 (1.11 - 1.72) | Reference |
| Age and nursing home adjusted HR (95% CI) | 2.91 (0.92 - 9.22) | 0.71 (0.59 - 0.85) | 0.92 (0.77 - 1.11) | 1.00 (0.77 - 1.31) | Reference | 0.71 (0.45 - 1.11) | 0.72 (0.63 - 0.82) | 0.92 (0.79 - 1.07) | 0.97 (0.74 - 1.26) | Reference | 0.70 (0.60 - 0.82) | 0.83 (0.74 - 0.92) | 1.10 (0.97 - 1.26) | 1.26 (1.01 - 1.58) | Reference | 0.75 (0.68 - 0.84) | 1.07 (0.97 - 1.19) | 1.19 (1.04 - 1.35) | 1.35 (1.09 - 1.69) | Reference |
|  | 2023 | | | | | | | | | | | | | | | | | | | |
|  | first quarter | | | | | second quarter | | | | | third quarter | | | | | fourth quarter | | | | |
|  | Four or more vaccine doses | Three vaccine doses | Two vaccine doses | One vaccine dose | Unvaccinated | Four or more vaccine doses | Three vaccine doses | Two vaccine doses | One vaccine dose | Unvaccinated | Four or more vaccine doses | Three vaccine doses | Two vaccine doses | One vaccine dose | Unvaccinated | Four or more vaccine doses | Three vaccine doses | Two vaccine doses | One vaccine dose | Unvaccinated |
|  | All-cause mortality | | | | | All-cause mortality | | | | | All-cause mortality | | | | | All-cause mortality | | | | |
| Deaths (n) | 1678 | 1526 | 431 | 77 | 580 | 1701 | 1396 | 400 | 79 | 544 | 1815 | 1361 | 371 | 85 | 536 | 2036 | 1534 | 385 | 70 | 563 |
| Events per 100,000 person days | 6.71 | 2.21 | 1.52 | 1.41 | 1.64 | 5.93 | 1.89 | 1.36 | 1.41 | 1.48 | 5.96 | 1.78 | 1.23 | 1.49 | 1.43 | 6.7 | 2.01 | 1.26 | 1.22 | 1.50 |
| Crude HR (95%CI) | 4.11 (3.74 - 4.51) | 1.35 (1.23 - 1.49) | 0.93 (0.82 - 1.05) | 0.86 (0.68 - 1.09) | Reference | 4.01 (3.64 - 4.41) | 1.28 (1.16 - 1.41) | 0.92 (0.81 - 1.04) | 0.95 (0.75 - 1.20) | Reference | 4.16 (3.78 - 4.58) | 1.25 (1.13 - 1.38) | 0.86 (0.75 - 0.98) | 1.04 (0.83 - 1.31) | Reference | 4.47 (4.07 - 4.90) | 1.34 (1.21 - 1.47) | 0.85 (0.74 - 0.96) | 0.82 (0.64 - 1.04) | Reference |
| Age adjusted HR (95% CI) | 0.86 (0.79 - 0.95) | 0.96 (0.87 - 1.05) | 1.12 (0.99 - 1.26) | 1.03 (0.81 - 1.30) | Reference | 0.90 (0.82 - 0.99) | 0.92 (0.83 - 1.01) | 1.12 (0.98 - 1.27) | 1.16 (0.92 - 1.47) | Reference | 0.97 (0.88 - 1.07) | 0.90 (0.81 - 0.99) | 1.06 (0.93 - 1.21) | 1.29 (1.02 - 1.62) | Reference | 0.99 (0.90 - 1.09) | 0.96 (0.87 - 1.05) | 1.05 (0.92 - 1.20) | 1.01 (0.79 - 1.29) | Reference |
| Age and nursing home adjusted HR (95% CI) | 0.73 (0.66 - 0.80) | 0.92 (0.83 - 1.01) | 1.09 (0.96 - 1.23) | 1.00 (0.79 - 1.27) | Reference | 0.79 (0.72 - 0.87) | 0.89 (0.81 - 0.98) | 1.10 (0.96 - 1.25) | 1.14 (0.90 - 1.45) | Reference | 0.87 (0.79 - 0.96) | 0.87 (0.79 - 0.97) | 1.02 (0.90 - 1.17) | 1.25 (1.00 - 1.57) | Reference | 0.88 (0.80 - 0.97) | 0.94 (0.85 - 1.03) | 1.02 (0.90 - 1.17) | 0.99 (0.77 - 1.27) | Reference |
|  | Non-COVID-19 mortality | | | | | Non-COVID-19 mortality | | | | | Non-COVID-19 mortality | | | | | Non-COVID-19 mortality | | | | |
| Deaths (n) | 1618 | 1498 | 420 | 77 | 563 | 1683 | 1383 | 394 | 78 | 538 | 1810 | 1353 | 369 | 83 | 534 | 1973 | 1493 | 371 | 69 | 549 |
| Events per 100,000 person days | 6.47 | 2.17 | 1.48 | 1.41 | 1.59 | 5.87 | 1.87 | 1.34 | 1.39 | 1.47 | 5.94 | 1.77 | 1.22 | 1.45 | 1.43 | 6.49 | 1.95 | 1.22 | 1.2 | 1.46 |
| Crude HR (95%CI) | 4.08 (3.71 - 4.49) | 1.37 (1.24 - 1.51) | 0.93 (0.82 - 1.06) | 0.89 (0.70 - 1.13) | Reference | 4.01 (3.64 - 4.42) | 1.28 (1.16 - 1.41) | 0.91 (0.80 - 1.04) | 0.95 (0.75 - 1.20) | Reference | 4.17 (3.78 - 4.59) | 1.24 (1.12 - 1.37) | 0.86 (0.75 - 0.98) | 1.02 (0.81 - 1.29) | Reference | 4.44 (4.04 - 4.88) | 1.34 (1.21 - 1.47) | 0.84 (0.73 - 0.95) | 0.82 (0.64 - 1.06) | Reference |
| Age adjusted HR (95% CI) | 0.86 (0.78 - 0.95) | 0.97 (0.88 - 1.07) | 1.12 (0.99 - 1.27) | 1.06 (0.83 - 1.34) | Reference | 0.90 (0.82 - 0.99) | 0.92 (0.83 - 1.01) | 1.12 (0.98 - 1.27) | 1.16 (0.92 - 1.47) | Reference | 0.97 (0.88 - 1.07) | 0.90 (0.81 - 0.99) | 1.05 (0.92 - 1.20) | 1.26 (1.00 - 1.59) | Reference | 0.98 (0.89 - 1.08) | 0.96 (0.87 - 1.05) | 1.04 (0.91 - 1.18) | 1.02 (0.79 - 1.31) | Reference |
| Age and nursing home adjusted HR (95% CI) | 0.73 (0.66 - 0.81) | 0.93 (0.84 - 1.02) | 1.09 (0.96 - 1.24) | 1.04 (0.82 - 1.31) | Reference | 0.79 (0.72 - 0.87) | 0.89 (0.81 - 0.98) | 1.09 (0.96 - 1.24) | 1.14 (0.90 - 1.45) | Reference | 0.87 (0.79 - 0.97) | 0.87 (0.79 - 0.96) | 1.02 (0.90 - 1.17) | 1.23 (0.97 - 1.55) | Reference | 0.88 (0.80 - 0.97) | 0.93 (0.85 - 1.03) | 1.01 (0.89 - 1.15) | 1.00 (0.78 - 1.29) | Reference |

| **Table S15:** Hazard ratios (HR) with 95% confidence intervals (95% CI) for non-COVID 19 and all-cause mortality according to number of SARS-CoV-2 vaccine doses for the period from 2021 to 2023 split into 3-month intervals for **18-39 year old** only. | | | | | | | | | | | | | | | | | | | | |
| --- | --- | --- | --- | --- | --- | --- | --- | --- | --- | --- | --- | --- | --- | --- | --- | --- | --- | --- | --- | --- |
|  | 2021 | | | | | | | | | | | | | | | | | | | |
|  | first quarter | | | | | second quarter | | | | | third quarter | | | | | fourth quarter | | | | |
|  | Four or more vaccine doses | Three vaccine doses | Two vaccine doses | One vaccine dose | Unvaccinated | Four or more vaccine doses | Three vaccine doses | Two vaccine doses | One vaccine dose | Unvaccinated | Four or more vaccine doses | Three vaccine doses | Two vaccine doses | One vaccine dose | Unvaccinated | Four or more vaccine doses | Three vaccine doses | Two vaccine doses | One vaccine dose | Unvaccinated |
|  | All-cause mortality | | | | | All-cause mortality | | | | | All-cause mortality | | | | | All-cause mortality | | | | |
| Deaths (n) |  |  |  |  | 2 |  |  | 1 | 2 | 9 |  |  | 7 | 1 | 16 |  | 1* | 5 | 3* | 6 |
| Events per 100,000 person days |  |  |  |  | 0.03 |  |  | 0.12 | 0.1 | 0.09 |  |  | 0.16 | 0.02 | 0.18 |  | 0.11 | 0.08 | 0.05 | 0.09 |
| Crude HR (95%CI) |  |  |  |  | Reference |  |  | 1.34 (0.16 - 10.91) | 1.34 (0.27 - 6.60) | Reference |  |  | 0.92 (0.37 - 2.25) | 0.13 (0.02 - 0.97) | Reference |  |  | 0.80 (0.24 - 2.64) |  | Reference |
| Age adjusted HR (95% CI) |  |  |  |  | Reference |  |  | 1.25 (0.15 - 10.21) | 1.27 (0.26 - 6.26) | Reference |  |  | 0.92 (0.37 - 2.25) | 0.13 (0.02 - 0.97) | Reference |  |  | 0.80 (0.24 - 2.63) |  | Reference |
| Age and gender adjusted HR (95% CI) |  |  |  |  | Reference |  |  | ( - ) | ( - ) | Reference |  |  | 0.91 (0.37 - 2.23) | ( - ) | Reference |  |  | 0.79 (0.24 - 2.61) |  | Reference |
| Age, gender and nursing home adjusted HR (95% CI) |  |  |  |  | Reference |  |  | 1.25 (0.15 - 10.21) | 1.27 (0.26 - 6.26) | Reference |  |  | 0.91 (0.37 - 2.23) | 0.13 (0.02 - 0.97) | Reference |  |  | 0.79 (0.24 - 2.61) |  | Reference |
|  | Non-COVID-19 mortality | | | | | Non-COVID-19 mortality | | | | | Non-COVID-19 mortality | | | | | Non-COVID-19 mortality | | | | |
| Deaths (n) |  |  |  |  | 2 |  |  | 1 | 2 | 9 |  |  | 7 | 1 | 15 |  | 1* | 5 | 3* | 5 |
| Events per 100,000 person days |  |  |  |  | 0.03 |  |  | 0.12 | 0.1 | 0.09 |  |  | 0.16 | 0.02 | 0.17 |  | 0.11 | 0.08 | 0.05 | 0.07 |
| Crude HR (95%CI) |  |  |  |  | Reference |  |  | 1.34 (0.16 - 10.91) | 1.34 (0.27 - 6.60) | Reference |  |  | 0.96 (0.39 - 2.39) | 0.14 (0.02 - 1.03) | Reference |  |  | 0.98 (0.28 - 3.41) |  | Reference |
| Age adjusted HR (95% CI) |  |  |  |  | Reference |  |  | 1.25 (0.15 - 10.21) | 1.27 (0.26 - 6.26) | Reference |  |  | 0.97 (0.39 - 2.40) | 0.14 (0.02 - 1.03) | Reference |  |  | 0.98 (0.28 - 3.39) |  | Reference |
| Age and gender adjusted HR (95% CI) |  |  |  |  | Reference |  |  | ( - ) | ( - ) | Reference |  |  | 0.95 (0.38 - 2.37) | ( - ) | Reference |  |  | 0.97 (0.28 - 3.37) |  | Reference |
| Age, gender and nursing home adjusted HR (95% CI) |  |  |  |  | Reference |  |  | 1.25 (0.15 - 10.21) | 1.27 (0.26 - 6.26) | Reference |  |  | 0.95 (0.38 - 2.37) | 0.14 (0.02 - 1.03) | Reference |  |  | 0.97 (0.28 - 3.37) |  | Reference |
|  | 2022 | | | | | | | | | | | | | | | | | | | |
|  | first quarter | | | | | second quarter | | | | | third quarter | | | | | fourth quarter | | | | |
|  | Four or more vaccine doses | Three vaccine doses | Two vaccine doses | One vaccine dose | Unvaccinated | Four or more vaccine doses | Three vaccine doses | Two vaccine doses | One vaccine dose | Unvaccinated | Four or more vaccine doses | Three vaccine doses | Two vaccine doses | One vaccine dose | Unvaccinated | Four or more vaccine doses | Three vaccine doses | Two vaccine doses | One vaccine dose | Unvaccinated |
|  | All-cause mortality | | | | | All-cause mortality | | | | | All-cause mortality | | | | | All-cause mortality | | | | |
| Deaths (n) |  | 5 | 7 | 3* | 6 |  | 9 | 22 | 4 | 28 | 1 | 31 | 36 | 6 | 38 | 4 | 45 | 42 | 5 | 40 |
| Events per 100,000 person days |  | 0.1 | 0.08 | 0.06 | 0.06 |  | 0.05 | 0.11 | 0.07 | 0.13 | 0.25 | 0.07 | 0.13 | 0.1 | 0.13 | 0.08 | 0.09 | 0.14 | 0.08 | 0.13 |
| Crude HR (95%CI) |  | 1.52 (0.46 - 4.97) | 1.21 (0.41 - 3.62) |  | Reference |  | 0.36 (0.17 - 0.76) | 0.88 (0.51 - 1.55) | 0.59 (0.21 - 1.68) | Reference | 1.97 (0.27 - 14.49) | 0.58 (0.36 - 0.94) | 1.00 (0.64 - 1.58) | 0.79 (0.34 - 1.88) | Reference | 0.68 (0.24 - 1.90) | 0.73 (0.48 - 1.12) | 1.12 (0.72 - 1.72) | 0.65 (0.26 - 1.64) | Reference |
| Age adjusted HR (95% CI) |  | 1.52 (0.46 - 4.97) | 1.23 (0.41 - 3.68) |  | Reference |  | 0.36 (0.17 - 0.76) | 0.89 (0.51 - 1.56) | 0.57 (0.20 - 1.63) | Reference | 1.86 (0.25 - 13.73) | 0.59 (0.36 - 0.94) | 1.05 (0.66 - 1.65) | 0.82 (0.35 - 1.94) | Reference | 0.64 (0.23 - 1.79) | 0.74 (0.48 - 1.13) | 1.15 (0.74 - 1.77) | 0.68 (0.27 - 1.73) | Reference |
| Age and gender adjusted HR (95% CI) |  | 1.51 (0.46 - 4.95) | 1.22 (0.41 - 3.65) |  | Reference |  | 0.36 (0.17 - 0.77) | 0.89 (0.51 - 1.56) | 0.56 (0.20 - 1.61) | Reference | ( - ) | 0.59 (0.37 - 0.95) | 1.05 (0.66 - 1.65) | 0.81 (0.34 - 1.92) | Reference | 0.65 (0.23 - 1.84) | 0.75 (0.49 - 1.15) | 1.14 (0.74 - 1.76) | 0.67 (0.27 - 1.71) | Reference |
| Age, gender and nursing home adjusted HR (95% CI) |  | 1.51 (0.46 - 4.95) | 1.22 (0.41 - 3.65) |  | Reference |  | 0.36 (0.17 - 0.77) | 0.89 (0.51 - 1.56) | 0.56 (0.20 - 1.61) | Reference | 1.86 (0.25 - 13.73) | 0.59 (0.37 - 0.95) | 1.05 (0.66 - 1.65) | 0.81 (0.34 - 1.92) | Reference | 0.65 (0.23 - 1.84) | 0.75 (0.49 - 1.15) | 1.14 (0.74 - 1.76) | 0.67 (0.27 - 1.71) | Reference |

| **Table S15 continued:** | | | | | | | | | | | | | | | | | | | | |
| --- | --- | --- | --- | --- | --- | --- | --- | --- | --- | --- | --- | --- | --- | --- | --- | --- | --- | --- | --- | --- |
|  | 2022 | | | | | | | | | | | | | | | | | | | |
|  | first quarter | | | | | second quarter | | | | | third quarter | | | | | fourth quarter | | | | |
|  | Four or more vaccine doses | Three vaccine doses | Two vaccine doses | One vaccine dose | Unvacc | Four or more vaccine doses | Three vaccine doses | Two vaccine doses | One vaccine dose | Unvacc | Four or more vaccine doses | Three vaccine doses | Two vaccine doses | One vaccine dose | Unvacc | Four or more vaccine doses | Three vaccine doses | Two vaccine doses | One vaccine dose | Unvacc |
|  | Non-COVID-19 mortality | | | | | Non-COVID-19 mortality | | | | | Non-COVID-19 mortality | | | | | Non-COVID-19 mortality | | | | |
| Deaths (n) |  | 5 | 7 | 3* | 6 |  | 9 | 20 | 4 | 28 | 1 | 29 | 35 | 6 | 38 | 4 | 44 | 42 | 5 | 39 |
| Events per 100,000 person days |  | 0.1 | 0.08 | 0.06 | 0.06 |  | 0.05 | 0.1 | 0.07 | 0.13 | 0.25 | 0.07 | 0.12 | 0.1 | 0.13 | 0.08 | 0.09 | 0.14 | 0.08 | 0.12 |
| Crude HR (95%CI) |  | 1.52 (0.46 - 4.97) | 1.21 (0.41 - 3.62) |  | Reference |  | 0.36 (0.17 - 0.76) | 0.80 (0.45 - 1.43) | 0.59 (0.21 - 1.68) | Reference | 1.97 (0.27 - 14.49) | 0.54 (0.34 - 0.88) | 0.97 (0.62 - 1.54) | 0.79 (0.34 - 1.88) | Reference | 0.68 (0.24 - 1.92) | 0.73 (0.48 - 1.13) | 1.14 (0.74 - 1.77) | 0.66 (0.26 - 1.68) | Reference |
| Age adjusted HR (95% CI) |  | 1.52 (0.46 - 4.97) | 1.23 (0.41 - 3.68) |  | Reference |  | 0.36 (0.17 - 0.76) | 0.81 (0.46 - 1.44) | 0.57 (0.20 - 1.63) | Reference | 1.86 (0.25 - 13.73) | 0.55 (0.34 - 0.89) | 1.02 (0.64 - 1.62) | 0.82 (0.35 - 1.94) | Reference | 0.64 (0.23 - 1.81) | 0.74 (0.48 - 1.14) | 1.18 (0.76 - 1.83) | 0.70 (0.28 - 1.78) | Reference |
| Age and gender adjusted HR (95% CI) |  | 1.51 (0.46 - 4.95) | 1.22 (0.41 - 3.65) |  | Reference |  | 0.36 (0.17 - 0.77) | 0.81 (0.46 - 1.44) | 0.56 (0.20 - 1.61) | Reference | ( - ) | 0.55 (0.34 - 0.90) | 1.02 (0.64 - 1.61) | 0.81 (0.34 - 1.92) | Reference | 0.66 (0.23 - 1.86) | 0.75 (0.49 - 1.16) | 1.17 (0.76 - 1.81) | 0.69 (0.27 - 1.76) | Reference |
| Age, gender and nursing home adjusted HR (95% CI) |  | 1.51 (0.46 - 4.95) | 1.22 (0.41 - 3.65) |  | Reference |  | 0.36 (0.17 - 0.77) | 0.81 (0.46 - 1.44) | 0.56 (0.20 - 1.61) | Reference | 1.86 (0.25 - 13.73) | 0.55 (0.34 - 0.90) | 1.02 (0.64 - 1.61) | 0.81 (0.34 - 1.92) | Reference | 0.66 (0.23 - 1.86) | 0.75 (0.49 - 1.16) | 1.17 (0.76 - 1.81) | 0.69 (0.27 - 1.76) | Reference |
|  | 2023 | | | | | | | | | | | | | | | | | | | |
|  | first quarter | | | | | second quarter | | | | | third quarter | | | | | fourth quarter | | | | |
|  | Four or more vaccine doses | Three vaccine doses | Two vaccine doses | One vaccine dose | Unvaccinated | Four or more vaccine doses | Three vaccine doses | Two vaccine doses | One vaccine dose | Unvaccinated | Four or more vaccine doses | Three vaccine doses | Two vaccine doses | One vaccine dose | Unvaccinated | Four or more vaccine doses | Three vaccine doses | Two vaccine doses | One vaccine dose | Unvaccinated |
|  | All-cause mortality | | | | | All-cause mortality | | | | | All-cause mortality | | | | | All-cause mortality | | | | |
| Deaths (n) | 10 | 55 | 32 | 7 | 41 | 8 | 53 | 29 | 4 | 43 | 21 | 59 | 45 | 14 | 56 | 9 | 61 | 35 | 8 | 42 |
| Events per 100,000 person days | 0.12 | 0.11 | 0.11 | 0.11 | 0.13 | 0.09 | 0.1 | 0.09 | 0.06 | 0.13 | 0.22 | 0.11 | 0.14 | 0.22 | 0.17 | 0.1 | 0.11 | 0.11 | 0.13 | 0.13 |
| Crude HR (95%CI) | 0.97 (0.48 - 1.93) | 0.84 (0.56 - 1.26) | 0.82 (0.52 - 1.30) | 0.90 (0.40 - 2.00) | Reference | 0.67 (0.32 - 1.43) | 0.75 (0.50 - 1.12) | 0.70 (0.44 - 1.13) | 0.49 (0.18 - 1.36) | Reference | 1.32 (0.80 - 2.18) | 0.64 (0.44 - 0.92) | 0.83 (0.56 - 1.23) | 1.31 (0.73 - 2.36) | Reference | 0.76 (0.37 - 1.56) | 0.88 (0.59 - 1.30) | 0.86 (0.55 - 1.34) | 1.00 (0.47 - 2.13) | Reference |
| Age adjusted HR (95% CI) | 0.96 (0.48 - 1.92) | 0.85 (0.56 - 1.27) | 0.81 (0.51 - 1.29) | 0.89 (0.40 - 1.98) | Reference | 0.64 (0.30 - 1.36) | 0.75 (0.50 - 1.13) | 0.72 (0.45 - 1.16) | 0.52 (0.19 - 1.44) | Reference | 1.24 (0.75 - 2.06) | 0.64 (0.44 - 0.92) | 0.85 (0.57 - 1.26) | 1.36 (0.76 - 2.45) | Reference | 0.71 (0.35 - 1.47) | 0.88 (0.60 - 1.31) | 0.88 (0.56 - 1.38) | 1.06 (0.50 - 2.26) | Reference |
| Age and gender adjusted HR (95% CI) | 1.01 (0.50 - 2.02) | 0.86 (0.57 - 1.28) | 0.81 (0.51 - 1.28) | 0.87 (0.39 - 1.94) | Reference | 0.65 (0.30 - 1.38) | 0.76 (0.51 - 1.14) | 0.72 (0.45 - 1.15) | 0.51 (0.18 - 1.42) | Reference | 1.27 (0.77 - 2.10) | 0.64 (0.45 - 0.93) | 0.84 (0.57 - 1.25) | 1.34 (0.75 - 2.41) | Reference | 0.72 (0.35 - 1.49) | 0.89 (0.60 - 1.31) | 0.87 (0.56 - 1.37) | 1.05 (0.49 - 2.24) | Reference |
| Age, gender and nursing home adjusted HR (95% CI) | 1.01 (0.50 - 2.02) | 0.86 (0.57 - 1.28) | 0.81 (0.51 - 1.28) | 0.87 (0.39 - 1.94) | Reference | 0.65 (0.30 - 1.38) | 0.76 (0.51 - 1.14) | 0.72 (0.45 - 1.15) | 0.51 (0.18 - 1.42) | Reference | 1.27 (0.77 - 2.10) | 0.64 (0.45 - 0.93) | 0.84 (0.57 - 1.25) | 1.34 (0.75 - 2.41) | Reference | 0.72 (0.35 - 1.49) | 0.89 (0.60 - 1.31) | 0.87 (0.56 - 1.37) | 1.05 (0.49 - 2.24) | Reference |
|  | Non-COVID-19 mortality | | | | | Non-COVID-19 mortality | | | | | Non-COVID-19 mortality | | | | | Non-COVID-19 mortality | | | | |
| Deaths (n) | 10 | 55 | 32 | 7 | 40 | 8 | 53 | 29 | 4 | 43 | 21 | 59 | 45 | 14 | 56 | 9 | 58 | 35 | 8 | 42 |
| Events per 100,000 person days | 0.12 | 0.11 | 0.11 | 0.11 | 0.13 | 0.09 | 0.1 | 0.09 | 0.06 | 0.13 | 0.22 | 0.11 | 0.14 | 0.22 | 0.17 | 0.1 | 0.11 | 0.11 | 0.13 | 0.13 |
| Crude HR (95%CI) | 0.99 (0.49 - 1.98) | 0.86 (0.57 - 1.30) | 0.84 (0.53 - 1.34) | 0.92 (0.41 - 2.05) | Reference | 0.67 (0.32 - 1.43) | 0.75 (0.50 - 1.12) | 0.70 (0.44 - 1.13) | 0.49 (0.18 - 1.36) | Reference | 1.32 (0.80 - 2.18) | 0.64 (0.44 - 0.92) | 0.83 (0.56 - 1.23) | 1.31 (0.73 - 2.36) | Reference | 0.76 (0.37 - 1.56) | 0.84 (0.56 - 1.24) | 0.86 (0.55 - 1.34) | 1.00 (0.47 - 2.13) | Reference |
| Age adjusted HR (95% CI) | 0.98 (0.49 - 1.97) | 0.87 (0.58 - 1.30) | 0.83 (0.52 - 1.33) | 0.91 (0.41 - 2.04) | Reference | 0.64 (0.30 - 1.36) | 0.75 (0.50 - 1.13) | 0.72 (0.45 - 1.16) | 0.52 (0.19 - 1.44) | Reference | 1.24 (0.75 - 2.06) | 0.64 (0.44 - 0.92) | 0.85 (0.57 - 1.26) | 1.36 (0.76 - 2.45) | Reference | 0.71 (0.35 - 1.47) | 0.84 (0.56 - 1.25) | 0.88 (0.56 - 1.38) | 1.06 (0.50 - 2.26) | Reference |
| Age and gender adjusted HR (95% CI) | 1.03 (0.51 - 2.07) | 0.88 (0.58 - 1.32) | 0.83 (0.52 - 1.32) | 0.89 (0.40 - 1.99) | Reference | 0.65 (0.30 - 1.38) | 0.76 (0.51 - 1.14) | 0.72 (0.45 - 1.15) | 0.51 (0.18 - 1.42) | Reference | 1.27 (0.77 - 2.10) | 0.64 (0.45 - 0.93) | 0.84 (0.57 - 1.25) | 1.34 (0.75 - 2.41) | Reference | 0.72 (0.35 - 1.49) | 0.84 (0.57 - 1.25) | 0.87 (0.56 - 1.37) | 1.05 (0.49 - 2.24) | Reference |
| Age, gender and nursing home adjusted HR (95% CI) | 1.03 (0.51 - 2.07) | 0.88 (0.58 - 1.32) | 0.83 (0.52 - 1.32) | 0.89 (0.40 - 1.99) | Reference | 0.65 (0.30 - 1.38) | 0.76 (0.51 - 1.14) | 0.72 (0.45 - 1.15) | 0.51 (0.18 - 1.42) | Reference | 1.27 (0.77 - 2.10) | 0.64 (0.45 - 0.93) | 0.84 (0.57 - 1.25) | 1.34 (0.75 - 2.41) | Reference | 0.72 (0.35 - 1.49) | 0.84 (0.57 - 1.25) | 0.87 (0.56 - 1.37) | 1.05 (0.49 - 2.24) | Reference |
| * HRs not reported because there were less than 10 events total (including reference group) | | | | | | | | | | | | | | | | | | | | |

| **Table S16:** Hazard ratios (HR) with 95% confidence intervals (95% CI) for non-COVID 19 and all-cause mortality according to number of SARS-CoV-2 vaccine doses for the period from 2021 to 2023 split into 3-month intervals for **40-59 year** old only. | | | | | | | | | | | | | | | | | | | | |
| --- | --- | --- | --- | --- | --- | --- | --- | --- | --- | --- | --- | --- | --- | --- | --- | --- | --- | --- | --- | --- |
|  | 2021 | | | | | | | | | | | | | | | | | | | |
|  | first quarter | | | | | second quarter | | | | | third quarter | | | | | fourth quarter | | | | |
|  | Four or more vaccine doses | Three vaccine doses | Two vaccine doses | One vaccine dose | Unvaccinated | Four or more vaccine doses | Three vaccine doses | Two vaccine doses | One vaccine dose | Unvaccinated | Four or more vaccine doses | Three vaccine doses | Two vaccine doses | One vaccine dose | Unvaccinated | Four or more vaccine doses | Three vaccine doses | Two vaccine doses | One vaccine dose | Unvaccinated |
|  | All-cause mortality | | | | | All-cause mortality | | | | | All-cause mortality | | | | | All-cause mortality | | | | |
| Deaths (n) |  |  | 2 |  | 23 |  |  | 9 | 4 | 47 |  |  | 36 | 20 | 68 |  | 7 | 36 | 22 | 44 |
| Events per 100,000 person days |  |  | 0.83 |  | 0.44 |  |  | 0.67 | 0.14 | 0.58 |  |  | 0.68 | 0.43 | 0.98 |  | 0.47 | 0.52 | 0.42 | 0.92 |
| Crude HR (95%CI) |  |  | 2.16 (0.50 - 9.34) |  | Reference |  |  | 1.10 (0.53 - 2.27) | 0.24 (0.09 - 0.67) | Reference |  |  | 0.69 (0.46 - 1.04) | 0.45 (0.27 - 0.74) | Reference |  | 0.51 (0.22 - 1.21) | 0.56 (0.36 - 0.87) | 0.46 (0.28 - 0.77) | Reference |
| Age adjusted HR (95% CI) |  |  | 2.09 (0.48 - 9.03) |  | Reference |  |  | 1.03 (0.50 - 2.13) | 0.23 (0.08 - 0.63) | Reference |  |  | 0.65 (0.43 - 0.97) | 0.42 (0.25 - 0.69) | Reference |  | 0.44 (0.19 - 1.04) | 0.51 (0.33 - 0.79) | 0.44 (0.26 - 0.74) | Reference |
| Age and gender adjusted HR (95% CI) |  |  | ( - ) |  | Reference |  |  | 1.08 (0.52 - 2.26) | 0.22 (0.08 - 0.63) | Reference |  |  | 0.64 (0.43 - 0.97) | 0.41 (0.25 - 0.68) | Reference |  | ( - ) | 0.50 (0.32 - 0.78) | 0.43 (0.26 - 0.73) | Reference |
| Age, gender and nursing home adjusted HR (95% CI) |  |  | 2.09 (0.48 - 9.03) |  | Reference |  |  | 1.08 (0.52 - 2.26) | 0.22 (0.08 - 0.63) | Reference |  |  | 0.64 (0.43 - 0.97) | 0.41 (0.25 - 0.68) | Reference |  | 0.44 (0.19 - 1.04) | 0.50 (0.32 - 0.78) | 0.43 (0.26 - 0.73) | Reference |
|  | Non-COVID-19 mortality | | | | | Non-COVID-19 mortality | | | | | Non-COVID-19 mortality | | | | | Non-COVID-19 mortality | | | | |
| Deaths (n) |  |  | 2 |  | 22 |  |  | 9 | 4 | 45 |  |  | 36 | 20 | 64 |  | 7 | 35 | 21 | 40 |
| Events per 100,000 person days |  |  | 0.83 |  | 0.42 |  |  | 0.67 | 0.14 | 0.55 |  |  | 0.68 | 0.43 | 0.92 |  | 0.47 | 0.5 | 0.4 | 0.84 |
| Crude HR (95%CI) |  |  | 2.32 (0.53 - 10.08) |  | Reference |  |  | 1.15 (0.55 - 2.39) | 0.25 (0.09 - 0.71) | Reference |  |  | 0.74 (0.49 - 1.12) | 0.48 (0.29 - 0.79) | Reference |  | 0.58 (0.24 - 1.37) | 0.60 (0.38 - 0.94) | 0.49 (0.29 - 0.83) | Reference |
| Age adjusted HR (95% CI) |  |  | 2.26 (0.52 - 9.80) |  | Reference |  |  | 1.08 (0.52 - 2.24) | 0.23 (0.08 - 0.66) | Reference |  |  | 0.69 (0.46 - 1.04) | 0.44 (0.27 - 0.74) | Reference |  | 0.50 (0.21 - 1.20) | 0.55 (0.35 - 0.86) | 0.47 (0.28 - 0.80) | Reference |
| Age and gender adjusted HR (95% CI) |  |  | ( - ) |  | Reference |  |  | 1.15 (0.55 - 2.39) | 0.23 (0.08 - 0.66) | Reference |  |  | 0.69 (0.45 - 1.04) | 0.44 (0.27 - 0.73) | Reference |  | ( - ) | 0.54 (0.34 - 0.85) | 0.46 (0.27 - 0.78) | Reference |
| Age, gender and nursing home adjusted HR (95% CI) |  |  | 2.26 (0.52 - 9.80) |  | Reference |  |  | 1.15 (0.55 - 2.39) | 0.23 (0.08 - 0.66) | Reference |  |  | 0.69 (0.45 - 1.04) | 0.44 (0.27 - 0.73) | Reference |  | 0.50 (0.21 - 1.20) | 0.54 (0.34 - 0.85) | 0.46 (0.27 - 0.78) | Reference |
|  | 2022 | | | | | | | | | | | | | | | | | | | |
|  | first quarter | | | | | second quarter | | | | | third quarter | | | | | fourth quarter | | | | |
|  | Four or more vaccine doses | Three vaccine doses | Two vaccine doses | One vaccine dose | Unvaccinated | Four or more vaccine doses | Three vaccine doses | Two vaccine doses | One vaccine dose | Unvaccinated | Four or more vaccine doses | Three vaccine doses | Two vaccine doses | One vaccine dose | Unvaccinated | Four or more vaccine doses | Three vaccine doses | Two vaccine doses | One vaccine dose | Unvaccinated |
|  | All-cause mortality | | | | | All-cause mortality | | | | | All-cause mortality | | | | | All-cause mortality | | | | |
| Deaths (n) |  | 37 | 43 | 19 | 39 | 3 | 76 | 64 | 23 | 73 | 9 | 220 | 120 | 20 | 103 | 59 | 233 | 107 | 24 | 144 |
| Events per 100,000 person days |  | 0.59 | 0.5 | 0.56 | 0.57 | 9.26 | 0.36 | 0.44 | 0.65 | 0.43 | 0.92 | 0.48 | 0.63 | 0.54 | 0.42 | 0.62 | 0.46 | 0.55 | 0.63 | 0.56 |
| Crude HR (95%CI) |  | 1.03 (0.66 - 1.62) | 0.87 (0.56 - 1.34) | 0.93 (0.53 - 1.62) | Reference | 22.37 (7.00 - 71.52) | 0.83 (0.60 - 1.15) | 1.03 (0.73 - 1.44) | 1.51 (0.94 - 2.42) | Reference | 2.22 (1.11 - 4.43) | 1.13 (0.90 - 1.43) | 1.48 (1.14 - 1.93) | 1.27 (0.79 - 2.06) | Reference | 1.15 (0.85 - 1.57) | 0.84 (0.68 - 1.03) | 0.99 (0.77 - 1.27) | 1.14 (0.74 - 1.76) | Reference |
| Age adjusted HR (95% CI) |  | 0.90 (0.57 - 1.42) | 0.80 (0.51 - 1.23) | 0.91 (0.52 - 1.59) | Reference | 20.02 (6.24 - 64.17) | 0.78 (0.56 - 1.07) | 1.01 (0.72 - 1.41) | 1.51 (0.94 - 2.42) | Reference | 1.77 (0.88 - 3.54) | 1.04 (0.82 - 1.31) | 1.50 (1.15 - 1.95) | 1.30 (0.81 - 2.10) | Reference | 0.96 (0.70 - 1.31) | 0.78 (0.63 - 0.96) | 1.01 (0.79 - 1.30) | 1.18 (0.76 - 1.81) | Reference |
| Age and gender adjusted HR (95% CI) |  | 0.89 (0.56 - 1.40) | 0.79 (0.51 - 1.22) | 0.90 (0.52 - 1.58) | Reference | 19.50 (6.08 - 62.54) | 0.77 (0.56 - 1.07) | 1.00 (0.71 - 1.39) | 1.50 (0.94 - 2.41) | Reference | 1.72 (0.86 - 3.45) | 1.03 (0.82 - 1.31) | 1.48 (1.14 - 1.93) | 1.29 (0.80 - 2.08) | Reference | 0.96 (0.70 - 1.30) | 0.78 (0.63 - 0.96) | 1.00 (0.78 - 1.28) | 1.17 (0.76 - 1.80) | Reference |
| Age, gender and nursing home adjusted HR (95% CI) |  | 0.89 (0.56 - 1.40) | 0.79 (0.51 - 1.22) | 0.90 (0.52 - 1.58) | Reference | 19.50 (6.08 - 62.54) | 0.77 (0.56 - 1.07) | 1.00 (0.71 - 1.39) | 1.50 (0.94 - 2.41) | Reference | 1.72 (0.86 - 3.45) | 1.03 (0.82 - 1.31) | 1.48 (1.14 - 1.93) | 1.29 (0.80 - 2.08) | Reference | 0.96 (0.70 - 1.30) | 0.78 (0.63 - 0.96) | 1.00 (0.78 - 1.28) | 1.17 (0.76 - 1.80) | Reference |

| **Table S16 continued:** | | | | | | | | | | | | | | | | | | | | |
| --- | --- | --- | --- | --- | --- | --- | --- | --- | --- | --- | --- | --- | --- | --- | --- | --- | --- | --- | --- | --- |
|  | 2022 | | | | | | | | | | | | | | | | | | | |
|  | first quarter | | | | | second quarter | | | | | third quarter | | | | | fourth quarter | | | | |
|  | Four or more vaccine doses | Three vaccine doses | Two vaccine doses | One vaccine dose | Unvacc | Four or more vaccine doses | Three vaccine doses | Two vaccine doses | One vaccine dose | Unvacc | Four or more vaccine doses | Three vaccine doses | Two vaccine doses | One vaccine dose | Unvacc | Four or more vaccine doses | Three vaccine doses | Two vaccine doses | One vaccine dose | Unvacc |
|  | Non-COVID-19 mortality | | | | | Non-COVID-19 mortality | | | | | Non-COVID-19 mortality | | | | | Non-COVID-19 mortality | | | | |
| Deaths (n) |  | 37 | 42 | 18 | 32 | 3 | 75 | 61 | 23 | 73 | 9 | 216 | 119 | 19 | 101 | 58 | 232 | 106 | 23 | 142 |
| Events per 100,000 person days |  | 0.59 | 0.49 | 0.53 | 0.47 | 9.26 | 0.35 | 0.42 | 0.65 | 0.43 | 0.92 | 0.47 | 0.62 | 0.51 | 0.42 | 0.61 | 0.46 | 0.54 | 0.61 | 0.55 |
| Crude HR (95%CI) |  | 1.25 (0.78 - 2.01) | 1.04 (0.65 - 1.64) | 1.05 (0.58 - 1.89) | Reference | 22.37 (7.00 - 71.52) | 0.82 (0.59 - 1.13) | 0.98 (0.70 - 1.38) | 1.51 (0.94 - 2.42) | Reference | 2.25 (1.13 - 4.50) | 1.13 (0.89 - 1.43) | 1.50 (1.15 - 1.95) | 1.23 (0.76 - 2.01) | Reference | 1.14 (0.84 - 1.56) | 0.85 (0.69 - 1.04) | 0.99 (0.77 - 1.27) | 1.11 (0.71 - 1.72) | Reference |
| Age adjusted HR (95% CI) |  | 1.11 (0.69 - 1.79) | 0.96 (0.60 - 1.52) | 1.03 (0.57 - 1.85) | Reference | 20.02 (6.24 - 64.17) | 0.76 (0.55 - 1.06) | 0.96 (0.69 - 1.36) | 1.51 (0.94 - 2.42) | Reference | 1.80 (0.90 - 3.61) | 1.04 (0.82 - 1.32) | 1.52 (1.17 - 1.98) | 1.26 (0.77 - 2.06) | Reference | 0.96 (0.70 - 1.31) | 0.79 (0.64 - 0.97) | 1.01 (0.79 - 1.30) | 1.14 (0.74 - 1.78) | Reference |
| Age and gender adjusted HR (95% CI) |  | 1.09 (0.68 - 1.76) | 0.95 (0.60 - 1.51) | 1.02 (0.57 - 1.84) | Reference | 19.50 (6.08 - 62.54) | 0.76 (0.55 - 1.05) | 0.95 (0.68 - 1.34) | 1.50 (0.94 - 2.41) | Reference | 1.75 (0.87 - 3.52) | 1.04 (0.82 - 1.31) | 1.50 (1.15 - 1.96) | 1.25 (0.77 - 2.04) | Reference | 0.95 (0.70 - 1.30) | 0.78 (0.64 - 0.97) | 1.00 (0.78 - 1.29) | 1.14 (0.73 - 1.76) | Reference |
| Age, gender and nursing home adjusted HR (95% CI) |  | 1.09 (0.68 - 1.76) | 0.95 (0.60 - 1.51) | 1.02 (0.57 - 1.84) | Reference | 19.50 (6.08 - 62.54) | 0.76 (0.55 - 1.05) | 0.95 (0.68 - 1.34) | 1.50 (0.94 - 2.41) | Reference | 1.75 (0.87 - 3.52) | 1.04 (0.82 - 1.31) | 1.50 (1.15 - 1.96) | 1.25 (0.77 - 2.04) | Reference | 0.95 (0.70 - 1.30) | 0.78 (0.64 - 0.97) | 1.00 (0.78 - 1.29) | 1.14 (0.73 - 1.76) | Reference |
|  | 2023 | | | | | | | | | | | | | | | | | | | |
|  | first quarter | | | | | second quarter | | | | | third quarter | | | | | fourth quarter | | | | |
|  | Four or more vaccine doses | Three vaccine doses | Two vaccine doses | One vaccine dose | Unvaccinated | Four or more vaccine doses | Three vaccine doses | Two vaccine doses | One vaccine dose | Unvaccinated | Four or more vaccine doses | Three vaccine doses | Two vaccine doses | One vaccine dose | Unvaccinated | Four or more vaccine doses | Three vaccine doses | Two vaccine doses | One vaccine dose | Unvaccinated |
|  | All-cause mortality | | | | | All-cause mortality | | | | | All-cause mortality | | | | | All-cause mortality | | | | |
| Deaths (n) | 105 | 252 | 104 | 7 | 138 | 106 | 267 | 104 | 22 | 142 | 130 | 296 | 117 | 19 | 175 | 128 | 259 | 115 | 29 | 165 |
| Events per 100,000 person days | 0.69 | 0.48 | 0.53 | 0.19 | 0.53 | 0.62 | 0.47 | 0.51 | 0.57 | 0.52 | 0.72 | 0.51 | 0.56 | 0.48 | 0.63 | 0.72 | 0.44 | 0.54 | 0.73 | 0.59 |
| Crude HR (95%CI) | 1.31 (1.02 - 1.69) | 0.90 (0.74 - 1.11) | 1.00 (0.78 - 1.30) | 0.36 (0.17 - 0.76) | Reference | 1.19 (0.92 - 1.53) | 0.90 (0.73 - 1.10) | 0.97 (0.75 - 1.25) | 1.09 (0.70 - 1.71) | Reference | 1.14 (0.91 - 1.43) | 0.80 (0.66 - 0.97) | 0.88 (0.70 - 1.11) | 0.77 (0.48 - 1.23) | Reference | 1.21 (0.96 - 1.52) | 0.75 (0.61 - 0.91) | 0.92 (0.72 - 1.16) | 1.24 (0.83 - 1.83) | Reference |
| Age adjusted HR (95% CI) | 1.10 (0.85 - 1.43) | 0.85 (0.69 - 1.05) | 1.03 (0.80 - 1.33) | 0.37 (0.17 - 0.78) | Reference | 1.02 (0.79 - 1.32) | 0.84 (0.69 - 1.03) | 0.99 (0.77 - 1.28) | 1.12 (0.71 - 1.75) | Reference | 0.99 (0.78 - 1.24) | 0.75 (0.62 - 0.91) | 0.90 (0.71 - 1.14) | 0.79 (0.49 - 1.26) | Reference | 1.01 (0.80 - 1.28) | 0.70 (0.57 - 0.85) | 0.94 (0.74 - 1.20) | 1.27 (0.86 - 1.89) | Reference |
| Age and gender adjusted HR (95% CI) | 1.10 (0.85 - 1.42) | 0.85 (0.69 - 1.05) | 1.01 (0.79 - 1.31) | 0.36 (0.17 - 0.78) | Reference | 1.02 (0.79 - 1.32) | 0.84 (0.69 - 1.04) | 0.98 (0.76 - 1.27) | 1.11 (0.71 - 1.74) | Reference | 0.99 (0.78 - 1.24) | 0.75 (0.62 - 0.91) | 0.90 (0.71 - 1.13) | 0.79 (0.49 - 1.26) | Reference | 1.01 (0.80 - 1.28) | 0.70 (0.57 - 0.85) | 0.93 (0.73 - 1.18) | 1.25 (0.84 - 1.86) | Reference |
| Age, gender and nursing home adjusted HR (95% CI) | 1.10 (0.85 - 1.42) | 0.85 (0.69 - 1.05) | 1.01 (0.79 - 1.31) | 0.36 (0.17 - 0.78) | Reference | 1.02 (0.79 - 1.32) | 0.84 (0.69 - 1.04) | 0.98 (0.76 - 1.27) | 1.11 (0.71 - 1.74) | Reference | 0.99 (0.78 - 1.24) | 0.75 (0.62 - 0.91) | 0.90 (0.71 - 1.13) | 0.79 (0.49 - 1.26) | Reference | 1.01 (0.80 - 1.28) | 0.70 (0.57 - 0.85) | 0.93 (0.73 - 1.18) | 1.25 (0.84 - 1.86) | Reference |
|  | Non-COVID-19 mortality | | | | | Non-COVID-19 mortality | | | | | Non-COVID-19 mortality | | | | | Non-COVID-19 mortality | | | | |
| Deaths (n) | 104 | 249 | 104 | 7 | 136 | 106 | 266 | 104 | 22 | 142 | 130 | 294 | 117 | 19 | 175 | 126 | 256 | 115 | 29 | 162 |
| Events per 100,000 person days | 0.68 | 0.47 | 0.53 | 0.19 | 0.52 | 0.62 | 0.47 | 0.51 | 0.57 | 0.52 | 0.72 | 0.5 | 0.56 | 0.48 | 0.63 | 0.71 | 0.44 | 0.54 | 0.73 | 0.58 |
| Crude HR (95%CI) | 1.32 (1.02 - 1.70) | 0.91 (0.74 - 1.12) | 1.02 (0.79 - 1.32) | 0.36 (0.17 - 0.77) | Reference | 1.19 (0.92 - 1.53) | 0.90 (0.73 - 1.10) | 0.97 (0.75 - 1.25) | 1.09 (0.70 - 1.71) | Reference | 1.14 (0.91 - 1.43) | 0.80 (0.66 - 0.96) | 0.88 (0.70 - 1.11) | 0.77 (0.48 - 1.23) | Reference | 1.21 (0.96 - 1.53) | 0.75 (0.62 - 0.92) | 0.93 (0.74 - 1.19) | 1.26 (0.85 - 1.87) | Reference |
| Age adjusted HR (95% CI) | 1.11 (0.86 - 1.44) | 0.86 (0.69 - 1.06) | 1.04 (0.81 - 1.35) | 0.37 (0.17 - 0.79) | Reference | 1.02 (0.79 - 1.32) | 0.84 (0.69 - 1.03) | 0.99 (0.77 - 1.28) | 1.12 (0.71 - 1.75) | Reference | 0.99 (0.78 - 1.24) | 0.75 (0.62 - 0.90) | 0.90 (0.71 - 1.14) | 0.79 (0.49 - 1.26) | Reference | 1.02 (0.80 - 1.29) | 0.70 (0.57 - 0.85) | 0.96 (0.76 - 1.22) | 1.29 (0.87 - 1.92) | Reference |
| Age and gender adjusted HR (95% CI) | 1.11 (0.86 - 1.43) | 0.86 (0.70 - 1.06) | 1.03 (0.80 - 1.33) | 0.37 (0.17 - 0.79) | Reference | 1.02 (0.79 - 1.32) | 0.84 (0.69 - 1.03) | 0.98 (0.76 - 1.27) | 1.11 (0.71 - 1.74) | Reference | 0.99 (0.78 - 1.24) | 0.75 (0.62 - 0.90) | 0.90 (0.71 - 1.13) | 0.79 (0.49 - 1.26) | Reference | 1.02 (0.80 - 1.29) | 0.70 (0.58 - 0.85) | 0.95 (0.74 - 1.20) | 1.27 (0.86 - 1.89) | Reference |
| Age, gender and nursing home adjusted HR (95% CI) | 1.11 (0.86 - 1.43) | 0.86 (0.70 - 1.06) | 1.03 (0.80 - 1.33) | 0.37 (0.17 - 0.79) | Reference | 1.02 (0.79 - 1.32) | 0.84 (0.69 - 1.03) | 0.98 (0.76 - 1.27) | 1.11 (0.71 - 1.74) | Reference | 0.99 (0.78 - 1.24) | 0.75 (0.62 - 0.90) | 0.90 (0.71 - 1.13) | 0.79 (0.49 - 1.26) | Reference | 1.02 (0.80 - 1.29) | 0.70 (0.58 - 0.85) | 0.95 (0.74 - 1.20) | 1.27 (0.86 - 1.89) | Reference |

| **Table S17:** Hazard ratios (HR) with 95% confidence intervals (95% CI) for non-COVID 19 and all-cause mortality according to number of SARS-CoV-2 vaccine doses for the period from 2021 to 2023 split into 3-month intervals for **60-74 year** old only. | | | | | | | | | | | | | | | | | | | | |
| --- | --- | --- | --- | --- | --- | --- | --- | --- | --- | --- | --- | --- | --- | --- | --- | --- | --- | --- | --- | --- |
|  | 2021 | | | | | | | | | | | | | | | | | | | |
|  | first quarter | | | | | second quarter | | | | | third quarter | | | | | fourth quarter | | | | |
|  | Four or more vaccine doses | Three vaccine doses | Two vaccine doses | One vaccine dose | Unvaccinated | Four or more vaccine doses | Three vaccine doses | Two vaccine doses | One vaccine dose | Unvaccinated | Four or more vaccine doses | Three vaccine doses | Two vaccine doses | One vaccine dose | Unvaccinated | Four or more vaccine doses | Three vaccine doses | Two vaccine doses | One vaccine dose | Unvaccinated |
|  | All-cause mortality | | | | | All-cause mortality | | | | | All-cause mortality | | | | | All-cause mortality | | | | |
| Deaths (n) |  |  | 12 | 11 | 91 |  |  | 63 | 34 | 158 |  | 3 | 116 | 48 | 113 |  | 44 | 91 | 61 | 91 |
| Events per 100,000 person days |  |  | 16.2 | 12.98 | 4.93 |  |  | 9.03 | 2.47 | 6.74 |  | 41.88 | 5.14 | 2.77 | 5.85 |  | 5.46 | 3.32 | 3.67 | 7.30 |
| Crude HR (95%CI) |  |  | 3.28 (1.78 - 6.02) | 2.65 (1.40 - 5.04) | Reference |  |  | 1.37 (1.01 - 1.87) | 0.35 (0.24 - 0.51) | Reference |  | 9.29 (2.75 - 31.40) | 0.90 (0.69 - 1.17) | 0.49 (0.35 - 0.68) | Reference |  | 0.69 (0.46 - 1.03) | 0.46 (0.34 - 0.61) | 0.51 (0.37 - 0.70) | Reference |
| Age adjusted HR (95% CI) |  |  | 2.89 (1.57 - 5.32) | 2.58 (1.36 - 4.91) | Reference |  |  | 1.12 (0.82 - 1.52) | 0.32 (0.22 - 0.47) | Reference |  | 6.80 (2.00 - 23.17) | 0.83 (0.64 - 1.08) | 0.46 (0.33 - 0.65) | Reference |  | 0.57 (0.38 - 0.86) | 0.42 (0.31 - 0.56) | 0.49 (0.35 - 0.68) | Reference |
| Age and gender adjusted HR (95% CI) |  |  | 2.88 (1.56 - 5.30) | 2.57 (1.35 - 4.89) | Reference |  |  | 1.10 (0.81 - 1.50) | 0.32 (0.22 - 0.46) | Reference |  | 6.42 (1.88 - 21.88) | 0.80 (0.61 - 1.04) | 0.46 (0.33 - 0.64) | Reference |  | 0.55 (0.36 - 0.83) | 0.41 (0.31 - 0.55) | 0.48 (0.35 - 0.67) | Reference |
| Age, gender and nursing home adjusted HR (95% CI) |  |  | 0.26 (0.13 - 0.53) | 1.02 (0.52 - 1.99) | Reference |  |  | 0.60 (0.43 - 0.84) | 0.34 (0.23 - 0.49) | Reference |  | 1.83 (0.50 - 6.79) | 0.67 (0.51 - 0.87) | 0.49 (0.35 - 0.69) | Reference |  | 0.37 (0.24 - 0.56) | 0.41 (0.30 - 0.55) | 0.52 (0.37 - 0.72) | Reference |
|  | Non-COVID-19 mortality | | | | | Non-COVID-19 mortality | | | | | Non-COVID-19 mortality | | | | | Non-COVID-19 mortality | | | | |
| Deaths (n) |  |  | 12 | 10 | 83 |  |  | 58 | 34 | 147 |  | 3 | 114 | 48 | 105 |  | 43 | 91 | 61 | 85 |
| Events per 100,000 person days |  |  | 16.2 | 11.8 | 4.49 |  |  | 8.32 | 2.47 | 6.27 |  | 41.88 | 5.05 | 2.77 | 5.43 |  | 5.34 | 3.32 | 3.67 | 6.82 |
| Crude HR (95%CI) |  |  | 3.54 (1.92 - 6.53) | 2.64 (1.35 - 5.17) | Reference |  |  | 1.31 (0.96 - 1.81) | 0.38 (0.26 - 0.55) | Reference |  | 9.33 (2.76 - 31.57) | 0.95 (0.73 - 1.24) | 0.52 (0.37 - 0.74) | Reference |  | 0.73 (0.48 - 1.10) | 0.49 (0.36 - 0.66) | 0.54 (0.39 - 0.76) | Reference |
| Age adjusted HR (95% CI) |  |  | 3.10 (1.68 - 5.73) | 2.57 (1.31 - 5.04) | Reference |  |  | 1.06 (0.77 - 1.46) | 0.34 (0.24 - 0.50) | Reference |  | 6.74 (1.98 - 22.98) | 0.87 (0.67 - 1.14) | 0.50 (0.35 - 0.70) | Reference |  | 0.60 (0.40 - 0.91) | 0.45 (0.33 - 0.61) | 0.52 (0.38 - 0.73) | Reference |
| Age and gender adjusted HR (95% CI) |  |  | 3.09 (1.67 - 5.71) | 2.55 (1.30 - 5.01) | Reference |  |  | 1.05 (0.76 - 1.44) | 0.34 (0.23 - 0.50) | Reference |  | 6.36 (1.86 - 21.69) | 0.84 (0.64 - 1.10) | 0.49 (0.35 - 0.69) | Reference |  | 0.58 (0.38 - 0.88) | 0.44 (0.33 - 0.59) | 0.52 (0.37 - 0.72) | Reference |
| Age, gender and nursing home adjusted HR (95% CI) |  |  | 0.30 (0.15 - 0.61) | 1.09 (0.54 - 2.20) | Reference |  |  | 0.57 (0.40 - 0.81) | 0.36 (0.25 - 0.53) | Reference |  | 1.72 (0.46 - 6.37) | 0.69 (0.53 - 0.91) | 0.53 (0.37 - 0.74) | Reference |  | 0.38 (0.25 - 0.58) | 0.44 (0.32 - 0.59) | 0.56 (0.40 - 0.78) | Reference |
|  | 2022 | | | | | | | | | | | | | | | | | | | |
|  | first quarter | | | | | second quarter | | | | | third quarter | | | | | fourth quarter | | | | |
|  | Four or more vaccine doses | Three vaccine doses | Two vaccine doses | One vaccine dose | Unvaccinated | Four or more vaccine doses | Three vaccine doses | Two vaccine doses | One vaccine dose | Unvaccinated | Four or more vaccine doses | Three vaccine doses | Two vaccine doses | One vaccine dose | Unvaccinated | Four or more vaccine doses | Three vaccine doses | Two vaccine doses | One vaccine dose | Unvaccinated |
|  | All-cause mortality | | | | | All-cause mortality | | | | | All-cause mortality | | | | | All-cause mortality | | | | |
| Deaths (n) |  | 110 | 121 | 31 | 82 | 4 | 301 | 189 | 27 | 120 | 70 | 589 | 198 | 42 | 187 | 312 | 638 | 191 | 34 | 205 |
| Events per 100,000 person days |  | 3.7 | 3.65 | 3.86 | 5.16 | 7.83 | 3.26 | 4.41 | 3.52 | 2.93 | 3.68 | 3 | 4 | 5.05 | 2.86 | 3.17 | 3.44 | 3.98 | 3.98 | 2.80 |
| Crude HR (95%CI) |  | 0.70 (0.53 - 0.93) | 0.70 (0.53 - 0.93) | 0.70 (0.46 - 1.07) | Reference | 2.97 (1.09 - 8.11) | 1.13 (0.92 - 1.40) | 1.52 (1.21 - 1.91) | 1.18 (0.78 - 1.80) | Reference | 1.33 (1.00 - 1.77) | 1.06 (0.90 - 1.25) | 1.41 (1.16 - 1.73) | 1.78 (1.28 - 2.49) | Reference | 1.12 (0.94 - 1.33) | 1.25 (1.07 - 1.46) | 1.43 (1.18 - 1.74) | 1.43 (0.99 - 2.05) | Reference |
| Age adjusted HR (95% CI) |  | 0.61 (0.45 - 0.81) | 0.63 (0.48 - 0.84) | 0.68 (0.45 - 1.04) | Reference | 1.97 (0.72 - 5.40) | 0.98 (0.79 - 1.21) | 1.39 (1.10 - 1.75) | 1.14 (0.75 - 1.74) | Reference | 0.99 (0.74 - 1.32) | 0.95 (0.81 - 1.12) | 1.36 (1.12 - 1.67) | 1.75 (1.25 - 2.44) | Reference | 0.91 (0.76 - 1.10) | 1.18 (1.00 - 1.38) | 1.42 (1.16 - 1.73) | 1.42 (0.99 - 2.04) | Reference |
| Age and gender adjusted HR (95% CI) |  | 0.59 (0.44 - 0.78) | 0.62 (0.47 - 0.82) | 0.68 (0.45 - 1.03) | Reference | 1.85 (0.67 - 5.07) | 0.94 (0.76 - 1.17) | 1.35 (1.07 - 1.70) | 1.12 (0.74 - 1.71) | Reference | 0.95 (0.71 - 1.27) | 0.93 (0.79 - 1.09) | 1.33 (1.09 - 1.63) | 1.72 (1.23 - 2.40) | Reference | 0.87 (0.73 - 1.05) | 1.15 (0.98 - 1.34) | 1.38 (1.13 - 1.68) | 1.39 (0.97 - 2.00) | Reference |
| Age, gender and nursing home adjusted HR (95% CI) |  | 0.49 (0.36 - 0.65) | 0.63 (0.47 - 0.83) | 0.69 (0.46 - 1.05) | Reference | 0.88 (0.31 - 2.50) | 0.80 (0.65 - 1.00) | 1.26 (1.00 - 1.59) | 1.06 (0.70 - 1.61) | Reference | 0.70 (0.52 - 0.96) | 0.87 (0.73 - 1.02) | 1.26 (1.03 - 1.55) | 1.60 (1.15 - 2.24) | Reference | 0.73 (0.60 - 0.88) | 1.11 (0.95 - 1.30) | 1.33 (1.09 - 1.62) | 1.32 (0.91 - 1.89) | Reference |

| **Table S17 continued:** | | | | | | | | | | | | | | | | | | | | |
| --- | --- | --- | --- | --- | --- | --- | --- | --- | --- | --- | --- | --- | --- | --- | --- | --- | --- | --- | --- | --- |
|  | 2022 | | | | | | | | | | | | | | | | | | | |
|  | first quarter | | | | | second quarter | | | | | third quarter | | | | | fourth quarter | | | | |
|  | Four or more vaccine doses | Three vaccine doses | Two vaccine doses | One vaccine dose | Unvacc | Four or more vaccine doses | Three vaccine doses | Two vaccine doses | One vaccine dose | Unvacc | Four or more vaccine doses | Three vaccine doses | Two vaccine doses | One vaccine dose | Unvacc | Four or more vaccine doses | Three vaccine doses | Two vaccine doses | One vaccine dose | Unvacc |
|  | Non-COVID-19 mortality | | | | | Non-COVID-19 mortality | | | | | Non-COVID-19 mortality | | | | | Non-COVID-19 mortality | | | | |
| Deaths (n) |  | 107 | 114 | 31 | 75 | 4 | 295 | 183 | 26 | 112 | 69 | 578 | 195 | 42 | 183 | 306 | 628 | 189 | 34 | 200 |
| Events per 100,000 person days |  | 3.6 | 3.44 | 3.86 | 4.72 | 7.83 | 3.19 | 4.27 | 3.39 | 2.73 | 3.63 | 2.94 | 3.94 | 5.05 | 2.79 | 3.11 | 3.39 | 3.94 | 3.98 | 2.73 |
| Crude HR (95%CI) |  | 0.74 (0.55 - 1.00) | 0.72 (0.54 - 0.97) | 0.77 (0.50 - 1.17) | Reference | 3.13 (1.15 - 8.56) | 1.19 (0.96 - 1.48) | 1.58 (1.25 - 2.00) | 1.23 (0.80 - 1.88) | Reference | 1.33 (1.00 - 1.77) | 1.06 (0.90 - 1.25) | 1.42 (1.16 - 1.74) | 1.82 (1.30 - 2.55) | Reference | 1.12 (0.94 - 1.34) | 1.26 (1.08 - 1.48) | 1.45 (1.19 - 1.77) | 1.46 (1.02 - 2.10) | Reference |
| Age adjusted HR (95% CI) |  | 0.65 (0.48 - 0.87) | 0.65 (0.49 - 0.87) | 0.75 (0.49 - 1.14) | Reference | 2.12 (0.77 - 5.82) | 1.03 (0.83 - 1.29) | 1.45 (1.14 - 1.84) | 1.19 (0.77 - 1.82) | Reference | 1.00 (0.74 - 1.34) | 0.96 (0.81 - 1.13) | 1.37 (1.12 - 1.68) | 1.79 (1.28 - 2.50) | Reference | 0.92 (0.77 - 1.10) | 1.19 (1.01 - 1.39) | 1.44 (1.18 - 1.75) | 1.46 (1.01 - 2.10) | Reference |
| Age and gender adjusted HR (95% CI) |  | 0.62 (0.46 - 0.84) | 0.64 (0.48 - 0.86) | 0.74 (0.48 - 1.13) | Reference | 2.01 (0.73 - 5.52) | 1.00 (0.80 - 1.24) | 1.41 (1.11 - 1.79) | 1.17 (0.76 - 1.80) | Reference | 0.96 (0.72 - 1.29) | 0.93 (0.79 - 1.10) | 1.34 (1.10 - 1.64) | 1.76 (1.26 - 2.46) | Reference | 0.88 (0.73 - 1.05) | 1.16 (0.99 - 1.36) | 1.40 (1.15 - 1.71) | 1.42 (0.99 - 2.05) | Reference |
| Age, gender and nursing home adjusted HR (95% CI) |  | 0.52 (0.38 - 0.71) | 0.65 (0.48 - 0.87) | 0.76 (0.50 - 1.16) | Reference | 0.95 (0.33 - 2.71) | 0.85 (0.68 - 1.06) | 1.32 (1.04 - 1.67) | 1.10 (0.72 - 1.70) | Reference | 0.72 (0.53 - 0.98) | 0.87 (0.74 - 1.03) | 1.28 (1.04 - 1.56) | 1.64 (1.17 - 2.30) | Reference | 0.74 (0.61 - 0.89) | 1.13 (0.96 - 1.32) | 1.35 (1.11 - 1.65) | 1.35 (0.94 - 1.94) | Reference |
|  | 2023 | | | | | | | | | | | | | | | | | | | |
|  | first quarter | | | | | second quarter | | | | | third quarter | | | | | fourth quarter | | | | |
|  | Four or more vaccine doses | Three vaccine doses | Two vaccine doses | One vaccine dose | Unvaccinated | Four or more vaccine doses | Three vaccine doses | Two vaccine doses | One vaccine dose | Unvaccinated | Four or more vaccine doses | Three vaccine doses | Two vaccine doses | One vaccine dose | Unvaccinated | Four or more vaccine doses | Three vaccine doses | Two vaccine doses | One vaccine dose | Unvaccinated |
|  | All-cause mortality | | | | | All-cause mortality | | | | | All-cause mortality | | | | | All-cause mortality | | | | |
| Deaths (n) | 490 | 649 | 175 | 28 | 230 | 592 | 594 | 185 | 30 | 260 | 590 | 624 | 175 | 27 | 239 | 587 | 628 | 173 | 28 | 262 |
| Events per 100,000 person days | 3.36 | 3.37 | 3.64 | 3.26 | 2.98 | 3.53 | 2.8 | 3.57 | 3.27 | 3.13 | 3.3 | 2.78 | 3.21 | 2.79 | 2.75 | 3.28 | 2.76 | 3.12 | 2.85 | 2.95 |
| Crude HR (95%CI) | 1.13 (0.97 - 1.32) | 1.14 (0.98 - 1.32) | 1.22 (1.00 - 1.49) | 1.09 (0.74 - 1.61) | Reference | 1.13 (0.98 - 1.31) | 0.90 (0.78 - 1.04) | 1.14 (0.95 - 1.38) | 1.05 (0.72 - 1.53) | Reference | 1.20 (1.03 - 1.39) | 1.01 (0.87 - 1.18) | 1.18 (0.97 - 1.43) | 1.02 (0.69 - 1.52) | Reference | 1.11 (0.96 - 1.28) | 0.93 (0.81 - 1.08) | 1.06 (0.87 - 1.28) | 0.97 (0.65 - 1.43) | Reference |
| Age adjusted HR (95% CI) | 0.96 (0.82 - 1.12) | 1.07 (0.92 - 1.25) | 1.22 (1.01 - 1.49) | 1.10 (0.74 - 1.63) | Reference | 0.96 (0.82 - 1.11) | 0.85 (0.74 - 0.99) | 1.15 (0.95 - 1.39) | 1.06 (0.73 - 1.55) | Reference | 1.04 (0.89 - 1.21) | 0.97 (0.83 - 1.12) | 1.19 (0.98 - 1.44) | 1.04 (0.70 - 1.54) | Reference | 0.94 (0.82 - 1.10) | 0.90 (0.78 - 1.04) | 1.07 (0.89 - 1.30) | 0.98 (0.66 - 1.45) | Reference |
| Age and gender adjusted HR (95% CI) | 0.91 (0.78 - 1.07) | 1.05 (0.90 - 1.22) | 1.20 (0.99 - 1.46) | 1.08 (0.73 - 1.60) | Reference | 0.91 (0.78 - 1.06) | 0.84 (0.72 - 0.97) | 1.13 (0.93 - 1.36) | 1.03 (0.71 - 1.51) | Reference | 1.00 (0.86 - 1.17) | 0.95 (0.82 - 1.10) | 1.16 (0.96 - 1.41) | 1.01 (0.68 - 1.51) | Reference | 0.91 (0.78 - 1.05) | 0.88 (0.77 - 1.02) | 1.05 (0.87 - 1.27) | 0.96 (0.65 - 1.42) | Reference |
| Age, gender and nursing home adjusted HR (95% CI) | 0.81 (0.69 - 0.95) | 1.02 (0.88 - 1.19) | 1.16 (0.95 - 1.42) | 1.05 (0.71 - 1.55) | Reference | 0.83 (0.72 - 0.97) | 0.82 (0.71 - 0.95) | 1.10 (0.91 - 1.33) | 1.01 (0.69 - 1.48) | Reference | 0.94 (0.81 - 1.10) | 0.93 (0.81 - 1.09) | 1.15 (0.94 - 1.39) | 1.00 (0.67 - 1.50) | Reference | 0.83 (0.72 - 0.97) | 0.87 (0.76 - 1.01) | 1.03 (0.85 - 1.24) | 0.95 (0.64 - 1.40) | Reference |
|  | Non-COVID-19 mortality | | | | | Non-COVID-19 mortality | | | | | Non-COVID-19 mortality | | | | | Non-COVID-19 mortality | | | | |
| Deaths (n) | 484 | 640 | 173 | 28 | 228 | 587 | 589 | 183 | 29 | 258 | 589 | 621 | 175 | 27 | 238 | 571 | 617 | 173 | 28 | 255 |
| Events per 100,000 person days | 3.32 | 3.33 | 3.6 | 3.26 | 2.95 | 3.5 | 2.78 | 3.53 | 3.16 | 3.11 | 3.29 | 2.77 | 3.21 | 2.79 | 2.73 | 3.19 | 2.71 | 3.12 | 2.85 | 2.87 |
| Crude HR (95%CI) | 1.13 (0.96 - 1.32) | 1.13 (0.97 - 1.32) | 1.22 (1.00 - 1.48) | 1.10 (0.74 - 1.63) | Reference | 1.13 (0.97 - 1.31) | 0.90 (0.77 - 1.04) | 1.14 (0.94 - 1.38) | 1.02 (0.69 - 1.50) | Reference | 1.20 (1.03 - 1.40) | 1.01 (0.87 - 1.18) | 1.18 (0.97 - 1.44) | 1.03 (0.69 - 1.53) | Reference | 1.11 (0.96 - 1.28) | 0.94 (0.81 - 1.09) | 1.09 (0.90 - 1.32) | 0.99 (0.67 - 1.47) | Reference |
| Age adjusted HR (95% CI) | 0.95 (0.81 - 1.12) | 1.07 (0.92 - 1.24) | 1.22 (1.00 - 1.49) | 1.11 (0.75 - 1.64) | Reference | 0.96 (0.82 - 1.11) | 0.85 (0.74 - 0.99) | 1.15 (0.95 - 1.39) | 1.03 (0.70 - 1.52) | Reference | 1.04 (0.89 - 1.21) | 0.97 (0.83 - 1.12) | 1.19 (0.98 - 1.45) | 1.04 (0.70 - 1.55) | Reference | 0.95 (0.82 - 1.10) | 0.91 (0.79 - 1.05) | 1.10 (0.91 - 1.34) | 1.01 (0.68 - 1.49) | Reference |
| Age and gender adjusted HR (95% CI) | 0.91 (0.78 - 1.07) | 1.04 (0.90 - 1.21) | 1.20 (0.98 - 1.46) | 1.09 (0.74 - 1.61) | Reference | 0.91 (0.78 - 1.06) | 0.84 (0.72 - 0.97) | 1.12 (0.93 - 1.36) | 1.01 (0.69 - 1.48) | Reference | 1.00 (0.86 - 1.17) | 0.95 (0.82 - 1.10) | 1.17 (0.96 - 1.42) | 1.02 (0.68 - 1.52) | Reference | 0.91 (0.78 - 1.06) | 0.89 (0.77 - 1.03) | 1.08 (0.89 - 1.31) | 0.98 (0.67 - 1.45) | Reference |
| Age, gender and nursing home adjusted HR (95% CI) | 0.81 (0.69 - 0.95) | 1.02 (0.88 - 1.18) | 1.16 (0.95 - 1.42) | 1.06 (0.72 - 1.57) | Reference | 0.83 (0.72 - 0.97) | 0.82 (0.71 - 0.95) | 1.10 (0.91 - 1.32) | 0.99 (0.67 - 1.45) | Reference | 0.94 (0.81 - 1.10) | 0.93 (0.80 - 1.09) | 1.15 (0.95 - 1.40) | 1.01 (0.68 - 1.50) | Reference | 0.84 (0.72 - 0.98) | 0.88 (0.76 - 1.02) | 1.05 (0.87 - 1.28) | 0.97 (0.66 - 1.44) | Reference |

| **Table S18:** Hazard ratios (HR) with 95% confidence intervals (95% CI) for non-COVID 19 and all-cause mortality according to number of SARS-CoV-2 vaccine doses for the period from 2021 to 2023 split into 3-month intervals for **75-84+ year** old only. | | | | | | | | | | | | | | | | | | | | |
| --- | --- | --- | --- | --- | --- | --- | --- | --- | --- | --- | --- | --- | --- | --- | --- | --- | --- | --- | --- | --- |
|  | 2021 | | | | | | | | | | | | | | | | | | | |
|  | first quarter | | | | | second quarter | | | | | third quarter | | | | | fourth quarter | | | | |
|  | Four or more vaccine doses | Three vaccine doses | Two vaccine doses | One vaccine dose | Unvaccinated | Four or more vaccine doses | Three vaccine doses | Two vaccine doses | One vaccine dose | Unvaccinated | Four or more vaccine doses | Three vaccine doses | Two vaccine doses | One vaccine dose | Unvaccinated | Four or more vaccine doses | Three vaccine doses | Two vaccine doses | One vaccine dose | Unvaccinated |
|  | All-cause mortality | | | | | All-cause mortality | | | | | All-cause mortality | | | | | All-cause mortality | | | | |
| Deaths (n) |  |  | 43 | 18 | 125 |  |  | 157 | 62 | 207 |  | 4 | 205 | 104 | 171 |  | 119 | 163 | 79 | 92 |
| Events per 100,000 person days |  |  | 36.23 | 19.74 | 20.69 |  |  | 23.8 | 13.78 | 29.06 |  | 33.55 | 19.12 | 17.1 | 29.68 |  | 21.72 | 16.02 | 15.62 | 26.46 |
| Crude HR (95%CI) |  |  | 1.74 (1.21 - 2.49) | 0.99 (0.60 - 1.64) | Reference |  |  | 0.83 (0.67 - 1.02) | 0.48 (0.36 - 0.64) | Reference |  | 1.06 (0.37 - 3.06) | 0.63 (0.51 - 0.77) | 0.57 (0.45 - 0.73) | Reference |  | 0.91 (0.67 - 1.22) | 0.60 (0.47 - 0.78) | 0.60 (0.44 - 0.81) | Reference |
| Age adjusted HR (95% CI) |  |  | 1.43 (0.99 - 2.07) | 0.80 (0.48 - 1.34) | Reference |  |  | 0.78 (0.63 - 0.97) | 0.50 (0.38 - 0.67) | Reference |  | 1.01 (0.35 - 2.91) | 0.62 (0.50 - 0.76) | 0.58 (0.45 - 0.74) | Reference |  | 0.86 (0.64 - 1.16) | 0.61 (0.47 - 0.79) | 0.62 (0.46 - 0.83) | Reference |
| Age and gender adjusted HR (95% CI) | |  | 1.48 (1.02 - 2.14) | 0.80 (0.48 - 1.33) | Reference |  |  | 0.78 (0.63 - 0.96) | 0.49 (0.37 - 0.65) | Reference |  | 1.01 (0.35 - 2.91) | 0.61 (0.50 - 0.75) | 0.57 (0.45 - 0.73) | Reference |  | 0.86 (0.64 - 1.15) | 0.61 (0.47 - 0.79) | 0.61 (0.45 - 0.82) | Reference |
| Age, gender and nursing home adjusted HR (95% CI) |  |  | 0.62 (0.40 - 0.96) | 0.67 (0.40 - 1.10) | Reference |  |  | 0.55 (0.44 - 0.69) | 0.51 (0.39 - 0.68) | Reference |  | 0.52 (0.18 - 1.54) | 0.49 (0.40 - 0.61) | 0.61 (0.48 - 0.78) | Reference |  | 0.55 (0.41 - 0.75) | 0.60 (0.46 - 0.77) | 0.71 (0.52 - 0.96) | Reference |
|  | Non-COVID-19 mortality | | | | | Non-COVID-19 mortality | | | | | Non-COVID-19 mortality | | | | | Non-COVID-19 mortality | | | | |
| Deaths (n) |  |  | 41 | 17 | 114 |  |  | 151 | 60 | 193 |  | 4 | 201 | 100 | 160 |  | 117 | 156 | 75 | 89 |
| Events per 100,000 person days |  |  | 34.55 | 18.65 | 18.87 |  |  | 22.89 | 13.34 | 27.10 |  | 33.55 | 18.75 | 16.45 | 27.77 |  | 21.36 | 15.34 | 14.82 | 25.59 |
| Crude HR (95%CI) |  |  | 1.79 (1.24 - 2.60) | 1.03 (0.61 - 1.73) | Reference |  |  | 0.86 (0.69 - 1.06) | 0.50 (0.37 - 0.67) | Reference |  | 1.20 (0.41 - 3.51) | 0.66 (0.54 - 0.81) | 0.59 (0.46 - 0.76) | Reference |  | 0.92 (0.68 - 1.24) | 0.60 (0.46 - 0.77) | 0.59 (0.43 - 0.80) | Reference |
| Age adjusted HR (95% CI) |  |  | 1.49 (1.02 - 2.18) | 0.83 (0.49 - 1.40) | Reference |  |  | 0.81 (0.65 - 1.01) | 0.52 (0.39 - 0.69) | Reference |  | 1.15 (0.39 - 3.37) | 0.65 (0.53 - 0.80) | 0.60 (0.46 - 0.77) | Reference |  | 0.88 (0.65 - 1.18) | 0.60 (0.46 - 0.78) | 0.61 (0.45 - 0.82) | Reference |
| Age and gender adjusted HR (95% CI) | |  | 1.53 (1.05 - 2.24) | 0.83 (0.49 - 1.40) | Reference |  |  | 0.81 (0.65 - 1.00) | 0.50 (0.38 - 0.68) | Reference |  | 1.15 (0.39 - 3.37) | 0.64 (0.52 - 0.79) | 0.59 (0.46 - 0.76) | Reference |  | 0.87 (0.64 - 1.18) | 0.60 (0.47 - 0.79) | 0.60 (0.44 - 0.81) | Reference |
| Age, gender and nursing home adjusted HR (95% CI) |  |  | 0.69 (0.44 - 1.08) | 0.69 (0.41 - 1.16) | Reference |  |  | 0.56 (0.45 - 0.70) | 0.53 (0.40 - 0.71) | Reference |  | 0.57 (0.19 - 1.71) | 0.52 (0.42 - 0.64) | 0.63 (0.49 - 0.81) | Reference |  | 0.56 (0.42 - 0.76) | 0.59 (0.46 - 0.77) | 0.70 (0.51 - 0.96) | Reference |
|  | 2022 | | | | | | | | | | | | | | | | | | | |
|  | first quarter | | | | | second quarter | | | | | third quarter | | | | | fourth quarter | | | | |
|  | Four or more vaccine doses | Three vaccine doses | Two vaccine doses | One vaccine dose | Unvaccinated | Four or more vaccine doses | Three vaccine doses | Two vaccine doses | One vaccine dose | Unvaccinated | Four or more vaccine doses | Three vaccine doses | Two vaccine doses | One vaccine dose | Unvaccinated | Four or more vaccine doses | Three vaccine doses | Two vaccine doses | One vaccine dose | Unvaccinated |
|  | All-cause mortality | | | | | All-cause mortality | | | | | All-cause mortality | | | | | All-cause mortality | | | | |
| Deaths (n) | 3 | 235 | 200 | 52 | 77 | 16 | 543 | 236 | 40 | 103 | 212 | 975 | 225 | 46 | 172 | 754 | 1018 | 225 | 39 | 210 |
| Events per 100,000 person days | 106.61 | 16.94 | 17.64 | 25.64 | 23.08 | 22.51 | 15.38 | 19.12 | 22.58 | 14.76 | 12.67 | 14.66 | 18.38 | 24.41 | 15.24 | 13.56 | 18.53 | 20.87 | 20.73 | 16.09 |
| Crude HR (95%CI) | 4.56 (1.44 - 14.50) | 0.73 (0.56 - 0.94) | 0.76 (0.58 - 0.99) | 1.09 (0.76 - 1.56) | Reference | 1.87 (1.08 - 3.23) | 1.07 (0.86 - 1.32) | 1.31 (1.04 - 1.65) | 1.48 (1.03 - 2.14) | Reference | 0.86 (0.70 - 1.06) | 0.97 (0.82 - 1.14) | 1.21 (0.99 - 1.47) | 1.61 (1.17 - 2.23) | Reference | 0.83 (0.71 - 0.97) | 1.17 (1.01 - 1.36) | 1.31 (1.09 - 1.58) | 1.28 (0.91 - 1.81) | Reference |
| Age adjusted HR (95% CI) | 4.50 (1.42 - 14.29) | 0.70 (0.54 - 0.91) | 0.76 (0.59 - 1.00) | 1.09 (0.76 - 1.56) | Reference | 1.60 (0.92 - 2.77) | 1.04 (0.84 - 1.28) | 1.29 (1.02 - 1.63) | 1.44 (1.00 - 2.08) | Reference | 0.80 (0.65 - 0.99) | 0.95 (0.81 - 1.12) | 1.19 (0.97 - 1.45) | 1.57 (1.14 - 2.18) | Reference | 0.79 (0.68 - 0.93) | 1.16 (1.00 - 1.35) | 1.30 (1.08 - 1.57) | 1.26 (0.89 - 1.77) | Reference |
| Age and gender adjusted HR (95% CI) | 4.31 (1.35 - 13.71) | 0.69 (0.53 - 0.89) | 0.75 (0.57 - 0.97) | 1.08 (0.76 - 1.54) | Reference | 1.62 (0.93 - 2.82) | 1.01 (0.82 - 1.25) | 1.27 (1.00 - 1.60) | 1.44 (1.00 - 2.08) | Reference | 0.79 (0.64 - 0.97) | 0.92 (0.78 - 1.08) | 1.17 (0.96 - 1.43) | 1.56 (1.13 - 2.17) | Reference | 0.78 (0.67 - 0.91) | 1.13 (0.97 - 1.31) | 1.29 (1.07 - 1.55) | 1.25 (0.89 - 1.76) | Reference |
| Age, gender and nursing home adjusted HR (95% CI) | 2.77 (0.86 - 8.90) | 0.53 (0.41 - 0.69) | 0.79 (0.60 - 1.02) | 1.19 (0.83 - 1.70) | Reference | 1.13 (0.64 - 1.99) | 0.83 (0.67 - 1.03) | 1.20 (0.95 - 1.52) | 1.42 (0.98 - 2.05) | Reference | 0.59 (0.47 - 0.72) | 0.83 (0.71 - 0.98) | 1.08 (0.89 - 1.32) | 1.51 (1.09 - 2.09) | Reference | 0.63 (0.54 - 0.74) | 1.08 (0.93 - 1.25) | 1.22 (1.01 - 1.47) | 1.19 (0.84 - 1.67) | Reference |

| **Table S18 continued:** | | | | | | | | | | | | | | | | | | | | |
| --- | --- | --- | --- | --- | --- | --- | --- | --- | --- | --- | --- | --- | --- | --- | --- | --- | --- | --- | --- | --- |
|  | 2022 | | | | | | | | | | | | | | | | | | | |
|  | first quarter | | | | | second quarter | | | | | third quarter | | | | | fourth quarter | | | | |
|  | Four or more vaccine doses | Three vaccine doses | Two vaccine doses | One vaccine dose | Unvacc | Four or more vaccine doses | Three vaccine doses | Two vaccine doses | One vaccine dose | Unvacc | Four or more vaccine doses | Three vaccine doses | Two vaccine doses | One vaccine dose | Unvacc | Four or more vaccine doses | Three vaccine doses | Two vaccine doses | One vaccine dose | Unvacc |
|  | Non-COVID-19 mortality | | | | | Non-COVID-19 mortality | | | | | Non-COVID-19 mortality | | | | | Non-COVID-19 mortality | | | | |
| Deaths (n) | 3 | 224 | 188 | 50 | 64 | 15 | 532 | 229 | 37 | 101 | 206 | 955 | 213 | 44 | 164 | 739 | 978 | 219 | 38 | 206 |
| Events per 100,000 person days | 106.61 | 16.15 | 16.58 | 24.66 | 19.18 | 21.1 | 15.07 | 18.55 | 20.89 | 14.47 | 12.32 | 14.36 | 17.4 | 23.35 | 14.53 | 13.29 | 17.8 | 20.31 | 20.2 | 15.78 |
| Crude HR (95%CI) | 5.65 (1.77 - 18.05) | 0.83 (0.63 - 1.10) | 0.86 (0.65 - 1.14) | 1.23 (0.85 - 1.79) | Reference | 1.75 (1.00 - 3.08) | 1.06 (0.86 - 1.32) | 1.30 (1.03 - 1.65) | 1.42 (0.97 - 2.07) | Reference | 0.88 (0.71 - 1.09) | 1.00 (0.84 - 1.17) | 1.20 (0.98 - 1.47) | 1.62 (1.16 - 2.26) | Reference | 0.83 (0.71 - 0.97) | 1.15 (0.99 - 1.34) | 1.30 (1.08 - 1.57) | 1.28 (0.90 - 1.80) | Reference |
| Age adjusted HR (95% CI) | 5.57 (1.74 - 17.79) | 0.80 (0.61 - 1.06) | 0.87 (0.65 - 1.15) | 1.23 (0.84 - 1.79) | Reference | 1.50 (0.85 - 2.64) | 1.03 (0.84 - 1.28) | 1.29 (1.02 - 1.63) | 1.38 (0.94 - 2.01) | Reference | 0.82 (0.66 - 1.01) | 0.98 (0.83 - 1.15) | 1.18 (0.96 - 1.44) | 1.57 (1.13 - 2.19) | Reference | 0.79 (0.68 - 0.93) | 1.14 (0.98 - 1.32) | 1.29 (1.07 - 1.56) | 1.25 (0.88 - 1.77) | Reference |
| Age and gender adjusted HR (95% CI) | 5.29 (1.65 - 16.95) | 0.78 (0.59 - 1.04) | 0.84 (0.63 - 1.12) | 1.22 (0.84 - 1.77) | Reference | 1.53 (0.86 - 2.70) | 1.01 (0.81 - 1.25) | 1.26 (1.00 - 1.60) | 1.38 (0.94 - 2.01) | Reference | 0.80 (0.65 - 1.00) | 0.94 (0.80 - 1.11) | 1.16 (0.95 - 1.43) | 1.56 (1.12 - 2.18) | Reference | 0.78 (0.67 - 0.91) | 1.11 (0.95 - 1.29) | 1.28 (1.06 - 1.55) | 1.24 (0.88 - 1.76) | Reference |
| Age, gender and nursing home adjusted HR (95% CI) | 3.50 (1.08 - 11.32) | 0.61 (0.46 - 0.80) | 0.89 (0.67 - 1.18) | 1.34 (0.92 - 1.95) | Reference | 1.08 (0.61 - 1.93) | 0.83 (0.67 - 1.03) | 1.20 (0.95 - 1.52) | 1.36 (0.93 - 1.98) | Reference | 0.60 (0.48 - 0.75) | 0.85 (0.72 - 1.01) | 1.07 (0.87 - 1.32) | 1.51 (1.08 - 2.10) | Reference | 0.63 (0.54 - 0.74) | 1.06 (0.91 - 1.23) | 1.21 (1.00 - 1.46) | 1.18 (0.83 - 1.67) | Reference |
|  | 2023 | | | | | | | | | | | | | | | | | | | |
|  | first quarter | | | | | second quarter | | | | | third quarter | | | | | fourth quarter | | | | |
|  | Four or more vaccine doses | Three vaccine doses | Two vaccine doses | One vaccine dose | Unvaccinated | Four or more vaccine doses | Three vaccine doses | Two vaccine doses | One vaccine dose | Unvaccinated | Four or more vaccine doses | Three vaccine doses | Two vaccine doses | One vaccine dose | Unvaccinated | Four or more vaccine doses | Three vaccine doses | Two vaccine doses | One vaccine dose | Unvaccinated |
|  | All-cause mortality | | | | | All-cause mortality | | | | | All-cause mortality | | | | | All-cause mortality | | | | |
| Deaths (n) | 1028 | 976 | 220 | 40 | 262 | 1050 | 868 | 230 | 60 | 234 | 1173 | 834 | 187 | 41 | 222 | 1381 | 1006 | 177 | 38 | 209 |
| Events per 100,000 person days | 13.26 | 17.43 | 20.55 | 21.28 | 18.35 | 11.61 | 14.16 | 20.2 | 30.31 | 15.09 | 12.1 | 12.99 | 15.83 | 20.22 | 13.63 | 14.08 | 15.44 | 14.79 | 18.71 | 12.56 |
| Crude HR (95%CI) | 0.72 (0.63 - 0.83) | 0.96 (0.83 - 1.10) | 1.12 (0.94 - 1.34) | 1.15 (0.83 - 1.61) | Reference | 0.77 (0.67 - 0.89) | 0.94 (0.81 - 1.09) | 1.34 (1.12 - 1.61) | 2.02 (1.52 - 2.68) | Reference | 0.88 (0.77 - 1.02) | 0.95 (0.82 - 1.10) | 1.16 (0.96 - 1.41) | 1.49 (1.07 - 2.08) | Reference | 1.11 (0.96 - 1.29) | 1.22 (1.05 - 1.42) | 1.17 (0.96 - 1.43) | 1.49 (1.05 - 2.10) | Reference |
| Age adjusted HR (95% CI) | 0.69 (0.61 - 0.80) | 0.95 (0.82 - 1.08) | 1.11 (0.93 - 1.33) | 1.13 (0.81 - 1.58) | Reference | 0.74 (0.64 - 0.85) | 0.93 (0.81 - 1.07) | 1.33 (1.11 - 1.60) | 2.00 (1.50 - 2.65) | Reference | 0.85 (0.73 - 0.98) | 0.94 (0.81 - 1.09) | 1.15 (0.95 - 1.40) | 1.47 (1.05 - 2.05) | Reference | 1.06 (0.91 - 1.22) | 1.20 (1.03 - 1.39) | 1.16 (0.95 - 1.42) | 1.47 (1.04 - 2.07) | Reference |
| Age and gender adjusted HR (95% CI) | 0.67 (0.59 - 0.77) | 0.92 (0.81 - 1.06) | 1.10 (0.92 - 1.32) | 1.13 (0.81 - 1.57) | Reference | 0.71 (0.62 - 0.82) | 0.91 (0.79 - 1.05) | 1.32 (1.10 - 1.59) | 1.99 (1.50 - 2.64) | Reference | 0.82 (0.71 - 0.95) | 0.92 (0.79 - 1.07) | 1.15 (0.94 - 1.39) | 1.47 (1.05 - 2.05) | Reference | 1.01 (0.88 - 1.17) | 1.17 (1.01 - 1.36) | 1.16 (0.95 - 1.41) | 1.46 (1.04 - 2.07) | Reference |
| Age, gender and nursing home adjusted HR (95% CI) | 0.58 (0.51 - 0.67) | 0.90 (0.79 - 1.03) | 1.07 (0.89 - 1.28) | 1.06 (0.76 - 1.48) | Reference | 0.65 (0.56 - 0.75) | 0.90 (0.77 - 1.03) | 1.28 (1.07 - 1.54) | 1.88 (1.42 - 2.50) | Reference | 0.75 (0.65 - 0.87) | 0.91 (0.78 - 1.05) | 1.11 (0.91 - 1.35) | 1.38 (0.99 - 1.92) | Reference | 0.93 (0.81 - 1.08) | 1.16 (1.00 - 1.34) | 1.13 (0.92 - 1.37) | 1.36 (0.96 - 1.92) | Reference |
|  | Non-COVID-19 mortality | | | | | Non-COVID-19 mortality | | | | | Non-COVID-19 mortality | | | | | Non-COVID-19 mortality | | | | |
| Deaths (n) | 996 | 949 | 212 | 40 | 256 | 1039 | 856 | 223 | 59 | 231 | 1170 | 826 | 187 | 39 | 218 | 1334 | 969 | 168 | 38 | 201 |
| Events per 100,000 person days | 12.85 | 16.95 | 19.8 | 21.28 | 17.93 | 11.49 | 13.96 | 19.59 | 29.81 | 14.89 | 12.07 | 12.87 | 15.83 | 19.24 | 13.38 | 13.6 | 14.88 | 14.04 | 18.71 | 12.08 |
| Crude HR (95%CI) | 0.72 (0.63 - 0.82) | 0.95 (0.83 - 1.09) | 1.10 (0.92 - 1.33) | 1.18 (0.85 - 1.65) | Reference | 0.77 (0.67 - 0.89) | 0.94 (0.81 - 1.09) | 1.32 (1.09 - 1.58) | 2.01 (1.51 - 2.68) | Reference | 0.90 (0.78 - 1.04) | 0.96 (0.83 - 1.11) | 1.18 (0.97 - 1.44) | 1.44 (1.03 - 2.03) | Reference | 1.12 (0.96 - 1.30) | 1.22 (1.05 - 1.42) | 1.16 (0.94 - 1.42) | 1.54 (1.09 - 2.19) | Reference |
| Age adjusted HR (95% CI) | 0.69 (0.60 - 0.79) | 0.94 (0.82 - 1.08) | 1.10 (0.91 - 1.31) | 1.16 (0.83 - 1.62) | Reference | 0.74 (0.64 - 0.86) | 0.93 (0.80 - 1.07) | 1.31 (1.09 - 1.57) | 1.99 (1.50 - 2.65) | Reference | 0.86 (0.74 - 0.99) | 0.95 (0.81 - 1.10) | 1.17 (0.97 - 1.43) | 1.42 (1.01 - 2.00) | Reference | 1.06 (0.91 - 1.23) | 1.20 (1.03 - 1.40) | 1.15 (0.94 - 1.41) | 1.53 (1.08 - 2.16) | Reference |
| Age and gender adjusted HR (95% CI) | 0.66 (0.58 - 0.76) | 0.92 (0.80 - 1.06) | 1.09 (0.91 - 1.31) | 1.15 (0.83 - 1.61) | Reference | 0.72 (0.62 - 0.83) | 0.91 (0.78 - 1.05) | 1.30 (1.08 - 1.56) | 1.98 (1.49 - 2.64) | Reference | 0.83 (0.72 - 0.96) | 0.93 (0.80 - 1.08) | 1.17 (0.96 - 1.42) | 1.42 (1.01 - 2.00) | Reference | 1.02 (0.88 - 1.18) | 1.17 (1.01 - 1.37) | 1.14 (0.93 - 1.40) | 1.52 (1.08 - 2.15) | Reference |
| Age, gender and nursing home adjusted HR (95% CI) | 0.58 (0.50 - 0.66) | 0.90 (0.78 - 1.03) | 1.05 (0.88 - 1.26) | 1.09 (0.78 - 1.52) | Reference | 0.65 (0.56 - 0.75) | 0.89 (0.77 - 1.03) | 1.26 (1.05 - 1.51) | 1.88 (1.41 - 2.50) | Reference | 0.76 (0.66 - 0.88) | 0.92 (0.79 - 1.06) | 1.13 (0.93 - 1.37) | 1.34 (0.95 - 1.88) | Reference | 0.94 (0.81 - 1.09) | 1.16 (1.00 - 1.35) | 1.11 (0.90 - 1.36) | 1.41 (1.00 - 2.00) | Reference |

| **Table S19:** Hazard ratios (HR) with 95% confidence intervals (95% CI) for non-COVID 19 and all-cause mortality according to number of SARS-CoV-2 vaccine doses for the period from 2021 to 2023 split into 3-month intervals for **85+ year** old only. | | | | | | | | | | | | | | | | | | | | |
| --- | --- | --- | --- | --- | --- | --- | --- | --- | --- | --- | --- | --- | --- | --- | --- | --- | --- | --- | --- | --- |
|  | 2021 | | | | | | | | | | | | | | | | | | | |
|  | first quarter | | | | | second quarter | | | | | third quarter | | | | | fourth quarter | | | | |
|  | Four or more vaccine doses | Three vaccine doses | Two vaccine doses | One vaccine dose | Unvaccinated | Four or more vaccine doses | Three vaccine doses | Two vaccine doses | One vaccine dose | Unvaccinated | Four or more vaccine doses | Three vaccine doses | Two vaccine doses | One vaccine dose | Unvaccinated | Four or more vaccine doses | Three vaccine doses | Two vaccine doses | One vaccine dose | Unvaccinated |
|  | All-cause mortality | | | | | All-cause mortality | | | | | All-cause mortality | | | | | All-cause mortality | | | | |
| Deaths (n) |  |  | 89 | 41 | 158 |  |  | 325 | 85 | 283 |  | 12 | 353 | 97 | 219 |  | 244 | 299 | 118 | 190 |
| Events per 100,000 person days |  |  | 53.77 | 66.56 | 77.90 |  |  | 59.12 | 64.83 | 82.60 |  | 67.08 | 54.54 | 45.61 | 72.87 |  | 60.45 | 71.46 | 65.45 | 93.10 |
| Crude HR (95%CI) |  |  | 0.68 (0.52 - 0.89) | 0.92 (0.65 - 1.31) | Reference |  |  | 0.70 (0.60 - 0.83) | 0.80 (0.63 - 1.03) | Reference |  | 0.91 (0.48 - 1.73) | 0.75 (0.63 - 0.88) | 0.63 (0.50 - 0.80) | Reference |  | 0.65 (0.54 - 0.79) | 0.80 (0.66 - 0.96) | 0.72 (0.58 - 0.91) | Reference |
| Age adjusted HR (95% CI) |  |  | 0.65 (0.49 - 0.85) | 0.92 (0.65 - 1.30) | Reference |  |  | 0.70 (0.59 - 0.82) | 0.82 (0.65 - 1.05) | Reference |  | 0.86 (0.46 - 1.64) | 0.75 (0.63 - 0.89) | 0.66 (0.52 - 0.84) | Reference |  | 0.66 (0.54 - 0.81) | 0.84 (0.70 - 1.01) | 0.78 (0.62 - 0.98) | Reference |
| Age and gender adjusted HR (95% CI) | |  | 0.66 (0.50 - 0.87) | 0.91 (0.65 - 1.30) | Reference |  |  | 0.70 (0.60 - 0.82) | 0.82 (0.64 - 1.05) | Reference |  | 0.87 (0.46 - 1.65) | 0.75 (0.63 - 0.89) | 0.66 (0.52 - 0.84) | Reference |  | 0.66 (0.55 - 0.81) | 0.84 (0.70 - 1.01) | 0.77 (0.61 - 0.97) | Reference |
| Age, gender and nursing home adjusted HR (95% CI) |  |  | 0.47 (0.35 - 0.63) | 0.79 (0.55 - 1.12) | Reference |  |  | 0.58 (0.49 - 0.68) | 0.79 (0.62 - 1.01) | Reference |  | 0.72 (0.38 - 1.38) | 0.65 (0.55 - 0.78) | 0.66 (0.52 - 0.83) | Reference |  | 0.53 (0.43 - 0.65) | 0.75 (0.63 - 0.91) | 0.78 (0.62 - 0.99) | Reference |
|  | Non-COVID-19 mortality | | | | | Non-COVID-19 mortality | | | | | Non-COVID-19 mortality | | | | | Non-COVID-19 mortality | | | | |
| Deaths (n) |  |  | 84 | 36 | 146 |  |  | 307 | 82 | 275 |  | 12 | 348 | 95 | 214 |  | 242 | 293 | 116 | 176 |
| Events per 100,000 person days |  |  | 50.75 | 58.44 | 71.98 |  |  | 55.85 | 62.54 | 80.27 |  | 67.08 | 53.77 | 44.67 | 71.21 |  | 59.95 | 70.03 | 64.34 | 86.24 |
| Crude HR (95%CI) |  |  | 0.69 (0.52 - 0.91) | 0.88 (0.61 - 1.28) | Reference |  |  | 0.68 (0.58 - 0.81) | 0.79 (0.62 - 1.02) | Reference |  | 0.91 (0.48 - 1.73) | 0.75 (0.63 - 0.89) | 0.63 (0.50 - 0.80) | Reference |  | 0.71 (0.58 - 0.87) | 0.84 (0.70 - 1.02) | 0.77 (0.61 - 0.97) | Reference |
| Age adjusted HR (95% CI) |  |  | 0.66 (0.50 - 0.87) | 0.88 (0.60 - 1.27) | Reference |  |  | 0.67 (0.57 - 0.79) | 0.81 (0.63 - 1.04) | Reference |  | 0.87 (0.46 - 1.65) | 0.76 (0.64 - 0.90) | 0.66 (0.52 - 0.84) | Reference |  | 0.72 (0.59 - 0.88) | 0.89 (0.74 - 1.07) | 0.82 (0.65 - 1.04) | Reference |
| Age and gender adjusted HR (95% CI) | |  | 0.67 (0.51 - 0.89) | 0.88 (0.60 - 1.27) | Reference |  |  | 0.68 (0.58 - 0.80) | 0.81 (0.63 - 1.04) | Reference |  | 0.87 (0.46 - 1.65) | 0.76 (0.64 - 0.90) | 0.66 (0.52 - 0.84) | Reference |  | 0.72 (0.59 - 0.88) | 0.89 (0.74 - 1.07) | 0.81 (0.64 - 1.02) | Reference |
| Age, gender and nursing home adjusted HR (95% CI) |  |  | 0.49 (0.36 - 0.66) | 0.76 (0.52 - 1.10) | Reference |  |  | 0.56 (0.47 - 0.67) | 0.78 (0.61 - 1.00) | Reference |  | 0.71 (0.37 - 1.35) | 0.65 (0.55 - 0.78) | 0.66 (0.51 - 0.84) | Reference |  | 0.57 (0.46 - 0.70) | 0.79 (0.66 - 0.96) | 0.83 (0.65 - 1.05) | Reference |
|  | 2022 | | | | | | | | | | | | | | | | | | | |
|  | first quarter | | | | | second quarter | | | | | third quarter | | | | | fourth quarter | | | | |
|  | Four or more vaccine doses | Three vaccine doses | Two vaccine doses | One vaccine dose | Unvaccinated | Four or more vaccine doses | Three vaccine doses | Two vaccine doses | One vaccine dose | Unvaccinated | Four or more vaccine doses | Three vaccine doses | Two vaccine doses | One vaccine dose | Unvaccinated | Four or more vaccine doses | Three vaccine doses | Two vaccine doses | One vaccine dose | Unvaccinated |
|  | All-cause mortality | | | | | All-cause mortality | | | | | All-cause mortality | | | | | All-cause mortality | | | | |
| Deaths (n) | 3 | 428 | 284 | 59 | 133 | 15 | 771 | 298 | 41 | 209 | 357 | 1367 | 341 | 62 | 279 | 1204 | 1278 | 315 | 65 | 315 |
| Events per 100,000 person days | 215.67 | 62.72 | 70.74 | 66.89 | 78.85 | 36.32 | 56.68 | 64.58 | 50.63 | 70.55 | 50.06 | 60.06 | 70.73 | 70.15 | 61.85 | 60.13 | 75.28 | 78.76 | 80.05 | 62.80 |
| Crude HR (95%CI) | 3.13 (0.99 - 9.87) | 0.79 (0.65 - 0.96) | 0.90 (0.74 - 1.11) | 0.84 (0.62 - 1.15) | Reference | 0.52 (0.31 - 0.89) | 0.81 (0.69 - 0.94) | 0.91 (0.76 - 1.09) | 0.72 (0.51 - 1.01) | Reference | 0.84 (0.71 - 0.98) | 0.99 (0.87 - 1.12) | 1.15 (0.98 - 1.35) | 1.14 (0.86 - 1.50) | Reference | 0.95 (0.84 - 1.08) | 1.24 (1.09 - 1.40) | 1.27 (1.08 - 1.48) | 1.26 (0.96 - 1.64) | Reference |
| Age adjusted HR (95% CI) | 3.29 (1.04 - 10.40) | 0.82 (0.68 - 1.00) | 0.98 (0.79 - 1.20) | 0.90 (0.66 - 1.23) | Reference | 0.54 (0.31 - 0.92) | 0.84 (0.72 - 0.98) | 0.96 (0.81 - 1.15) | 0.75 (0.54 - 1.05) | Reference | 0.86 (0.73 - 1.01) | 1.05 (0.92 - 1.19) | 1.18 (1.01 - 1.38) | 1.16 (0.88 - 1.53) | Reference | 1.01 (0.90 - 1.15) | 1.34 (1.19 - 1.52) | 1.31 (1.12 - 1.53) | 1.32 (1.01 - 1.72) | Reference |
| Age and gender adjusted HR (95% CI) | ( - ) | 0.82 (0.68 - 1.00) | 0.97 (0.79 - 1.19) | 0.89 (0.65 - 1.21) | Reference | 0.53 (0.31 - 0.91) | 0.84 (0.72 - 0.98) | 0.95 (0.80 - 1.14) | 0.75 (0.53 - 1.05) | Reference | 0.87 (0.74 - 1.02) | 1.04 (0.91 - 1.18) | 1.17 (1.00 - 1.38) | 1.17 (0.89 - 1.54) | Reference | 1.02 (0.90 - 1.15) | 1.33 (1.17 - 1.50) | 1.30 (1.11 - 1.52) | 1.31 (1.01 - 1.72) | Reference |
| Age, gender and nursing home adjusted HR (95% CI) | 2.59 (0.82 - 8.24) | 0.67 (0.55 - 0.81) | 0.92 (0.75 - 1.14) | 0.93 (0.68 - 1.27) | Reference | 0.43 (0.25 - 0.73) | 0.66 (0.57 - 0.78) | 0.88 (0.74 - 1.06) | 0.74 (0.53 - 1.03) | Reference | 0.63 (0.53 - 0.74) | 0.85 (0.75 - 0.97) | 1.09 (0.93 - 1.28) | 1.12 (0.85 - 1.48) | Reference | 0.78 (0.68 - 0.88) | 1.18 (1.04 - 1.34) | 1.26 (1.08 - 1.47) | 1.30 (1.00 - 1.70) | Reference |

| **Table S19 continued:** | | | | | | | | | | | | | | | | | | | | |
| --- | --- | --- | --- | --- | --- | --- | --- | --- | --- | --- | --- | --- | --- | --- | --- | --- | --- | --- | --- | --- |
|  | 2022 | | | | | | | | | | | | | | | | | | | |
|  | first quarter | | | | | second quarter | | | | | third quarter | | | | | fourth quarter | | | | |
|  | Four or more vaccine doses | Three vaccine doses | Two vaccine doses | One vaccine dose | Unvacc | Four or more vaccine doses | Three vaccine doses | Two vaccine doses | One vaccine dose | Unvacc | Four or more vaccine doses | Three vaccine doses | Two vaccine doses | One vaccine dose | Unvacc | Four or more vaccine doses | Three vaccine doses | Two vaccine doses | One vaccine dose | Unvacc |
|  | Non-COVID-19 mortality | | | | | Non-COVID-19 mortality | | | | | Non-COVID-19 mortality | | | | | Non-COVID-19 mortality | | | | |
| Deaths (n) | 1 | 403 | 267 | 58 | 121 | 15 | 749 | 284 | 40 | 203 | 354 | 1333 | 332 | 59 | 274 | 1176 | 1249 | 308 | 65 | 305 |
| Events per 100,000 person days | 71.89 | 59.06 | 66.51 | 65.76 | 71.74 | 36.32 | 55.07 | 61.54 | 49.39 | 68.52 | 49.64 | 58.57 | 68.86 | 66.76 | 60.74 | 58.73 | 73.57 | 77.01 | 80.05 | 60.81 |
| Crude HR (95%CI) | 1.17 (0.16 - 8.37) | 0.82 (0.67 - 1.00) | 0.93 (0.75 - 1.16) | 0.90 (0.66 - 1.24) | Reference | 0.54 (0.32 - 0.92) | 0.81 (0.69 - 0.94) | 0.90 (0.75 - 1.08) | 0.72 (0.51 - 1.01) | Reference | 0.84 (0.72 - 0.99) | 0.98 (0.86 - 1.12) | 1.14 (0.97 - 1.34) | 1.10 (0.83 - 1.46) | Reference | 0.96 (0.85 - 1.09) | 1.25 (1.10 - 1.41) | 1.28 (1.09 - 1.50) | 1.30 (0.99 - 1.70) | Reference |
| Age adjusted HR (95% CI) | 1.23 (0.17 - 8.87) | 0.85 (0.69 - 1.04) | 1.01 (0.81 - 1.25) | 0.98 (0.71 - 1.34) | Reference | 0.55 (0.32 - 0.94) | 0.84 (0.72 - 0.99) | 0.95 (0.79 - 1.14) | 0.76 (0.54 - 1.06) | Reference | 0.87 (0.74 - 1.02) | 1.04 (0.91 - 1.19) | 1.17 (0.99 - 1.37) | 1.13 (0.85 - 1.49) | Reference | 1.02 (0.90 - 1.16) | 1.36 (1.20 - 1.54) | 1.32 (1.13 - 1.55) | 1.36 (1.04 - 1.78) | Reference |
| Age and gender adjusted HR (95% CI) | ( - ) | 0.85 (0.69 - 1.04) | 1.00 (0.80 - 1.24) | 0.96 (0.70 - 1.32) | Reference | 0.55 (0.32 - 0.94) | 0.84 (0.72 - 0.98) | 0.94 (0.78 - 1.13) | 0.75 (0.54 - 1.06) | Reference | 0.88 (0.75 - 1.03) | 1.03 (0.91 - 1.18) | 1.16 (0.99 - 1.36) | 1.13 (0.85 - 1.50) | Reference | 1.03 (0.90 - 1.16) | 1.34 (1.18 - 1.52) | 1.32 (1.12 - 1.54) | 1.36 (1.04 - 1.78) | Reference |
| Age, gender and nursing home adjusted HR (95% CI) | 1.00 (0.14 - 7.19) | 0.69 (0.56 - 0.85) | 0.96 (0.77 - 1.19) | 1.00 (0.73 - 1.37) | Reference | 0.44 (0.25 - 0.75) | 0.66 (0.57 - 0.78) | 0.87 (0.73 - 1.05) | 0.74 (0.53 - 1.04) | Reference | 0.63 (0.53 - 0.75) | 0.85 (0.74 - 0.97) | 1.08 (0.92 - 1.27) | 1.09 (0.82 - 1.44) | Reference | 0.79 (0.69 - 0.90) | 1.20 (1.05 - 1.36) | 1.27 (1.09 - 1.49) | 1.35 (1.03 - 1.76) | Reference |
|  | 2023 | | | | | | | | | | | | | | | | | | | |
|  | first quarter | | | | | second quarter | | | | | third quarter | | | | | fourth quarter | | | | |
|  | Four or more vaccine doses | Three vaccine doses | Two vaccine doses | One vaccine dose | Unvaccinated | Four or more vaccine doses | Three vaccine doses | Two vaccine doses | One vaccine dose | Unvaccinated | Four or more vaccine doses | Three vaccine doses | Two vaccine doses | One vaccine dose | Unvaccinated | Four or more vaccine doses | Three vaccine doses | Two vaccine doses | One vaccine dose | Unvaccinated |
|  | All-cause mortality | | | | | All-cause mortality | | | | | All-cause mortality | | | | | All-cause mortality | | | | |
| Deaths (n) | 1552 | 1130 | 301 | 53 | 378 | 1611 | 990 | 251 | 49 | 332 | 1579 | 1013 | 246 | 48 | 294 | 1846 | 1130 | 272 | 49 | 363 |
| Events per 100,000 person days | 60.42 | 66.17 | 77.19 | 63.39 | 69.68 | 53.67 | 51.96 | 60.23 | 56.79 | 56.09 | 48.38 | 50.23 | 57.1 | 54.57 | 47.78 | 56.51 | 56.19 | 63.94 | 57.54 | 58.98 |
| Crude HR (95%CI) | 0.87 (0.77 - 0.97) | 0.95 (0.84 - 1.06) | 1.10 (0.95 - 1.28) | 0.92 (0.69 - 1.23) | Reference | 0.96 (0.85 - 1.07) | 0.93 (0.82 - 1.05) | 1.07 (0.91 - 1.26) | 1.01 (0.75 - 1.36) | Reference | 1.02 (0.90 - 1.16) | 1.06 (0.93 - 1.20) | 1.20 (1.01 - 1.42) | 1.15 (0.85 - 1.56) | Reference | 0.96 (0.86 - 1.07) | 0.95 (0.85 - 1.07) | 1.09 (0.93 - 1.27) | 0.96 (0.71 - 1.29) | Reference |
| Age adjusted HR (95% CI) | 0.94 (0.84 - 1.05) | 1.04 (0.92 - 1.17) | 1.13 (0.97 - 1.32) | 0.96 (0.72 - 1.28) | Reference | 1.03 (0.91 - 1.16) | 1.02 (0.90 - 1.15) | 1.10 (0.93 - 1.29) | 1.05 (0.78 - 1.42) | Reference | 1.10 (0.97 - 1.25) | 1.14 (1.00 - 1.30) | 1.22 (1.03 - 1.45) | 1.18 (0.87 - 1.60) | Reference | 1.05 (0.94 - 1.17) | 1.03 (0.92 - 1.16) | 1.12 (0.95 - 1.31) | 0.98 (0.73 - 1.33) | Reference |
| Age and gender adjusted HR (95% CI) | 0.94 (0.84 - 1.05) | 1.03 (0.91 - 1.15) | 1.12 (0.97 - 1.31) | 0.95 (0.72 - 1.27) | Reference | 1.01 (0.90 - 1.14) | 1.00 (0.88 - 1.14) | 1.09 (0.92 - 1.28) | 1.04 (0.77 - 1.41) | Reference | 1.10 (0.97 - 1.24) | 1.12 (0.98 - 1.28) | 1.21 (1.02 - 1.43) | 1.18 (0.87 - 1.60) | Reference | 1.05 (0.94 - 1.18) | 1.02 (0.91 - 1.15) | 1.11 (0.95 - 1.30) | 0.98 (0.73 - 1.32) | Reference |
| Age, gender and nursing home adjusted HR (95% CI) | 0.73 (0.65 - 0.82) | 0.94 (0.84 - 1.06) | 1.08 (0.93 - 1.26) | 0.95 (0.71 - 1.26) | Reference | 0.81 (0.72 - 0.91) | 0.93 (0.82 - 1.05) | 1.05 (0.89 - 1.24) | 1.04 (0.77 - 1.40) | Reference | 0.89 (0.78 - 1.01) | 1.05 (0.92 - 1.20) | 1.16 (0.98 - 1.38) | 1.17 (0.86 - 1.59) | Reference | 0.85 (0.76 - 0.96) | 0.98 (0.87 - 1.10) | 1.08 (0.92 - 1.27) | 0.98 (0.73 - 1.32) | Reference |
|  | Non-COVID-19 mortality | | | | | Non-COVID-19 mortality | | | | | Non-COVID-19 mortality | | | | | Non-COVID-19 mortality | | | | |
| Deaths (n) | 1493 | 1104 | 292 | 53 | 361 | 1594 | 983 | 251 | 49 | 327 | 1572 | 1008 | 243 | 48 | 293 | 1783 | 1081 | 258 | 46 | 354 |
| Events per 100,000 person days | 58.12 | 64.65 | 74.89 | 63.39 | 66.55 | 53.1 | 51.59 | 60.23 | 56.79 | 55.25 | 48.17 | 49.98 | 56.4 | 54.57 | 47.62 | 54.59 | 53.76 | 60.65 | 54.01 | 57.52 |
| Crude HR (95%CI) | 0.87 (0.78 - 0.98) | 0.97 (0.86 - 1.09) | 1.12 (0.96 - 1.30) | 0.97 (0.73 - 1.29) | Reference | 0.96 (0.85 - 1.08) | 0.93 (0.82 - 1.06) | 1.09 (0.92 - 1.28) | 1.03 (0.76 - 1.38) | Reference | 1.02 (0.90 - 1.15) | 1.06 (0.93 - 1.20) | 1.19 (1.00 - 1.41) | 1.16 (0.85 - 1.57) | Reference | 0.95 (0.85 - 1.07) | 0.93 (0.83 - 1.05) | 1.06 (0.90 - 1.24) | 0.92 (0.68 - 1.25) | Reference |
| Age adjusted HR (95% CI) | 0.95 (0.84 - 1.06) | 1.06 (0.94 - 1.20) | 1.15 (0.99 - 1.34) | 1.00 (0.75 - 1.34) | Reference | 1.03 (0.92 - 1.16) | 1.02 (0.90 - 1.16) | 1.11 (0.94 - 1.31) | 1.07 (0.79 - 1.44) | Reference | 1.10 (0.97 - 1.25) | 1.14 (1.00 - 1.30) | 1.21 (1.02 - 1.44) | 1.19 (0.87 - 1.61) | Reference | 1.04 (0.93 - 1.17) | 1.01 (0.90 - 1.14) | 1.08 (0.92 - 1.27) | 0.95 (0.70 - 1.29) | Reference |
| Age and gender adjusted HR (95% CI) | 0.94 (0.84 - 1.06) | 1.05 (0.93 - 1.19) | 1.14 (0.98 - 1.33) | 1.00 (0.75 - 1.33) | Reference | 1.02 (0.90 - 1.15) | 1.01 (0.89 - 1.15) | 1.10 (0.94 - 1.30) | 1.06 (0.79 - 1.43) | Reference | 1.09 (0.97 - 1.24) | 1.12 (0.98 - 1.28) | 1.20 (1.01 - 1.42) | 1.18 (0.87 - 1.60) | Reference | 1.04 (0.93 - 1.17) | 1.01 (0.89 - 1.14) | 1.08 (0.92 - 1.27) | 0.95 (0.70 - 1.29) | Reference |
| Age, gender and nursing home adjusted HR (95% CI) | 0.74 (0.66 - 0.83) | 0.96 (0.86 - 1.09) | 1.10 (0.94 - 1.28) | 0.99 (0.74 - 1.32) | Reference | 0.82 (0.72 - 0.92) | 0.94 (0.83 - 1.06) | 1.07 (0.91 - 1.26) | 1.05 (0.78 - 1.42) | Reference | 0.88 (0.78 - 1.00) | 1.05 (0.92 - 1.19) | 1.15 (0.97 - 1.37) | 1.18 (0.87 - 1.59) | Reference | 0.85 (0.76 - 0.96) | 0.96 (0.85 - 1.08) | 1.05 (0.89 - 1.23) | 0.94 (0.69 - 1.28) | Reference |

| **Table S20:** Hazard ratios (HR) with 95% confidence intervals (95% CI) for non-COVID 19 and all-cause mortality according to number of SARS-CoV-2 vaccine doses for the period from 2021 to 2023 split into 3-month intervals for **nursing home residents** only. | | | | | | | | | | | | | | | | | | | | |
| --- | --- | --- | --- | --- | --- | --- | --- | --- | --- | --- | --- | --- | --- | --- | --- | --- | --- | --- | --- | --- |
|  | 2021 | | | | | | | | | | | | | | | | | | | |
|  | first quarter |  |  |  | 0 | second quarter | | | | | third quarter |  |  |  | 0 | fourth quarter | | | | |
|  | Four or more vaccine doses | Three vaccine doses | Two vaccine doses | One vaccine dose | Unvaccinated | Four or more vaccine doses | Three vaccine doses | Two vaccine doses | One vaccine dose | Unvaccinated | Four or more vaccine doses | Three vaccine doses | Two vaccine doses | One vaccine dose | Unvaccinated | Four or more vaccine doses | Three vaccine doses | Two vaccine doses | One vaccine dose | Unvaccinated |
|  | All-cause mortality | | | | | All-cause mortality | | | | | All-cause mortality | | | | | All-cause mortality | | | | |
| Deaths (n) |  |  | 122 | 48 | 142 |  |  | 424 | 81 | 229 |  | 17 | 421 | 91 | 161 |  | 315 | 290 | 112 | 131 |
| Events per 100,000 person days | |  | 48.75 | 86.14 | 113.81 |  |  | 61.18 | 78.98 | 93.88 |  | 59.28 | 57.22 | 69.08 | 82.68 |  | 65.31 | 79.89 | 115.39 | 101.42 |
| Crude HR (95%CI) |  |  | 0.41 (0.32 - 0.53) | 0.88 (0.62 - 1.24) | Reference |  |  | 0.64 (0.55 - 0.76) | 0.86 (0.66 - 1.11) | Reference |  | 0.78 (0.42 - 1.43) | 0.69 (0.58 - 0.83) | 0.84 (0.65 - 1.09) | Reference |  | 0.61 (0.49 - 0.75) | 0.82 (0.66 - 1.00) | 1.17 (0.91 - 1.50) | Reference |
| Age adjusted HR (95% CI) | |  | 0.41 (0.32 - 0.53) | 0.87 (0.62 - 1.23) | Reference |  |  | 0.65 (0.55 - 0.76) | 0.87 (0.67 - 1.12) | Reference |  | 0.78 (0.42 - 1.43) | 0.70 (0.58 - 0.84) | 0.85 (0.66 - 1.10) | Reference |  | 0.62 (0.51 - 0.77) | 0.84 (0.68 - 1.03) | 1.19 (0.93 - 1.54) | Reference |
| Age and gender adjusted HR (95% CI) |  |  | 0.41 (0.32 - 0.53) | 0.87 (0.62 - 1.23) | Reference |  |  | 0.65 (0.55 - 0.76) | 0.86 (0.67 - 1.11) | Reference |  | 0.77 (0.42 - 1.42) | 0.70 (0.58 - 0.83) | 0.84 (0.65 - 1.09) | Reference |  | 0.62 (0.50 - 0.76) | 0.84 (0.68 - 1.03) | 1.18 (0.91 - 1.51) | Reference |
|  | Non-COVID-19 mortality | | | | | Non-COVID-19 mortality | | | | | Non-COVID-19 mortality | | | | | Non-COVID-19 mortality | | | | |
| Deaths (n) |  |  | 116 | 44 | 126 |  |  | 405 | 80 | 221 |  | 17 | 415 | 90 | 160 |  | 314 | 283 | 111 | 123 |
| Events per 100,000 person days | |  | 46.35 | 78.96 | 100.98 |  |  | 58.44 | 78 | 90.60 |  | 59.28 | 56.4 | 68.32 | 82.17 |  | 65.1 | 77.97 | 114.36 | 95.23 |
| Crude HR (95%CI) |  |  | 0.44 (0.34 - 0.57) | 0.92 (0.64 - 1.32) | Reference |  |  | 0.64 (0.54 - 0.75) | 0.88 (0.68 - 1.13) | Reference |  | 0.82 (0.44 - 1.50) | 0.69 (0.57 - 0.82) | 0.84 (0.65 - 1.09) | Reference |  | 0.65 (0.53 - 0.80) | 0.85 (0.68 - 1.05) | 1.23 (0.95 - 1.59) | Reference |
| Age adjusted HR (95% CI) | |  | 0.44 (0.34 - 0.57) | 0.92 (0.64 - 1.32) | Reference |  |  | 0.64 (0.54 - 0.76) | 0.89 (0.69 - 1.15) | Reference |  | 0.81 (0.44 - 1.50) | 0.69 (0.58 - 0.83) | 0.84 (0.65 - 1.09) | Reference |  | 0.67 (0.54 - 0.83) | 0.87 (0.71 - 1.08) | 1.25 (0.97 - 1.62) | Reference |
| Age and gender adjusted HR (95% CI) |  |  | 0.44 (0.34 - 0.57) | 0.92 (0.64 - 1.31) | Reference |  |  | 0.64 (0.54 - 0.76) | 0.88 (0.68 - 1.14) | Reference |  | 0.81 (0.44 - 1.49) | 0.69 (0.58 - 0.83) | 0.84 (0.65 - 1.08) | Reference |  | 0.66 (0.53 - 0.82) | 0.87 (0.70 - 1.07) | 1.23 (0.95 - 1.60) | Reference |
|  | 2022 | | | | | | | | | | | | | | | | | | | |
|  | first quarter |  |  |  | 0 | second quarter | | | | | third quarter |  |  |  | 0 | fourth quarter | | | | |
|  | Four or more vaccine doses | Three vaccine doses | Two vaccine doses | One vaccine dose | Unvaccinated | Four or more vaccine doses | Three vaccine doses | Two vaccine doses | One vaccine dose | Unvaccinated | Four or more vaccine doses | Three vaccine doses | Two vaccine doses | One vaccine dose | Unvaccinated | Four or more vaccine doses | Three vaccine doses | Two vaccine doses | One vaccine dose | Unvaccinated |
|  | All-cause mortality | | | | | All-cause mortality | | | | | All-cause mortality | | | | | All-cause mortality | | | | |
| Deaths (n) | 4 | 492 | 226 | 41 | 103 | 23 | 861 | 253 | 31 | 124 | 413 | 1287 | 250 | 42 | 179 | 1253 | 1026 | 222 | 39 | 178 |
| Events per 100,000 person days | 240.24 | 72.33 | 94.7 | 102.95 | 109.38 | 57.68 | 78.58 | 93.92 | 80.53 | 94.28 | 67.62 | 85.89 | 92.08 | 103.04 | 97.49 | 87.63 | 117.57 | 119.95 | 116.66 | 94.11 |
| Crude HR (95%CI) | 2.26 (0.83 - 6.19) | 0.66 (0.53 - 0.81) | 0.87 (0.69 - 1.10) | 0.96 (0.66 - 1.37) | Reference | 0.66 (0.42 - 1.05) | 0.84 (0.69 - 1.01) | 1.00 (0.80 - 1.24) | 0.86 (0.58 - 1.28) | Reference | 0.71 (0.59 - 0.85) | 0.90 (0.77 - 1.06) | 0.97 (0.80 - 1.17) | 1.05 (0.75 - 1.46) | Reference | 0.92 (0.78 - 1.07) | 1.27 (1.08 - 1.49) | 1.28 (1.05 - 1.56) | 1.21 (0.86 - 1.71) | Reference |
| Age adjusted HR (95% CI) | 2.27 (0.83 - 6.21) | 0.68 (0.55 - 0.84) | 0.89 (0.71 - 1.13) | 0.98 (0.68 - 1.40) | Reference | 0.65 (0.41 - 1.03) | 0.84 (0.70 - 1.01) | 1.01 (0.81 - 1.25) | 0.88 (0.60 - 1.31) | Reference | 0.70 (0.59 - 0.84) | 0.91 (0.77 - 1.06) | 0.98 (0.81 - 1.19) | 1.07 (0.76 - 1.50) | Reference | 0.92 (0.79 - 1.07) | 1.29 (1.10 - 1.51) | 1.30 (1.07 - 1.58) | 1.24 (0.88 - 1.76) | Reference |
| Age and gender adjusted HR (95% CI) | (-) | 0.67 (0.54 - 0.83) | 0.87 (0.69 - 1.10) | 0.97 (0.67 - 1.39) | Reference | 0.65 (0.41 - 1.03) | 0.82 (0.68 - 0.99) | 0.98 (0.79 - 1.22) | 0.88 (0.59 - 1.30) | Reference | 0.69 (0.57 - 0.82) | 0.88 (0.75 - 1.03) | 0.96 (0.79 - 1.16) | 1.07 (0.76 - 1.49) | Reference | 0.91 (0.77 - 1.06) | 1.26 (1.08 - 1.48) | 1.28 (1.05 - 1.56) | 1.24 (0.88 - 1.76) | Reference |

| **Table S20 continued:** | | | | | | | | | | | | | | | | | | | | |
| --- | --- | --- | --- | --- | --- | --- | --- | --- | --- | --- | --- | --- | --- | --- | --- | --- | --- | --- | --- | --- |
|  | 2022 | | | | | | | | | | | | | | | | | | | |
|  | first quarter |  |  |  | 0 | second quarter | | | | | third quarter |  |  |  | 0 | fourth quarter | | | | |
|  | Four or more vaccine doses | Three vaccine doses | Two vaccine doses | One vaccine dose | Unvacc | Four or more vaccine doses | Three vaccine doses | Two vaccine doses | One vaccine dose | Unvacc | Four or more vaccine doses | Three vaccine doses | Two vaccine doses | One vaccine dose | Unvacc | Four or more vaccine doses | Three vaccine doses | Two vaccine doses | One vaccine dose | Unvacc |
|  | Non-COVID-19 mortality | | | | | Non-COVID-19 mortality | | | | | Non-COVID-19 mortality | | | | | Non-COVID-19 mortality | | | | |
| Deaths (n) | 2 | 461 | 210 | 39 | 90 | 23 | 836 | 241 | 29 | 120 | 406 | 1253 | 240 | 41 | 173 | 1216 | 982 | 215 | 38 | 171 |
| Events per 100,000 person days | 120.12 | 67.77 | 88 | 97.92 | 95.58 | 57.68 | 76.3 | 89.47 | 75.34 | 91.23 | 66.48 | 83.62 | 88.4 | 100.59 | 94.22 | 85.05 | 112.53 | 116.17 | 113.67 | 90.41 |
| Crude HR (95%CI) | 1.32 (0.32 - 5.39) | 0.71 (0.56 - 0.88) | 0.93 (0.73 - 1.19) | 1.03 (0.71 - 1.51) | Reference | 0.67 (0.42 - 1.06) | 0.84 (0.69 - 1.01) | 0.98 (0.79 - 1.23) | 0.84 (0.56 - 1.26) | Reference | 0.72 (0.60 - 0.87) | 0.91 (0.78 - 1.07) | 0.96 (0.79 - 1.17) | 1.06 (0.75 - 1.48) | Reference | 0.93 (0.79 - 1.09) | 1.27 (1.08 - 1.49) | 1.29 (1.06 - 1.58) | 1.23 (0.86 - 1.74) | Reference |
| Age adjusted HR (95% CI) | 1.33 (0.32 - 5.43) | 0.73 (0.58 - 0.91) | 0.95 (0.74 - 1.22) | 1.06 (0.73 - 1.55) | Reference | 0.66 (0.41 - 1.04) | 0.84 (0.70 - 1.02) | 0.99 (0.80 - 1.24) | 0.86 (0.57 - 1.29) | Reference | 0.72 (0.60 - 0.86) | 0.91 (0.78 - 1.07) | 0.97 (0.80 - 1.19) | 1.08 (0.77 - 1.52) | Reference | 0.93 (0.79 - 1.09) | 1.29 (1.09 - 1.51) | 1.31 (1.07 - 1.60) | 1.26 (0.89 - 1.80) | Reference |
| Age and gender adjusted HR (95% CI) | (-) | 0.72 (0.57 - 0.90) | 0.92 (0.72 - 1.18) | 1.05 (0.72 - 1.54) | Reference | 0.65 (0.41 - 1.04) | 0.82 (0.68 - 0.99) | 0.97 (0.78 - 1.20) | 0.86 (0.57 - 1.28) | Reference | 0.70 (0.58 - 0.84) | 0.89 (0.76 - 1.04) | 0.95 (0.78 - 1.16) | 1.08 (0.77 - 1.51) | Reference | 0.91 (0.78 - 1.07) | 1.26 (1.07 - 1.49) | 1.29 (1.06 - 1.58) | 1.26 (0.89 - 1.79) | Reference |
|  | 2023 | | | | | | | | | | | | | | | | | | | |
|  | first quarter |  |  |  | 0 | second quarter | | | | | third quarter |  |  |  | 0 | fourth quarter | | | | |
|  | Four or more vaccine doses | Three vaccine doses | Two vaccine doses | One vaccine dose | Unvaccinated | Four or more vaccine doses | Three vaccine doses | Two vaccine doses | One vaccine dose | Unvaccinated | Four or more vaccine doses | Three vaccine doses | Two vaccine doses | One vaccine dose | Unvaccinated | Four or more vaccine doses | Three vaccine doses | Two vaccine doses | One vaccine dose | Unvaccinated |
|  | All-cause mortality | | | | | All-cause mortality | | | | | All-cause mortality | | | | | All-cause mortality | | | | |
| Deaths (n) | 1494 | 851 | 186 | 29 | 212 | 1366 | 663 | 140 | 33 | 168 | 1294 | 608 | 133 | 19 | 159 | 1511 | 644 | 144 | 24 | 146 |
| Events per 100,000 person days | 92.88 | 111.27 | 110.84 | 87 | 110.32 | 78.88 | 83.84 | 80.55 | 100.49 | 83.18 | 74.04 | 77.5 | 77.87 | 61.58 | 80.03 | 92.16 | 87.81 | 89.3 | 83.24 | 76.61 |
| Crude HR (95%CI) | 0.84 (0.73 - 0.97) | 1.01 (0.87 - 1.17) | 1.00 (0.82 - 1.22) | 0.81 (0.55 - 1.19) | Reference | 0.94 (0.80 - 1.11) | 1.00 (0.85 - 1.19) | 0.96 (0.77 - 1.20) | 1.21 (0.83 - 1.76) | Reference | 0.93 (0.79 - 1.10) | 0.98 (0.82 - 1.16) | 0.98 (0.78 - 1.23) | 0.76 (0.47 - 1.23) | Reference | 1.20 (1.01 - 1.42) | 1.13 (0.95 - 1.36) | 1.16 (0.92 - 1.47) | 1.06 (0.69 - 1.63) | Reference |
| Age adjusted HR (95% CI) | 0.84 (0.73 - 0.97) | 1.02 (0.88 - 1.19) | 1.03 (0.84 - 1.25) | 0.84 (0.57 - 1.24) | Reference | 0.94 (0.80 - 1.10) | 1.02 (0.86 - 1.21) | 0.98 (0.79 - 1.23) | 1.25 (0.86 - 1.82) | Reference | 0.92 (0.78 - 1.09) | 0.99 (0.83 - 1.18) | 1.00 (0.79 - 1.25) | 0.79 (0.49 - 1.26) | Reference | 1.19 (1.00 - 1.41) | 1.15 (0.96 - 1.37) | 1.19 (0.95 - 1.50) | 1.09 (0.71 - 1.67) | Reference |
| Age and gender adjusted HR (95% CI) | 0.82 (0.71 - 0.95) | 1.00 (0.86 - 1.17) | 1.01 (0.83 - 1.23) | 0.84 (0.57 - 1.25) | Reference | 0.91 (0.78 - 1.07) | 1.00 (0.84 - 1.18) | 0.96 (0.77 - 1.20) | 1.27 (0.87 - 1.84) | Reference | 0.91 (0.77 - 1.07) | 0.97 (0.81 - 1.15) | 0.98 (0.78 - 1.24) | 0.79 (0.49 - 1.27) | Reference | 1.16 (0.98 - 1.38) | 1.13 (0.95 - 1.36) | 1.18 (0.94 - 1.49) | 1.09 (0.71 - 1.68) | Reference |
|  | Non-COVID-19 mortality | | | | | Non-COVID-19 mortality | | | | | Non-COVID-19 mortality | | | | | Non-COVID-19 mortality | | | | |
| Deaths (n) | 1420 | 827 | 179 | 29 | 201 | 1346 | 653 | 140 | 31 | 165 | 1290 | 605 | 132 | 19 | 157 | 1445 | 626 | 140 | 24 | 142 |
| Events per 100,000 person days | 88.28 | 108.13 | 106.67 | 87 | 104.59 | 77.73 | 82.58 | 80.55 | 94.4 | 81.70 | 73.81 | 77.11 | 77.28 | 61.58 | 79.02 | 88.13 | 85.36 | 86.81 | 83.24 | 74.51 |
| Crude HR (95%CI) | 0.85 (0.73 - 0.98) | 1.03 (0.89 - 1.20) | 1.02 (0.83 - 1.25) | 0.85 (0.58 - 1.26) | Reference | 0.95 (0.81 - 1.11) | 1.01 (0.85 - 1.19) | 0.98 (0.78 - 1.23) | 1.16 (0.79 - 1.70) | Reference | 0.94 (0.80 - 1.11) | 0.98 (0.83 - 1.17) | 0.98 (0.78 - 1.24) | 0.77 (0.48 - 1.24) | Reference | 1.18 (0.99 - 1.40) | 1.13 (0.95 - 1.36) | 1.16 (0.92 - 1.47) | 1.09 (0.70 - 1.67) | Reference |
| Age adjusted HR (95% CI) | 0.85 (0.73 - 0.98) | 1.05 (0.90 - 1.22) | 1.04 (0.85 - 1.28) | 0.89 (0.60 - 1.31) | Reference | 0.94 (0.80 - 1.11) | 1.03 (0.86 - 1.22) | 1.00 (0.80 - 1.26) | 1.20 (0.82 - 1.77) | Reference | 0.93 (0.79 - 1.10) | 1.00 (0.84 - 1.19) | 1.00 (0.79 - 1.26) | 0.80 (0.49 - 1.28) | Reference | 1.17 (0.98 - 1.39) | 1.15 (0.96 - 1.38) | 1.19 (0.94 - 1.51) | 1.12 (0.72 - 1.72) | Reference |
| Age and gender adjusted HR (95% CI) | 0.83 (0.71 - 0.96) | 1.03 (0.88 - 1.20) | 1.03 (0.84 - 1.26) | 0.89 (0.60 - 1.32) | Reference | 0.92 (0.78 - 1.08) | 1.00 (0.85 - 1.19) | 0.98 (0.78 - 1.23) | 1.21 (0.83 - 1.78) | Reference | 0.92 (0.78 - 1.08) | 0.98 (0.82 - 1.17) | 0.99 (0.79 - 1.25) | 0.80 (0.50 - 1.28) | Reference | 1.14 (0.96 - 1.36) | 1.13 (0.94 - 1.36) | 1.18 (0.94 - 1.50) | 1.12 (0.73 - 1.73) | Reference |

| **Table S21:** Hazard ratios (HR) with 95% confidence intervals (95% CI) for non-COVID 19 and all-cause mortality according to number of SARS-CoV-2 vaccine doses for the period from 2021 to 2023 split into 3-month intervals for **communiy dwelling** individuals only. | | | | | | | | | | | | | | | | | | | | |
| --- | --- | --- | --- | --- | --- | --- | --- | --- | --- | --- | --- | --- | --- | --- | --- | --- | --- | --- | --- | --- |
|  | 2021 | | | | | | | | | | | | | | | | | | | |
|  | first quarter | | | | | second quarter | | | | | third quarter | | | | | fourth quarter | | | | |
|  | Four or more vaccine doses | Three vaccine doses | Two vaccine doses | One vaccine dose | Unvaccinated | Four or more vaccine doses | Three vaccine doses | Two vaccine doses | One vaccine dose | Unvaccinated | Four or more vaccine doses | Three vaccine doses | Two vaccine doses | One vaccine dose | Unvaccinated | Four or more vaccine doses | Three vaccine doses | Two vaccine doses | One vaccine dose | Unvaccinated |
|  | All-cause mortality | | | | | All-cause mortality | | | | | All-cause mortality | | | | | All-cause mortality | | | | |
| Deaths (n) |  |  | 24 | 22 | 257 |  |  | 131 | 106 | 475 |  | 2 | 296 | 179 | 426 |  | 100 | 304 | 171 | 292 |
| Events per 100,000 person days | |  | 5.03 | 3.13 | 1.90 |  |  | 3.89 | 1.58 | 2.27 |  | 10.13 | 2.34 | 1.57 | 2.30 |  | 2.72 | 1.75 | 1.3 | 2.22 |
| Crude HR (95%CI) |  |  | 2.62 (1.72 - 4.01) | 1.69 (1.08 - 2.62) | Reference |  |  | 1.79 (1.47 - 2.18) | 0.73 (0.59 - 0.91) | Reference |  | 4.74 (1.17 - 19.29) | 1.03 (0.89 - 1.20) | 0.70 (0.59 - 0.83) | Reference |  | 1.50 (1.16 - 1.94) | 0.79 (0.68 - 0.93) | 0.59 (0.49 - 0.72) | Reference |
| Age adjusted HR (95% CI) | |  | 0.83 (0.54 - 1.29) | 0.65 (0.42 - 1.02) | Reference |  |  | 0.50 (0.41 - 0.61) | 0.45 (0.37 - 0.56) | Reference |  | 0.81 (0.20 - 3.31) | 0.57 (0.49 - 0.66) | 0.50 (0.42 - 0.59) | Reference |  | 0.41 (0.32 - 0.53) | 0.52 (0.44 - 0.61) | 0.50 (0.41 - 0.60) | Reference |
| Age and gender adjusted HR (95% CI) |  |  | 0.86 (0.55 - 1.32) | 0.65 (0.41 - 1.01) | Reference |  |  | 0.49 (0.40 - 0.60) | 0.44 (0.36 - 0.55) | Reference |  | (-) | 0.55 (0.47 - 0.64) | 0.49 (0.41 - 0.58) | Reference |  | 0.40 (0.31 - 0.51) | 0.51 (0.43 - 0.59) | 0.49 (0.41 - 0.59) | Reference |
|  | Non-COVID-19 mortality | | | | | Non-COVID-19 mortality | | | | | Non-COVID-19 mortality | | | | | Non-COVID-19 mortality | | | | |
| Deaths (n) |  |  | 23 | 19 | 241 |  |  | 121 | 102 | 448 |  | 2 | 291 | 174 | 398 |  | 96 | 297 | 165 | 272 |
| Events per 100,000 person days | |  | 4.82 | 2.7 | 1.78 |  |  | 3.59 | 1.52 | 2.14 |  | 10.13 | 2.3 | 1.53 | 2.15 |  | 2.61 | 1.71 | 1.25 | 2.07 |
| Crude HR (95%CI) |  |  | 2.68 (1.74 - 4.13) | 1.58 (0.98 - 2.53) | Reference |  |  | 1.72 (1.40 - 2.12) | 0.74 (0.59 - 0.92) | Reference |  | 4.94 (1.21 - 20.10) | 1.08 (0.93 - 1.26) | 0.72 (0.61 - 0.87) | Reference |  | 1.58 (1.21 - 2.06) | 0.83 (0.71 - 0.98) | 0.62 (0.51 - 0.75) | Reference |
| Age adjusted HR (95% CI) | |  | 0.86 (0.55 - 1.34) | 0.61 (0.38 - 0.99) | Reference |  |  | 0.48 (0.39 - 0.59) | 0.46 (0.37 - 0.57) | Reference |  | 0.84 (0.20 - 3.43) | 0.59 (0.51 - 0.69) | 0.52 (0.43 - 0.62) | Reference |  | 0.43 (0.34 - 0.56) | 0.55 (0.46 - 0.64) | 0.52 (0.43 - 0.63) | Reference |
| Age and gender adjusted HR (95% CI) |  |  | 0.88 (0.56 - 1.38) | 0.61 (0.37 - 0.98) | Reference |  |  | 0.47 (0.38 - 0.58) | 0.45 (0.36 - 0.56) | Reference |  | (-) | 0.58 (0.50 - 0.67) | 0.51 (0.43 - 0.61) | Reference |  | 0.42 (0.33 - 0.54) | 0.53 (0.45 - 0.63) | 0.51 (0.42 - 0.62) | Reference |
|  | 2022 | | | | | | | | | | | | | | | | | | | |
|  | first quarter | | | | | second quarter | | | | | third quarter | | | | | fourth quarter | | | | |
|  | Four or more vaccine doses | Three vaccine doses | Two vaccine doses | One vaccine dose | Unvaccinated | Four or more vaccine doses | Three vaccine doses | Two vaccine doses | One vaccine dose | Unvaccinated | Four or more vaccine doses | Three vaccine doses | Two vaccine doses | One vaccine dose | Unvaccinated | Four or more vaccine doses | Three vaccine doses | Two vaccine doses | One vaccine dose | Unvaccinated |
|  | All-cause mortality | | | | | All-cause mortality | | | | | All-cause mortality | | | | | All-cause mortality | | | | |
| Deaths (n) | 2 | 323 | 429 | 123 | 234 | 15 | 839 | 556 | 104 | 409 | 236 | 1895 | 670 | 134 | 600 | 1080 | 2186 | 658 | 128 | 736 |
| Events per 100,000 person days | 18.04 | 2.06 | 1.93 | 1.34 | 1.29 | 8.85 | 1.57 | 1.39 | 1.04 | 0.93 | 4.71 | 1.65 | 1.24 | 1.25 | 0.97 | 3.6 | 1.76 | 1.18 | 1.16 | 1.11 |
| Crude HR (95%CI) | 15.28 (3.80 - 61.50) | 1.58 (1.33 - 1.86) | 1.48 (1.27 - 1.74) | 0.98 (0.79 - 1.22) | Reference | 10.07 (5.98 - 16.94) | 1.72 (1.52 - 1.93) | 1.50 (1.32 - 1.71) | 1.12 (0.90 - 1.38) | Reference | 5.11 (4.37 - 5.97) | 1.72 (1.57 - 1.88) | 1.29 (1.16 - 1.44) | 1.29 (1.07 - 1.56) | Reference | 3.23 (2.94 - 3.55) | 1.60 (1.48 - 1.74) | 1.07 (0.97 - 1.19) | 1.05 (0.87 - 1.27) | Reference |
| Age adjusted HR (95% CI) | 2.78 (0.69 - 11.21) | 0.60 (0.50 - 0.71) | 0.80 (0.68 - 0.94) | 0.93 (0.75 - 1.16) | Reference | 0.90 (0.53 - 1.53) | 0.72 (0.64 - 0.81) | 1.10 (0.97 - 1.25) | 1.12 (0.90 - 1.39) | Reference | 0.62 (0.53 - 0.73) | 0.87 (0.80 - 0.96) | 1.28 (1.15 - 1.43) | 1.44 (1.19 - 1.74) | Reference | 0.66 (0.60 - 0.72) | 1.03 (0.95 - 1.12) | 1.23 (1.10 - 1.36) | 1.25 (1.03 - 1.51) | Reference |
| Age and gender adjusted HR (95% CI) | 2.58 (0.64 - 10.40) | 0.57 (0.48 - 0.68) | 0.79 (0.67 - 0.92) | 0.92 (0.74 - 1.15) | Reference | 0.84 (0.50 - 1.42) | 0.69 (0.61 - 0.78) | 1.07 (0.94 - 1.22) | 1.11 (0.89 - 1.38) | Reference | 0.59 (0.50 - 0.70) | 0.84 (0.77 - 0.92) | 1.26 (1.13 - 1.41) | 1.43 (1.18 - 1.72) | Reference | 0.62 (0.56 - 0.68) | 1.00 (0.92 - 1.09) | 1.21 (1.09 - 1.34) | 1.24 (1.03 - 1.49) | Reference |

| **Table S21 continued:** | | | | | | | | | | | | | | | | | | | | |
| --- | --- | --- | --- | --- | --- | --- | --- | --- | --- | --- | --- | --- | --- | --- | --- | --- | --- | --- | --- | --- |
|  | 2022 | | | | | | | | | | | | | | | | | | | |
|  | first quarter | | | | | second quarter | | | | | third quarter | | | | | fourth quarter | | | | |
|  | Four or more vaccine doses | Three vaccine doses | Two vaccine doses | One vaccine dose | Unvacc | Four or more vaccine doses | Three vaccine doses | Two vaccine doses | One vaccine dose | Unvacc | Four or more vaccine doses | Three vaccine doses | Two vaccine doses | One vaccine dose | Unvacc | Four or more vaccine doses | Three vaccine doses | Two vaccine doses | One vaccine dose | Unvacc |
|  | Non-COVID-19 mortality | | | | | Non-COVID-19 mortality | | | | | Non-COVID-19 mortality | | | | | Non-COVID-19 mortality | | | | |
| Deaths (n) | 2 | 315 | 408 | 121 | 208 | 14 | 824 | 536 | 101 | 397 | 233 | 1858 | 654 | 129 | 587 | 1067 | 2149 | 649 | 127 | 721 |
| Events per 100,000 person days | 18.04 | 2.01 | 1.83 | 1.32 | 1.15 | 8.26 | 1.54 | 1.34 | 1.01 | 0.90 | 4.65 | 1.62 | 1.21 | 1.2 | 0.95 | 3.55 | 1.73 | 1.17 | 1.15 | 1.09 |
| Crude HR (95%CI) | 17.35 (4.31 - 69.88) | 1.72 (1.45 - 2.05) | 1.59 (1.34 - 1.87) | 1.07 (0.85 - 1.34) | Reference | 9.63 (5.62 - 16.50) | 1.74 (1.54 - 1.96) | 1.49 (1.31 - 1.70) | 1.12 (0.90 - 1.39) | Reference | 5.15 (4.40 - 6.02) | 1.72 (1.57 - 1.89) | 1.29 (1.15 - 1.44) | 1.27 (1.05 - 1.54) | Reference | 3.25 (2.96 - 3.58) | 1.61 (1.48 - 1.75) | 1.08 (0.97 - 1.20) | 1.07 (0.88 - 1.29) | Reference |
| Age adjusted HR (95% CI) | 3.18 (0.79 - 12.83) | 0.66 (0.55 - 0.78) | 0.86 (0.73 - 1.02) | 1.02 (0.81 - 1.28) | Reference | 0.87 (0.51 - 1.50) | 0.73 (0.65 - 0.82) | 1.10 (0.96 - 1.25) | 1.12 (0.90 - 1.40) | Reference | 0.63 (0.54 - 0.74) | 0.88 (0.80 - 0.96) | 1.28 (1.14 - 1.43) | 1.42 (1.17 - 1.71) | Reference | 0.66 (0.60 - 0.73) | 1.04 (0.95 - 1.13) | 1.24 (1.11 - 1.37) | 1.26 (1.05 - 1.53) | Reference |
| Age and gender adjusted HR (95% CI) | 2.98 (0.74 - 12.03) | 0.63 (0.52 - 0.75) | 0.84 (0.71 - 1.00) | 1.01 (0.80 - 1.26) | Reference | 0.82 (0.48 - 1.42) | 0.70 (0.62 - 0.79) | 1.07 (0.94 - 1.22) | 1.11 (0.89 - 1.39) | Reference | 0.60 (0.51 - 0.71) | 0.84 (0.77 - 0.93) | 1.26 (1.13 - 1.41) | 1.41 (1.16 - 1.70) | Reference | 0.63 (0.57 - 0.69) | 1.01 (0.92 - 1.09) | 1.22 (1.10 - 1.35) | 1.25 (1.04 - 1.51) | Reference |
|  | 2023 | | | | | | | | | | | | | | | | | | | |
|  | first quarter | | | | | second quarter | | | | | third quarter | | | | | fourth quarter | | | | |
|  | Four or more vaccine doses | Three vaccine doses | Two vaccine doses | One vaccine dose | Unvaccinated | Four or more vaccine doses | Three vaccine doses | Two vaccine doses | One vaccine dose | Unvaccinated | Four or more vaccine doses | Three vaccine doses | Two vaccine doses | One vaccine dose | Unvaccinated | Four or more vaccine doses | Three vaccine doses | Two vaccine doses | One vaccine dose | Unvaccinated |
|  | All-cause mortality | | | | | All-cause mortality | | | | | All-cause mortality | | | | | All-cause mortality | | | | |
| Deaths (n) | 1691 | 2211 | 646 | 106 | 837 | 2001 | 2109 | 659 | 132 | 843 | 2199 | 2218 | 637 | 130 | 827 | 2440 | 2440 | 628 | 128 | 895 |
| Events per 100,000 person days | 3.66 | 1.72 | 1.16 | 0.98 | 1.25 | 3.79 | 1.53 | 1.13 | 1.18 | 1.21 | 3.93 | 1.56 | 1.07 | 1.14 | 1.17 | 4.36 | 1.71 | 1.05 | 1.12 | 1.26 |
| Crude HR (95%CI) | 2.93 (2.70 - 3.19) | 1.38 (1.27 - 1.49) | 0.93 (0.84 - 1.03) | 0.78 (0.64 - 0.96) | Reference | 3.13 (2.89 - 3.39) | 1.26 (1.17 - 1.37) | 0.94 (0.85 - 1.04) | 0.98 (0.81 - 1.17) | Reference | 3.37 (3.11 - 3.65) | 1.34 (1.24 - 1.45) | 0.92 (0.83 - 1.02) | 0.98 (0.82 - 1.18) | Reference | 3.47 (3.21 - 3.74) | 1.36 (1.26 - 1.47) | 0.84 (0.75 - 0.92) | 0.89 (0.74 - 1.07) | Reference |
| Age adjusted HR (95% CI) | 0.69 (0.63 - 0.75) | 0.93 (0.86 - 1.00) | 1.12 (1.01 - 1.24) | 0.97 (0.79 - 1.18) | Reference | 0.76 (0.70 - 0.82) | 0.86 (0.80 - 0.93) | 1.15 (1.04 - 1.27) | 1.23 (1.02 - 1.47) | Reference | 0.86 (0.79 - 0.94) | 0.92 (0.85 - 1.00) | 1.13 (1.02 - 1.25) | 1.24 (1.03 - 1.49) | Reference | 0.82 (0.76 - 0.88) | 0.92 (0.85 - 0.99) | 1.04 (0.94 - 1.15) | 1.14 (0.95 - 1.37) | Reference |
| Age and gender adjusted HR (95% CI) | 0.65 (0.60 - 0.71) | 0.90 (0.83 - 0.98) | 1.11 (1.00 - 1.23) | 0.96 (0.78 - 1.17) | Reference | 0.72 (0.66 - 0.78) | 0.84 (0.78 - 0.91) | 1.13 (1.02 - 1.25) | 1.21 (1.01 - 1.46) | Reference | 0.83 (0.76 - 0.90) | 0.90 (0.83 - 0.97) | 1.11 (1.00 - 1.23) | 1.23 (1.02 - 1.48) | Reference | 0.78 (0.72 - 0.84) | 0.90 (0.83 - 0.97) | 1.02 (0.92 - 1.13) | 1.13 (0.94 - 1.36) | Reference |
|  | Non-COVID-19 mortality | | | | | Non-COVID-19 mortality | | | | | Non-COVID-19 mortality | | | | | Non-COVID-19 mortality | | | | |
| Deaths (n) | 1667 | 2170 | 634 | 106 | 820 | 1988 | 2094 | 650 | 132 | 836 | 2192 | 2203 | 635 | 128 | 823 | 2378 | 2355 | 609 | 125 | 872 |
| Events per 100,000 person days | 3.61 | 1.68 | 1.13 | 0.98 | 1.22 | 3.77 | 1.52 | 1.12 | 1.18 | 1.20 | 3.92 | 1.55 | 1.07 | 1.13 | 1.16 | 4.25 | 1.65 | 1.01 | 1.09 | 1.22 |
| Crude HR (95%CI) | 2.95 (2.71 - 3.21) | 1.38 (1.28 - 1.50) | 0.93 (0.84 - 1.03) | 0.80 (0.65 - 0.98) | Reference | 3.14 (2.89 - 3.40) | 1.27 (1.17 - 1.37) | 0.93 (0.84 - 1.03) | 0.99 (0.82 - 1.18) | Reference | 3.38 (3.12 - 3.66) | 1.34 (1.23 - 1.45) | 0.92 (0.83 - 1.02) | 0.97 (0.81 - 1.17) | Reference | 3.47 (3.21 - 3.75) | 1.35 (1.25 - 1.46) | 0.83 (0.75 - 0.92) | 0.90 (0.74 - 1.08) | Reference |
| Age adjusted HR (95% CI) | 0.69 (0.64 - 0.76) | 0.93 (0.86 - 1.01) | 1.12 (1.01 - 1.24) | 0.99 (0.81 - 1.21) | Reference | 0.76 (0.70 - 0.83) | 0.86 (0.80 - 0.94) | 1.14 (1.03 - 1.26) | 1.24 (1.03 - 1.49) | Reference | 0.87 (0.80 - 0.94) | 0.92 (0.85 - 1.00) | 1.13 (1.02 - 1.25) | 1.23 (1.02 - 1.48) | Reference | 0.82 (0.76 - 0.89) | 0.91 (0.85 - 0.99) | 1.03 (0.93 - 1.14) | 1.14 (0.95 - 1.38) | Reference |
| Age and gender adjusted HR (95% CI) | 0.66 (0.60 - 0.72) | 0.91 (0.84 - 0.98) | 1.11 (1.00 - 1.23) | 0.98 (0.80 - 1.20) | Reference | 0.72 (0.66 - 0.78) | 0.84 (0.78 - 0.91) | 1.12 (1.02 - 1.25) | 1.22 (1.02 - 1.47) | Reference | 0.83 (0.76 - 0.90) | 0.90 (0.83 - 0.97) | 1.11 (1.00 - 1.24) | 1.22 (1.01 - 1.47) | Reference | 0.78 (0.72 - 0.85) | 0.89 (0.83 - 0.96) | 1.02 (0.92 - 1.13) | 1.13 (0.94 - 1.36) | Reference |

| **Table S22:** Hazard ratios (HR) with 95% confidence intervals (95% CI) for non-COVID 19 and all-cause mortality according to number of SARS-CoV-2 vaccine doses during different time periods for **males** only. | | | | | | | | | | |
| --- | --- | --- | --- | --- | --- | --- | --- | --- | --- | --- |
|  | 2021 | | | | | | | | | |
|  | June and July 2021 (low COVID-19 disease burden) | | | | | October and November 2021 (high COVID-19 disease burden) | | | | |
|  | Four or more vaccine doses | Three vaccine doses | Two vaccine doses | One vaccine dose | Unvaccinated | Four or more vaccine doses | Three vaccine doses | Two vaccine doses | One vaccine dose | Unvaccinated |
|  | All-cause mortality | | | | | All-cause mortality | | | | |
| Deaths (n) | 0 | 0 | 137 | 67 | 173 | 0 | 67 | 151 | 102 | 116 |
| Events per 100,000 person days |  |  | 5.29 | 1.8 | 3.15 |  | 14.5 | 2.43 | 2.07 | 2.46 |
| Crude HR (95%CI) |  |  | 1.70 (1.35 - 2.13) | 0.56 (0.42 - 0.75) | Reference |  | 6.58 (4.71 - 9.19) | 0.98 (0.77 - 1.25) | 0.84 (0.64 - 1.10) | Reference |
| Age adjusted HR (95% CI) |  |  | 0.58 (0.46 - 0.73) | 0.44 (0.33 - 0.58) | Reference |  | 0.78 (0.56 - 1.08) | 0.53 (0.42 - 0.68) | 0.67 (0.51 - 0.87) | Reference |
| Age and nursing home adjusted HR (95% CI) |  |  | 0.48 (0.38 - 0.61) | 0.45 (0.34 - 0.59) | Reference |  | 0.57 (0.40 - 0.80) | 0.52 (0.41 - 0.66) | 0.70 (0.54 - 0.92) | Reference |
| Age, nursing home and year of last infection adjusted HR (95% CI) |  |  | 0.50 (0.39 - 0.63) | 0.45 (0.34 - 0.60) | Reference |  | 0.59 (0.42 - 0.85) | 0.52 (0.41 - 0.66) | 0.70 (0.54 - 0.91) | Reference |
|  | Non-COVID-19 mortality | | | | | Non-COVID-19 mortality | | | | |
| Deaths (n) | 0 | 0 | 132 | 66 | 172 | 0 | 65 | 146 | 98 | 108 |
| Events per 100,000 person days |  |  | 5.1 | 1.78 | 3.13 |  | 14.07 | 2.35 | 1.99 | 2.29 |
| Crude HR (95%CI) |  |  | 1.64 (1.30 - 2.07) | 0.56 (0.42 - 0.74) | Reference |  | 7.09 (5.03 - 9.98) | 1.02 (0.79 - 1.31) | 0.87 (0.66 - 1.15) | Reference |
| Age adjusted HR (95% CI) |  |  | 0.56 (0.44 - 0.70) | 0.43 (0.33 - 0.58) | Reference |  | 0.82 (0.58 - 1.15) | 0.55 (0.43 - 0.71) | 0.69 (0.53 - 0.91) | Reference |
| Age and nursing home adjusted HR (95% CI) |  |  | 0.47 (0.37 - 0.59) | 0.44 (0.33 - 0.59) | Reference |  | 0.58 (0.41 - 0.84) | 0.54 (0.42 - 0.69) | 0.73 (0.55 - 0.96) | Reference |
| Age, nursing home and year of last infection adjusted HR (95% CI) |  |  | 0.48 (0.38 - 0.61) | 0.45 (0.34 - 0.60) | Reference |  | 0.61 (0.42 - 0.88) | 0.54 (0.42 - 0.69) | 0.73 (0.55 - 0.96) | Reference |
|  | May and June 2022 (low COVID-19 disease burden) | | | | | February and March 2022 (high COVID-19 disease burden) | | | | |
|  | Four or more vaccine doses | Three vaccine doses | Two vaccine doses | One vaccine dose | Unvaccinated | Four or more vaccine doses | Three vaccine doses | Two vaccine doses | One vaccine dose | Unvaccinated |
|  | All-cause mortality | | | | | All-cause mortality | | | | |
| Deaths (n) | 9 | 306 | 197 | 31 | 85 | 0 | 187 | 152 | 40 | 76 |
| Events per 100,000 person days | 16.44 | 2.61 | 1.77 | 1.06 | 0.74 |  | 3.21 | 2.36 | 1.54 | 1.97 |
| Crude HR (95%CI) | 20.75 (10.37 - 41.50) | 3.51 (2.76 - 4.47) | 2.39 (1.85 - 3.08) | 1.43 (0.95 - 2.16) | Reference |  | 1.62 (1.24 - 2.12) | 1.20 (0.91 - 1.58) | 0.79 (0.54 - 1.15) | Reference |
| Age adjusted HR (95% CI) | 1.36 (0.66 - 2.80) | 1.07 (0.84 - 1.37) | 1.44 (1.12 - 1.87) | 1.33 (0.88 - 2.01) | Reference |  | 0.57 (0.43 - 0.75) | 0.65 (0.50 - 0.86) | 0.81 (0.55 - 1.19) | Reference |
| Age and nursing home adjusted HR (95% CI) | 1.04 (0.48 - 2.25) | 0.99 (0.77 - 1.27) | 1.41 (1.09 - 1.83) | 1.33 (0.88 - 2.00) | Reference |  | 0.50 (0.38 - 0.67) | 0.65 (0.50 - 0.86) | 0.86 (0.58 - 1.26) | Reference |
| Age, nursing home and year of last infection adjusted HR (95% CI) | 1.06 (0.49 - 2.29) | 1.01 (0.78 - 1.30) | 1.40 (1.07 - 1.82) | 1.40 (0.92 - 2.15) | Reference |  | 0.49 (0.37 - 0.65) | 0.66 (0.50 - 0.87) | 0.86 (0.59 - 1.26) | Reference |
|  | Non-COVID-19 mortality | | | | | Non-COVID-19 mortality | | | | |
| Deaths (n) | 9 | 301 | 196 | 30 | 81 | 0 | 180 | 146 | 39 | 68 |
| Events per 100,000 person days | 16.44 | 2.57 | 1.76 | 1.03 | 0.71 |  | 3.09 | 2.27 | 1.5 | 1.76 |
| Crude HR (95%CI) | 21.85 (10.90 - 43.80) | 3.63 (2.84 - 4.64) | 2.49 (1.92 - 3.23) | 1.46 (0.96 - 2.21) | Reference |  | 1.75 (1.32 - 2.31) | 1.29 (0.97 - 1.72) | 0.86 (0.58 - 1.27) | Reference |
| Age adjusted HR (95% CI) | 1.50 (0.72 - 3.11) | 1.11 (0.87 - 1.43) | 1.51 (1.16 - 1.96) | 1.36 (0.89 - 2.07) | Reference |  | 0.62 (0.47 - 0.82) | 0.70 (0.53 - 0.94) | 0.88 (0.60 - 1.31) | Reference |
| Age and nursing home adjusted HR (95% CI) | 1.12 (0.52 - 2.44) | 1.03 (0.80 - 1.33) | 1.48 (1.14 - 1.92) | 1.35 (0.89 - 2.06) | Reference |  | 0.55 (0.41 - 0.74) | 0.70 (0.53 - 0.94) | 0.94 (0.63 - 1.39) | Reference |
| Age, nursing home and year of last infection adjusted HR (95% CI) | 1.15 (0.53 - 2.51) | 1.06 (0.81 - 1.37) | 1.47 (1.12 - 1.92) | 1.43 (0.93 - 2.21) | Reference |  | 0.53 (0.39 - 0.71) | 0.71 (0.53 - 0.95) | 0.94 (0.63 - 1.39) | Reference |

| **Table S22 continued:** | | | | | | | | | | |
| --- | --- | --- | --- | --- | --- | --- | --- | --- | --- | --- |
|  | July and August 2023 (low COVID-19 disease burden) | | | | | February and March 2023 (high COVID-19 disease burden) | | | | |
|  | Four or more vaccine doses | Three vaccine doses | Two vaccine doses | One vaccine dose | Unvaccinated | Four or more vaccine doses | Three vaccine doses | Two vaccine doses | One vaccine dose | Unvaccinated |
|  | All-cause mortality | | | | | All-cause mortality | | | | |
| Deaths (n) | 1075 | 975 | 263 | 45 | 280 | 901 | 870 | 232 | 34 | 264 |
| Events per 100,000 person days | 5.2 | 1.92 | 1.17 | 1.04 | 1.09 | 5.4 | 2 | 1.16 | 0.86 | 1.13 |
| Crude HR (95%CI) | 4.79 (4.20 - 5.46) | 1.77 (1.55 - 2.02) | 1.08 (0.91 - 1.28) | 0.96 (0.70 - 1.31) | Reference | 4.78 (4.17 - 5.48) | 1.77 (1.55 - 2.04) | 1.03 (0.86 - 1.23) | 0.76 (0.53 - 1.09) | Reference |
| Age adjusted HR (95% CI) | 0.93 (0.81 - 1.07) | 1.07 (0.94 - 1.23) | 1.26 (1.07 - 1.50) | 1.20 (0.88 - 1.64) | Reference | 0.88 (0.76 - 1.02) | 1.03 (0.90 - 1.18) | 1.18 (0.99 - 1.41) | 0.93 (0.65 - 1.34) | Reference |
| Age and nursing home adjusted HR (95% CI) | 0.92 (0.80 - 1.06) | 1.06 (0.93 - 1.21) | 1.24 (1.05 - 1.47) | 1.19 (0.87 - 1.64) | Reference | 0.86 (0.74 - 1.00) | 1.01 (0.88 - 1.17) | 1.15 (0.96 - 1.37) | 0.93 (0.65 - 1.33) | Reference |
| Age, nursing home and year of last infection adjusted HR (95% CI) | 0.90 (0.78 - 1.04) | 1.04 (0.91 - 1.19) | 1.28 (1.08 - 1.52) | 1.22 (0.89 - 1.69) | Reference | 0.86 (0.74 - 1.00) | 1.01 (0.88 - 1.16) | 1.16 (0.97 - 1.38) | 0.92 (0.64 - 1.33) | Reference |
|  | Non-COVID-19 mortality | | | | | Non-COVID-19 mortality | | | | |
| Deaths (n) | 1073 | 970 | 263 | 45 | 277 | 876 | 848 | 227 | 34 | 258 |
| Events per 100,000 person days | 5.19 | 1.91 | 1.17 | 1.04 | 1.07 | 5.25 | 1.95 | 1.14 | 0.86 | 1.10 |
| Crude HR (95%CI) | 4.83 (4.23 - 5.51) | 1.78 (1.56 - 2.03) | 1.09 (0.92 - 1.29) | 0.97 (0.70 - 1.32) | Reference | 4.76 (4.14 - 5.46) | 1.77 (1.54 - 2.03) | 1.03 (0.86 - 1.23) | 0.78 (0.55 - 1.12) | Reference |
| Age adjusted HR (95% CI) | 0.95 (0.82 - 1.09) | 1.08 (0.94 - 1.23) | 1.28 (1.08 - 1.51) | 1.21 (0.88 - 1.66) | Reference | 0.89 (0.77 - 1.03) | 1.03 (0.90 - 1.18) | 1.18 (0.99 - 1.41) | 0.96 (0.67 - 1.37) | Reference |
| Age and nursing home adjusted HR (95% CI) | 0.93 (0.81 - 1.08) | 1.07 (0.93 - 1.22) | 1.25 (1.06 - 1.48) | 1.21 (0.88 - 1.65) | Reference | 0.87 (0.75 - 1.01) | 1.02 (0.88 - 1.17) | 1.15 (0.96 - 1.37) | 0.95 (0.66 - 1.36) | Reference |
| Age, nursing home and year of last infection adjusted HR (95% CI) | 0.91 (0.79 - 1.05) | 1.05 (0.92 - 1.20) | 1.29 (1.09 - 1.53) | 1.23 (0.89 - 1.70) | Reference | 0.87 (0.75 - 1.01) | 1.02 (0.88 - 1.17) | 1.16 (0.97 - 1.39) | 0.95 (0.66 - 1.37) | Reference |

| **Table S23:** Hazard ratios (HR) with 95% confidence intervals (95% CI) for non-COVID 19 and all-cause mortality according to number of SARS-CoV-2 vaccine doses during different time periods for **females** only. | | | | | | | | | | |
| --- | --- | --- | --- | --- | --- | --- | --- | --- | --- | --- |
|  | 2021 | | | | | | | | | |
|  | June and July 2021 (low COVID-19 disease burden) | | | | | October and November 2021 (high COVID-19 disease burden) | | | | |
|  | Four or more vaccine doses | Three vaccine doses | Two vaccine doses | One vaccine dose | Unvaccinated | Four or more vaccine doses | Three vaccine doses | Two vaccine doses | One vaccine dose | Unvaccinated |
|  | All-cause mortality | | | | | All-cause mortality | | | | |
| Deaths (n) | 0 | 0 | 242 | 74 | 203 | 0 | 124 | 249 | 95 | 173 |
| Events per 100,000 person days |  |  | 7.85 | 2.08 | 3.36 |  | 17.38 | 4.01 | 1.94 | 3.29 |
| Crude HR (95%CI) |  |  | 2.46 (2.03 - 2.97) | 0.61 (0.47 - 0.80) | Reference |  | 5.73 (4.47 - 7.34) | 1.21 (1.00 - 1.47) | 0.60 (0.46 - 0.77) | Reference |
| Age adjusted HR (95% CI) |  |  | 0.72 (0.60 - 0.87) | 0.59 (0.45 - 0.77) | Reference |  | 0.71 (0.55 - 0.90) | 0.76 (0.63 - 0.93) | 0.57 (0.45 - 0.74) | Reference |
| Age and nursing home adjusted HR (95% CI) |  |  | 0.61 (0.50 - 0.74) | 0.60 (0.46 - 0.78) | Reference |  | 0.56 (0.43 - 0.73) | 0.69 (0.57 - 0.84) | 0.60 (0.47 - 0.77) | Reference |
| Age, nursing home and year of last infection adjusted HR (95% CI) |  |  | 0.63 (0.51 - 0.76) | 0.60 (0.46 - 0.79) | Reference |  | 0.57 (0.44 - 0.73) | 0.70 (0.58 - 0.85) | 0.60 (0.46 - 0.77) | Reference |
|  | Non-COVID-19 mortality | | | | | Non-COVID-19 mortality | | | | |
| Deaths (n) | 0 | 0 | 237 | 73 | 195 | 0 | 124 | 244 | 95 | 168 |
| Events per 100,000 person days |  |  | 7.69 | 2.05 | 3.22 |  | 17.38 | 3.93 | 1.94 | 3.20 |
| Crude HR (95%CI) |  |  | 2.51 (2.07 - 3.04) | 0.63 (0.48 - 0.83) | Reference |  | 5.95 (4.63 - 7.63) | 1.22 (1.01 - 1.49) | 0.62 (0.48 - 0.79) | Reference |
| Age adjusted HR (95% CI) |  |  | 0.74 (0.61 - 0.89) | 0.61 (0.47 - 0.80) | Reference |  | 0.74 (0.57 - 0.94) | 0.77 (0.63 - 0.94) | 0.59 (0.46 - 0.76) | Reference |
| Age and nursing home adjusted HR (95% CI) |  |  | 0.61 (0.50 - 0.75) | 0.62 (0.47 - 0.81) | Reference |  | 0.58 (0.45 - 0.75) | 0.70 (0.57 - 0.85) | 0.62 (0.48 - 0.80) | Reference |
| Age, nursing home and year of last infection adjusted HR (95% CI) |  |  | 0.63 (0.52 - 0.77) | 0.62 (0.47 - 0.81) | Reference |  | 0.58 (0.45 - 0.75) | 0.71 (0.58 - 0.86) | 0.62 (0.48 - 0.79) | Reference |
|  | May and June 2022 (low COVID-19 disease burden) | | | | | February and March 2022 (high COVID-19 disease burden) | | | | |
|  | Four or more vaccine doses | Three vaccine doses | Two vaccine doses | One vaccine dose | Unvaccinated | Four or more vaccine doses | Three vaccine doses | Two vaccine doses | One vaccine dose | Unvaccinated |
|  | All-cause mortality | | | | | All-cause mortality | | | | |
| Deaths (n) | 13 | 353 | 190 | 31 | 145 | 5 | 285 | 204 | 36 | 89 |
| Events per 100,000 person days | 21.18 | 2.9 | 1.71 | 1.12 | 1.22 | 106.56 | 4.79 | 3.09 | 1.34 | 2.16 |
| Crude HR (95%CI) | 17.06 (9.60 - 30.29) | 2.38 (1.96 - 2.89) | 1.41 (1.13 - 1.74) | 0.92 (0.62 - 1.35) | Reference | 48.86 (19.83 - 120.41) | 2.21 (1.74 - 2.80) | 1.43 (1.11 - 1.83) | 0.62 (0.42 - 0.91) | Reference |
| Age adjusted HR (95% CI) | 0.83 (0.46 - 1.49) | 0.79 (0.65 - 0.96) | 0.89 (0.72 - 1.11) | 0.82 (0.55 - 1.20) | Reference | 6.73 (2.72 - 16.64) | 0.82 (0.65 - 1.04) | 0.94 (0.73 - 1.21) | 0.76 (0.52 - 1.12) | Reference |
| Age and nursing home adjusted HR (95% CI) | 0.56 (0.30 - 1.02) | 0.66 (0.54 - 0.81) | 0.86 (0.69 - 1.07) | 0.81 (0.55 - 1.19) | Reference | 4.96 (1.97 - 12.47) | 0.67 (0.53 - 0.86) | 0.95 (0.74 - 1.22) | 0.83 (0.56 - 1.22) | Reference |
| Age, nursing home and year of last infection adjusted HR (95% CI) | 0.53 (0.29 - 0.97) | 0.66 (0.54 - 0.81) | 0.84 (0.68 - 1.05) | 0.78 (0.53 - 1.16) | Reference | 4.84 (1.92 - 12.18) | 0.68 (0.54 - 0.88) | 0.95 (0.74 - 1.21) | 0.83 (0.56 - 1.22) | Reference |
|  | Non-COVID-19 mortality | | | | | Non-COVID-19 mortality | | | | |
| Deaths (n) | 13 | 348 | 186 | 31 | 144 | 3 | 265 | 193 | 35 | 82 |
| Events per 100,000 person days | 21.18 | 2.86 | 1.67 | 1.12 | 1.21 | 63.94 | 4.45 | 2.92 | 1.3 | 1.99 |
| Crude HR (95%CI) | 17.14 (9.65 - 30.45) | 2.36 (1.94 - 2.87) | 1.39 (1.11 - 1.72) | 0.93 (0.63 - 1.36) | Reference | 32.07 (10.12 - 101.61) | 2.23 (1.74 - 2.86) | 1.47 (1.13 - 1.90) | 0.65 (0.44 - 0.97) | Reference |
| Age adjusted HR (95% CI) | 0.83 (0.46 - 1.50) | 0.79 (0.65 - 0.96) | 0.88 (0.71 - 1.09) | 0.82 (0.56 - 1.21) | Reference | 4.42 (1.39 - 14.03) | 0.83 (0.65 - 1.06) | 0.97 (0.75 - 1.25) | 0.80 (0.54 - 1.19) | Reference |
| Age and nursing home adjusted HR (95% CI) | 0.56 (0.30 - 1.02) | 0.66 (0.54 - 0.81) | 0.85 (0.68 - 1.06) | 0.82 (0.55 - 1.20) | Reference | 3.22 (1.00 - 10.36) | 0.68 (0.53 - 0.88) | 0.97 (0.75 - 1.26) | 0.87 (0.58 - 1.30) | Reference |
| Age, nursing home and year of last infection adjusted HR (95% CI) | 0.53 (0.29 - 0.97) | 0.66 (0.54 - 0.81) | 0.83 (0.67 - 1.04) | 0.79 (0.53 - 1.17) | Reference | 3.15 (0.98 - 10.16) | 0.70 (0.54 - 0.90) | 0.97 (0.75 - 1.26) | 0.87 (0.58 - 1.30) | Reference |

| **Table S23 continued:** | | | | | | | | | | |
| --- | --- | --- | --- | --- | --- | --- | --- | --- | --- | --- |
|  | July and August 2023 (low COVID-19 disease burden) | | | | | February and March 2023 (high COVID-19 disease burden) | | | | |
|  | Four or more vaccine doses | Three vaccine doses | Two vaccine doses | One vaccine dose | Unvaccinated | Four or more vaccine doses | Three vaccine doses | Two vaccine doses | One vaccine dose | Unvaccinated |
|  | All-cause mortality | | | | | All-cause mortality | | | | |
| Deaths (n) | 1155 | 895 | 247 | 56 | 347 | 959 | 822 | 220 | 48 | 322 |
| Events per 100,000 person days | 5.02 | 1.54 | 1.08 | 1.29 | 1.21 | 5.27 | 1.68 | 1.08 | 1.23 | 1.25 |
| Crude HR (95%CI) | 4.14 (3.67 - 4.67) | 1.27 (1.12 - 1.44) | 0.89 (0.75 - 1.04) | 1.06 (0.80 - 1.41) | Reference | 4.21 (3.71 - 4.78) | 1.34 (1.18 - 1.52) | 0.87 (0.73 - 1.03) | 0.98 (0.72 - 1.33) | Reference |
| Age adjusted HR (95% CI) | 0.96 (0.85 - 1.09) | 0.91 (0.81 - 1.03) | 1.09 (0.92 - 1.28) | 1.31 (0.99 - 1.73) | Reference | 0.90 (0.79 - 1.02) | 0.94 (0.83 - 1.07) | 1.03 (0.87 - 1.23) | 1.17 (0.86 - 1.58) | Reference |
| Age and nursing home adjusted HR (95% CI) | 0.85 (0.75 - 0.97) | 0.89 (0.78 - 1.01) | 1.05 (0.89 - 1.24) | 1.27 (0.96 - 1.68) | Reference | 0.76 (0.67 - 0.87) | 0.90 (0.79 - 1.02) | 1.00 (0.85 - 1.19) | 1.15 (0.85 - 1.56) | Reference |
| Age, nursing home and year of last infection adjusted HR (95% CI) | 0.85 (0.75 - 0.96) | 0.88 (0.77 - 0.99) | 1.08 (0.92 - 1.27) | 1.35 (1.01 - 1.79) | Reference | 0.76 (0.67 - 0.87) | 0.90 (0.79 - 1.03) | 1.01 (0.85 - 1.21) | 1.18 (0.86 - 1.60) | Reference |
|  | Non-COVID-19 mortality | | | | | Non-COVID-19 mortality | | | | |
| Deaths (n) | 1154 | 893 | 247 | 55 | 346 | 928 | 813 | 217 | 48 | 312 |
| Events per 100,000 person days | 5.01 | 1.54 | 1.08 | 1.26 | 1.21 | 5.1 | 1.66 | 1.07 | 1.23 | 1.21 |
| Crude HR (95%CI) | 4.15 (3.68 - 4.68) | 1.27 (1.12 - 1.44) | 0.89 (0.76 - 1.05) | 1.05 (0.79 - 1.39) | Reference | 4.21 (3.70 - 4.78) | 1.37 (1.20 - 1.56) | 0.88 (0.74 - 1.05) | 1.01 (0.75 - 1.37) | Reference |
| Age adjusted HR (95% CI) | 0.96 (0.85 - 1.09) | 0.91 (0.81 - 1.03) | 1.09 (0.93 - 1.28) | 1.29 (0.97 - 1.71) | Reference | 0.90 (0.79 - 1.03) | 0.96 (0.84 - 1.09) | 1.05 (0.88 - 1.25) | 1.21 (0.89 - 1.63) | Reference |
| Age and nursing home adjusted HR (95% CI) | 0.86 (0.75 - 0.97) | 0.89 (0.78 - 1.01) | 1.05 (0.89 - 1.24) | 1.25 (0.94 - 1.66) | Reference | 0.76 (0.67 - 0.87) | 0.92 (0.81 - 1.05) | 1.02 (0.86 - 1.22) | 1.19 (0.88 - 1.61) | Reference |
| Age, nursing home and year of last infection adjusted HR (95% CI) | 0.85 (0.75 - 0.96) | 0.88 (0.78 - 0.99) | 1.08 (0.92 - 1.28) | 1.32 (0.99 - 1.76) | Reference | 0.76 (0.67 - 0.87) | 0.92 (0.81 - 1.05) | 1.04 (0.87 - 1.23) | 1.22 (0.89 - 1.66) | Reference |

| **Table S24:** Hazard ratios (HR) with 95% confidence intervals (95% CI) for non-COVID 19 and all-cause mortality according to number of SARS-CoV-2 vaccine doses during different time periods for **18-39 year olds** only. | | | | | | | | | | |
| --- | --- | --- | --- | --- | --- | --- | --- | --- | --- | --- |
|  | 2021 | | | | | | | | | |
|  | June and July 2021 (low COVID-19 disease burden) | | | | | October and November 2021 (high COVID-19 disease burden) | | | | |
|  | Four or more vaccine doses | Three vaccine doses | Two vaccine doses | One vaccine dose | Unvaccinated | Four or more vaccine doses | Three vaccine doses | Two vaccine doses | One vaccine dose | Unvaccinated |
|  | All-cause mortality | | | | | All-cause mortality | | | | |
| Deaths (n) | 0 | 0 | 3* | 0 | 8 | 0 | 0 | 3* | 2* | 2 |
| Events per 100,000 person days |  |  | 0.21 |  | 0.14 |  |  | 0.07 | 0.05 | 0.04 |
| Crude HR (95%CI) |  |  |  |  | Reference |  |  |  |  | Reference |
| Age adjusted HR (95% CI) |  |  |  |  | Reference |  |  |  |  | Reference |
| Age and gender adjusted HR (95% CI) |  |  |  |  | Reference |  |  |  |  | Reference |
| Age, gender and nursing home adjusted HR (95% CI) |  |  |  |  | Reference |  |  |  |  | Reference |
| Age, gender, nursing home and year of last infection adjusted HR (95% CI) |  |  |  |  | Reference |  |  |  |  | Reference |
|  | Non-COVID-19 mortality | | | | | Non-COVID-19 mortality | | | | |
| Deaths (n) | 0 | 0 | 3 | 0 | 7 | 0 | 0 | 3* | 2* | 2 |
| Events per 100.000 person days |  |  | 0.21 |  | 0.12 |  |  | 0.07 | 0.05 | 0.04 |
| Crude HR (95%CI) |  |  | 1.40 (0.35 - 5.64) |  | Reference |  |  |  |  | Reference |
| Age adjusted HR (95% CI) |  |  | 1.39 (0.34 - 5.58) |  | Reference |  |  |  |  | Reference |
| Age and gender adjusted HR (95% CI) |  |  | 1.38 (0.34 - 5.59) |  | Reference |  |  |  |  | Reference |
| Age, gender and nursing home adjusted HR (95% CI) |  |  | 1.38 (0.34 - 5.59) |  | Reference |  |  |  |  | Reference |
| Age, gender, nursing home and year of last infection adjusted HR (95% CI) | | | 1.27 (0.31 - 5.17) |  | Reference |  |  |  |  | Reference |
|  | May and June 2022 (low COVID-19 disease burden) | | | | | February and March 2022 (high COVID-19 disease burden) | | | | |
|  | Four or more vaccine doses | Three vaccine doses | Two vaccine doses | One vaccine dose | Unvaccinated | Four or more vaccine doses | Three vaccine doses | Two vaccine doses | One vaccine dose | Unvaccinated |
|  | All-cause mortality | | | | | All-cause mortality | | | | |
| Deaths (n) | 0 | 5 | 7 | 3 | 9 | 0 | 4 | 5 | 3 | 2 |
| Events per 100,000 person days |  | 0.06 | 0.07 | 0.1 | 0.08 |  | 0.1 | 0.1 | 0.11 | 0.05 |
| Crude HR (95%CI) |  | 0.81 (0.27 - 2.41) | 0.90 (0.34 - 2.42) | 1.31 (0.35 - 4.84) | Reference |  | 2.08 (0.38 - 11.35) | 1.90 (0.37 - 9.82) | 2.23 (0.37 - 13.32) | Reference |
| Age adjusted HR (95% CI) |  | 0.80 (0.27 - 2.38) | 0.92 (0.34 - 2.48) | 1.33 (0.36 - 4.92) | Reference |  | 2.08 (0.38 - 11.35) | 1.94 (0.38 - 9.98) | 2.15 (0.36 - 12.86) | Reference |
| Age and gender adjusted HR (95% CI) |  | 0.79 (0.27 - 2.37) | 0.92 (0.34 - 2.47) | 1.33 (0.36 - 4.92) | Reference |  | 2.06 (0.38 - 11.27) | 1.92 (0.37 - 9.90) | 2.14 (0.36 - 12.85) | Reference |
| Age, gender and nursing home adjusted HR (95% CI) |  | 0.79 (0.27 - 2.37) | 0.92 (0.34 - 2.47) | 1.33 (0.36 - 4.92) | Reference |  | 2.06 (0.38 - 11.27) | 1.92 (0.37 - 9.90) | 2.14 (0.36 - 12.85) | Reference |
| Age, gender, nursing home and year of last infection adjusted HR (95% CI) |  | 0.71 (0.23 - 2.18) | 0.77 (0.28 - 2.12) | 1.14 (0.29 - 4.43) | Reference |  | 1.08 (0.18 - 6.43) | 1.84 (0.35 - 9.79) | 2.05 (0.34 - 12.37) | Reference |
|  | Non-COVID-19 mortality | | | | | Non-COVID-19 mortality | | | | |
| Deaths (n) | 0 | 5 | 7 | 3 | 9 | 0 | 4* | 5* | 3* | 2 |
| Events per 100,000 person days |  | 0.06 | 0.07 | 0.1 | 0.08 |  | 0.1 | 0.1 | 0.11 | 0.05 |
| Crude HR (95%CI) |  | 0.81 (0.27 - 2.41) | 0.90 (0.34 - 2.42) | 1.31 (0.35 - 4.84) | Reference |  |  |  |  | Reference |
| Age adjusted HR (95% CI) |  | 0.80 (0.27 - 2.38) | 0.92 (0.34 - 2.48) | 1.33 (0.36 - 4.92) | Reference |  |  |  |  | Reference |
| Age and gender adjusted HR (95% CI) |  | 0.79 (0.27 - 2.37) | 0.92 (0.34 - 2.47) | 1.33 (0.36 - 4.92) | Reference |  |  |  |  | Reference |
| Age, gender and nursing home adjusted HR (95% CI) |  | 0.79 (0.27 - 2.37) | 0.92 (0.34 - 2.47) | 1.33 (0.36 - 4.92) | Reference |  |  |  |  | Reference |
| Age, gender, nursing home and year of last infection adjusted HR (95% CI) | | 0.71 (0.23 - 2.18) | 0.77 (0.28 - 2.12) | 1.14 (0.29 - 4.43) | Reference |  |  |  |  | Reference |

| **Table S24 continued:** | | | | | | | | | | |
| --- | --- | --- | --- | --- | --- | --- | --- | --- | --- | --- |
|  | July and August 2023 (low COVID-19 disease burden) | | | | | February and March 2023 (high COVID-19 disease burden) | | | | |
|  | Four or more vaccine doses | Three vaccine doses | Two vaccine doses | One vaccine dose | Unvaccinated | Four or more vaccine doses | Three vaccine doses | Two vaccine doses | One vaccine dose | Unvaccinated |
|  | All-cause mortality | | | | | All-cause mortality | | | | |
| Deaths (n) | 20 | 55 | 33 | 10 | 41 | 8 | 35 | 20 | 5 | 23 |
| Events per 100,000 person days | 0.25 | 0.13 | 0.14 | 0.21 | 0.16 | 0.12 | 0.09 | 0.09 | 0.12 | 0.10 |
| Crude HR (95%CI) | 1.59 (0.93 - 2.71) | 0.81 (0.54 - 1.21) | 0.89 (0.56 - 1.40) | 1.35 (0.68 - 2.69) | Reference | 1.27 (0.57 - 2.84) | 0.97 (0.58 - 1.65) | 0.97 (0.53 - 1.77) | 1.20 (0.46 - 3.16) | Reference |
| Age adjusted HR (95% CI) | 1.45 (0.85 - 2.49) | 0.81 (0.54 - 1.21) | 0.91 (0.58 - 1.45) | 1.43 (0.72 - 2.86) | Reference | 1.26 (0.56 - 2.82) | 0.97 (0.58 - 1.65) | 1.00 (0.55 - 1.83) | 1.20 (0.46 - 3.16) | Reference |
| Age and gender adjusted HR (95% CI) | 1.46 (0.85 - 2.50) | 0.81 (0.54 - 1.22) | 0.91 (0.57 - 1.44) | 1.42 (0.71 - 2.83) | Reference | 1.28 (0.57 - 2.87) | 0.98 (0.58 - 1.65) | 1.00 (0.55 - 1.82) | 1.19 (0.45 - 3.13) | Reference |
| Age, gender and nursing home adjusted HR (95% CI) | 1.46 (0.85 - 2.50) | 0.81 (0.54 - 1.22) | 0.91 (0.57 - 1.44) | 1.42 (0.71 - 2.83) | Reference | 1.28 (0.57 - 2.87) | 0.98 (0.58 - 1.65) | 1.00 (0.55 - 1.82) | 1.19 (0.45 - 3.13) | Reference |
| Age, gender, nursing home and year of last infection adjusted HR (95% CI) | 1.52 (0.88 - 2.64) | 0.84 (0.56 - 1.26) | 0.91 (0.57 - 1.44) | 1.37 (0.68 - 2.76) | Reference | 1.32 (0.58 - 2.98) | 0.98 (0.58 - 1.67) | 1.00 (0.55 - 1.82) | 1.24 (0.46 - 3.31) | Reference |
|  | Non-COVID-19 mortality | | | | | Non-COVID-19 mortality | | | | |
| Deaths (n) | 20 | 55 | 33 | 10 | 41 | 8 | 35 | 20 | 5 | 23 |
| Events per 100,000 person days | 0.25 | 0.13 | 0.14 | 0.21 | 0.16 | 0.12 | 0.09 | 0.09 | 0.12 | 0.10 |
| Crude HR (95%CI) | 1.59 (0.93 - 2.71) | 0.81 (0.54 - 1.21) | 0.89 (0.56 - 1.40) | 1.35 (0.68 - 2.69) | Reference | 1.27 (0.57 - 2.84) | 0.97 (0.58 - 1.65) | 0.97 (0.53 - 1.77) | 1.20 (0.46 - 3.16) | Reference |
| Age adjusted HR (95% CI) | 1.45 (0.85 - 2.49) | 0.81 (0.54 - 1.21) | 0.91 (0.58 - 1.45) | 1.43 (0.72 - 2.86) | Reference | 1.26 (0.56 - 2.82) | 0.97 (0.58 - 1.65) | 1.00 (0.55 - 1.83) | 1.20 (0.46 - 3.16) | Reference |
| Age and gender adjusted HR (95% CI) | 1.46 (0.85 - 2.50) | 0.81 (0.54 - 1.22) | 0.91 (0.57 - 1.44) | 1.42 (0.71 - 2.83) | Reference | 1.28 (0.57 - 2.87) | 0.98 (0.58 - 1.65) | 1.00 (0.55 - 1.82) | 1.19 (0.45 - 3.13) | Reference |
| Age, gender and nursing home adjusted HR (95% CI) | 1.46 (0.85 - 2.50) | 0.81 (0.54 - 1.22) | 0.91 (0.57 - 1.44) | 1.42 (0.71 - 2.83) | Reference | 1.28 (0.57 - 2.87) | 0.98 (0.58 - 1.65) | 1.00 (0.55 - 1.82) | 1.19 (0.45 - 3.13) | Reference |
| Age, gender, nursing home and year of last infection adjusted HR (95% CI) | 1.52 (0.88 - 2.64) | 0.84 (0.56 - 1.26) | 0.91 (0.57 - 1.44) | 1.37 (0.68 - 2.76) | Reference | 1.32 (0.58 - 2.98) | 0.98 (0.58 - 1.67) | 1.00 (0.55 - 1.82) | 1.24 (0.46 - 3.31) | Reference |
| * HRs not reported because there were less than 10 events total (including reference group) | | | | | | | | | | |

| **Table S25:** Hazard ratios (HR) with 95% confidence intervals (95% CI) for non-COVID 19 and all-cause mortality according to number of SARS-CoV-2 vaccine doses during different time periods for **40-59 year olds** only. | | | | | | | | | | |
| --- | --- | --- | --- | --- | --- | --- | --- | --- | --- | --- |
|  | 2021 | | | | | | | | | |
|  | June and July 2021 (low COVID-19 disease burden) | | | | | October and November 2021 (high COVID-19 disease burden) | | | | |
|  | Four or more vaccine doses | Three vaccine doses | Two vaccine doses | One vaccine dose | Unvaccinated | Four or more vaccine doses | Three vaccine doses | Two vaccine doses | One vaccine dose | Unvaccinated |
|  | All-cause mortality | | | | | All-cause mortality | | | | |
| Deaths (n) | 0 | 0 | 13 | 8 | 29 | 0 | 0 | 23 | 16 | 32 |
| Events per 100,000 person days |  |  | 0.61 | 0.26 | 0.70 |  |  | 0.47 | 0.4 | 0.89 |
| Crude HR (95%CI) |  |  | 0.81 (0.42 - 1.59) | 0.35 (0.16 - 0.77) | Reference |  |  | 0.51 (0.30 - 0.88) | 0.46 (0.25 - 0.83) | Reference |
| Age adjusted HR (95% CI) |  |  | 0.73 (0.37 - 1.43) | 0.32 (0.15 - 0.70) | Reference |  |  | 0.46 (0.27 - 0.79) | 0.43 (0.23 - 0.78) | Reference |
| Age and gender adjusted HR (95% CI) |  |  | 0.74 (0.38 - 1.44) | 0.30 (0.14 - 0.67) | Reference |  |  | 0.45 (0.26 - 0.77) | 0.42 (0.23 - 0.76) | Reference |
| Age, gender and nursing home adjusted HR (95% CI) |  |  | 0.74 (0.38 - 1.44) | 0.30 (0.14 - 0.67) | Reference |  |  | 0.45 (0.26 - 0.77) | 0.42 (0.23 - 0.76) | Reference |
| Age, gender, nursing home and year of last infection adjusted HR (95% CI) |  |  | 0.73 (0.37 - 1.44) | 0.30 (0.14 - 0.67) | Reference |  |  | 0.46 (0.26 - 0.79) | 0.42 (0.23 - 0.77) | Reference |
|  | Non-COVID-19 mortality | | | | | Non-COVID-19 mortality | | | | |
| Deaths (n) | 0 | 0 | 13 | 8 | 28 | 0 | 0 | 22 | 16 | 30 |
| Events per 100,000 person days |  |  | 0.61 | 0.26 | 0.68 |  |  | 0.45 | 0.4 | 0.83 |
| Crude HR (95%CI) |  |  | 0.84 (0.43 - 1.64) | 0.37 (0.17 - 0.80) | Reference |  |  | 0.52 (0.30 - 0.91) | 0.49 (0.26 - 0.89) | Reference |
| Age adjusted HR (95% CI) |  |  | 0.74 (0.38 - 1.46) | 0.33 (0.15 - 0.73) | Reference |  |  | 0.47 (0.27 - 0.82) | 0.46 (0.25 - 0.84) | Reference |
| Age and gender adjusted HR (95% CI) |  |  | 0.75 (0.38 - 1.48) | 0.31 (0.14 - 0.69) | Reference |  |  | 0.46 (0.26 - 0.80) | 0.45 (0.24 - 0.82) | Reference |
| Age, gender and nursing home adjusted HR (95% CI) |  |  | 0.75 (0.38 - 1.48) | 0.31 (0.14 - 0.69) | Reference |  |  | 0.46 (0.26 - 0.80) | 0.45 (0.24 - 0.82) | Reference |
| Age, gender, nursing home and year of last infection adjusted HR (95% CI) | | | 0.75 (0.38 - 1.49) | 0.31 (0.14 - 0.69) | Reference |  |  | 0.46 (0.26 - 0.81) | 0.45 (0.24 - 0.83) | Reference |
|  | May and June 2022 (low COVID-19 disease burden) | | | | | February and March 2022 (high COVID-19 disease burden) | | | | |
|  | Four or more vaccine doses | Three vaccine doses | Two vaccine doses | One vaccine dose | Unvaccinated | Four or more vaccine doses | Three vaccine doses | Two vaccine doses | One vaccine dose | Unvaccinated |
|  | All-cause mortality | | | | | All-cause mortality | | | | |
| Deaths (n) | 2 | 44 | 27 | 10 | 32 | 0 | 24 | 20 | 7 | 22 |
| Events per 100,000 person days | 11.84 | 0.47 | 0.34 | 0.52 | 0.37 |  | 0.53 | 0.4 | 0.37 | 0.76 |
| Crude HR (95%CI) | 28.87 (6.87 - 121.32) | 1.27 (0.80 - 2.00) | 0.90 (0.54 - 1.50) | 1.40 (0.69 - 2.85) | Reference |  | 0.69 (0.39 - 1.23) | 0.53 (0.29 - 0.97) | 0.49 (0.21 - 1.14) | Reference |
| Age adjusted HR (95% CI) | 24.26 (5.73 - 102.69) | 1.15 (0.73 - 1.81) | 0.87 (0.52 - 1.45) | 1.39 (0.68 - 2.83) | Reference |  | 0.60 (0.34 - 1.08) | 0.49 (0.27 - 0.91) | 0.48 (0.21 - 1.13) | Reference |
| Age and gender adjusted HR (95% CI) | 23.57 (5.56 - 99.91) | 1.13 (0.72 - 1.79) | 0.86 (0.51 - 1.43) | 1.38 (0.68 - 2.80) | Reference |  | 0.59 (0.33 - 1.05) | 0.49 (0.27 - 0.90) | 0.48 (0.21 - 1.12) | Reference |
| Age, gender and nursing home adjusted HR (95% CI) | 23.63 (5.57 - 100.16) | 1.13 (0.72 - 1.79) | 0.86 (0.51 - 1.43) | 1.38 (0.68 - 2.80) | Reference |  | 0.59 (0.33 - 1.05) | 0.49 (0.27 - 0.90) | 0.48 (0.21 - 1.12) | Reference |
| Age, gender, nursing home and year of last infection adjusted HR (95% CI) | 26.47 (6.15 - 113.97) | 1.12 (0.70 - 1.79) | 0.85 (0.50 - 1.44) | 1.47 (0.70 - 3.10) | Reference |  | 0.62 (0.33 - 1.16) | 0.49 (0.27 - 0.92) | 0.47 (0.20 - 1.10) | Reference |
|  | Non-COVID-19 mortality | | | | | Non-COVID-19 mortality | | | | |
| Deaths (n) | 2 | 44 | 27 | 10 | 32 | 0 | 24 | 20 | 6 | 19 |
| Events per 100,000 person days | 11.84 | 0.47 | 0.34 | 0.52 | 0.37 |  | 0.53 | 0.4 | 0.32 | 0.66 |
| Crude HR (95%CI) | 28.87 (6.87 - 121.32) | 1.27 (0.80 - 2.00) | 0.90 (0.54 - 1.50) | 1.40 (0.69 - 2.85) | Reference |  | 0.80 (0.44 - 1.46) | 0.61 (0.33 - 1.15) | 0.48 (0.19 - 1.21) | Reference |
| Age adjusted HR (95% CI) | 24.26 (5.73 - 102.69) | 1.15 (0.73 - 1.81) | 0.87 (0.52 - 1.45) | 1.39 (0.68 - 2.83) | Reference |  | 0.69 (0.38 - 1.27) | 0.57 (0.30 - 1.07) | 0.48 (0.19 - 1.20) | Reference |
| Age and gender adjusted HR (95% CI) | 23.57 (5.56 - 99.91) | 1.13 (0.72 - 1.79) | 0.86 (0.51 - 1.43) | 1.38 (0.68 - 2.80) | Reference |  | 0.68 (0.37 - 1.24) | 0.56 (0.30 - 1.06) | 0.47 (0.19 - 1.19) | Reference |
| Age, gender and nursing home adjusted HR (95% CI) | 23.63 (5.57 - 100.16) | 1.13 (0.72 - 1.79) | 0.86 (0.51 - 1.43) | 1.38 (0.68 - 2.80) | Reference |  | 0.68 (0.37 - 1.24) | 0.56 (0.30 - 1.06) | 0.47 (0.19 - 1.19) | Reference |
| Age, gender, nursing home and year of last infection adjusted HR (95% CI) | 26.47 (6.15 - 113.97) | 1.12 (0.70 - 1.79) | 0.85 (0.50 - 1.44) | 1.47 (0.70 - 3.10) | Reference |  | 0.69 (0.36 - 1.32) | 0.56 (0.30 - 1.06) | 0.46 (0.18 - 1.15) | Reference |

| **Table S25 continued:** | | | | | | | | | | |
| --- | --- | --- | --- | --- | --- | --- | --- | --- | --- | --- |
|  | July and August 2023 (low COVID-19 disease burden) | | | | | February and March 2023 (high COVID-19 disease burden) | | | | |
|  | Four or more vaccine doses | Three vaccine doses | Two vaccine doses | One vaccine dose | Unvaccinated | Four or more vaccine doses | Three vaccine doses | Two vaccine doses | One vaccine dose | Unvaccinated |
|  | All-cause mortality | | | | | All-cause mortality | | | | |
| Deaths (n) | 112 | 228 | 87 | 15 | 128 | 90 | 179 | 64 | 8 | 96 |
| Events per 100,000 person days | 0.75 | 0.53 | 0.59 | 0.55 | 0.65 | 0.76 | 0.49 | 0.49 | 0.32 | 0.53 |
| Crude HR (95%CI) | 1.16 (0.90 - 1.50) | 0.81 (0.65 - 1.01) | 0.92 (0.70 - 1.21) | 0.85 (0.50 - 1.45) | Reference | 1.42 (1.06 - 1.89) | 0.91 (0.71 - 1.17) | 0.91 (0.67 - 1.26) | 0.60 (0.29 - 1.24) | Reference |
| Age adjusted HR (95% CI) | 0.97 (0.75 - 1.25) | 0.77 (0.62 - 0.95) | 0.94 (0.72 - 1.24) | 0.88 (0.52 - 1.51) | Reference | 1.18 (0.88 - 1.57) | 0.87 (0.68 - 1.11) | 0.94 (0.69 - 1.29) | 0.62 (0.30 - 1.28) | Reference |
| Age and gender adjusted HR (95% CI) | 0.96 (0.74 - 1.24) | 0.77 (0.62 - 0.95) | 0.93 (0.71 - 1.22) | 0.88 (0.51 - 1.50) | Reference | 1.16 (0.87 - 1.56) | 0.87 (0.68 - 1.11) | 0.93 (0.67 - 1.27) | 0.62 (0.30 - 1.27) | Reference |
| Age, gender and nursing home adjusted HR (95% CI) | 0.96 (0.74 - 1.24) | 0.76 (0.61 - 0.95) | 0.93 (0.71 - 1.22) | 0.88 (0.51 - 1.50) | Reference | 1.14 (0.85 - 1.54) | 0.86 (0.67 - 1.11) | 0.93 (0.67 - 1.27) | 0.62 (0.30 - 1.27) | Reference |
| Age, gender, nursing home and year of last infection adjusted HR (95% CI) | 0.92 (0.70 - 1.19) | 0.75 (0.60 - 0.93) | 0.95 (0.72 - 1.24) | 0.99 (0.58 - 1.69) | Reference | 1.14 (0.85 - 1.53) | 0.86 (0.67 - 1.10) | 0.94 (0.68 - 1.30) | 0.67 (0.32 - 1.40) | Reference |
|  | Non-COVID-19 mortality | | | | | Non-COVID-19 mortality | | | | |
| Deaths (n) | 112 | 227 | 87 | 15 | 128 | 90 | 178 | 64 | 8 | 95 |
| Events per 100,000 person days | 0.75 | 0.52 | 0.59 | 0.55 | 0.65 | 0.76 | 0.48 | 0.49 | 0.32 | 0.53 |
| Crude HR (95%CI) | 1.16 (0.90 - 1.50) | 0.81 (0.65 - 1.01) | 0.92 (0.70 - 1.21) | 0.85 (0.50 - 1.45) | Reference | 1.43 (1.07 - 1.91) | 0.92 (0.72 - 1.18) | 0.92 (0.67 - 1.27) | 0.61 (0.30 - 1.26) | Reference |
| Age adjusted HR (95% CI) | 0.97 (0.75 - 1.25) | 0.77 (0.62 - 0.95) | 0.94 (0.72 - 1.24) | 0.88 (0.52 - 1.51) | Reference | 1.19 (0.89 - 1.59) | 0.87 (0.68 - 1.12) | 0.95 (0.69 - 1.30) | 0.63 (0.31 - 1.29) | Reference |
| Age and gender adjusted HR (95% CI) | 0.96 (0.74 - 1.24) | 0.76 (0.61 - 0.95) | 0.93 (0.71 - 1.22) | 0.88 (0.51 - 1.50) | Reference | 1.18 (0.88 - 1.58) | 0.87 (0.68 - 1.12) | 0.94 (0.68 - 1.28) | 0.62 (0.30 - 1.28) | Reference |
| Age, gender and nursing home adjusted HR (95% CI) | 0.96 (0.74 - 1.24) | 0.76 (0.61 - 0.94) | 0.93 (0.71 - 1.22) | 0.88 (0.51 - 1.50) | Reference | 1.16 (0.86 - 1.56) | 0.87 (0.68 - 1.11) | 0.94 (0.68 - 1.29) | 0.62 (0.30 - 1.28) | Reference |
| Age, gender, nursing home and year of last infection adjusted HR (95% CI) | 0.92 (0.70 - 1.19) | 0.75 (0.60 - 0.93) | 0.95 (0.72 - 1.24) | 0.99 (0.58 - 1.69) | Reference | 1.15 (0.86 - 1.55) | 0.86 (0.67 - 1.11) | 0.95 (0.69 - 1.31) | 0.68 (0.32 - 1.41) | Reference |

| **Table S26:** Hazard ratios (HR) with 95% confidence intervals (95% CI) for non-COVID 19 and all-cause mortality according to number of SARS-CoV-2 vaccine doses during different time periods for **60-74 year olds** only. | | | | | | | | | | |
| --- | --- | --- | --- | --- | --- | --- | --- | --- | --- | --- |
|  | 2021 | | | | | | | | | |
|  | June and July 2021 (low COVID-19 disease burden) | | | | | October and November 2021 (high COVID-19 disease burden) | | | | |
|  | Four or more vaccine doses | Three vaccine doses | Two vaccine doses | One vaccine dose | Unvaccinated | Four or more vaccine doses | Three vaccine doses | Two vaccine doses | One vaccine dose | Unvaccinated |
|  | All-cause mortality | | | | | All-cause mortality | | | | |
| Deaths (n) | 0 | 0 | 48 | 25 | 93 | 0 | 12 | 68 | 40 | 65 |
| Events per 100,000 person days |  |  | 4.68 | 2.28 | 8.60 |  | 6.01 | 3.62 | 3.25 | 7.21 |
| Crude HR (95%CI) |  |  | 0.52 (0.37 - 0.74) | 0.26 (0.17 - 0.41) | Reference |  | 0.73 (0.38 - 1.41) | 0.49 (0.35 - 0.69) | 0.45 (0.30 - 0.67) | Reference |
| Age adjusted HR (95% CI) |  |  | 0.48 (0.33 - 0.68) | 0.26 (0.17 - 0.40) | Reference |  | 0.57 (0.30 - 1.10) | 0.45 (0.32 - 0.64) | 0.43 (0.29 - 0.64) | Reference |
| Age and gender adjusted HR (95% CI) |  |  | 0.46 (0.32 - 0.66) | 0.25 (0.16 - 0.39) | Reference |  | 0.55 (0.28 - 1.06) | 0.45 (0.32 - 0.63) | 0.42 (0.29 - 0.63) | Reference |
| Age, gender and nursing home adjusted HR (95% CI) |  |  | 0.35 (0.24 - 0.50) | 0.28 (0.18 - 0.43) | Reference |  | 0.24 (0.12 - 0.49) | 0.44 (0.31 - 0.62) | 0.47 (0.32 - 0.70) | Reference |
| Age, gender, nursing home and year of last infection adjusted HR (95% CI) |  |  | 0.34 (0.23 - 0.49) | 0.27 (0.17 - 0.42) | Reference |  | 0.25 (0.12 - 0.51) | 0.43 (0.30 - 0.61) | 0.48 (0.32 - 0.71) | Reference |
|  | Non-COVID-19 mortality | | | | | Non-COVID-19 mortality | | | | |
| Deaths (n) | 0 | 0 | 47 | 25 | 92 | 0 | 12 | 68 | 40 | 62 |
| Events per 100,000 person days |  |  | 4.58 | 2.28 | 8.51 |  | 6.01 | 3.62 | 3.25 | 6.88 |
| Crude HR (95%CI) |  |  | 0.52 (0.36 - 0.74) | 0.27 (0.17 - 0.41) | Reference |  | 0.79 (0.41 - 1.54) | 0.52 (0.37 - 0.73) | 0.47 (0.32 - 0.70) | Reference |
| Age adjusted HR (95% CI) |  |  | 0.47 (0.33 - 0.68) | 0.26 (0.17 - 0.41) | Reference |  | 0.61 (0.31 - 1.18) | 0.48 (0.34 - 0.67) | 0.45 (0.30 - 0.68) | Reference |
| Age and gender adjusted HR (95% CI) |  |  | 0.46 (0.32 - 0.65) | 0.26 (0.16 - 0.40) | Reference |  | 0.59 (0.30 - 1.14) | 0.47 (0.33 - 0.66) | 0.45 (0.30 - 0.67) | Reference |
| Age, gender and nursing home adjusted HR (95% CI) |  |  | 0.34 (0.24 - 0.50) | 0.28 (0.18 - 0.43) | Reference |  | 0.25 (0.13 - 0.52) | 0.46 (0.33 - 0.65) | 0.50 (0.33 - 0.74) | Reference |
| Age, gender, nursing home and year of last infection adjusted HR (95% CI) | | | 0.33 (0.23 - 0.48) | 0.27 (0.17 - 0.43) | Reference |  | 0.26 (0.13 - 0.54) | 0.45 (0.32 - 0.64) | 0.50 (0.34 - 0.75) | Reference |
|  | May and June 2022 (low COVID-19 disease burden) | | | | | February and March 2022 (high COVID-19 disease burden) | | | | |
|  | Four or more vaccine doses | Three vaccine doses | Two vaccine doses | One vaccine dose | Unvaccinated | Four or more vaccine doses | Three vaccine doses | Two vaccine doses | One vaccine dose | Unvaccinated |
|  | All-cause mortality | | | | | All-cause mortality | | | | |
| Deaths (n) | 2 | 120 | 106 | 15 | 56 | 0 | 66 | 70 | 16 | 41 |
| Events per 100,000 person days | 6.95 | 3.02 | 4.62 | 3.68 | 3.08 |  | 3.4 | 3.99 | 3.72 | 6.01 |
| Crude HR (95%CI) | 2.25 (0.55 - 9.29) | 0.98 (0.71 - 1.34) | 1.50 (1.09 - 2.08) | 1.20 (0.68 - 2.13) | Reference | 0.00 (0.00 - Inf) | 0.57 (0.38 - 0.84) | 0.67 (0.45 - 0.98) | 0.62 (0.35 - 1.10) | Reference |
| Age adjusted HR (95% CI) | 1.49 (0.36 - 6.18) | 0.85 (0.62 - 1.18) | 1.36 (0.98 - 1.88) | 1.14 (0.64 - 2.01) | Reference | 0.00 (0.00 - Inf) | 0.50 (0.34 - 0.75) | 0.61 (0.41 - 0.89) | 0.61 (0.34 - 1.08) | Reference |
| Age and gender adjusted HR (95% CI) | 1.40 (0.34 - 5.82) | 0.82 (0.59 - 1.13) | 1.32 (0.95 - 1.82) | 1.12 (0.63 - 1.99) | Reference | 0.00 (0.00 - Inf) | 0.49 (0.33 - 0.72) | 0.60 (0.40 - 0.88) | 0.60 (0.34 - 1.07) | Reference |
| Age, gender and nursing home adjusted HR (95% CI) | 0.45 (0.10 - 2.00) | 0.67 (0.48 - 0.93) | 1.26 (0.91 - 1.74) | 1.06 (0.60 - 1.87) | Reference | 0.00 (0.00 - Inf) | 0.40 (0.26 - 0.59) | 0.60 (0.41 - 0.88) | 0.62 (0.35 - 1.11) | Reference |
| Age, gender, nursing home and year of last infection adjusted HR (95% CI) | 0.42 (0.09 - 1.88) | 0.70 (0.50 - 0.98) | 1.18 (0.84 - 1.66) | 1.02 (0.57 - 1.83) | Reference | 0.00 (0.00 - Inf) | 0.40 (0.27 - 0.61) | 0.60 (0.41 - 0.89) | 0.62 (0.35 - 1.11) | Reference |
|  | Non-COVID-19 mortality | | | | | Non-COVID-19 mortality | | | | |
| Deaths (n) | 2 | 117 | 105 | 15 | 55 | 0 | 64 | 66 | 16 | 37 |
| Events per 100,000 person days | 6.95 | 2.95 | 4.58 | 3.68 | 3.02 |  | 3.3 | 3.77 | 3.72 | 5.42 |
| Crude HR (95%CI) | 2.28 (0.55 - 9.40) | 0.97 (0.71 - 1.34) | 1.52 (1.09 - 2.10) | 1.23 (0.69 - 2.17) | Reference | 0.00 (0.00 - Inf) | 0.61 (0.41 - 0.91) | 0.70 (0.47 - 1.04) | 0.69 (0.38 - 1.23) | Reference |
| Age adjusted HR (95% CI) | 1.49 (0.36 - 6.18) | 0.85 (0.61 - 1.17) | 1.37 (0.99 - 1.90) | 1.16 (0.65 - 2.05) | Reference | 0.00 (0.00 - Inf) | 0.55 (0.36 - 0.82) | 0.64 (0.43 - 0.96) | 0.67 (0.37 - 1.21) | Reference |
| Age and gender adjusted HR (95% CI) | 1.41 (0.34 - 5.85) | 0.81 (0.59 - 1.13) | 1.33 (0.96 - 1.84) | 1.14 (0.65 - 2.02) | Reference | 0.00 (0.00 - Inf) | 0.53 (0.35 - 0.79) | 0.63 (0.42 - 0.94) | 0.67 (0.37 - 1.20) | Reference |
| Age, gender and nursing home adjusted HR (95% CI) | 0.44 (0.10 - 1.99) | 0.67 (0.48 - 0.93) | 1.27 (0.91 - 1.76) | 1.08 (0.61 - 1.91) | Reference | 0.00 (0.00 - Inf) | 0.43 (0.29 - 0.66) | 0.63 (0.42 - 0.94) | 0.69 (0.38 - 1.24) | Reference |
| Age, gender, nursing home and year of last infection adjusted HR (95% CI) | 0.42 (0.09 - 1.88) | 0.69 (0.49 - 0.97) | 1.19 (0.84 - 1.68) | 1.04 (0.58 - 1.87) | Reference | 0.00 (0.00 - Inf) | 0.44 (0.29 - 0.68) | 0.63 (0.42 - 0.95) | 0.69 (0.38 - 1.24) | Reference |

| **Table S26 continued:** | | | | | | | | | | |
| --- | --- | --- | --- | --- | --- | --- | --- | --- | --- | --- |
|  | July and August 2023 (low COVID-19 disease burden) | | | | | February and March 2023 (high COVID-19 disease burden) | | | | |
|  | Four or more vaccine doses | Three vaccine doses | Two vaccine doses | One vaccine dose | Unvaccinated | Four or more vaccine doses | Three vaccine doses | Two vaccine doses | One vaccine dose | Unvaccinated |
|  | All-cause mortality | | | | | All-cause mortality | | | | |
| Deaths (n) | 475 | 475 | 133 | 22 | 154 | 362 | 427 | 114 | 19 | 144 |
| Events per 100,000 person days | 3.72 | 3.52 | 4.33 | 4.06 | 3.08 | 3.56 | 3.69 | 4.13 | 3.84 | 3.19 |
| Crude HR (95%CI) | 1.21 (1.01 - 1.45) | 1.14 (0.95 - 1.37) | 1.41 (1.12 - 1.78) | 1.32 (0.84 - 2.06) | Reference | 1.11 (0.92 - 1.35) | 1.16 (0.96 - 1.40) | 1.29 (1.01 - 1.65) | 1.20 (0.75 - 1.94) | Reference |
| Age adjusted HR (95% CI) | 1.03 (0.86 - 1.24) | 1.06 (0.88 - 1.27) | 1.38 (1.10 - 1.74) | 1.30 (0.83 - 2.04) | Reference | 0.92 (0.76 - 1.12) | 1.06 (0.88 - 1.29) | 1.26 (0.99 - 1.61) | 1.17 (0.73 - 1.89) | Reference |
| Age and gender adjusted HR (95% CI) | 0.99 (0.82 - 1.19) | 1.03 (0.86 - 1.24) | 1.36 (1.08 - 1.72) | 1.29 (0.82 - 2.01) | Reference | 0.87 (0.71 - 1.06) | 1.04 (0.86 - 1.25) | 1.24 (0.97 - 1.58) | 1.15 (0.71 - 1.86) | Reference |
| Age, gender and nursing home adjusted HR (95% CI) | 0.91 (0.76 - 1.10) | 1.02 (0.85 - 1.22) | 1.31 (1.04 - 1.66) | 1.24 (0.79 - 1.93) | Reference | 0.78 (0.64 - 0.95) | 1.01 (0.83 - 1.22) | 1.19 (0.93 - 1.53) | 1.09 (0.68 - 1.76) | Reference |
| Age, gender, nursing home and year of last infection adjusted HR (95% CI) | 0.89 (0.74 - 1.07) | 1.00 (0.83 - 1.20) | 1.39 (1.10 - 1.76) | 1.34 (0.85 - 2.12) | Reference | 0.78 (0.64 - 0.95) | 1.01 (0.83 - 1.22) | 1.25 (0.97 - 1.61) | 1.14 (0.70 - 1.87) | Reference |
|  | Non-COVID-19 mortality | | | | | Non-COVID-19 mortality | | | | |
| Deaths (n) | 475 | 473 | 133 | 22 | 152 | 355 | 418 | 112 | 19 | 142 |
| Events per 100,000 person days | 3.72 | 3.51 | 4.33 | 4.06 | 3.04 | 3.49 | 3.62 | 4.06 | 3.84 | 3.15 |
| Crude HR (95%CI) | 1.23 (1.02 - 1.47) | 1.15 (0.96 - 1.39) | 1.43 (1.13 - 1.80) | 1.34 (0.85 - 2.09) | Reference | 1.11 (0.91 - 1.34) | 1.15 (0.95 - 1.39) | 1.29 (1.00 - 1.65) | 1.22 (0.76 - 1.97) | Reference |
| Age adjusted HR (95% CI) | 1.04 (0.87 - 1.25) | 1.07 (0.89 - 1.28) | 1.40 (1.11 - 1.77) | 1.32 (0.84 - 2.06) | Reference | 0.92 (0.75 - 1.12) | 1.06 (0.87 - 1.28) | 1.26 (0.98 - 1.61) | 1.19 (0.74 - 1.92) | Reference |
| Age and gender adjusted HR (95% CI) | 1.00 (0.83 - 1.21) | 1.04 (0.87 - 1.25) | 1.38 (1.09 - 1.74) | 1.30 (0.83 - 2.04) | Reference | 0.87 (0.71 - 1.06) | 1.03 (0.85 - 1.25) | 1.24 (0.96 - 1.58) | 1.17 (0.72 - 1.88) | Reference |
| Age, gender and nursing home adjusted HR (95% CI) | 0.93 (0.77 - 1.12) | 1.03 (0.85 - 1.23) | 1.33 (1.05 - 1.68) | 1.25 (0.80 - 1.96) | Reference | 0.78 (0.64 - 0.95) | 1.00 (0.83 - 1.21) | 1.19 (0.93 - 1.52) | 1.11 (0.68 - 1.79) | Reference |
| Age, gender, nursing home and year of last infection adjusted HR (95% CI) | 0.90 (0.75 - 1.09) | 1.01 (0.84 - 1.21) | 1.41 (1.11 - 1.78) | 1.35 (0.85 - 2.13) | Reference | 0.78 (0.64 - 0.95) | 1.00 (0.83 - 1.21) | 1.24 (0.96 - 1.60) | 1.16 (0.71 - 1.90) | Reference |

| **Table S27:** Hazard ratios (HR) with 95% confidence intervals (95% CI) for non-COVID 19 and all-cause mortality according to number of SARS-CoV-2 vaccine doses during different time periods for **75-84 year olds** only. | | | | | | | | | | |
| --- | --- | --- | --- | --- | --- | --- | --- | --- | --- | --- |
|  | 2021 | | | | | | | | | |
|  | June and July 2021 (low COVID-19 disease burden) | | | | | October and November 2021 (high COVID-19 disease burden) | | | | |
|  | Four or more vaccine doses | Three vaccine doses | Two vaccine doses | One vaccine dose | Unvaccinated | Four or more vaccine doses | Three vaccine doses | Two vaccine doses | One vaccine dose | Unvaccinated |
|  | All-cause mortality | | | | | All-cause mortality | | | | |
| Deaths (n) | 0 | 0 | 112 | 54 | 98 | 0 | 72 | 110 | 63 | 66 |
| Events per 100,000 person days |  |  | 17.59 | 15.57 | 25.26 |  | 30.14 | 14.89 | 16.07 | 24.48 |
| Crude HR (95%CI) |  |  | 0.70 (0.53 - 0.91) | 0.61 (0.44 - 0.85) | Reference |  | 1.20 (0.85 - 1.70) | 0.60 (0.44 - 0.82) | 0.66 (0.46 - 0.93) | Reference |
| Age adjusted HR (95% CI) |  |  | 0.68 (0.52 - 0.90) | 0.63 (0.45 - 0.88) | Reference |  | 1.10 (0.77 - 1.56) | 0.62 (0.46 - 0.84) | 0.69 (0.49 - 0.98) | Reference |
| Age and gender adjusted HR (95% CI) |  |  | 0.68 (0.51 - 0.89) | 0.63 (0.45 - 0.87) | Reference |  | 1.10 (0.77 - 1.56) | 0.62 (0.46 - 0.85) | 0.68 (0.48 - 0.97) | Reference |
| Age, gender and nursing home adjusted HR (95% CI) |  |  | 0.54 (0.41 - 0.71) | 0.66 (0.48 - 0.93) | Reference |  | 0.73 (0.51 - 1.06) | 0.59 (0.44 - 0.80) | 0.77 (0.55 - 1.10) | Reference |
| Age, gender, nursing home and year of last infection adjusted HR (95% CI) |  |  | 0.58 (0.43 - 0.77) | 0.69 (0.49 - 0.96) | Reference |  | 0.72 (0.50 - 1.05) | 0.59 (0.43 - 0.80) | 0.77 (0.55 - 1.10) | Reference |
|  | Non-COVID-19 mortality | | | | | Non-COVID-19 mortality | | | | |
| Deaths (n) | 0 | 0 | 107 | 53 | 95 | 0 | 71 | 106 | 60 | 64 |
| Events per 100,000 person days |  |  | 16.8 | 15.28 | 24.49 |  | 29.72 | 14.35 | 15.31 | 23.74 |
| Crude HR (95%CI) |  |  | 0.68 (0.52 - 0.90) | 0.62 (0.44 - 0.86) | Reference |  | 1.22 (0.85 - 1.73) | 0.60 (0.44 - 0.82) | 0.64 (0.45 - 0.92) | Reference |
| Age adjusted HR (95% CI) |  |  | 0.67 (0.51 - 0.89) | 0.64 (0.46 - 0.90) | Reference |  | 1.11 (0.78 - 1.59) | 0.62 (0.45 - 0.84) | 0.68 (0.48 - 0.97) | Reference |
| Age and gender adjusted HR (95% CI) |  |  | 0.67 (0.50 - 0.88) | 0.63 (0.45 - 0.89) | Reference |  | 1.11 (0.78 - 1.59) | 0.62 (0.45 - 0.85) | 0.68 (0.48 - 0.96) | Reference |
| Age, gender and nursing home adjusted HR (95% CI) |  |  | 0.53 (0.40 - 0.70) | 0.67 (0.48 - 0.95) | Reference |  | 0.73 (0.50 - 1.06) | 0.59 (0.43 - 0.80) | 0.77 (0.54 - 1.10) | Reference |
| Age, gender, nursing home and year of last infection adjusted HR (95% CI) | | | 0.56 (0.42 - 0.75) | 0.69 (0.49 - 0.98) | Reference |  | 0.71 (0.49 - 1.04) | 0.59 (0.43 - 0.80) | 0.77 (0.54 - 1.10) | Reference |
|  | May and June 2022 (low COVID-19 disease burden) | | | | | February and March 2022 (high COVID-19 disease burden) | | | | |
|  | Four or more vaccine doses | Three vaccine doses | Two vaccine doses | One vaccine dose | Unvaccinated | Four or more vaccine doses | Three vaccine doses | Two vaccine doses | One vaccine dose | Unvaccinated |
|  | All-cause mortality | | | | | All-cause mortality | | | | |
| Deaths (n) | 11 | 220 | 123 | 16 | 48 | 2 | 148 | 104 | 26 | 35 |
| Events per 100,000 person days | 27.24 | 14.24 | 17.9 | 16.1 | 14.47 | 91.49 | 16.31 | 16.53 | 23.57 | 21.09 |
| Crude HR (95%CI) | 2.06 (1.05 - 4.03) | 0.98 (0.72 - 1.35) | 1.24 (0.88 - 1.72) | 1.11 (0.63 - 1.95) | Reference | 4.41 (1.06 - 18.38) | 0.77 (0.53 - 1.11) | 0.78 (0.53 - 1.15) | 1.12 (0.67 - 1.86) | Reference |
| Age adjusted HR (95% CI) | 1.77 (0.90 - 3.48) | 0.98 (0.72 - 1.34) | 1.24 (0.89 - 1.73) | 1.07 (0.61 - 1.89) | Reference | 4.30 (1.03 - 17.92) | 0.75 (0.52 - 1.09) | 0.79 (0.54 - 1.16) | 1.11 (0.67 - 1.85) | Reference |
| Age and gender adjusted HR (95% CI) | 1.83 (0.93 - 3.62) | 0.97 (0.71 - 1.33) | 1.22 (0.87 - 1.71) | 1.07 (0.61 - 1.89) | Reference | 4.11 (0.98 - 17.18) | 0.74 (0.51 - 1.08) | 0.77 (0.53 - 1.14) | 1.11 (0.67 - 1.85) | Reference |
| Age, gender and nursing home adjusted HR (95% CI) | 1.23 (0.60 - 2.52) | 0.81 (0.59 - 1.11) | 1.19 (0.85 - 1.67) | 1.08 (0.61 - 1.91) | Reference | 2.68 (0.63 - 11.40) | 0.59 (0.41 - 0.86) | 0.82 (0.56 - 1.20) | 1.29 (0.77 - 2.15) | Reference |
| Age, gender, nursing home and year of last infection adjusted HR (95% CI) | 1.16 (0.56 - 2.40) | 0.81 (0.59 - 1.12) | 1.18 (0.84 - 1.67) | 1.07 (0.60 - 1.92) | Reference | 2.65 (0.62 - 11.34) | 0.58 (0.40 - 0.84) | 0.82 (0.56 - 1.20) | 1.28 (0.77 - 2.14) | Reference |
|  | Non-COVID-19 mortality | | | | | Non-COVID-19 mortality | | | | |
| Deaths (n) | 11 | 218 | 123 | 16 | 47 | 2 | 140 | 99 | 25 | 33 |
| Events per 100,000 person days | 27.24 | 14.11 | 17.9 | 16.1 | 14.17 | 91.49 | 15.43 | 15.73 | 22.67 | 19.89 |
| Crude HR (95%CI) | 2.12 (1.08 - 4.15) | 1.00 (0.73 - 1.37) | 1.26 (0.90 - 1.77) | 1.13 (0.64 - 1.99) | Reference | 4.67 (1.12 - 19.51) | 0.77 (0.53 - 1.13) | 0.79 (0.53 - 1.17) | 1.14 (0.68 - 1.92) | Reference |
| Age adjusted HR (95% CI) | 1.83 (0.93 - 3.61) | 0.99 (0.72 - 1.36) | 1.26 (0.90 - 1.77) | 1.09 (0.62 - 1.93) | Reference | 4.54 (1.09 - 18.99) | 0.76 (0.52 - 1.11) | 0.80 (0.54 - 1.18) | 1.13 (0.67 - 1.91) | Reference |
| Age and gender adjusted HR (95% CI) | 1.91 (0.97 - 3.78) | 0.99 (0.72 - 1.35) | 1.25 (0.89 - 1.75) | 1.10 (0.62 - 1.94) | Reference | 4.34 (1.04 - 18.18) | 0.74 (0.51 - 1.09) | 0.78 (0.53 - 1.16) | 1.13 (0.67 - 1.90) | Reference |
| Age, gender and nursing home adjusted HR (95% CI) | 1.27 (0.62 - 2.61) | 0.82 (0.59 - 1.12) | 1.22 (0.87 - 1.71) | 1.11 (0.63 - 1.96) | Reference | 2.74 (0.64 - 11.70) | 0.59 (0.40 - 0.87) | 0.82 (0.55 - 1.22) | 1.31 (0.77 - 2.21) | Reference |
| Age, gender, nursing home and year of last infection adjusted HR (95% CI) | 1.20 (0.58 - 2.49) | 0.83 (0.60 - 1.14) | 1.21 (0.86 - 1.71) | 1.10 (0.62 - 1.96) | Reference | 2.72 (0.64 - 11.65) | 0.58 (0.40 - 0.86) | 0.82 (0.55 - 1.22) | 1.31 (0.77 - 2.21) | Reference |

| **Table S27 continued:** | | | | | | | | | | |
| --- | --- | --- | --- | --- | --- | --- | --- | --- | --- | --- |
|  | July and August 2023 (low COVID-19 disease burden) | | | | | February and March 2023 (high COVID-19 disease burden) | | | | |
|  | Four or more vaccine doses | Three vaccine doses | Two vaccine doses | One vaccine dose | Unvaccinated | Four or more vaccine doses | Three vaccine doses | Two vaccine doses | One vaccine dose | Unvaccinated |
|  | All-cause mortality | | | | | All-cause mortality | | | | |
| Deaths (n) | 829 | 631 | 128 | 29 | 141 | 650 | 539 | 126 | 19 | 146 |
| Events per 100,000 person days | 13.56 | 16.34 | 18.28 | 23.17 | 14.65 | 13.52 | 16.37 | 19.94 | 16.37 | 17.33 |
| Crude HR (95%CI) | 0.93 (0.77 - 1.11) | 1.12 (0.93 - 1.34) | 1.25 (0.98 - 1.59) | 1.58 (1.06 - 2.36) | Reference | 0.78 (0.65 - 0.93) | 0.94 (0.79 - 1.13) | 1.15 (0.91 - 1.46) | 0.94 (0.59 - 1.52) | Reference |
| Age adjusted HR (95% CI) | 0.92 (0.77 - 1.10) | 1.13 (0.94 - 1.35) | 1.25 (0.99 - 1.59) | 1.54 (1.03 - 2.30) | Reference | 0.78 (0.65 - 0.93) | 0.95 (0.79 - 1.14) | 1.15 (0.91 - 1.46) | 0.92 (0.57 - 1.49) | Reference |
| Age and gender adjusted HR (95% CI) | 0.90 (0.75 - 1.07) | 1.11 (0.92 - 1.33) | 1.24 (0.98 - 1.58) | 1.54 (1.03 - 2.30) | Reference | 0.75 (0.63 - 0.90) | 0.93 (0.77 - 1.11) | 1.14 (0.90 - 1.45) | 0.92 (0.57 - 1.49) | Reference |
| Age, gender and nursing home adjusted HR (95% CI) | 0.79 (0.66 - 0.95) | 1.08 (0.90 - 1.29) | 1.20 (0.94 - 1.52) | 1.47 (0.99 - 2.20) | Reference | 0.64 (0.54 - 0.77) | 0.89 (0.74 - 1.07) | 1.10 (0.87 - 1.40) | 0.87 (0.54 - 1.40) | Reference |
| Age, gender, nursing home and year of last infection adjusted HR (95% CI) | 0.79 (0.66 - 0.95) | 1.06 (0.89 - 1.28) | 1.26 (0.99 - 1.61) | 1.52 (1.01 - 2.29) | Reference | 0.64 (0.54 - 0.77) | 0.90 (0.75 - 1.08) | 1.09 (0.85 - 1.39) | 0.81 (0.50 - 1.31) | Reference |
|  | Non-COVID-19 mortality | | | | | Non-COVID-19 mortality | | | | |
| Deaths (n) | 827 | 628 | 128 | 28 | 139 | 632 | 527 | 123 | 19 | 142 |
| Events per 100,000 person days | 13.53 | 16.27 | 18.28 | 22.37 | 14.44 | 13.14 | 16.01 | 19.47 | 16.37 | 16.86 |
| Crude HR (95%CI) | 0.94 (0.78 - 1.12) | 1.13 (0.94 - 1.35) | 1.27 (1.00 - 1.61) | 1.55 (1.03 - 2.33) | Reference | 0.78 (0.65 - 0.93) | 0.95 (0.79 - 1.14) | 1.15 (0.91 - 1.47) | 0.97 (0.60 - 1.57) | Reference |
| Age adjusted HR (95% CI) | 0.93 (0.78 - 1.12) | 1.14 (0.95 - 1.37) | 1.27 (1.00 - 1.62) | 1.51 (1.01 - 2.27) | Reference | 0.78 (0.65 - 0.93) | 0.96 (0.79 - 1.15) | 1.16 (0.91 - 1.47) | 0.95 (0.59 - 1.53) | Reference |
| Age and gender adjusted HR (95% CI) | 0.91 (0.76 - 1.09) | 1.12 (0.93 - 1.34) | 1.26 (0.99 - 1.61) | 1.51 (1.01 - 2.27) | Reference | 0.75 (0.63 - 0.91) | 0.93 (0.77 - 1.12) | 1.15 (0.90 - 1.46) | 0.95 (0.59 - 1.53) | Reference |
| Age, gender and nursing home adjusted HR (95% CI) | 0.80 (0.67 - 0.96) | 1.09 (0.91 - 1.31) | 1.22 (0.96 - 1.55) | 1.44 (0.96 - 2.16) | Reference | 0.64 (0.54 - 0.77) | 0.90 (0.75 - 1.08) | 1.10 (0.87 - 1.41) | 0.90 (0.56 - 1.45) | Reference |
| Age, gender, nursing home and year of last infection adjusted HR (95% CI) | 0.80 (0.67 - 0.96) | 1.08 (0.89 - 1.29) | 1.28 (1.01 - 1.64) | 1.49 (0.98 - 2.25) | Reference | 0.65 (0.54 - 0.78) | 0.90 (0.75 - 1.09) | 1.09 (0.85 - 1.39) | 0.82 (0.51 - 1.34) | Reference |

| **Table S28:** Hazard ratios (HR) with 95% confidence intervals (95% CI) for non-COVID 19 and all-cause mortality according to number of SARS-CoV-2 vaccine doses during different time periods for **85+ year olds** only. | | | | | | | | | | |
| --- | --- | --- | --- | --- | --- | --- | --- | --- | --- | --- |
|  | 2021 | | | | | | | | | |
|  | June and July 2021 (low COVID-19 disease burden) | | | | | October and November 2021 (high COVID-19 disease burden) | | | | |
|  | Four or more vaccine doses | Three vaccine doses | Two vaccine doses | One vaccine dose | Unvaccinated | Four or more vaccine doses | Three vaccine doses | Two vaccine doses | One vaccine dose | Unvaccinated |
|  | All-cause mortality | | | | | All-cause mortality | | | | |
| Deaths (n) | 0 | 0 | 203 | 54 | 148 | 0 | 107 | 196 | 76 | 123 |
| Events per 100,000 person days |  |  | 48.49 | 48.93 | 70.78 |  | 51.32 | 67.43 | 59.2 | 82.23 |
| Crude HR (95%CI) |  |  | 0.69 (0.56 - 0.85) | 0.70 (0.51 - 0.96) | Reference |  | 0.62 (0.48 - 0.81) | 0.82 (0.65 - 1.03) | 0.72 (0.54 - 0.96) | Reference |
| Age adjusted HR (95% CI) |  |  | 0.69 (0.56 - 0.85) | 0.72 (0.53 - 0.98) | Reference |  | 0.62 (0.48 - 0.81) | 0.86 (0.69 - 1.08) | 0.77 (0.58 - 1.03) | Reference |
| Age and gender adjusted HR (95% CI) |  |  | 0.69 (0.56 - 0.85) | 0.71 (0.52 - 0.98) | Reference |  | 0.63 (0.48 - 0.82) | 0.86 (0.69 - 1.08) | 0.75 (0.57 - 1.01) | Reference |
| Age, gender and nursing home adjusted HR (95% CI) |  |  | 0.57 (0.46 - 0.71) | 0.70 (0.51 - 0.96) | Reference |  | 0.54 (0.41 - 0.71) | 0.77 (0.61 - 0.97) | 0.75 (0.57 - 1.01) | Reference |
| Age, gender, nursing home and year of last infection adjusted HR (95% CI) |  |  | 0.59 (0.48 - 0.74) | 0.70 (0.51 - 0.96) | Reference |  | 0.55 (0.42 - 0.73) | 0.77 (0.62 - 0.98) | 0.75 (0.56 - 1.00) | Reference |
|  | Non-COVID-19 mortality | | | | | Non-COVID-19 mortality | | | | |
| Deaths (n) | 0 | 0 | 199 | 53 | 145 | 0 | 106 | 191 | 75 | 117 |
| Events per 100,000 person days |  |  | 47.53 | 48.02 | 69.35 |  | 50.84 | 65.71 | 58.42 | 78.22 |
| Crude HR (95%CI) |  |  | 0.69 (0.56 - 0.86) | 0.70 (0.51 - 0.96) | Reference |  | 0.65 (0.50 - 0.85) | 0.84 (0.67 - 1.06) | 0.75 (0.56 - 1.00) | Reference |
| Age adjusted HR (95% CI) |  |  | 0.69 (0.56 - 0.85) | 0.72 (0.53 - 0.99) | Reference |  | 0.65 (0.50 - 0.85) | 0.88 (0.70 - 1.11) | 0.80 (0.60 - 1.07) | Reference |
| Age and gender adjusted HR (95% CI) |  |  | 0.69 (0.56 - 0.86) | 0.72 (0.52 - 0.98) | Reference |  | 0.66 (0.51 - 0.86) | 0.88 (0.70 - 1.11) | 0.78 (0.58 - 1.05) | Reference |
| Age, gender and nursing home adjusted HR (95% CI) |  |  | 0.57 (0.46 - 0.71) | 0.70 (0.51 - 0.97) | Reference |  | 0.56 (0.42 - 0.74) | 0.78 (0.62 - 0.99) | 0.78 (0.58 - 1.05) | Reference |
| Age, gender, nursing home and year of last infection adjusted HR (95% CI) | | | 0.59 (0.47 - 0.74) | 0.70 (0.51 - 0.96) | Reference |  | 0.57 (0.43 - 0.76) | 0.79 (0.62 - 1.00) | 0.77 (0.58 - 1.04) | Reference |
|  | May and June 2022 (low COVID-19 disease burden) | | | | | February and March 2022 (high COVID-19 disease burden) | | | | |
|  | Four or more vaccine doses | Three vaccine doses | Two vaccine doses | One vaccine dose | Unvaccinated | Four or more vaccine doses | Three vaccine doses | Two vaccine doses | One vaccine dose | Unvaccinated |
|  | All-cause mortality | | | | | All-cause mortality | | | | |
| Deaths (n) | 7 | 270 | 122 | 18 | 85 | 3 | 229 | 157 | 24 | 65 |
| Events per 100,000 person days | 30.95 | 51.53 | 53.9 | 45.4 | 65.12 | 346.02 | 55.57 | 71.69 | 55.46 | 75.19 |
| Crude HR (95%CI) | 0.43 (0.20 - 0.94) | 0.79 (0.62 - 1.01) | 0.83 (0.63 - 1.10) | 0.70 (0.42 - 1.16) | Reference | 4.40 (1.38 - 14.01) | 0.74 (0.56 - 0.97) | 0.95 (0.72 - 1.27) | 0.74 (0.46 - 1.18) | Reference |
| Age adjusted HR (95% CI) | 0.43 (0.20 - 0.93) | 0.81 (0.64 - 1.04) | 0.89 (0.67 - 1.17) | 0.73 (0.44 - 1.21) | Reference | 4.55 (1.43 - 14.51) | 0.78 (0.59 - 1.03) | 1.03 (0.77 - 1.38) | 0.79 (0.49 - 1.26) | Reference |
| Age and gender adjusted HR (95% CI) | 0.43 (0.20 - 0.93) | 0.81 (0.64 - 1.04) | 0.89 (0.67 - 1.18) | 0.73 (0.44 - 1.22) | Reference | 4.72 (1.48 - 15.06) | 0.78 (0.59 - 1.03) | 1.03 (0.77 - 1.37) | 0.76 (0.47 - 1.22) | Reference |
| Age, gender and nursing home adjusted HR (95% CI) | 0.34 (0.15 - 0.76) | 0.65 (0.50 - 0.83) | 0.84 (0.63 - 1.11) | 0.72 (0.43 - 1.19) | Reference | 3.63 (1.12 - 11.73) | 0.65 (0.49 - 0.86) | 0.99 (0.74 - 1.32) | 0.81 (0.50 - 1.30) | Reference |
| Age, gender, nursing home and year of last infection adjusted HR (95% CI) | 0.34 (0.15 - 0.76) | 0.65 (0.51 - 0.84) | 0.85 (0.64 - 1.12) | 0.72 (0.43 - 1.21) | Reference | 3.51 (1.08 - 11.38) | 0.65 (0.49 - 0.86) | 0.97 (0.73 - 1.30) | 0.80 (0.50 - 1.29) | Reference |
|  | Non-COVID-19 mortality | | | | | Non-COVID-19 mortality | | | | |
| Deaths (n) | 7 | 265 | 118 | 17 | 82 | 1 | 212 | 149 | 24 | 59 |
| Events per 100,000 person days | 30.95 | 50.58 | 52.13 | 42.88 | 62.82 | 115.34 | 51.45 | 68.03 | 55.46 | 68.25 |
| Crude HR (95%CI) | 0.45 (0.20 - 0.98) | 0.80 (0.63 - 1.03) | 0.83 (0.63 - 1.11) | 0.68 (0.41 - 1.15) | Reference | 1.63 (0.23 - 11.77) | 0.75 (0.56 - 1.00) | 1.00 (0.74 - 1.35) | 0.81 (0.51 - 1.31) | Reference |
| Age adjusted HR (95% CI) | 0.44 (0.20 - 0.97) | 0.83 (0.65 - 1.06) | 0.89 (0.67 - 1.18) | 0.71 (0.42 - 1.20) | Reference | 1.70 (0.23 - 12.25) | 0.80 (0.60 - 1.06) | 1.08 (0.79 - 1.46) | 0.87 (0.54 - 1.40) | Reference |
| Age and gender adjusted HR (95% CI) | 0.44 (0.20 - 0.97) | 0.83 (0.64 - 1.06) | 0.90 (0.67 - 1.19) | 0.72 (0.43 - 1.21) | Reference | 1.76 (0.24 - 12.73) | 0.79 (0.59 - 1.06) | 1.07 (0.79 - 1.45) | 0.83 (0.52 - 1.34) | Reference |
| Age, gender and nursing home adjusted HR (95% CI) | 0.34 (0.15 - 0.77) | 0.66 (0.51 - 0.85) | 0.84 (0.63 - 1.12) | 0.71 (0.42 - 1.19) | Reference | 1.33 (0.18 - 9.69) | 0.66 (0.49 - 0.89) | 1.03 (0.76 - 1.39) | 0.89 (0.55 - 1.44) | Reference |
| Age, gender, nursing home and year of last infection adjusted HR (95% CI) | 0.35 (0.15 - 0.78) | 0.67 (0.52 - 0.86) | 0.85 (0.64 - 1.14) | 0.71 (0.42 - 1.20) | Reference | 1.28 (0.18 - 9.35) | 0.67 (0.49 - 0.90) | 1.01 (0.75 - 1.37) | 0.89 (0.55 - 1.43) | Reference |

| **Table S28 continued:** | | | | | | | | | | |
| --- | --- | --- | --- | --- | --- | --- | --- | --- | --- | --- |
|  | July and August 2023 (low COVID-19 disease burden) | | | | | February and March 2023 (high COVID-19 disease burden) | | | | |
|  | Four or more vaccine doses | Three vaccine doses | Two vaccine doses | One vaccine dose | Unvaccinated | Four or more vaccine doses | Three vaccine doses | Two vaccine doses | One vaccine dose | Unvaccinated |
|  | All-cause mortality | | | | | All-cause mortality | | | | |
| Deaths (n) | 792 | 480 | 125 | 24 | 160 | 750 | 511 | 126 | 31 | 174 |
| Events per 100,000 person days | 52.73 | 51.67 | 58.2 | 57.1 | 52.57 | 60.56 | 62.55 | 63.49 | 77.12 | 63.37 |
| Crude HR (95%CI) | 1.00 (0.85 - 1.19) | 0.98 (0.82 - 1.18) | 1.11 (0.88 - 1.40) | 1.09 (0.71 - 1.67) | Reference | 0.96 (0.81 - 1.13) | 0.99 (0.83 - 1.17) | 1.00 (0.80 - 1.26) | 1.22 (0.83 - 1.79) | Reference |
| Age adjusted HR (95% CI) | 1.07 (0.90 - 1.27) | 1.06 (0.88 - 1.27) | 1.14 (0.90 - 1.44) | 1.10 (0.72 - 1.70) | Reference | 1.02 (0.86 - 1.20) | 1.05 (0.88 - 1.25) | 1.03 (0.82 - 1.30) | 1.24 (0.85 - 1.82) | Reference |
| Age and gender adjusted HR (95% CI) | 1.06 (0.90 - 1.26) | 1.04 (0.87 - 1.25) | 1.13 (0.89 - 1.42) | 1.10 (0.71 - 1.68) | Reference | 1.02 (0.86 - 1.20) | 1.04 (0.88 - 1.24) | 1.02 (0.81 - 1.29) | 1.24 (0.85 - 1.82) | Reference |
| Age, gender and nursing home adjusted HR (95% CI) | 0.85 (0.72 - 1.02) | 0.97 (0.81 - 1.17) | 1.09 (0.86 - 1.38) | 1.09 (0.71 - 1.67) | Reference | 0.80 (0.68 - 0.95) | 0.96 (0.81 - 1.15) | 0.99 (0.79 - 1.24) | 1.24 (0.85 - 1.82) | Reference |
| Age, gender, nursing home and year of last infection adjusted HR (95% CI) | 0.85 (0.71 - 1.01) | 0.97 (0.81 - 1.16) | 1.11 (0.88 - 1.40) | 1.12 (0.72 - 1.73) | Reference | 0.80 (0.68 - 0.95) | 0.96 (0.81 - 1.15) | 0.99 (0.78 - 1.25) | 1.28 (0.87 - 1.89) | Reference |
|  | Non-COVID-19 mortality | | | | | Non-COVID-19 mortality | | | | |
| Deaths (n) | 791 | 479 | 125 | 24 | 160 | 719 | 502 | 123 | 31 | 165 |
| Events per 100,000 person days | 52.66 | 51.56 | 58.2 | 57.1 | 52.57 | 58.06 | 61.44 | 61.98 | 77.12 | 60.09 |
| Crude HR (95%CI) | 1.00 (0.85 - 1.19) | 0.98 (0.82 - 1.17) | 1.11 (0.88 - 1.40) | 1.09 (0.71 - 1.67) | Reference | 0.97 (0.82 - 1.14) | 1.02 (0.86 - 1.22) | 1.03 (0.82 - 1.30) | 1.29 (0.88 - 1.89) | Reference |
| Age adjusted HR (95% CI) | 1.07 (0.90 - 1.26) | 1.06 (0.88 - 1.27) | 1.14 (0.90 - 1.44) | 1.10 (0.72 - 1.70) | Reference | 1.03 (0.87 - 1.22) | 1.09 (0.91 - 1.30) | 1.06 (0.84 - 1.34) | 1.31 (0.89 - 1.93) | Reference |
| Age and gender adjusted HR (95% CI) | 1.06 (0.90 - 1.26) | 1.04 (0.87 - 1.25) | 1.13 (0.89 - 1.42) | 1.10 (0.71 - 1.68) | Reference | 1.03 (0.87 - 1.22) | 1.08 (0.91 - 1.29) | 1.05 (0.83 - 1.33) | 1.31 (0.89 - 1.93) | Reference |
| Age, gender and nursing home adjusted HR (95% CI) | 0.85 (0.72 - 1.01) | 0.97 (0.81 - 1.17) | 1.09 (0.86 - 1.38) | 1.09 (0.71 - 1.67) | Reference | 0.82 (0.69 - 0.98) | 1.00 (0.84 - 1.20) | 1.02 (0.81 - 1.29) | 1.31 (0.89 - 1.92) | Reference |
| Age, gender, nursing home and year of last infection adjusted HR (95% CI) | 0.84 (0.71 - 1.01) | 0.97 (0.81 - 1.16) | 1.11 (0.88 - 1.40) | 1.12 (0.72 - 1.73) | Reference | 0.82 (0.69 - 0.97) | 1.00 (0.84 - 1.20) | 1.03 (0.81 - 1.31) | 1.37 (0.93 - 2.03) | Reference |

| **Table S29:** Hazard ratios (HR) with 95% confidence intervals (95% CI) for non-COVID 19 and all-cause mortality according to number of SARS-CoV-2 vaccine doses during different time periods for **nursing home residents** only. | | | | | | | | | | |
| --- | --- | --- | --- | --- | --- | --- | --- | --- | --- | --- |
|  | 2021 | | | | | | | | | |
|  | June and July 2021 (low COVID-19 disease burden) | | | | | October and November 2021 (high COVID-19 disease burden) | | | | |
|  | Four or more vaccine doses | Three vaccine doses | Two vaccine doses | One vaccine dose | Unvaccinated | Four or more vaccine doses | Three vaccine doses | Two vaccine doses | One vaccine dose | Unvaccinated |
|  | All-cause mortality | | | | | All-cause mortality | | | | |
| Deaths (n) | 0 | 0 | 266 | 51 | 118 | 0 | 152 | 207 | 81 | 83 |
| Events per 100,000 person days |  |  | 48.49 | 58.05 | 74.53 |  | 51.91 | 67.9 | 99.06 | 79.58 |
| Crude HR (95%CI) |  |  | 0.65 (0.53 - 0.81) | 0.78 (0.56 - 1.08) | Reference |  | 0.63 (0.48 - 0.82) | 0.86 (0.67 - 1.11) | 1.24 (0.91 - 1.69) | Reference |
| Age adjusted HR (95% CI) |  |  | 0.66 (0.53 - 0.82) | 0.79 (0.57 - 1.09) | Reference |  | 0.64 (0.49 - 0.84) | 0.89 (0.69 - 1.14) | 1.26 (0.92 - 1.71) | Reference |
| Age and gender adjusted HR (95% CI) |  |  | 0.66 (0.53 - 0.82) | 0.78 (0.56 - 1.09) | Reference |  | 0.63 (0.48 - 0.83) | 0.88 (0.68 - 1.14) | 1.24 (0.91 - 1.68) | Reference |
| Age, gender and year of last infection adjusted HR (95% CI) |  |  | 0.69 (0.56 - 0.86) | 0.77 (0.55 - 1.06) | Reference |  | 0.64 (0.48 - 0.83) | 0.89 (0.69 - 1.14) | 1.20 (0.88 - 1.63) | Reference |
|  | Non-COVID-19 mortality | | | | | Non-COVID-19 mortality | | | | |
| Deaths (n) | 0 | 0 | 260 | 50 | 118 | 0 | 152 | 203 | 81 | 81 |
| Events per 100,000 person days |  |  | 47.4 | 56.92 | 74.53 |  | 51.91 | 66.59 | 99.06 | 77.66 |
| Crude HR (95%CI) |  |  | 0.64 (0.51 - 0.79) | 0.76 (0.55 - 1.06) | Reference |  | 0.64 (0.49 - 0.84) | 0.86 (0.67 - 1.12) | 1.27 (0.93 - 1.73) | Reference |
| Age adjusted HR (95% CI) |  |  | 0.65 (0.52 - 0.81) | 0.77 (0.55 - 1.07) | Reference |  | 0.65 (0.50 - 0.86) | 0.89 (0.69 - 1.15) | 1.28 (0.94 - 1.75) | Reference |
| Age and gender adjusted HR (95% CI) |  |  | 0.65 (0.52 - 0.80) | 0.77 (0.55 - 1.07) | Reference |  | 0.65 (0.49 - 0.85) | 0.89 (0.68 - 1.15) | 1.27 (0.93 - 1.73) | Reference |
| Age, gender and year of last infection adjusted HR (95% CI) |  |  | 0.68 (0.54 - 0.84) | 0.75 (0.54 - 1.04) | Reference |  | 0.65 (0.50 - 0.86) | 0.89 (0.69 - 1.15) | 1.22 (0.90 - 1.67) | Reference |
|  | May and June 2022 (low COVID-19 disease burden) | | | | | February and March 2022 (high COVID-19 disease burden) | | | | |
|  | Four or more vaccine doses | Three vaccine doses | Two vaccine doses | One vaccine dose | Unvaccinated | Four or more vaccine doses | Three vaccine doses | Two vaccine doses | One vaccine dose | Unvaccinated |
|  | All-cause mortality | | | | | All-cause mortality | | | | |
| Deaths (n) | 14 | 308 | 95 | 13 | 49 | 4 | 288 | 134 | 20 | 53 |
| Events per 100,000 person days | 46.11 | 61.28 | 69.86 | 70.86 | 80.80 | 250.16 | 59.49 | 83.78 | 90.62 | 91.20 |
| Crude HR (95%CI) | 0.60 (0.32 - 1.11) | 0.76 (0.56 - 1.02) | 0.87 (0.61 - 1.22) | 0.87 (0.47 - 1.61) | Reference | 2.64 (0.95 - 7.30) | 0.65 (0.49 - 0.87) | 0.92 (0.67 - 1.26) | 0.99 (0.59 - 1.66) | Reference |
| Age adjusted HR (95% CI) | 0.59 (0.32 - 1.10) | 0.76 (0.57 - 1.03) | 0.88 (0.62 - 1.24) | 0.88 (0.48 - 1.62) | Reference | 2.69 (0.97 - 7.45) | 0.68 (0.50 - 0.91) | 0.96 (0.70 - 1.32) | 1.02 (0.61 - 1.71) | Reference |
| Age and gender adjusted HR (95% CI) | 0.60 (0.32 - 1.11) | 0.75 (0.55 - 1.01) | 0.87 (0.61 - 1.23) | 0.88 (0.47 - 1.61) | Reference | 2.77 (1.00 - 7.68) | 0.67 (0.50 - 0.89) | 0.92 (0.67 - 1.27) | 1.01 (0.60 - 1.69) | Reference |
| Age, gender and year of last infection adjusted HR (95% CI) | 0.61 (0.33 - 1.14) | 0.76 (0.56 - 1.03) | 0.87 (0.62 - 1.23) | 0.86 (0.47 - 1.60) | Reference | 2.83 (1.02 - 7.86) | 0.67 (0.50 - 0.89) | 0.91 (0.66 - 1.26) | 1.03 (0.61 - 1.74) | Reference |
|  | Non-COVID-19 mortality | | | | | Non-COVID-19 mortality | | | | |
| Deaths (n) | 14 | 303 | 93 | 12 | 49 | 2 | 265 | 125 | 19 | 50 |
| Events per 100,000 person days | 46.11 | 60.29 | 68.39 | 65.41 | 80.80 | 125.08 | 54.74 | 78.16 | 86.09 | 86.04 |
| Crude HR (95%CI) | 0.60 (0.32 - 1.11) | 0.75 (0.55 - 1.01) | 0.85 (0.60 - 1.20) | 0.81 (0.43 - 1.52) | Reference | 1.41 (0.34 - 5.78) | 0.64 (0.47 - 0.86) | 0.91 (0.66 - 1.26) | 1.00 (0.59 - 1.70) | Reference |
| Age adjusted HR (95% CI) | 0.59 (0.32 - 1.10) | 0.75 (0.56 - 1.02) | 0.86 (0.61 - 1.21) | 0.81 (0.43 - 1.52) | Reference | 1.43 (0.35 - 5.89) | 0.66 (0.49 - 0.89) | 0.95 (0.68 - 1.32) | 1.03 (0.60 - 1.74) | Reference |
| Age and gender adjusted HR (95% CI) | 0.60 (0.32 - 1.11) | 0.73 (0.54 - 0.99) | 0.85 (0.60 - 1.20) | 0.81 (0.43 - 1.52) | Reference | 1.48 (0.36 - 6.08) | 0.65 (0.48 - 0.88) | 0.91 (0.66 - 1.27) | 1.01 (0.60 - 1.72) | Reference |
| Age, gender and year of last infection adjusted HR (95% CI) | 0.61 (0.33 - 1.14) | 0.75 (0.55 - 1.02) | 0.85 (0.60 - 1.20) | 0.80 (0.43 - 1.51) | Reference | 1.54 (0.37 - 6.34) | 0.65 (0.48 - 0.88) | 0.89 (0.64 - 1.24) | 1.05 (0.61 - 1.78) | Reference |

| **Table S29 continued:** | | | | | | | | | | |
| --- | --- | --- | --- | --- | --- | --- | --- | --- | --- | --- |
|  | July and August 2023 (low COVID-19 disease burden) | | | | | February and March 2023 (high COVID-19 disease burden) | | | | |
|  | Four or more vaccine doses | Three vaccine doses | Two vaccine doses | One vaccine dose | Unvaccinated | Four or more vaccine doses | Three vaccine doses | Two vaccine doses | One vaccine dose | Unvaccinated |
|  | All-cause mortality | | | | | All-cause mortality | | | | |
| Deaths (n) | 843 | 394 | 86 | 12 | 109 | 821 | 433 | 91 | 18 | 109 |
| Events per 100,000 person days | 63.59 | 65.95 | 65.98 | 50.39 | 72.00 | 74.65 | 83.06 | 80 | 82.21 | 84.42 |
| Crude HR (95%CI) | 0.88 (0.72 - 1.08) | 0.92 (0.74 - 1.13) | 0.92 (0.69 - 1.22) | 0.70 (0.39 - 1.27) | Reference | 0.88 (0.72 - 1.08) | 0.98 (0.80 - 1.21) | 0.95 (0.72 - 1.25) | 0.97 (0.59 - 1.61) | Reference |
| Age adjusted HR (95% CI) | 0.88 (0.72 - 1.07) | 0.93 (0.75 - 1.15) | 0.93 (0.70 - 1.24) | 0.72 (0.40 - 1.30) | Reference | 0.89 (0.73 - 1.08) | 1.00 (0.81 - 1.23) | 0.97 (0.73 - 1.28) | 1.01 (0.62 - 1.67) | Reference |
| Age and gender adjusted HR (95% CI) | 0.86 (0.71 - 1.05) | 0.91 (0.73 - 1.12) | 0.93 (0.70 - 1.23) | 0.72 (0.40 - 1.30) | Reference | 0.87 (0.71 - 1.06) | 0.98 (0.80 - 1.21) | 0.95 (0.72 - 1.26) | 1.02 (0.62 - 1.69) | Reference |
| Age, gender and year of last infection adjusted HR (95% CI) | 0.86 (0.70 - 1.05) | 0.91 (0.73 - 1.12) | 0.93 (0.70 - 1.23) | 0.72 (0.40 - 1.31) | Reference | 0.87 (0.71 - 1.06) | 0.98 (0.80 - 1.21) | 0.95 (0.72 - 1.25) | 1.02 (0.62 - 1.69) | Reference |
|  | Non-COVID-19 mortality | | | | | Non-COVID-19 mortality | | | | |
| Deaths (n) | 843 | 392 | 86 | 12 | 108 | 778 | 421 | 88 | 18 | 103 |
| Events per 100,000 person days | 63.59 | 65.62 | 65.98 | 50.39 | 71.34 | 70.74 | 80.76 | 77.36 | 82.21 | 79.78 |
| Crude HR (95%CI) | 0.89 (0.73 - 1.09) | 0.92 (0.74 - 1.14) | 0.92 (0.70 - 1.23) | 0.71 (0.39 - 1.28) | Reference | 0.89 (0.72 - 1.09) | 1.01 (0.82 - 1.26) | 0.97 (0.73 - 1.29) | 1.03 (0.63 - 1.70) | Reference |
| Age adjusted HR (95% CI) | 0.88 (0.72 - 1.08) | 0.93 (0.75 - 1.15) | 0.94 (0.71 - 1.25) | 0.72 (0.40 - 1.32) | Reference | 0.89 (0.72 - 1.09) | 1.03 (0.83 - 1.28) | 0.99 (0.74 - 1.32) | 1.07 (0.65 - 1.77) | Reference |
| Age and gender adjusted HR (95% CI) | 0.87 (0.71 - 1.06) | 0.91 (0.74 - 1.13) | 0.94 (0.70 - 1.24) | 0.72 (0.40 - 1.32) | Reference | 0.88 (0.71 - 1.07) | 1.01 (0.82 - 1.26) | 0.97 (0.73 - 1.30) | 1.08 (0.65 - 1.78) | Reference |
| Age, gender and year of last infection adjusted HR (95% CI) | 0.87 (0.71 - 1.06) | 0.91 (0.74 - 1.13) | 0.94 (0.71 - 1.25) | 0.73 (0.40 - 1.33) | Reference | 0.87 (0.71 - 1.07) | 1.01 (0.82 - 1.26) | 0.97 (0.73 - 1.29) | 1.08 (0.66 - 1.79) | Reference |

| **Table S30:** Hazard ratios (HR) with 95% confidence intervals (95% CI) for non-COVID 19 and all-cause mortality according to number of SARS-CoV-2 vaccine doses during different time periods for **community dwelling (not in a nursing home)** individuals only. | | | | | | | | | | |
| --- | --- | --- | --- | --- | --- | --- | --- | --- | --- | --- |
|  | 2021 | | | | | | | | | |
|  | June and July 2021 (low COVID-19 disease burden) | | | | | October and November 2021 (high COVID-19 disease burden) | | | | |
|  | Four or more vaccine doses | Three vaccine doses | Two vaccine doses | One vaccine dose | Unvaccinated | Four or more vaccine doses | Three vaccine doses | Two vaccine doses | One vaccine dose | Unvaccinated |
|  | All-cause mortality | | | | | All-cause mortality | | | | |
| Deaths (n) | 0 | 0 | 113 | 90 | 258 | 0 | 39 | 193 | 116 | 206 |
| Events per 100,000 person days |  |  | 2.21 | 1.25 | 2.27 |  | 4.42 | 1.59 | 1.19 | 2.09 |
| Crude HR (95%CI) |  |  | 0.95 (0.75 - 1.18) | 0.54 (0.43 - 0.69) | Reference |  | 2.14 (1.47 - 3.10) | 0.75 (0.62 - 0.92) | 0.57 (0.45 - 0.72) | Reference |
| Age adjusted HR (95% CI) |  |  | 0.42 (0.34 - 0.53) | 0.45 (0.35 - 0.57) | Reference |  | 0.42 (0.29 - 0.61) | 0.50 (0.41 - 0.60) | 0.47 (0.38 - 0.60) | Reference |
| Age and gender adjusted HR (95% CI) |  |  | 0.41 (0.33 - 0.51) | 0.43 (0.34 - 0.55) | Reference |  | 0.41 (0.29 - 0.59) | 0.48 (0.39 - 0.59) | 0.46 (0.37 - 0.58) | Reference |
| Age, gender and year of last infection adjusted HR (95% CI) |  |  | 0.41 (0.33 - 0.51) | 0.43 (0.34 - 0.55) | Reference |  | 0.42 (0.29 - 0.61) | 0.48 (0.39 - 0.59) | 0.46 (0.37 - 0.58) | Reference |
|  | Non-COVID-19 mortality | | | | | Non-COVID-19 mortality | | | | |
| Deaths (n) | 0 | 0 | 109 | 89 | 249 | 0 | 37 | 187 | 112 | 195 |
| Events per 100,000 person days |  |  | 2.13 | 1.24 | 2.19 |  | 4.19 | 1.54 | 1.15 | 1.98 |
| Crude HR (95%CI) |  |  | 0.95 (0.75 - 1.19) | 0.56 (0.44 - 0.71) | Reference |  | 2.17 (1.48 - 3.18) | 0.77 (0.63 - 0.94) | 0.58 (0.46 - 0.74) | Reference |
| Age adjusted HR (95% CI) |  |  | 0.42 (0.34 - 0.53) | 0.46 (0.36 - 0.58) | Reference |  | 0.43 (0.30 - 0.62) | 0.51 (0.42 - 0.62) | 0.48 (0.38 - 0.61) | Reference |
| Age and gender adjusted HR (95% CI) |  |  | 0.41 (0.33 - 0.51) | 0.44 (0.35 - 0.56) | Reference |  | 0.42 (0.29 - 0.60) | 0.49 (0.40 - 0.60) | 0.47 (0.37 - 0.60) | Reference |
| Age, gender and year of last infection adjusted HR (95% CI) |  |  | 0.41 (0.32 - 0.52) | 0.44 (0.35 - 0.56) | Reference |  | 0.43 (0.29 - 0.62) | 0.49 (0.40 - 0.60) | 0.47 (0.37 - 0.60) | Reference |
|  | May and June 2022 (low COVID-19 disease burden) | | | | | February and March 2022 (high COVID-19 disease burden) | | | | |
|  | Four or more vaccine doses | Three vaccine doses | Two vaccine doses | One vaccine dose | Unvaccinated | Four or more vaccine doses | Three vaccine doses | Two vaccine doses | One vaccine dose | Unvaccinated |
|  | All-cause mortality | | | | | All-cause mortality | | | | |
| Deaths (n) | 8 | 351 | 292 | 49 | 181 | 1 | 184 | 222 | 56 | 112 |
| Events per 100,000 person days | 9.33 | 1.5 | 1.32 | 0.87 | 0.78 | 12.09 | 1.63 | 1.72 | 1.06 | 1.41 |
| Crude HR (95%CI) | 11.39 (5.59 - 23.21) | 1.93 (1.61 - 2.31) | 1.70 (1.41 - 2.05) | 1.11 (0.81 - 1.53) | Reference | 8.47 (1.18 - 60.67) | 1.15 (0.91 - 1.45) | 1.22 (0.97 - 1.53) | 0.75 (0.55 - 1.04) | Reference |
| Age adjusted HR (95% CI) | 0.89 (0.44 - 1.84) | 0.75 (0.63 - 0.90) | 1.12 (0.93 - 1.35) | 1.04 (0.76 - 1.43) | Reference | 1.99 (0.28 - 14.25) | 0.53 (0.42 - 0.67) | 0.74 (0.59 - 0.93) | 0.79 (0.57 - 1.09) | Reference |
| Age and gender adjusted HR (95% CI) | 0.86 (0.42 - 1.77) | 0.73 (0.61 - 0.87) | 1.09 (0.91 - 1.32) | 1.03 (0.75 - 1.42) | Reference | 1.78 (0.25 - 12.80) | 0.50 (0.40 - 0.64) | 0.73 (0.58 - 0.91) | 0.78 (0.57 - 1.07) | Reference |
| Age, gender and year of last infection adjusted HR (95% CI) | 0.83 (0.40 - 1.70) | 0.71 (0.59 - 0.86) | 1.06 (0.88 - 1.29) | 1.00 (0.72 - 1.39) | Reference | 1.95 (0.27 - 14.09) | 0.50 (0.39 - 0.64) | 0.74 (0.59 - 0.92) | 0.79 (0.57 - 1.09) | Reference |
|  | Non-COVID-19 mortality | | | | | Non-COVID-19 mortality | | | | |
| Deaths (n) | 8 | 346 | 289 | 49 | 176 | 1 | 180 | 214 | 55 | 100 |
| Events per 100,000 person days | 9.33 | 1.48 | 1.31 | 0.87 | 0.76 | 12.09 | 1.59 | 1.66 | 1.04 | 1.26 |
| Crude HR (95%CI) | 11.72 (5.75 - 23.89) | 1.95 (1.63 - 2.34) | 1.73 (1.44 - 2.09) | 1.15 (0.84 - 1.57) | Reference | 9.59 (1.34 - 68.81) | 1.26 (0.99 - 1.61) | 1.32 (1.04 - 1.67) | 0.83 (0.60 - 1.15) | Reference |
| Age adjusted HR (95% CI) | 0.93 (0.45 - 1.92) | 0.77 (0.64 - 0.92) | 1.14 (0.95 - 1.38) | 1.07 (0.78 - 1.47) | Reference | 2.25 (0.31 - 16.15) | 0.58 (0.45 - 0.74) | 0.80 (0.63 - 1.02) | 0.87 (0.62 - 1.20) | Reference |
| Age and gender adjusted HR (95% CI) | 0.91 (0.44 - 1.87) | 0.74 (0.62 - 0.89) | 1.12 (0.92 - 1.35) | 1.07 (0.78 - 1.46) | Reference | 2.03 (0.28 - 14.62) | 0.55 (0.43 - 0.71) | 0.79 (0.62 - 1.00) | 0.86 (0.62 - 1.19) | Reference |
| Age, gender and year of last infection adjusted HR (95% CI) | 0.87 (0.42 - 1.80) | 0.73 (0.60 - 0.88) | 1.09 (0.89 - 1.32) | 1.04 (0.75 - 1.44) | Reference | 2.20 (0.30 - 15.91) | 0.55 (0.43 - 0.71) | 0.79 (0.63 - 1.01) | 0.86 (0.62 - 1.20) | Reference |

| **Table S30 continued:** | | | | | | | | | | |
| --- | --- | --- | --- | --- | --- | --- | --- | --- | --- | --- |
|  | July and August 2023 (low COVID-19 disease burden) | | | | | February and March 2023 (high COVID-19 disease burden) | | | | |
|  | Four or more vaccine doses | Three vaccine doses | Two vaccine doses | One vaccine dose | Unvaccinated | Four or more vaccine doses | Three vaccine doses | Two vaccine doses | One vaccine dose | Unvaccinated |
|  | All-cause mortality | | | | | All-cause mortality | | | | |
| Deaths (n) | 1387 | 1476 | 424 | 89 | 518 | 1039 | 1259 | 361 | 64 | 477 |
| Events per 100,000 person days | 3.27 | 1.36 | 0.94 | 1.03 | 0.95 | 3.07 | 1.37 | 0.9 | 0.82 | 0.97 |
| Crude HR (95%CI) | 3.43 (3.10 - 3.79) | 1.43 (1.29 - 1.58) | 0.98 (0.86 - 1.12) | 1.08 (0.86 - 1.35) | Reference | 3.16 (2.83 - 3.52) | 1.41 (1.27 - 1.56) | 0.92 (0.80 - 1.06) | 0.84 (0.65 - 1.09) | Reference |
| Age adjusted HR (95% CI) | 0.88 (0.79 - 0.98) | 0.98 (0.89 - 1.08) | 1.20 (1.05 - 1.36) | 1.36 (1.08 - 1.70) | Reference | 0.75 (0.67 - 0.84) | 0.94 (0.85 - 1.05) | 1.11 (0.97 - 1.27) | 1.03 (0.79 - 1.34) | Reference |
| Age and gender adjusted HR (95% CI) | 0.84 (0.76 - 0.93) | 0.96 (0.87 - 1.06) | 1.18 (1.04 - 1.34) | 1.35 (1.08 - 1.69) | Reference | 0.71 (0.63 - 0.79) | 0.91 (0.82 - 1.02) | 1.09 (0.95 - 1.25) | 1.02 (0.79 - 1.33) | Reference |
| Age, gender and year of last infection adjusted HR (95% CI) | 0.82 (0.74 - 0.92) | 0.94 (0.85 - 1.04) | 1.23 (1.08 - 1.40) | 1.43 (1.14 - 1.80) | Reference | 0.71 (0.64 - 0.80) | 0.92 (0.82 - 1.02) | 1.12 (0.97 - 1.28) | 1.03 (0.79 - 1.35) | Reference |
|  | Non-COVID-19 mortality | | | | | Non-COVID-19 mortality | | | | |
| Deaths (n) | 1384 | 1471 | 424 | 88 | 515 | 1026 | 1240 | 356 | 64 | 467 |
| Events per 100,000 person days | 3.27 | 1.36 | 0.94 | 1.02 | 0.95 | 3.04 | 1.35 | 0.89 | 0.82 | 0.95 |
| Crude HR (95%CI) | 3.44 (3.11 - 3.81) | 1.43 (1.30 - 1.58) | 0.99 (0.87 - 1.12) | 1.07 (0.85 - 1.34) | Reference | 3.18 (2.85 - 3.55) | 1.41 (1.27 - 1.57) | 0.93 (0.81 - 1.07) | 0.86 (0.66 - 1.11) | Reference |
| Age adjusted HR (95% CI) | 0.88 (0.80 - 0.98) | 0.98 (0.89 - 1.09) | 1.20 (1.06 - 1.37) | 1.35 (1.08 - 1.69) | Reference | 0.76 (0.68 - 0.85) | 0.95 (0.85 - 1.05) | 1.12 (0.97 - 1.28) | 1.05 (0.81 - 1.37) | Reference |
| Age and gender adjusted HR (95% CI) | 0.84 (0.76 - 0.94) | 0.96 (0.87 - 1.06) | 1.19 (1.05 - 1.35) | 1.34 (1.07 - 1.68) | Reference | 0.72 (0.64 - 0.80) | 0.92 (0.83 - 1.02) | 1.10 (0.96 - 1.26) | 1.04 (0.80 - 1.36) | Reference |
| Age, gender and year of last infection adjusted HR (95% CI) | 0.83 (0.74 - 0.92) | 0.94 (0.85 - 1.04) | 1.24 (1.09 - 1.41) | 1.42 (1.13 - 1.79) | Reference | 0.72 (0.64 - 0.81) | 0.92 (0.83 - 1.03) | 1.12 (0.98 - 1.29) | 1.06 (0.81 - 1.39) | Reference |

| **Table S31:** Hazard ratios (HR) with 95% confidence intervals (95% CI) for non-COVID 19 and all-cause mortality according to number of SARS-CoV-2 vaccine doses during different time periods for individuals **with an infection in the observation period** only. | | | | | | | | | | |
| --- | --- | --- | --- | --- | --- | --- | --- | --- | --- | --- |
|  | 2021 | | | | | | | | | |
|  | June and July 2021 (low COVID-19 disease burden) | | | | | October and November 2021 (high COVID-19 disease burden) | | | | |
|  | Four or more vaccine doses | Three vaccine doses | Two vaccine doses | One vaccine dose | Unvaccinated | Four or more vaccine doses | Three vaccine doses | Two vaccine doses | One vaccine dose | Unvaccinated |
|  | All-cause mortality | | | | | All-cause mortality | | | | |
| Deaths (n) |  |  | 2* |  | 2 |  | 2* | 7 | 3 | 7 |
| Events per 100,000 person days |  |  | 196.46 |  | 29.62 |  | 60.98 | 12.96 | 8.7 | 2.77 |
| Crude HR (95%CI) |  |  |  |  | Reference |  |  | 4.60 (1.61 - 13.11) | 2.95 (0.76 - 11.44) | Reference |
| Age adjusted HR (95% CI) |  |  |  |  | Reference |  |  | 0.96 (0.33 - 2.78) | 1.03 (0.27 - 4.00) | Reference |
| Age and gender adjusted HR (95% CI) |  |  |  |  | Reference |  |  | 1.15 (0.40 - 3.33) | 1.57 (0.38 - 6.39) | Reference |
| Age, gender and nursing home adjusted HR (95% CI) |  |  |  |  | Reference |  |  | 1.15 (0.40 - 3.32) | 1.43 (0.35 - 5.88) | Reference |
| Age, gender, nursing home and year of last infection adjusted HR (95% CI) |  |  |  |  | Reference |  |  | 1.12 (0.39 - 3.25) | 1.47 (0.35 - 6.19) | Reference |
|  | Non-COVID-19 mortality | | | | | Non-COVID-19 mortality | | | | |
| Deaths (n) |  |  | 1* |  | 2 |  | 1* | 2* | 2* | 1 |
| Events per 100,000 person days |  |  | 98.23 |  | 29.62 |  | 30.49 | 3.7 | 5.8 | 0.40 |
| Crude HR (95%CI) |  |  |  |  | Reference |  |  |  |  | Reference |
| Age adjusted HR (95% CI) |  |  |  |  | Reference |  |  |  |  | Reference |
| Age and gender adjusted HR (95% CI) |  |  |  |  | Reference |  |  |  |  | Reference |
| Age, gender and nursing home adjusted HR (95% CI) |  |  |  |  | Reference |  |  |  |  | Reference |
| Age, gender, nursing home and year of last infection adjusted HR (95% CI) | | |  |  | Reference |  |  |  |  | Reference |
|  | May and June 2022 (low COVID-19 disease burden) | | | | | February and March 2022 (high COVID-19 disease burden) | | | | |
|  | Four or more vaccine doses | Three vaccine doses | Two vaccine doses | One vaccine dose | Unvaccinated | Four or more vaccine doses | Three vaccine doses | Two vaccine doses | One vaccine dose | Unvaccinated |
|  | All-cause mortality | | | | | All-cause mortality | | | | |
| Deaths (n) |  | 15 | 10 | 4* | 4 | 2 | 45 | 19 | 6 | 27 |
| Events per 100,000 person days |  | 2.94 | 2.11 | 2.51 | 0.65 | 155.76 | 2.82 | 1.05 | 0.6 | 1.14 |
| Crude HR (95%CI) |  | 4.51 (1.50 - 13.60) | 3.26 (1.02 - 10.39) |  | Reference | 125.23 (29.77 - 526.85) | 2.43 (1.51 - 3.92) | 0.93 (0.51 - 1.66) | 0.53 (0.22 - 1.28) | Reference |
| Age adjusted HR (95% CI) |  | 1.30 (0.43 - 3.93) | 2.13 (0.66 - 6.86) |  | Reference | 16.54 (3.92 - 69.80) | 0.67 (0.41 - 1.09) | 0.63 (0.35 - 1.14) | 0.79 (0.33 - 1.93) | Reference |
| Age and gender adjusted HR (95% CI) |  | 1.23 (0.41 - 3.72) | 2.11 (0.66 - 6.78) |  | Reference | 12.90 (3.01 - 55.22) | 0.66 (0.40 - 1.07) | 0.62 (0.34 - 1.11) | 0.76 (0.31 - 1.84) | Reference |
| Age, gender and nursing home adjusted HR (95% CI) |  | 1.12 (0.37 - 3.44) | 2.14 (0.67 - 6.89) |  | Reference | 11.66 (2.69 - 50.48) | 0.53 (0.32 - 0.89) | 0.60 (0.33 - 1.09) | 0.75 (0.31 - 1.83) | Reference |
| Age, gender, nursing home and year of last infection adjusted HR (95% CI) |  | 1.27 (0.41 - 3.94) | 2.01 (0.60 - 6.70) |  | Reference | 11.01 (2.48 - 48.95) | 0.53 (0.32 - 0.89) | 0.60 (0.33 - 1.09) | 0.74 (0.30 - 1.80) | Reference |
|  | Non-COVID-19 mortality | | | | | Non-COVID-19 mortality | | | | |
| Deaths (n) |  | 10 | 8 | 4* | 3 |  | 21 | 9 | 5 | 16 |
| Events per 100,000 person days |  | 1.96 | 1.69 | 2.51 | 0.49 |  | 1.31 | 0.5 | 0.5 | 0.67 |
| Crude HR (95%CI) |  | 4.01 (1.10 - 14.57) | 3.48 (0.92 - 13.10) |  | Reference |  | 1.91 (1.00 - 3.67) | 0.74 (0.33 - 1.67) | 0.74 (0.27 - 2.03) | Reference |
| Age adjusted HR (95% CI) |  | 1.17 (0.32 - 4.27) | 2.43 (0.64 - 9.28) |  | Reference |  | 0.53 (0.27 - 1.04) | 0.51 (0.22 - 1.16) | 1.08 (0.39 - 2.96) | Reference |
| Age and gender adjusted HR (95% CI) |  | 1.08 (0.30 - 3.95) | 2.40 (0.63 - 9.17) |  | Reference |  | 0.52 (0.27 - 1.01) | 0.49 (0.22 - 1.12) | 1.02 (0.37 - 2.79) | Reference |
| Age, gender and nursing home adjusted HR (95% CI) |  | 0.94 (0.25 - 3.51) | 2.42 (0.64 - 9.25) |  | Reference |  | 0.41 (0.21 - 0.83) | 0.48 (0.21 - 1.11) | 1.00 (0.36 - 2.74) | Reference |
| Age, gender, nursing home and year of last infection adjusted HR (95% CI) | | 1.18 (0.31 - 4.51) | 2.46 (0.61 - 9.89) |  | Reference |  | 0.41 (0.21 - 0.83) | 0.48 (0.21 - 1.10) | 0.99 (0.36 - 2.72) | Reference |

| **Table S31 continued:** | | | | | | | | | | |
| --- | --- | --- | --- | --- | --- | --- | --- | --- | --- | --- |
|  | July and August 2023 (low COVID-19 disease burden) | | | | | February and March 2023 (high COVID-19 disease burden) | | | | |
|  | Four or more vaccine doses | Three vaccine doses | Two vaccine doses | One vaccine dose | Unvaccinated | Four or more vaccine doses | Three vaccine doses | Two vaccine doses | One vaccine dose | Unvaccinated |
|  | All-cause mortality | | | | | All-cause mortality | | | | |
| Deaths (n) |  |  |  |  | 0 | 127 | 94 | 24 | 2 | 33 |
| Events per 100,000 person days |  |  |  |  |  | 10.06 | 2.88 | 2.12 | 1.04 | 4.83 |
| Crude HR (95%CI) |  |  |  |  | Reference | 2.08 (1.42 - 3.05) | 0.60 (0.40 - 0.89) | 0.44 (0.26 - 0.74) | 0.22 (0.05 - 0.90) | Reference |
| Age adjusted HR (95% CI) |  |  |  |  | Reference | 0.59 (0.40 - 0.86) | 0.66 (0.44 - 0.98) | 0.69 (0.41 - 1.17) | 0.31 (0.07 - 1.31) | Reference |
| Age and gender adjusted HR (95% CI) |  |  |  |  | Reference | 0.55 (0.37 - 0.81) | 0.62 (0.42 - 0.92) | 0.65 (0.38 - 1.10) | 0.31 (0.07 - 1.29) | Reference |
| Age, gender and nursing home adjusted HR (95% CI) |  |  |  |  | Reference | 0.52 (0.35 - 0.77) | 0.61 (0.41 - 0.91) | 0.64 (0.38 - 1.09) | 0.31 (0.07 - 1.29) | Reference |
| Age, gender, nursing home and year of last infection adjusted HR (95% CI) |  |  |  |  | Reference | 0.52 (0.35 - 0.77) | 0.62 (0.42 - 0.92) | 0.62 (0.36 - 1.07) | 0.29 (0.07 - 1.23) | Reference |
|  | Non-COVID-19 mortality | | | | | Non-COVID-19 mortality | | | | |
| Deaths (n) |  |  |  |  | 0 | 81 | 67 | 19 | 2 | 17 |
| Events per 100,000 person days |  |  |  |  |  | 6.42 | 2.05 | 1.67 | 1.04 | 2.49 |
| Crude HR (95%CI) |  |  |  |  | Reference | 2.57 (1.53 - 4.34) | 0.83 (0.49 - 1.41) | 0.67 (0.35 - 1.30) | 0.42 (0.10 - 1.81) | Reference |
| Age adjusted HR (95% CI) |  |  |  |  | Reference | 0.76 (0.45 - 1.30) | 0.90 (0.53 - 1.53) | 1.04 (0.54 - 2.00) | 0.58 (0.13 - 2.51) | Reference |
| Age and gender adjusted HR (95% CI) |  |  |  |  | Reference | 0.71 (0.42 - 1.21) | 0.85 (0.50 - 1.46) | 0.98 (0.50 - 1.89) | 0.57 (0.13 - 2.47) | Reference |
| Age, gender and nursing home adjusted HR (95% CI) |  |  |  |  | Reference | 0.70 (0.41 - 1.19) | 0.85 (0.50 - 1.45) | 0.97 (0.50 - 1.87) | 0.57 (0.13 - 2.46) | Reference |
| Age, gender, nursing home and year of last infection adjusted HR (95% CI) | | |  |  | Reference | 0.70 (0.41 - 1.19) | 0.85 (0.50 - 1.46) | 0.96 (0.49 - 1.90) | 0.54 (0.12 - 2.40) | Reference |
| * HRs not reported because there were less than 10 events total (including reference group) | | | | | | | | | | |
| Note that the offical documentation of infections stopped on July 30th 2023. | | | | | | | | | | |

| **Table S32:** Hazard ratios (HR) with 95% confidence intervals (95% CI) for non-COVID 19 and all-cause mortality according to number of SARS-CoV-2 vaccine doses during different time periods for individuals **with no infections in the observation period** only. | | | | | | | | | | |
| --- | --- | --- | --- | --- | --- | --- | --- | --- | --- | --- |
|  | 2021 | | | | | | | | | |
|  | June and July 2021 (low COVID-19 disease burden) | | | | | October and November 2021 (high COVID-19 disease burden) | | | | |
|  | Four or more vaccine doses | Three vaccine doses | Two vaccine doses | One vaccine dose | Unvaccinated | Four or more vaccine doses | Three vaccine doses | Two vaccine doses | One vaccine dose | Unvaccinated |
|  | All-cause mortality | | | | | All-cause mortality | | | | |
| Deaths (n) |  |  | 377 | 141 | 374 |  | 189 | 393 | 194 | 282 |
| Events per 100,000 person days |  |  | 6.65 | 1.94 | 3.24 |  | 16.13 | 3.18 | 1.98 | 2.90 |
| Crude HR (95%CI) |  |  | 2.13 (1.84 - 2.47) | 0.59 (0.49 - 0.72) | Reference |  | 6.16 (5.04 - 7.53) | 1.09 (0.94 - 1.27) | 0.69 (0.57 - 0.83) | Reference |
| Age adjusted HR (95% CI) |  |  | 0.66 (0.57 - 0.77) | 0.52 (0.43 - 0.64) | Reference |  | 0.74 (0.61 - 0.91) | 0.67 (0.58 - 0.78) | 0.63 (0.53 - 0.76) | Reference |
| Age and gender adjusted HR (95% CI) |  |  | 0.66 (0.57 - 0.77) | 0.51 (0.42 - 0.62) | Reference |  | 0.74 (0.61 - 0.91) | 0.67 (0.57 - 0.78) | 0.62 (0.51 - 0.74) | Reference |
| Age, gender and nursing home adjusted HR (95% CI) |  |  | 0.55 (0.47 - 0.64) | 0.52 (0.43 - 0.63) | Reference |  | 0.57 (0.46 - 0.70) | 0.62 (0.53 - 0.72) | 0.65 (0.54 - 0.78) | Reference |
| Age, gender, nursing home and year of last infection adjusted HR (95% CI) |  |  | 0.57 (0.49 - 0.66) | 0.53 (0.43 - 0.64) | Reference |  | 0.58 (0.47 - 0.72) | 0.63 (0.54 - 0.73) | 0.65 (0.54 - 0.78) | Reference |
|  | Non-COVID-19 mortality | | | | | Non-COVID-19 mortality | | | | |
| Deaths (n) |  |  | 368 | 139 | 365 |  | 188 | 388 | 191 | 275 |
| Events per 100,000 person days |  |  | 6.49 | 1.91 | 3.16 |  | 16.04 | 3.14 | 1.95 | 2.83 |
| Crude HR (95%CI) |  |  | 2.14 (1.84 - 2.48) | 0.60 (0.49 - 0.73) | Reference |  | 6.31 (5.15 - 7.72) | 1.10 (0.95 - 1.29) | 0.70 (0.58 - 0.84) | Reference |
| Age adjusted HR (95% CI) |  |  | 0.66 (0.57 - 0.77) | 0.53 (0.44 - 0.65) | Reference |  | 0.76 (0.62 - 0.92) | 0.68 (0.58 - 0.80) | 0.64 (0.53 - 0.77) | Reference |
| Age and gender adjusted HR (95% CI) |  |  | 0.66 (0.57 - 0.77) | 0.52 (0.42 - 0.63) | Reference |  | 0.76 (0.62 - 0.92) | 0.68 (0.58 - 0.79) | 0.62 (0.52 - 0.75) | Reference |
| Age, gender and nursing home adjusted HR (95% CI) |  |  | 0.55 (0.47 - 0.64) | 0.53 (0.43 - 0.64) | Reference |  | 0.57 (0.47 - 0.71) | 0.63 (0.54 - 0.73) | 0.66 (0.55 - 0.79) | Reference |
| Age, gender, nursing home and year of last infection adjusted HR (95% CI) | | | 0.56 (0.48 - 0.66) | 0.53 (0.44 - 0.65) | Reference |  | 0.58 (0.47 - 0.72) | 0.63 (0.54 - 0.74) | 0.66 (0.54 - 0.79) | Reference |
|  | May and June 2022 (low COVID-19 disease burden) | | | | | February and March 2022 (high COVID-19 disease burden) | | | | |
|  | Four or more vaccine doses | Three vaccine doses | Two vaccine doses | One vaccine dose | Unvaccinated | Four or more vaccine doses | Three vaccine doses | Two vaccine doses | One vaccine dose | Unvaccinated |
|  | All-cause mortality | | | | | All-cause mortality | | | | |
| Deaths (n) | 22 | 644 | 377 | 58 | 226 | 3 | 427 | 337 | 70 | 138 |
| Events per 100,000 person days | 19.19 | 2.75 | 1.73 | 1.05 | 0.99 | 34.94 | 4.19 | 3 | 1.64 | 2.46 |
| Crude HR (95%CI) | 18.69 (12.01 - 29.10) | 2.77 (2.38 - 3.22) | 1.74 (1.48 - 2.06) | 1.06 (0.79 - 1.41) | Reference | 14.21 (4.52 - 44.64) | 1.71 (1.41 - 2.07) | 1.22 (1.00 - 1.49) | 0.66 (0.50 - 0.88) | Reference |
| Age adjusted HR (95% CI) | 1.01 (0.64 - 1.60) | 0.90 (0.77 - 1.05) | 1.09 (0.92 - 1.29) | 0.96 (0.72 - 1.29) | Reference | 2.45 (0.78 - 7.71) | 0.71 (0.59 - 0.87) | 0.83 (0.68 - 1.01) | 0.80 (0.60 - 1.06) | Reference |
| Age and gender adjusted HR (95% CI) | 1.01 (0.64 - 1.59) | 0.88 (0.76 - 1.03) | 1.07 (0.91 - 1.27) | 0.96 (0.72 - 1.28) | Reference | 2.43 (0.77 - 7.65) | 0.70 (0.58 - 0.85) | 0.81 (0.67 - 0.99) | 0.78 (0.59 - 1.05) | Reference |
| Age, gender and nursing home adjusted HR (95% CI) | 0.69 (0.43 - 1.11) | 0.77 (0.66 - 0.90) | 1.04 (0.88 - 1.23) | 0.95 (0.71 - 1.27) | Reference | 1.70 (0.54 - 5.39) | 0.59 (0.48 - 0.72) | 0.82 (0.67 - 1.00) | 0.84 (0.63 - 1.13) | Reference |
| Age, gender, nursing home and year of last infection adjusted HR (95% CI) | 0.67 (0.42 - 1.08) | 0.78 (0.66 - 0.91) | 1.03 (0.87 - 1.22) | 0.95 (0.71 - 1.27) | Reference | 1.70 (0.54 - 5.39) | 0.59 (0.48 - 0.72) | 0.82 (0.67 - 1.00) | 0.84 (0.63 - 1.13) | Reference |
|  | Non-COVID-19 mortality | | | | | Non-COVID-19 mortality | | | | |
| Deaths (n) | 22 | 639 | 374 | 57 | 222 | 3 | 424 | 330 | 69 | 134 |
| Events per 100,000 person days | 19.19 | 2.73 | 1.72 | 1.03 | 0.98 | 34.94 | 4.16 | 2.94 | 1.61 | 2.39 |
| Crude HR (95%CI) | 18.99 (12.19 - 29.57) | 2.80 (2.40 - 3.26) | 1.76 (1.49 - 2.08) | 1.06 (0.79 - 1.41) | Reference | 14.67 (4.67 - 46.10) | 1.74 (1.44 - 2.12) | 1.23 (1.01 - 1.50) | 0.67 (0.50 - 0.90) | Reference |
| Age adjusted HR (95% CI) | 1.04 (0.66 - 1.63) | 0.91 (0.78 - 1.06) | 1.10 (0.93 - 1.30) | 0.96 (0.72 - 1.29) | Reference | 2.52 (0.80 - 7.93) | 0.73 (0.60 - 0.89) | 0.83 (0.68 - 1.02) | 0.81 (0.61 - 1.08) | Reference |
| Age and gender adjusted HR (95% CI) | 1.03 (0.66 - 1.63) | 0.90 (0.77 - 1.05) | 1.09 (0.92 - 1.28) | 0.96 (0.72 - 1.29) | Reference | 2.50 (0.79 - 7.87) | 0.72 (0.59 - 0.87) | 0.82 (0.67 - 1.00) | 0.80 (0.60 - 1.07) | Reference |
| Age, gender and nursing home adjusted HR (95% CI) | 0.70 (0.43 - 1.12) | 0.78 (0.67 - 0.91) | 1.06 (0.89 - 1.25) | 0.96 (0.71 - 1.28) | Reference | 1.73 (0.55 - 5.48) | 0.60 (0.49 - 0.73) | 0.83 (0.67 - 1.01) | 0.86 (0.64 - 1.15) | Reference |
| Age, gender, nursing home and year of last infection adjusted HR (95% CI) | 0.68 (0.42 - 1.10) | 0.79 (0.67 - 0.92) | 1.04 (0.87 - 1.23) | 0.94 (0.70 - 1.27) | Reference | 1.73 (0.55 - 5.48) | 0.60 (0.49 - 0.74) | 0.83 (0.68 - 1.01) | 0.86 (0.64 - 1.15) | Reference |

| **Table S32 continued:** | | | | | | | | | | |
| --- | --- | --- | --- | --- | --- | --- | --- | --- | --- | --- |
|  | July and August 2023 (low COVID-19 disease burden) | | | | | February and March 2023 (high COVID-19 disease burden) | | | | |
|  | Four or more vaccine doses | Three vaccine doses | Two vaccine doses | One vaccine dose | Unvaccinated | Four or more vaccine doses | Three vaccine doses | Two vaccine doses | One vaccine dose | Unvaccinated |
|  | All-cause mortality | | | | | All-cause mortality | | | | |
| Deaths (n) | 2230 | 1870 | 510 | 101 | 627 | 1733 | 1598 | 428 | 80 | 553 |
| Events per 100,000 person days | 5.1 | 1.72 | 1.12 | 1.16 | 1.15 | 5.15 | 1.79 | 1.09 | 1.04 | 1.14 |
| Crude HR (95%CI) | 4.43 (4.05 - 4.84) | 1.49 (1.36 - 1.63) | 0.97 (0.87 - 1.10) | 1.01 (0.82 - 1.24) | Reference | 4.51 (4.10 - 4.97) | 1.57 (1.42 - 1.73) | 0.96 (0.84 - 1.09) | 0.91 (0.72 - 1.16) | Reference |
| Age adjusted HR (95% CI) | 0.96 (0.88 - 1.05) | 1.00 (0.91 - 1.09) | 1.18 (1.05 - 1.33) | 1.26 (1.03 - 1.56) | Reference | 0.90 (0.82 - 1.00) | 1.00 (0.91 - 1.11) | 1.13 (0.99 - 1.28) | 1.11 (0.88 - 1.40) | Reference |
| Age and gender adjusted HR (95% CI) | 0.94 (0.86 - 1.03) | 0.98 (0.89 - 1.07) | 1.17 (1.04 - 1.31) | 1.26 (1.02 - 1.55) | Reference | 0.88 (0.80 - 0.97) | 0.98 (0.89 - 1.08) | 1.11 (0.98 - 1.27) | 1.10 (0.87 - 1.39) | Reference |
| Age, gender and nursing home adjusted HR (95% CI) | 0.89 (0.81 - 0.97) | 0.96 (0.88 - 1.05) | 1.14 (1.01 - 1.28) | 1.23 (1.00 - 1.52) | Reference | 0.80 (0.73 - 0.89) | 0.96 (0.87 - 1.06) | 1.09 (0.96 - 1.23) | 1.09 (0.86 - 1.38) | Reference |
| Age, gender, nursing home and year of last infection adjusted HR (95% CI) | 0.87 (0.80 - 0.96) | 0.95 (0.87 - 1.04) | 1.17 (1.04 - 1.32) | 1.30 (1.05 - 1.61) | Reference | 0.80 (0.73 - 0.89) | 0.96 (0.87 - 1.06) | 1.10 (0.97 - 1.25) | 1.11 (0.87 - 1.41) | Reference |
|  | Non-COVID-19 mortality | | | | | Non-COVID-19 mortality | | | | |
| Deaths (n) | 2227 | 1863 | 510 | 100 | 623 | 1723 | 1594 | 425 | 80 | 553 |
| Events per 100,000 person days | 5.1 | 1.71 | 1.12 | 1.15 | 1.14 | 5.12 | 1.79 | 1.08 | 1.04 | 1.14 |
| Crude HR (95%CI) | 4.45 (4.07 - 4.87) | 1.50 (1.37 - 1.64) | 0.98 (0.87 - 1.10) | 1.01 (0.81 - 1.24) | Reference | 4.49 (4.08 - 4.94) | 1.57 (1.42 - 1.72) | 0.95 (0.84 - 1.08) | 0.91 (0.72 - 1.16) | Reference |
| Age adjusted HR (95% CI) | 0.97 (0.88 - 1.06) | 1.00 (0.91 - 1.09) | 1.19 (1.06 - 1.34) | 1.26 (1.02 - 1.56) | Reference | 0.90 (0.82 - 0.99) | 1.00 (0.91 - 1.10) | 1.12 (0.99 - 1.27) | 1.11 (0.88 - 1.40) | Reference |
| Age and gender adjusted HR (95% CI) | 0.94 (0.86 - 1.03) | 0.98 (0.89 - 1.07) | 1.18 (1.05 - 1.32) | 1.25 (1.02 - 1.55) | Reference | 0.87 (0.79 - 0.97) | 0.98 (0.89 - 1.08) | 1.11 (0.98 - 1.26) | 1.10 (0.87 - 1.39) | Reference |
| Age, gender and nursing home adjusted HR (95% CI) | 0.89 (0.81 - 0.98) | 0.96 (0.88 - 1.06) | 1.14 (1.02 - 1.29) | 1.23 (1.00 - 1.52) | Reference | 0.80 (0.72 - 0.89) | 0.96 (0.87 - 1.05) | 1.08 (0.95 - 1.23) | 1.09 (0.86 - 1.38) | Reference |
| Age, gender, nursing home and year of last infection adjusted HR (95% CI) | 0.88 (0.80 - 0.97) | 0.95 (0.87 - 1.04) | 1.18 (1.05 - 1.33) | 1.29 (1.04 - 1.60) | Reference | 0.80 (0.72 - 0.89) | 0.96 (0.87 - 1.06) | 1.09 (0.96 - 1.24) | 1.11 (0.87 - 1.41) | Reference |

| **Table S33:** Hazard ratios (HR) with 95% confidence intervals (95% CI) for non-COVID 19 and all-cause mortality according to number of SARS-CoV-2 vaccine doses during different time periods for individuals with their **last infection in 2020** only. | | | | | | | | | | |
| --- | --- | --- | --- | --- | --- | --- | --- | --- | --- | --- |
|  | 2021 | | | | | | | | | |
|  | June and July 2021 (low COVID-19 disease burden) | | | | | October and November 2021 (high COVID-19 disease burden) | | | | |
|  | Four or more vaccine doses | Three vaccine doses | Two vaccine doses | One vaccine dose | Unvaccinated | Four or more vaccine doses | Three vaccine doses | Two vaccine doses | One vaccine dose | Unvaccinated |
|  | All-cause mortality | | | | | All-cause mortality | | | | |
| Deaths (n) |  |  | 303 | 111 | 247 |  | 149 | 274 | 105 | 158 |
| Events per 100,000 person days |  |  | 6.22 | 1.82 | 2.95 |  | 16.22 | 3.26 | 2.09 | 3.21 |
| Crude HR (95%CI) |  |  | 2.18 (1.84 - 2.59) | 0.61 (0.49 - 0.76) | Reference |  | 5.70 (4.47 - 7.27) | 1.01 (0.83 - 1.23) | 0.65 (0.51 - 0.84) | Reference |
| Age adjusted HR (95% CI) |  |  | 0.67 (0.56 - 0.80) | 0.55 (0.44 - 0.69) | Reference |  | 0.75 (0.59 - 0.96) | 0.71 (0.58 - 0.86) | 0.64 (0.50 - 0.82) | Reference |
| Age and gender adjusted HR (95% CI) |  |  | 0.67 (0.56 - 0.79) | 0.53 (0.42 - 0.67) | Reference |  | 0.75 (0.59 - 0.95) | 0.70 (0.58 - 0.85) | 0.62 (0.48 - 0.80) | Reference |
| Age, gender and nursing home adjusted HR (95% CI) |  |  | 0.57 (0.48 - 0.68) | 0.56 (0.45 - 0.70) | Reference |  | 0.60 (0.47 - 0.77) | 0.67 (0.55 - 0.81) | 0.67 (0.52 - 0.86) | Reference |
|  | Non-COVID-19 mortality | | | | | Non-COVID-19 mortality | | | | |
| Deaths (n) |  |  | 294 | 109 | 242 |  | 148 | 269 | 104 | 153 |
| Events per 100,000 person days |  |  | 6.04 | 1.79 | 2.89 |  | 16.11 | 3.2 | 2.07 | 3.11 |
| Crude HR (95%CI) |  |  | 2.16 (1.82 - 2.57) | 0.61 (0.49 - 0.76) | Reference |  | 5.90 (4.62 - 7.54) | 1.02 (0.84 - 1.25) | 0.67 (0.52 - 0.86) | Reference |
| Age adjusted HR (95% CI) |  |  | 0.67 (0.56 - 0.79) | 0.55 (0.44 - 0.69) | Reference |  | 0.78 (0.61 - 0.99) | 0.72 (0.59 - 0.88) | 0.65 (0.51 - 0.84) | Reference |
| Age and gender adjusted HR (95% CI) |  |  | 0.66 (0.56 - 0.79) | 0.53 (0.42 - 0.67) | Reference |  | 0.77 (0.61 - 0.99) | 0.71 (0.58 - 0.87) | 0.64 (0.49 - 0.82) | Reference |
| Age, gender and nursing home adjusted HR (95% CI) |  |  | 0.57 (0.47 - 0.68) | 0.56 (0.45 - 0.70) | Reference |  | 0.61 (0.48 - 0.79) | 0.68 (0.56 - 0.83) | 0.69 (0.53 - 0.88) | Reference |
|  | May and June 2022 (low COVID-19 disease burden) | | | | | February and March 2022 (high COVID-19 disease burden) | | | | |
|  | Four or more vaccine doses | Three vaccine doses | Two vaccine doses | One vaccine dose | Unvaccinated | Four or more vaccine doses | Three vaccine doses | Two vaccine doses | One vaccine dose | Unvaccinated |
|  | All-cause mortality | | | | | All-cause mortality | | | | |
| Deaths (n) | 15 | 268 | 146 | 14 | 47 | 2 | 331 | 170 | 32 | 73 |
| Events per 100,000 person days | 29.66 | 4.06 | 3.01 | 1.25 | 3.47 | 29.76 | 4.5 | 2.9 | 1.86 | 3.45 |
| Crude HR (95%CI) | 9.37 (5.18 - 16.97) | 1.17 (0.86 - 1.59) | 0.87 (0.62 - 1.20) | 0.36 (0.20 - 0.65) | Reference | 8.37 (2.05 - 34.11) | 1.30 (1.01 - 1.68) | 0.84 (0.64 - 1.11) | 0.54 (0.36 - 0.82) | Reference |
| Age adjusted HR (95% CI) | 1.31 (0.71 - 2.40) | 0.81 (0.60 - 1.11) | 0.98 (0.70 - 1.36) | 0.57 (0.31 - 1.04) | Reference | 2.37 (0.58 - 9.67) | 0.76 (0.59 - 0.98) | 0.81 (0.62 - 1.07) | 0.75 (0.49 - 1.13) | Reference |
| Age and gender adjusted HR (95% CI) | 1.33 (0.72 - 2.44) | 0.81 (0.59 - 1.10) | 0.96 (0.69 - 1.34) | 0.58 (0.32 - 1.06) | Reference | ( - ) | 0.74 (0.58 - 0.96) | 0.79 (0.60 - 1.04) | 0.72 (0.48 - 1.10) | Reference |
| Age, gender and nursing home adjusted HR (95% CI) | 1.06 (0.56 - 2.00) | 0.73 (0.53 - 0.99) | 1.02 (0.73 - 1.42) | 0.65 (0.35 - 1.19) | Reference | 1.92 (0.47 - 7.84) | 0.65 (0.50 - 0.84) | 0.85 (0.65 - 1.13) | 0.89 (0.58 - 1.36) | Reference |
|  | Non-COVID-19 mortality | | | | | Non-COVID-19 mortality | | | | |
| Deaths (n) | 15 | 263 | 144 | 14 | 46 | 2 | 310 | 160 | 32 | 68 |
| Events per 100,000 person days | 29.66 | 3.98 | 2.97 | 1.25 | 3.40 | 29.76 | 4.22 | 2.73 | 1.86 | 3.21 |
| Crude HR (95%CI) | 9.72 (5.36 - 17.63) | 1.17 (0.86 - 1.60) | 0.87 (0.63 - 1.22) | 0.37 (0.20 - 0.67) | Reference | 8.99 (2.20 - 36.71) | 1.31 (1.01 - 1.70) | 0.85 (0.64 - 1.13) | 0.58 (0.38 - 0.89) | Reference |
| Age adjusted HR (95% CI) | 1.37 (0.74 - 2.53) | 0.81 (0.60 - 1.11) | 0.98 (0.70 - 1.37) | 0.58 (0.32 - 1.06) | Reference | 2.57 (0.63 - 10.52) | 0.77 (0.59 - 1.00) | 0.82 (0.61 - 1.09) | 0.79 (0.52 - 1.21) | Reference |
| Age and gender adjusted HR (95% CI) | 1.40 (0.76 - 2.58) | 0.81 (0.59 - 1.11) | 0.96 (0.69 - 1.35) | 0.59 (0.32 - 1.08) | Reference | ( - ) | 0.75 (0.57 - 0.97) | 0.80 (0.60 - 1.06) | 0.77 (0.51 - 1.18) | Reference |
| Age, gender and nursing home adjusted HR (95% CI) | 1.10 (0.58 - 2.08) | 0.73 (0.53 - 1.00) | 1.02 (0.73 - 1.43) | 0.66 (0.36 - 1.22) | Reference | 2.06 (0.50 - 8.44) | 0.66 (0.50 - 0.86) | 0.86 (0.65 - 1.14) | 0.96 (0.62 - 1.47) | Reference |

| **Table S33 continued:** | | | | | | | | | | |
| --- | --- | --- | --- | --- | --- | --- | --- | --- | --- | --- |
|  | July and August 2023 (low COVID-19 disease burden) | | | | | February and March 2023 (high COVID-19 disease burden) | | | | |
|  | Four or more vaccine doses | Three vaccine doses | Two vaccine doses | One vaccine dose | Unvaccinated | Four or more vaccine doses | Three vaccine doses | Two vaccine doses | One vaccine dose | Unvaccinated |
|  | All-cause mortality | | | | | All-cause mortality | | | | |
| Deaths (n) | 147 | 123 | 57 | 13 | 26 | 165 | 125 | 65 | 9 | 42 |
| Events per 100,000 person days | 8.74 | 2.96 | 1.82 | 1.58 | 2.27 | 9.7 | 3.01 | 2.1 | 1.11 | 3.78 |
| Crude HR (95%CI) | 3.85 (2.54 - 5.85) | 1.31 (0.86 - 2.00) | 0.80 (0.51 - 1.28) | 0.70 (0.36 - 1.35) | Reference | 2.56 (1.83 - 3.59) | 0.80 (0.56 - 1.13) | 0.56 (0.38 - 0.82) | 0.29 (0.14 - 0.60) | Reference |
| Age adjusted HR (95% CI) | 1.26 (0.83 - 1.92) | 1.28 (0.83 - 1.95) | 1.09 (0.68 - 1.74) | 1.01 (0.52 - 1.97) | Reference | 0.82 (0.58 - 1.15) | 0.81 (0.57 - 1.15) | 0.83 (0.56 - 1.23) | 0.47 (0.23 - 0.97) | Reference |
| Age and gender adjusted HR (95% CI) | 1.29 (0.84 - 1.96) | 1.25 (0.82 - 1.92) | 1.06 (0.67 - 1.70) | 0.97 (0.50 - 1.90) | Reference | 0.81 (0.57 - 1.13) | 0.79 (0.56 - 1.13) | 0.79 (0.53 - 1.17) | 0.45 (0.22 - 0.94) | Reference |
| Age, gender and nursing home adjusted HR (95% CI) | 1.14 (0.74 - 1.74) | 1.27 (0.83 - 1.95) | 1.17 (0.73 - 1.89) | 1.04 (0.53 - 2.06) | Reference | 0.67 (0.47 - 0.95) | 0.79 (0.56 - 1.13) | 0.89 (0.60 - 1.32) | 0.51 (0.24 - 1.06) | Reference |
|  | Non-COVID-19 mortality | | | | | Non-COVID-19 mortality | | | | |
| Deaths (n) | 146 | 122 | 57 | 13 | 26 | 156 | 123 | 63 | 9 | 40 |
| Events per 100,000 person days | 8.68 | 2.94 | 1.82 | 1.58 | 2.27 | 9.17 | 2.97 | 2.04 | 1.11 | 3.60 |
| Crude HR (95%CI) | 3.83 (2.52 - 5.81) | 1.30 (0.85 - 1.98) | 0.80 (0.51 - 1.28) | 0.70 (0.36 - 1.35) | Reference | 2.54 (1.80 - 3.60) | 0.82 (0.58 - 1.18) | 0.57 (0.38 - 0.84) | 0.31 (0.15 - 0.63) | Reference |
| Age adjusted HR (95% CI) | 1.26 (0.83 - 1.91) | 1.26 (0.83 - 1.94) | 1.09 (0.68 - 1.74) | 1.01 (0.52 - 1.97) | Reference | 0.81 (0.57 - 1.15) | 0.83 (0.58 - 1.20) | 0.83 (0.56 - 1.25) | 0.49 (0.24 - 1.02) | Reference |
| Age and gender adjusted HR (95% CI) | 1.28 (0.84 - 1.94) | 1.24 (0.81 - 1.90) | 1.06 (0.67 - 1.70) | 0.97 (0.50 - 1.90) | Reference | 0.80 (0.56 - 1.13) | 0.82 (0.57 - 1.17) | 0.80 (0.53 - 1.20) | 0.48 (0.23 - 0.98) | Reference |
| Age, gender and nursing home adjusted HR (95% CI) | 1.13 (0.74 - 1.72) | 1.26 (0.82 - 1.93) | 1.17 (0.73 - 1.89) | 1.04 (0.53 - 2.06) | Reference | 0.80 (0.56 - 1.13) | 0.82 (0.57 - 1.17) | 0.89 (0.59 - 1.34) | 0.53 (0.25 - 1.10) | Reference |

| **Table S34:** Hazard ratios (HR) with 95% confidence intervals (95% CI) for non-COVID 19 and all-cause mortality according to number of SARS-CoV-2 vaccine doses during different time periods for individuals with their **last infection in 2021** only. | | | | | | | | | | |
| --- | --- | --- | --- | --- | --- | --- | --- | --- | --- | --- |
|  | 2021 | | | | | | | | | |
|  | June and July 2021 (low COVID-19 disease burden) | | | | | October and November 2021 (high COVID-19 disease burden) | | | | |
|  | Four or more vaccine doses | Three vaccine doses | Two vaccine doses | One vaccine dose | Unvaccinated | Four or more vaccine doses | Three vaccine doses | Two vaccine doses | One vaccine dose | Unvaccinated |
|  | All-cause mortality | | | | | All-cause mortality | | | | |
| Deaths (n) |  |  | 76 | 30 | 129 |  | 42 | 126 | 92 | 131 |
| Events per 100,000 person days |  |  | 9.49 | 2.55 | 4.07 |  | 16.36 | 3.15 | 1.91 | 2.60 |
| Crude HR (95%CI) |  |  | 2.47 (1.85 - 3.30) | 0.63 (0.42 - 0.94) | Reference |  | 6.62 (4.56 - 9.61) | 1.21 (0.94 - 1.54) | 0.75 (0.57 - 0.98) | Reference |
| Age adjusted HR (95% CI) |  |  | 0.73 (0.55 - 0.98) | 0.48 (0.32 - 0.71) | Reference |  | 0.71 (0.49 - 1.03) | 0.63 (0.49 - 0.80) | 0.63 (0.48 - 0.82) | Reference |
| Age and gender adjusted HR (95% CI) |  |  | 0.74 (0.55 - 0.99) | 0.47 (0.32 - 0.70) | Reference |  | 0.72 (0.49 - 1.04) | 0.62 (0.49 - 0.79) | 0.61 (0.47 - 0.80) | Reference |
| Age, gender and nursing home adjusted HR (95% CI) |  |  | 0.54 (0.40 - 0.74) | 0.42 (0.28 - 0.63) | Reference |  | 0.53 (0.35 - 0.79) | 0.55 (0.43 - 0.71) | 0.60 (0.46 - 0.79) | Reference |
|  | Non-COVID-19 mortality | | | | | Non-COVID-19 mortality | | | | |
| Deaths (n) |  |  | 75 | 30 | 125 |  | 41 | 121 | 89 | 123 |
| Events per 100,000 person days |  |  | 9.37 | 2.55 | 3.94 |  | 15.97 | 3.03 | 1.85 | 2.44 |
| Crude HR (95%CI) |  |  | 2.52 (1.88 - 3.37) | 0.65 (0.44 - 0.97) | Reference |  | 7.08 (4.84 - 10.36) | 1.24 (0.96 - 1.60) | 0.78 (0.59 - 1.03) | Reference |
| Age adjusted HR (95% CI) |  |  | 0.74 (0.55 - 0.99) | 0.49 (0.33 - 0.73) | Reference |  | 0.74 (0.51 - 1.09) | 0.64 (0.50 - 0.82) | 0.65 (0.49 - 0.85) | Reference |
| Age and gender adjusted HR (95% CI) |  |  | 0.75 (0.56 - 1.00) | 0.49 (0.33 - 0.73) | Reference |  | 0.75 (0.52 - 1.10) | 0.64 (0.49 - 0.82) | 0.64 (0.49 - 0.84) | Reference |
| Age, gender and nursing home adjusted HR (95% CI) |  |  | 0.55 (0.40 - 0.74) | 0.43 (0.29 - 0.65) | Reference |  | 0.53 (0.35 - 0.80) | 0.56 (0.43 - 0.72) | 0.62 (0.48 - 0.82) | Reference |
|  | May and June 2022 (low COVID-19 disease burden) | | | | | February and March 2022 (high COVID-19 disease burden) | | | | |
|  | Four or more vaccine doses | Three vaccine doses | Two vaccine doses | One vaccine dose | Unvaccinated | Four or more vaccine doses | Three vaccine doses | Two vaccine doses | One vaccine dose | Unvaccinated |
|  | All-cause mortality | | | | | All-cause mortality | | | | |
| Deaths (n) | 2 | 240 | 200 | 42 | 132 | 3 | 141 | 186 | 44 | 92 |
| Events per 100,000 person days | 4.58 | 2.65 | 1.97 | 1.19 | 1.09 | 95.24 | 3.19 | 2.59 | 1.23 | 1.57 |
| Crude HR (95%CI) | 3.88 (0.96 - 15.73) | 2.43 (1.96 - 3.00) | 1.82 (1.46 - 2.26) | 1.09 (0.77 - 1.55) | Reference | 60.07 (18.99 - 189.94) | 2.02 (1.56 - 2.63) | 1.65 (1.29 - 2.12) | 0.79 (0.55 - 1.12) | Reference |
| Age adjusted HR (95% CI) | 0.25 (0.06 - 1.01) | 0.76 (0.61 - 0.94) | 1.03 (0.83 - 1.29) | 1.20 (0.84 - 1.69) | Reference | 5.33 (1.68 - 16.94) | 0.61 (0.47 - 0.80) | 0.81 (0.63 - 1.05) | 0.84 (0.59 - 1.20) | Reference |
| Age and gender adjusted HR (95% CI) | 0.25 (0.06 - 1.00) | 0.75 (0.60 - 0.93) | 1.01 (0.81 - 1.27) | 1.19 (0.84 - 1.68) | Reference | ( - ) | 0.61 (0.47 - 0.80) | 0.80 (0.63 - 1.03) | 0.83 (0.58 - 1.19) | Reference |
| Age, gender and nursing home adjusted HR (95% CI) | 0.17 (0.04 - 0.71) | 0.67 (0.54 - 0.83) | 0.98 (0.79 - 1.22) | 1.16 (0.82 - 1.64) | Reference | 4.13 (1.25 - 13.68) | 0.52 (0.39 - 0.68) | 0.76 (0.59 - 0.98) | 0.84 (0.58 - 1.20) | Reference |
|  | Non-COVID-19 mortality | | | | | Non-COVID-19 mortality | | | | |
| Deaths (n) | 2 | 236 | 197 | 42 | 128 | 1 | 135 | 179 | 42 | 82 |
| Events per 100,000 person days | 4.58 | 2.6 | 1.94 | 1.19 | 1.05 | 31.75 | 3.05 | 2.49 | 1.18 | 1.40 |
| Crude HR (95%CI) | 3.98 (0.98 - 16.14) | 2.46 (1.99 - 3.05) | 1.84 (1.48 - 2.30) | 1.13 (0.79 - 1.60) | Reference | 22.75 (3.16 - 163.62) | 2.18 (1.66 - 2.87) | 1.78 (1.37 - 2.31) | 0.84 (0.58 - 1.22) | Reference |
| Age adjusted HR (95% CI) | 0.26 (0.06 - 1.05) | 0.77 (0.62 - 0.96) | 1.05 (0.84 - 1.31) | 1.23 (0.87 - 1.75) | Reference | 1.97 (0.27 - 14.19) | 0.66 (0.50 - 0.88) | 0.88 (0.68 - 1.14) | 0.90 (0.62 - 1.30) | Reference |
| Age and gender adjusted HR (95% CI) | 0.26 (0.06 - 1.04) | 0.76 (0.61 - 0.95) | 1.03 (0.83 - 1.29) | 1.23 (0.87 - 1.74) | Reference | ( - ) | 0.66 (0.50 - 0.87) | 0.87 (0.67 - 1.13) | 0.89 (0.61 - 1.29) | Reference |
| Age, gender and nursing home adjusted HR (95% CI) | 0.17 (0.04 - 0.72) | 0.68 (0.54 - 0.85) | 1.00 (0.80 - 1.25) | 1.19 (0.84 - 1.69) | Reference | 1.55 (0.21 - 11.48) | 0.56 (0.42 - 0.75) | 0.82 (0.63 - 1.07) | 0.90 (0.62 - 1.30) | Reference |

| **Table S34 continued:** | | | | | | | | | | |
| --- | --- | --- | --- | --- | --- | --- | --- | --- | --- | --- |
|  | July and August 2023 (low COVID-19 disease burden) | | | | | February and March 2023 (high COVID-19 disease burden) | | | | |
|  | Four or more vaccine doses | Three vaccine doses | Two vaccine doses | One vaccine dose | Unvaccinated | Four or more vaccine doses | Three vaccine doses | Two vaccine doses | One vaccine dose | Unvaccinated |
|  | All-cause mortality | | | | | All-cause mortality | | | | |
| Deaths (n) | 149 | 133 | 78 | 25 | 109 | 164 | 175 | 86 | 21 | 105 |
| Events per 100,000 person days | 6.42 | 1.97 | 1.14 | 0.91 | 1.04 | 7.11 | 2.59 | 1.28 | 0.79 | 1.05 |
| Crude HR (95%CI) | 6.17 (4.82 - 7.89) | 1.89 (1.47 - 2.44) | 1.10 (0.82 - 1.47) | 0.87 (0.57 - 1.35) | Reference | 6.78 (5.30 - 8.66) | 2.48 (1.95 - 3.16) | 1.22 (0.92 - 1.63) | 0.75 (0.47 - 1.20) | Reference |
| Age adjusted HR (95% CI) | 1.03 (0.79 - 1.33) | 0.88 (0.68 - 1.14) | 0.83 (0.62 - 1.12) | 1.02 (0.66 - 1.57) | Reference | 1.12 (0.87 - 1.45) | 1.13 (0.88 - 1.44) | 0.91 (0.69 - 1.22) | 0.89 (0.56 - 1.42) | Reference |
| Age and gender adjusted HR (95% CI) | 1.00 (0.77 - 1.30) | 0.86 (0.67 - 1.12) | 0.82 (0.62 - 1.10) | 1.01 (0.65 - 1.56) | Reference | 1.09 (0.85 - 1.42) | 1.10 (0.86 - 1.41) | 0.91 (0.68 - 1.20) | 0.88 (0.55 - 1.41) | Reference |
| Age, gender and nursing home adjusted HR (95% CI) | 0.90 (0.69 - 1.18) | 0.83 (0.64 - 1.07) | 0.82 (0.61 - 1.09) | 1.00 (0.65 - 1.54) | Reference | 0.98 (0.75 - 1.28) | 1.05 (0.82 - 1.34) | 0.90 (0.68 - 1.20) | 0.87 (0.54 - 1.38) | Reference |
|  | Non-COVID-19 mortality | | | | | Non-COVID-19 mortality | | | | |
| Deaths (n) | 149 | 132 | 78 | 25 | 109 | 158 | 173 | 85 | 21 | 102 |
| Events per 100,000 person days | 6.42 | 1.96 | 1.14 | 0.91 | 1.04 | 6.85 | 2.57 | 1.27 | 0.79 | 1.02 |
| Crude HR (95%CI) | 6.17 (4.82 - 7.89) | 1.88 (1.46 - 2.42) | 1.10 (0.82 - 1.47) | 0.87 (0.57 - 1.35) | Reference | 6.72 (5.24 - 8.62) | 2.52 (1.97 - 3.22) | 1.25 (0.93 - 1.66) | 0.78 (0.49 - 1.24) | Reference |
| Age adjusted HR (95% CI) | 1.03 (0.79 - 1.33) | 0.87 (0.68 - 1.13) | 0.83 (0.62 - 1.12) | 1.02 (0.66 - 1.57) | Reference | 1.13 (0.87 - 1.46) | 1.15 (0.90 - 1.47) | 0.93 (0.70 - 1.24) | 0.91 (0.57 - 1.46) | Reference |
| Age and gender adjusted HR (95% CI) | 1.00 (0.77 - 1.30) | 0.86 (0.66 - 1.11) | 0.82 (0.62 - 1.10) | 1.01 (0.65 - 1.56) | Reference | 1.10 (0.84 - 1.43) | 1.13 (0.88 - 1.44) | 0.92 (0.69 - 1.23) | 0.91 (0.57 - 1.45) | Reference |
| Age, gender and nursing home adjusted HR (95% CI) | 0.90 (0.69 - 1.18) | 0.82 (0.64 - 1.07) | 0.82 (0.61 - 1.09) | 1.00 (0.65 - 1.54) | Reference | 1.10 (0.84 - 1.43) | 1.07 (0.84 - 1.37) | 0.92 (0.69 - 1.22) | 0.89 (0.56 - 1.42) | Reference |

| **Table S35:** Hazard ratios (HR) with 95% confidence intervals (95% CI) for non-COVID 19 and all-cause mortality according to number of SARS-CoV-2 vaccine doses during different time periods for individuals with their **last infection in 2022** only. | | | | | | | | | | |
| --- | --- | --- | --- | --- | --- | --- | --- | --- | --- | --- |
|  | 2021 | | | | | | | | | |
|  | June and July 2021 (low COVID-19 disease burden) | | | | | October and November 2021 (high COVID-19 disease burden) | | | | |
|  | Four or more vaccine doses | Three vaccine doses | Two vaccine doses | One vaccine dose | Unvaccinated | Four or more vaccine doses | Three vaccine doses | Two vaccine doses | One vaccine dose | Unvaccinated |
|  | All-cause mortality | | | | | All-cause mortality | | | | |
| Deaths (n) |  |  |  |  | 0 |  |  |  |  | 0 |
| Events per 100,000 person days |  |  |  |  |  |  |  |  |  |  |
| Crude HR (95%CI) |  |  |  |  | Reference |  |  |  |  | Reference |
| Age adjusted HR (95% CI) |  |  |  |  | Reference |  |  |  |  | Reference |
| Age and gender adjusted HR (95% CI) |  |  |  |  | Reference |  |  |  |  | Reference |
| Age, gender and nursing home adjusted HR (95% CI) |  |  |  |  | Reference |  |  |  |  | Reference |
|  | Non-COVID-19 mortality | | | | | Non-COVID-19 mortality | | | | |
| Deaths (n) |  |  |  |  | 0 |  |  |  |  | 0 |
| Events per 100,000 person days |  |  |  |  |  |  |  |  |  |  |
| Crude HR (95%CI) |  |  |  |  | Reference |  |  |  |  | Reference |
| Age adjusted HR (95% CI) |  |  |  |  | Reference |  |  |  |  | Reference |
| Age and gender adjusted HR (95% CI) |  |  |  |  | Reference |  |  |  |  | Reference |
| Age, gender and nursing home adjusted HR (95% CI) |  |  |  |  | Reference |  |  |  |  | Reference |
|  | May and June 2022 (low COVID-19 disease burden) | | | | | February and March 2022 (high COVID-19 disease burden) | | | | |
|  | Four or more vaccine doses | Three vaccine doses | Two vaccine doses | One vaccine dose | Unvaccinated | Four or more vaccine doses | Three vaccine doses | Two vaccine doses | One vaccine dose | Unvaccinated |
|  | All-cause mortality | | | | | All-cause mortality | | | | |
| Deaths (n) | 5 | 151 | 41 | 6 | 51 |  |  |  |  | 0 |
| Events per 100,000 person days | 22.82 | 1.83 | 0.57 | 0.59 | 0.52 |  |  |  |  |  |
| Crude HR (95%CI) | 39.79 (15.67 - 101.04) | 3.54 (2.58 - 4.86) | 1.09 (0.73 - 1.65) | 1.14 (0.49 - 2.64) | Reference |  |  |  |  | Reference |
| Age adjusted HR (95% CI) | 1.96 (0.74 - 5.15) | 1.26 (0.91 - 1.75) | 1.42 (0.94 - 2.15) | 1.23 (0.53 - 2.88) | Reference |  |  |  |  | Reference |
| Age and gender adjusted HR (95% CI) | 1.86 (0.70 - 4.90) | 1.21 (0.87 - 1.68) | 1.40 (0.93 - 2.12) | 1.24 (0.53 - 2.88) | Reference |  |  |  |  | Reference |
| Age, gender and nursing home adjusted HR (95% CI) | 1.18 (0.43 - 3.26) | 1.00 (0.71 - 1.41) | 1.19 (0.78 - 1.82) | 0.92 (0.38 - 2.18) | Reference |  |  |  |  | Reference |
|  | Non-COVID-19 mortality | | | | | Non-COVID-19 mortality | | | | |
| Deaths (n) | 5 | 150 | 41 | 5 | 51 |  |  |  |  | 0 |
| Events per 100,000 person days | 22.82 | 1.82 | 0.57 | 0.49 | 0.52 |  |  |  |  |  |
| Crude HR (95%CI) | 39.79 (15.67 - 101.04) | 3.52 (2.56 - 4.83) | 1.09 (0.73 - 1.65) | 0.95 (0.38 - 2.37) | Reference |  |  |  |  | Reference |
| Age adjusted HR (95% CI) | 1.96 (0.74 - 5.15) | 1.25 (0.90 - 1.74) | 1.42 (0.94 - 2.15) | 1.03 (0.41 - 2.59) | Reference |  |  |  |  | Reference |
| Age and gender adjusted HR (95% CI) | 1.86 (0.70 - 4.90) | 1.20 (0.87 - 1.67) | 1.40 (0.93 - 2.12) | 1.03 (0.41 - 2.59) | Reference |  |  |  |  | Reference |
| Age, gender and nursing home adjusted HR (95% CI) | 1.18 (0.43 - 3.26) | 0.99 (0.71 - 1.40) | 1.19 (0.78 - 1.82) | 0.78 (0.30 - 1.99) | Reference |  |  |  |  | Reference |

| **Table S35 continued:** | | | | | | | | | | |
| --- | --- | --- | --- | --- | --- | --- | --- | --- | --- | --- |
|  | July and August 2023 (low COVID-19 disease burden) | | | | | February and March 2023 (high COVID-19 disease burden) | | | | |
|  | Four or more vaccine doses | Three vaccine doses | Two vaccine doses | One vaccine dose | Unvaccinated | Four or more vaccine doses | Three vaccine doses | Two vaccine doses | One vaccine dose | Unvaccinated |
|  | All-cause mortality | | | | | All-cause mortality | | | | |
| Deaths (n) | 1542 | 1338 | 320 | 51 | 434 | 1531 | 1392 | 301 | 52 | 439 |
| Events per 100,000 person days | 4.46 | 1.5 | 0.97 | 1.07 | 1.06 | 4.96 | 1.71 | 0.99 | 1.18 | 1.15 |
| Crude HR (95%CI) | 4.22 (3.80 - 4.70) | 1.42 (1.27 - 1.58) | 0.91 (0.79 - 1.06) | 1.01 (0.76 - 1.35) | Reference | 4.29 (3.86 - 4.77) | 1.48 (1.33 - 1.64) | 0.85 (0.74 - 0.99) | 1.03 (0.77 - 1.37) | Reference |
| Age adjusted HR (95% CI) | 0.91 (0.82 - 1.02) | 0.96 (0.86 - 1.07) | 1.32 (1.15 - 1.53) | 1.43 (1.07 - 1.91) | Reference | 0.87 (0.78 - 0.97) | 0.99 (0.89 - 1.10) | 1.25 (1.08 - 1.45) | 1.43 (1.07 - 1.91) | Reference |
| Age and gender adjusted HR (95% CI) | 0.89 (0.79 - 0.99) | 0.93 (0.84 - 1.04) | 1.32 (1.14 - 1.52) | 1.43 (1.07 - 1.91) | Reference | 0.85 (0.76 - 0.94) | 0.96 (0.87 - 1.07) | 1.24 (1.07 - 1.44) | 1.43 (1.07 - 1.91) | Reference |
| Age, gender and nursing home adjusted HR (95% CI) | 0.85 (0.76 - 0.95) | 0.93 (0.83 - 1.03) | 1.23 (1.07 - 1.43) | 1.31 (0.98 - 1.75) | Reference | 0.79 (0.70 - 0.88) | 0.95 (0.85 - 1.06) | 1.17 (1.01 - 1.35) | 1.33 (0.99 - 1.77) | Reference |
|  | Non-COVID-19 mortality | | | | | Non-COVID-19 mortality | | | | |
| Deaths (n) | 1541 | 1334 | 320 | 50 | 431 | 1490 | 1365 | 296 | 52 | 428 |
| Events per 100,000 person days | 4.46 | 1.49 | 0.97 | 1.05 | 1.05 | 4.82 | 1.67 | 0.97 | 1.18 | 1.13 |
| Crude HR (95%CI) | 4.25 (3.82 - 4.73) | 1.43 (1.28 - 1.59) | 0.92 (0.80 - 1.06) | 1.00 (0.75 - 1.34) | Reference | 4.28 (3.85 - 4.77) | 1.49 (1.33 - 1.66) | 0.86 (0.74 - 1.00) | 1.05 (0.79 - 1.40) | Reference |
| Age adjusted HR (95% CI) | 0.92 (0.82 - 1.02) | 0.96 (0.86 - 1.07) | 1.33 (1.15 - 1.54) | 1.41 (1.05 - 1.89) | Reference | 0.88 (0.79 - 0.98) | 0.99 (0.89 - 1.11) | 1.26 (1.09 - 1.46) | 1.47 (1.10 - 1.96) | Reference |
| Age and gender adjusted HR (95% CI) | 0.89 (0.80 - 1.00) | 0.94 (0.84 - 1.05) | 1.32 (1.15 - 1.53) | 1.41 (1.05 - 1.89) | Reference | 0.85 (0.76 - 0.95) | 0.97 (0.87 - 1.08) | 1.25 (1.08 - 1.45) | 1.47 (1.10 - 1.96) | Reference |
| Age, gender and nursing home adjusted HR (95% CI) | 0.86 (0.76 - 0.96) | 0.93 (0.84 - 1.04) | 1.24 (1.07 - 1.44) | 1.29 (0.96 - 1.73) | Reference | 0.85 (0.76 - 0.95) | 0.96 (0.86 - 1.07) | 1.18 (1.02 - 1.37) | 1.36 (1.02 - 1.82) | Reference |

| **Table S36:** Hazard ratios (HR) with 95% confidence intervals (95% CI) for non-COVID 19 and all-cause mortality according to number of SARS-CoV-2 vaccine doses during different time periods for **mRNA vaccinated** only. | | | | | | | | | | |
| --- | --- | --- | --- | --- | --- | --- | --- | --- | --- | --- |
|  | 2021 | | | | | | | | | |
|  | June and July 2021 (low COVID-19 disease burden) | | | | | October and November 2021 (high COVID-19 disease burden) | | | | |
|  | Four or more vaccine doses | Three vaccine doses | Two vaccine doses | One vaccine dose | Unvaccinated | Four or more vaccine doses | Three vaccine doses | Two vaccine doses | One vaccine dose | Unvaccinated |
|  | All-cause mortality | | | | | All-cause mortality | | | | |
| Deaths (n) |  |  | 378 | 121 | (-) |  | 191 | 387 | 163 | (-) |
| Events per 100,000 person days |  |  | 7.88 | 2.04 | (-) |  | 16.27 | 3.5 | 2.11 | (-) |
| Crude HR (95%CI) |  |  | 4.11 (3.35 - 5.06) | Ref | (-) |  | 9.23 (7.42 - 11.49) | 1.65 (1.38 - 1.99) | Ref | (-) |
| Age adjusted HR (95% CI) |  |  | 1.26 (1.02 - 1.56) | Ref | (-) |  | 1.11 (0.88 - 1.41) | 1.08 (0.90 - 1.30) | Ref | (-) |
| Age and gender adjusted HR (95% CI) |  |  | 1.27 (1.02 - 1.57) | Ref | (-) |  | 1.14 (0.90 - 1.44) | 1.08 (0.90 - 1.29) | Ref | (-) |
| Age, gender and nursing home adjusted HR (95% CI) |  |  | 1.02 (0.82 - 1.27) | Ref | (-) |  | 0.67 (0.52 - 0.86) | 0.93 (0.77 - 1.11) | Ref | (-) |
| Age, gender, nursing home and year of last infection adjusted HR (95% CI) |  |  | 1.03 (0.83 - 1.29) | Ref | (-) |  | 0.69 (0.54 - 0.89) | 0.94 (0.78 - 1.13) | Ref | (-) |
|  | Non-COVID-19 mortality | | | | | Non-COVID-19 mortality | | | | |
| Deaths (n) |  |  | 368 | 120 | (-) |  | 189 | 377 | 160 | (-) |
| Events per 100,000 person days |  |  | 7.67 | 2.02 | (-) |  | 16.1 | 3.41 | 2.07 | (-) |
| Crude HR (95%CI) |  |  | 4.03 (3.28 - 4.96) | Ref | (-) |  | 9.36 (7.51 - 11.67) | 1.64 (1.36 - 1.97) | Ref | (-) |
| Age adjusted HR (95% CI) |  |  | 1.24 (1.00 - 1.53) | Ref | (-) |  | 1.13 (0.89 - 1.43) | 1.07 (0.89 - 1.29) | Ref | (-) |
| Age and gender adjusted HR (95% CI) |  |  | 1.25 (1.00 - 1.54) | Ref | (-) |  | 1.15 (0.91 - 1.46) | 1.07 (0.89 - 1.29) | Ref | (-) |
| Age, gender and nursing home adjusted HR (95% CI) |  |  | 1.00 (0.80 - 1.25) | Ref | (-) |  | 0.67 (0.52 - 0.86) | 0.92 (0.76 - 1.10) | Ref | (-) |
| Age, gender, nursing home and year of last infection adjusted HR (95% CI) | | | 1.02 (0.81 - 1.27) | Ref | (-) |  | 0.68 (0.53 - 0.88) | 0.93 (0.77 - 1.12) | Ref | (-) |
|  | May and June 2022 (low COVID-19 disease burden) | | | | | February and March 2022 (high COVID-19 disease burden) | | | | |
|  | Four or more vaccine doses | Three vaccine doses | Two vaccine doses | One vaccine dose | Unvaccinated | Four or more vaccine doses | Three vaccine doses | Two vaccine doses | One vaccine dose | Unvaccinated |
|  | All-cause mortality | | | | | All-cause mortality | | | | |
| Deaths (n) | 22 | 657 | 373 | 53 | (-) | 5 | 472 | 350 | 74 | (-) |
| Events per 100,000 person days | 19.09 | 2.75 | 1.76 | 1.09 | (-) | 51.19 | 4.01 | 2.78 | 1.48 | (-) |
| Crude HR (95%CI) | 18.32 (10.99 - 30.55) | 2.54 (1.92 - 3.36) | 1.62 (1.22 - 2.16) | Ref | (-) | 35.99 (14.51 - 89.23) | 2.69 (2.11 - 3.44) | 1.87 (1.46 - 2.40) | Ref | (-) |
| Age adjusted HR (95% CI) | 1.43 (0.82 - 2.49) | 0.95 (0.72 - 1.26) | 1.17 (0.88 - 1.56) | Ref | (-) | 4.60 (1.84 - 11.49) | 0.86 (0.67 - 1.10) | 0.98 (0.76 - 1.26) | Ref | (-) |
| Age and gender adjusted HR (95% CI) | 1.41 (0.81 - 2.46) | 0.94 (0.71 - 1.24) | 1.15 (0.86 - 1.54) | Ref | (-) | 4.63 (1.86 - 11.56) | 0.85 (0.66 - 1.09) | 0.97 (0.76 - 1.25) | Ref | (-) |
| Age, gender and nursing home adjusted HR (95% CI) | 0.94 (0.51 - 1.72) | 0.82 (0.62 - 1.09) | 1.13 (0.84 - 1.50) | Ref | (-) | 2.79 (1.09 - 7.14) | 0.66 (0.51 - 0.86) | 0.91 (0.71 - 1.17) | Ref | (-) |
| Age, gender, nursing home and year of last infection adjusted HR (95% CI) | 0.96 (0.52 - 1.75) | 0.82 (0.62 - 1.09) | 1.13 (0.84 - 1.50) | Ref | (-) | 2.80 (1.09 - 7.18) | 0.66 (0.51 - 0.86) | 0.91 (0.71 - 1.18) | Ref | (-) |
|  | Non-COVID-19 mortality | | | | | Non-COVID-19 mortality | | | | |
| Deaths (n) | 22 | 647 | 368 | 52 | (-) | 3 | 445 | 333 | 72 | (-) |
| Events per 100,000 person days | 19.09 | 2.71 | 1.74 | 1.07 | (-) | 30.71 | 3.78 | 2.64 | 1.44 | (-) |
| Crude HR (95%CI) | 18.74 (11.22 - 31.30) | 2.55 (1.92 - 3.38) | 1.63 (1.22 - 2.18) | Ref | (-) | 22.48 (7.07 - 71.48) | 2.62 (2.04 - 3.35) | 1.83 (1.42 - 2.36) | Ref | (-) |
| Age adjusted HR (95% CI) | 1.48 (0.84 - 2.59) | 0.96 (0.72 - 1.27) | 1.17 (0.88 - 1.57) | Ref | (-) | 2.86 (0.89 - 9.15) | 0.84 (0.65 - 1.08) | 0.96 (0.74 - 1.24) | Ref | (-) |
| Age and gender adjusted HR (95% CI) | 1.46 (0.84 - 2.56) | 0.94 (0.71 - 1.25) | 1.16 (0.87 - 1.55) | Ref | (-) | 2.89 (0.90 - 9.23) | 0.83 (0.65 - 1.07) | 0.95 (0.74 - 1.23) | Ref | (-) |
| Age, gender and nursing home adjusted HR (95% CI) | 1.00 (0.54 - 1.84) | 0.83 (0.62 - 1.10) | 1.13 (0.85 - 1.52) | Ref | (-) | 1.79 (0.55 - 5.86) | 0.66 (0.51 - 0.85) | 0.89 (0.69 - 1.15) | Ref | (-) |
| Age, gender, nursing home and year of last infection adjusted HR (95% CI) | 1.01 (0.55 - 1.86) | 0.82 (0.62 - 1.10) | 1.13 (0.85 - 1.52) | Ref | (-) | 1.79 (0.55 - 5.87) | 0.66 (0.50 - 0.85) | 0.90 (0.69 - 1.16) | Ref | (-) |

| **Table S36 continued:** | | | | | | | | | | |
| --- | --- | --- | --- | --- | --- | --- | --- | --- | --- | --- |
|  | July and August 2023 (low COVID-19 disease burden) | | | | | February and March 2023 (high COVID-19 disease burden) | | | | |
|  | Four or more vaccine doses | Three vaccine doses | Two vaccine doses | One vaccine dose | Unvaccinated | Four or more vaccine doses | Three vaccine doses | Two vaccine doses | One vaccine dose | Unvaccinated |
|  | All-cause mortality | | | | | All-cause mortality | | | | |
| Deaths (n) | 2223 | 1869 | 493 | 87 | (-) | 1856 | 1688 | 436 | 71 | (-) |
| Events per 100,000 person days | 5.11 | 1.72 | 1.12 | 1.17 | (-) | 5.34 | 1.83 | 1.12 | 1.05 | (-) |
| Crude HR (95%CI) | 4.38 (3.53 - 5.43) | 1.47 (1.19 - 1.83) | 0.96 (0.77 - 1.21) | Ref | (-) | 5.07 (4.00 - 6.42) | 1.74 (1.37 - 2.20) | 1.06 (0.83 - 1.36) | Ref | (-) |
| Age adjusted HR (95% CI) | 0.76 (0.61 - 0.94) | 0.80 (0.65 - 1.00) | 0.96 (0.76 - 1.21) | Ref | (-) | 0.85 (0.67 - 1.07) | 0.95 (0.75 - 1.21) | 1.07 (0.83 - 1.38) | Ref | (-) |
| Age and gender adjusted HR (95% CI) | 0.74 (0.60 - 0.92) | 0.79 (0.64 - 0.98) | 0.96 (0.76 - 1.20) | Ref | (-) | 0.83 (0.65 - 1.05) | 0.94 (0.74 - 1.19) | 1.07 (0.83 - 1.37) | Ref | (-) |
| Age, gender and nursing home adjusted HR (95% CI) | 0.75 (0.60 - 0.93) | 0.80 (0.64 - 0.99) | 0.95 (0.75 - 1.19) | Ref | (-) | 0.80 (0.63 - 1.02) | 0.93 (0.74 - 1.18) | 1.05 (0.82 - 1.35) | Ref | (-) |
| Age, gender, nursing home and year of last infection adjusted HR (95% CI) | 0.71 (0.57 - 0.89) | 0.74 (0.59 - 0.91) | 0.90 (0.72 - 1.14) | Ref | (-) | 0.80 (0.62 - 1.01) | 0.92 (0.72 - 1.17) | 1.02 (0.80 - 1.32) | Ref | (-) |
|  | Non-COVID-19 mortality | | | | | Non-COVID-19 mortality | | | | |
| Deaths (n) | 2220 | 1862 | 493 | 86 | (-) | 1800 | 1657 | 428 | 71 | (-) |
| Events per 100,000 person days | 5.1 | 1.71 | 1.12 | 1.15 | (-) | 5.17 | 1.79 | 1.1 | 1.05 | (-) |
| Crude HR (95%CI) | 4.42 (3.57 - 5.49) | 1.49 (1.20 - 1.84) | 0.97 (0.77 - 1.22) | Ref | (-) | 4.91 (3.88 - 6.23) | 1.70 (1.34 - 2.16) | 1.04 (0.81 - 1.34) | Ref | (-) |
| Age adjusted HR (95% CI) | 0.77 (0.62 - 0.95) | 0.81 (0.65 - 1.01) | 0.97 (0.77 - 1.22) | Ref | (-) | 0.82 (0.65 - 1.05) | 0.94 (0.74 - 1.19) | 1.05 (0.82 - 1.35) | Ref | (-) |
| Age and gender adjusted HR (95% CI) | 0.75 (0.61 - 0.93) | 0.80 (0.64 - 0.99) | 0.97 (0.77 - 1.21) | Ref | (-) | 0.81 (0.64 - 1.02) | 0.92 (0.73 - 1.17) | 1.05 (0.81 - 1.35) | Ref | (-) |
| Age, gender and nursing home adjusted HR (95% CI) | 0.76 (0.61 - 0.94) | 0.80 (0.65 - 1.00) | 0.96 (0.76 - 1.21) | Ref | (-) | 0.78 (0.61 - 0.99) | 0.92 (0.72 - 1.16) | 1.03 (0.80 - 1.33) | Ref | (-) |
| Age, gender, nursing home and year of last infection adjusted HR (95% CI) | 0.72 (0.58 - 0.90) | 0.74 (0.60 - 0.92) | 0.92 (0.73 - 1.15) | Ref | (-) | 0.77 (0.61 - 0.99) | 0.91 (0.71 - 1.15) | 1.01 (0.78 - 1.29) | Ref | (-) |

| **Table S37:** Hazard ratios (HR) with 95% confidence intervals (95% CI) for non-COVID 19 and all-cause mortality according to number of SARS-CoV-2 vaccine doses during different time periods for **non-mRNA** vaccinated only. | | | | | | | | | | |
| --- | --- | --- | --- | --- | --- | --- | --- | --- | --- | --- |
|  | 2021 | | | | | | | | | |
|  | June and July 2021 (low COVID-19 disease burden) | | | | | October and November 2021 (high COVID-19 disease burden) | | | | |
|  | Four or more vaccine doses | Three vaccine doses | Two vaccine doses | One vaccine dose | Unvaccinated | Four or more vaccine doses | Three vaccine doses | Two vaccine doses | One vaccine dose | Unvaccinated |
|  | All-cause mortality | | | | | All-cause mortality | | | | |
| Deaths (n) |  |  | 1 | 20 | (-) |  |  | 13 | 34 | (-) |
| Events per 100,000 person days |  |  | 0.11 | 1.48 | (-) |  |  | 0.95 | 1.60 | (-) |
| Crude HR (95%CI) |  |  | 0.07 (0.01 - 0.56) | Ref | (-) |  |  | 0.59 (0.31 - 1.13) | Ref | (-) |
| Age adjusted HR (95% CI) |  |  | 0.09 (0.01 - 0.70) | Ref | (-) |  |  | 0.53 (0.28 - 1.00) | Ref | (-) |
| Age and gender adjusted HR (95% CI) |  |  | 0.09 (0.01 - 0.68) | Ref | (-) |  |  | 0.51 (0.27 - 0.98) | Ref | (-) |
| Age, gender and nursing home adjusted HR (95% CI) |  |  | 0.10 (0.01 - 0.78) | Ref | (-) |  |  | 0.52 (0.27 - 0.99) | Ref | (-) |
| Age, gender, nursing home and year of last infection adjusted HR (95% CI) |  |  | 0.10 (0.01 - 0.79) | Ref | (-) |  |  | 0.52 (0.27 - 0.99) | Ref | (-) |
|  | Non-COVID-19 mortality | | | | | Non-COVID-19 mortality | | | | |
| Deaths (n) |  |  | 1 | 19 | (-) |  |  | 13 | 33 | (-) |
| Events per 100,000 person days |  |  | 0.11 | 1.41 | (-) |  |  | 0.95 | 1.56 | (-) |
| Crude HR (95%CI) |  |  | 0.08 (0.01 - 0.59) | Ref | (-) |  |  | 0.61 (0.32 - 1.17) | Ref | (-) |
| Age adjusted HR (95% CI) |  |  | 0.10 (0.01 - 0.73) | Ref | (-) |  |  | 0.54 (0.28 - 1.04) | Ref | (-) |
| Age and gender adjusted HR (95% CI) |  |  | 0.09 (0.01 - 0.71) | Ref | (-) |  |  | 0.53 (0.28 - 1.01) | Ref | (-) |
| Age, gender and nursing home adjusted HR (95% CI) |  |  | 0.11 (0.01 - 0.83) | Ref | (-) |  |  | 0.54 (0.28 - 1.03) | Ref | (-) |
| Age, gender, nursing home and year of last infection adjusted HR (95% CI) | | | 0.11 (0.01 - 0.83) | Ref | (-) |  |  | 0.54 (0.28 - 1.03) | Ref | (-) |
|  | May and June 2022 (low COVID-19 disease burden) | | | | | February and March 2022 (high COVID-19 disease burden) | | | | |
|  | Four or more vaccine doses | Three vaccine doses | Two vaccine doses | One vaccine dose | Unvaccinated | Four or more vaccine doses | Three vaccine doses | Two vaccine doses | One vaccine dose | Unvaccinated |
|  | All-cause mortality | | | | | All-cause mortality | | | | |
| Deaths (n) |  | 2 | 14 | 9 | (-) |  |  | 6* | 2 | (-) |
| Events per 100,000 person days |  | 6.03 | 1.34 | 1.12 | (-) |  |  | 1.33 | 0.67 | (-) |
| Crude HR (95%CI) |  | 5.31 (1.15 - 24.64) | 1.18 (0.51 - 2.72) | Ref | (-) |  |  |  | Ref | (-) |
| Age adjusted HR (95% CI) |  | 1.77 (0.38 - 8.23) | 0.61 (0.26 - 1.41) | Ref | (-) |  |  |  | Ref | (-) |
| Age and gender adjusted HR (95% CI) |  | 1.75 (0.38 - 8.15) | 0.61 (0.26 - 1.42) | Ref | (-) |  |  |  | Ref | (-) |
| Age, gender and nursing home adjusted HR (95% CI) |  | 1.77 (0.38 - 8.25) | 0.63 (0.27 - 1.46) | Ref | (-) |  |  |  | Ref | (-) |
| Age, gender, nursing home and year of last infection adjusted HR (95% CI) |  | 1.95 (0.41 - 9.17) | 0.62 (0.27 - 1.45) | Ref | (-) |  |  |  | Ref | (-) |
|  | Non-COVID-19 mortality | | | | | Non-COVID-19 mortality | | | | |
| Deaths (n) |  | 2 | 14 | 9 | (-) |  |  | 6 | 2 | (-) |
| Events per 100,000 person days |  | 6.03 | 1.34 | 1.12 | (-) |  |  | 1.33 | 0.67 | (-) |
| Crude HR (95%CI) |  | 5.31 (1.15 - 24.64) | 1.18 (0.51 - 2.72) | Ref | (-) |  |  | 1.91 (0.39 - 9.48) | Ref | (-) |
| Age adjusted HR (95% CI) |  | 1.77 (0.38 - 8.23) | 0.61 (0.26 - 1.41) | Ref | (-) |  |  | 1.32 (0.26 - 6.85) | Ref | (-) |
| Age and gender adjusted HR (95% CI) |  | 1.75 (0.38 - 8.15) | 0.61 (0.26 - 1.42) | Ref | (-) |  |  | 1.31 (0.25 - 6.79) | Ref | (-) |
| Age, gender and nursing home adjusted HR (95% CI) |  | 1.77 (0.38 - 8.25) | 0.63 (0.27 - 1.46) | Ref | (-) |  |  | 1.35 (0.26 - 6.98) | Ref | (-) |
| Age, gender, nursing home and year of last infection adjusted HR (95% CI) | | 1.95 (0.41 - 9.17) | 0.62 (0.27 - 1.45) | Ref | (-) |  |  | 1.13 (0.20 - 6.52) | Ref | (-) |

| **Table S37 continued:** | | | | | | | | | | | |
| --- | --- | --- | --- | --- | --- | --- | --- | --- | --- | --- | --- |
|  | | July and August 2023 (low COVID-19 disease burden) | | | | | February and March 2023 (high COVID-19 disease burden) | | | | |
|  | Four or more vaccine doses | | Three vaccine doses | Two vaccine doses | One vaccine dose | Unvaccinated | Four or more vaccine doses | Three vaccine doses | Two vaccine doses | One vaccine dose | Unvaccinated |
|  | | All-cause mortality | | | | | All-cause mortality | | | | |
| Deaths (n) | | 6 | 1 | 17 | 14 | (-) | 4 | 4 | 16 | 11 | (-) |
| Events per 100,000 person days | | 6.27 | 0.61 | 1.14 | 1.14 | (-) | 5.57 | 2.96 | 1.21 | 0.98 | (-) |
| Crude HR (95%CI) | | 5.52 (2.12 - 14.35) | 0.54 (0.07 - 4.11) | 1.00 (0.49 - 2.03) | Ref | (-) | 5.67 (1.80 - 17.80) | 3.02 (0.96 - 9.47) | 1.23 (0.57 - 2.65) | Ref | (-) |
| Age adjusted HR (95% CI) | | 1.06 (0.37 - 3.01) | 0.31 (0.04 - 2.34) | 0.71 (0.35 - 1.44) | Ref | (-) | 0.96 (0.29 - 3.23) | 1.49 (0.47 - 4.70) | 0.87 (0.41 - 1.88) | Ref | (-) |
| Age and gender adjusted HR (95% CI) | | 1.05 (0.37 - 2.99) | 0.27 (0.04 - 2.05) | 0.71 (0.35 - 1.44) | Ref | (-) | 0.94 (0.28 - 3.19) | 1.52 (0.48 - 4.81) | 0.87 (0.40 - 1.87) | Ref | (-) |
| Age, gender and nursing home adjusted HR (95% CI) | | 1.05 (0.37 - 2.99) | 0.27 (0.04 - 2.04) | 0.70 (0.34 - 1.42) | Ref | (-) | 0.93 (0.27 - 3.16) | 1.53 (0.48 - 4.84) | 0.87 (0.40 - 1.88) | Ref | (-) |
| Age, gender, nursing home and year of last infection adjusted HR (95% CI) | | 1.09 (0.37 - 3.26) | 0.27 (0.04 - 2.06) | 0.70 (0.34 - 1.43) | Ref | (-) | 0.85 (0.24 - 2.98) | 1.51 (0.48 - 4.79) | 0.87 (0.40 - 1.88) | Ref | (-) |
|  | | Non-COVID-19 mortality | | | | | Non-COVID-19 mortality | | | | |
| Deaths (n) | | 6 | 1 | 17 | 14 | (-) | 4 | 4 | 16 | 11 | (-) |
| Events per 100,000 person days | | 6.27 | 0.61 | 1.14 | 1.14 | (-) | 5.57 | 2.96 | 1.21 | 0.98 | (-) |
| Crude HR (95%CI) | | 5.52 (2.12 - 14.35) | 0.54 (0.07 - 4.11) | 1.00 (0.49 - 2.03) | Ref | (-) | 5.67 (1.80 - 17.80) | 3.02 (0.96 - 9.47) | 1.23 (0.57 - 2.65) | Ref | (-) |
| Age adjusted HR (95% CI) | | 1.06 (0.37 - 3.01) | 0.31 (0.04 - 2.34) | 0.71 (0.35 - 1.44) | Ref | (-) | 0.96 (0.29 - 3.23) | 1.49 (0.47 - 4.70) | 0.87 (0.41 - 1.88) | Ref | (-) |
| Age and gender adjusted HR (95% CI) | | 1.05 (0.37 - 2.99) | 0.27 (0.04 - 2.05) | 0.71 (0.35 - 1.44) | Ref | (-) | 0.94 (0.28 - 3.19) | 1.52 (0.48 - 4.81) | 0.87 (0.40 - 1.87) | Ref | (-) |
| Age, gender and nursing home adjusted HR (95% CI) | | 1.05 (0.37 - 2.99) | 0.27 (0.04 - 2.04) | 0.70 (0.34 - 1.42) | Ref | (-) | 0.93 (0.27 - 3.16) | 1.53 (0.48 - 4.84) | 0.87 (0.40 - 1.88) | Ref | (-) |
| Age, gender, nursing home and year of last infection adjusted HR (95% CI) | | 1.09 (0.37 - 3.26) | 0.27 (0.04 - 2.06) | 0.70 (0.34 - 1.43) | Ref | (-) | 0.85 (0.24 - 2.98) | 1.51 (0.48 - 4.79) | 0.87 (0.40 - 1.88) | Ref | (-) |

### **Additional Outcomes**

| **Table S38:** Hazard ratios (HR) with 95% confidence intervals (95% CI) for cancer and transport accident mortality according to number of SARS-CoV-2 vaccine doses over the entire period. | | | | | |
| --- | --- | --- | --- | --- | --- |
|  | Cancer | | | | |
|  | Four or more vaccine doses | Three vaccine doses | Two vaccine doses | One vaccine dose | Unvaccinated |
| Deaths (n) | 3622 | 5406 | 1956 | 484 | 2321 |
| Events per 100,000 person days | 2.73 | 1.39 | 1.07 | 0.87 | 1.14 |
| Crude HR (95%CI) | 3.65 (3.43 - 3.88) | 1.65 (1.57 - 1.75) | 1.09 (1.02 - 1.15) | 0.84 (0.76 - 0.93) | Reference |
| Age adjusted HR (95% CI) | 0.95 (0.89 - 1.01) | 1.01 (0.96 - 1.06) | 0.95 (0.89 - 1.01) | 0.81 (0.73 - 0.90) | Reference |
| Age and gender adjusted HR (95% CI) | 0.91 (0.85 - 0.97) | 0.99 (0.94 - 1.04) | 0.94 (0.89 - 1.00) | 0.81 (0.73 - 0.90) | Reference |
| Age, gender and nursing home adjusted HR (95% CI) | 0.90 (0.85 - 0.96) | 0.98 (0.93 - 1.04) | 0.94 (0.88 - 1.00) | 0.81 (0.73 - 0.90) | Reference |
|  | Transport accidents | | | | |
|  | Four or more vaccine doses | Three vaccine doses | Two vaccine doses | One vaccine dose | Unvaccinated |
| Deaths (n) | 35 | 104 | 65 | 14 | 85 |
| Events per 100,000 person days | 0.03 | 0.03 | 0.04 | 0.03 | 0.04 |
| Crude HR (95%CI) | 0.91 (0.60 - 1.40) | 0.73 (0.54 - 0.98) | 0.94 (0.68 - 1.30) | 0.80 (0.45 - 1.42) | Reference |
| Age adjusted HR (95% CI) | 0.79 (0.50 - 1.26) | 0.66 (0.49 - 0.89) | 0.94 (0.68 - 1.29) | 0.80 (0.45 - 1.42) | Reference |
| Age and gender adjusted HR (95% CI) | 0.77 (0.48 - 1.24) | 0.66 (0.49 - 0.89) | 0.92 (0.66 - 1.27) | 0.78 (0.44 - 1.39) | Reference |
| Age, gender and nursing home adjusted HR (95% CI) | 0.77 (0.48 - 1.23) | 0.66 (0.49 - 0.89) | 0.89 (0.65 - 1.24) | 0.78 (0.44 - 1.38) | Reference |

| **Table S39:** Hazard ratios (HR) with 95% confidence intervals (95% CI) for **cancer and transport accidents** mortality according to number of SARS-CoV-2 vaccine doses for the period from 2021 to 2023 split into 3-month intervals. | | | | | | | | | | | | | | | | | | | | |
| --- | --- | --- | --- | --- | --- | --- | --- | --- | --- | --- | --- | --- | --- | --- | --- | --- | --- | --- | --- | --- |
|  | 2021 | | | | | | | | | | | | | | | | | | | |
|  | first quarter | | | | | second quarter | | | | | third quarter | | | | | fourth quarter | | | | |
|  | Four or more vaccine doses | Three vaccine doses | Two vaccine doses | One vaccine dose | Unvaccinated | Four or more vaccine doses | Three vaccine doses | Two vaccine doses | One vaccine dose | Unvaccinated | Four or more vaccine doses | Three vaccine doses | Two vaccine doses | One vaccine dose | Unvaccinated | Four or more vaccine doses | Three vaccine doses | Two vaccine doses | One vaccine dose | Unvaccinated |
|  | Cancer | | | | | Cancer | | | | | Cancer | | | | | Cancer | | | | |
| Deaths (n) |  |  | 14 | 11 | 96 |  |  | 73 | 34 | 184 |  | 5 | 137 | 60 | 157 |  | 46 | 130 | 72 | 108 |
| Events per 100,000 person days |  |  | 1.92 | 1.45 | 0.70 |  |  | 1.8 | 0.5 | 0.87 |  | 10.33 | 1.02 | 0.52 | 0.84 |  | 1.11 | 0.73 | 0.54 | 0.81 |
| Crude HR (95%CI) |  |  | 2.71 (1.54 - 4.79) | 2.13 (1.13 - 4.02) | Reference |  |  | 2.23 (1.69 - 2.94) | 0.63 (0.43 - 0.91) | Reference |  | 14.72 (5.61 - 38.58) | 1.25 (0.99 - 1.57) | 0.63 (0.47 - 0.85) | Reference |  | 1.42 (0.97 - 2.09) | 0.91 (0.70 - 1.17) | 0.69 (0.51 - 0.92) | Reference |
| Age adjusted HR (95% CI) |  |  | 0.45 (0.25 - 0.83) | 0.76 (0.40 - 1.45) | Reference |  |  | 0.54 (0.40 - 0.72) | 0.40 (0.27 - 0.57) | Reference |  | 1.53 (0.57 - 4.08) | 0.59 (0.47 - 0.74) | 0.48 (0.35 - 0.64) | Reference |  | 0.34 (0.24 - 0.50) | 0.59 (0.45 - 0.76) | 0.60 (0.45 - 0.81) | Reference |
| Age and gender adjusted HR (95% CI) |  |  | 0.47 (0.26 - 0.86) | 0.77 (0.41 - 1.46) | Reference |  |  | 0.55 (0.41 - 0.73) | 0.39 (0.27 - 0.56) | Reference |  | 1.59 (0.60 - 4.24) | 0.58 (0.46 - 0.73) | 0.47 (0.35 - 0.63) | Reference |  | 0.34 (0.23 - 0.50) | 0.58 (0.45 - 0.75) | 0.59 (0.44 - 0.80) | Reference |
| Age, gender and nursing home adjusted HR (95% CI) |  |  | 0.60 (0.30 - 1.18) | 0.79 (0.41 - 1.50) | Reference |  |  | 0.51 (0.38 - 0.70) | 0.39 (0.27 - 0.56) | Reference |  | 1.38 (0.50 - 3.80) | 0.56 (0.44 - 0.71) | 0.47 (0.35 - 0.63) | Reference |  | 0.33 (0.23 - 0.49) | 0.58 (0.45 - 0.75) | 0.60 (0.45 - 0.82) | Reference |
|  | Transport Accidents | | | | | Transport Accidents | | | | | Transport Accidents | | | | | Transport Accidents | | | | |
| Deaths (n) |  |  |  |  | 1 |  |  | 1* |  | 2 |  |  | 3* | 2* | 6 |  |  | 1 |  | 0 |
| Events per 100,000 person days |  |  |  |  | 0.01 |  |  | 0.02 |  | 0.01 |  |  | 0.02 | 0.02 | 0.03 |  |  | 0.01 |  | 0.00 |
| Crude HR (95%CI) |  |  |  |  | Reference |  |  |  |  | Reference |  |  |  |  | Reference |  |  |  |  | Reference |
| Age adjusted HR (95% CI) |  |  |  |  | Reference |  |  |  |  | Reference |  |  |  |  | Reference |  |  |  |  | Reference |
| Age and gender adjusted HR (95% CI) |  |  |  |  | Reference |  |  |  |  | Reference |  |  |  |  | Reference |  |  |  |  | Reference |
| Age, gender and nursing home adjusted HR (95% CI) |  |  |  |  | Reference |  |  |  |  | Reference |  |  |  |  | Reference |  |  |  |  | Reference |
|  | 2022 | | | | | | | | | | | | | | | | | | | |
|  | first quarter | | | | | second quarter | | | | | third quarter | | | | | fourth quarter | | | | |
|  | Four or more vaccine doses | Three vaccine doses | Two vaccine doses | One vaccine dose | Unvaccinated | Four or more vaccine doses | Three vaccine doses | Two vaccine doses | One vaccine dose | Unvaccinated | Four or more vaccine doses | Three vaccine doses | Two vaccine doses | One vaccine dose | Unvaccinated | Four or more vaccine doses | Three vaccine doses | Two vaccine doses | One vaccine dose | Unvaccinated |
|  | Cancer | | | | | Cancer | | | | | Cancer | | | | | Cancer | | | | |
| Deaths (n) | 1 | 132 | 154 | 46 | 84 | 11 | 376 | 213 | 38 | 125 | 99 | 795 | 225 | 54 | 193 | 408 | 829 | 213 | 41 | 243 |
| Events per 100,000 person days | 7.84 | 0.81 | 0.69 | 0.5 | 0.46 | 5.26 | 0.69 | 0.53 | 0.38 | 0.28 | 1.76 | 0.68 | 0.42 | 0.5 | 0.31 | 1.3 | 0.66 | 0.38 | 0.37 | 0.37 |
| Crude HR (95%CI) | 19.03 (2.65 - 136.84) | 1.71 (1.30 - 2.24) | 1.49 (1.14 - 1.95) | 1.01 (0.70 - 1.45) | Reference | 20.51 (10.94 - 38.45) | 2.49 (2.03 - 3.05) | 1.88 (1.51 - 2.35) | 1.34 (0.93 - 1.92) | Reference | 5.74 (4.47 - 7.38) | 2.22 (1.89 - 2.59) | 1.35 (1.11 - 1.63) | 1.62 (1.20 - 2.19) | Reference | 3.54 (3.02 - 4.16) | 1.83 (1.59 - 2.12) | 1.05 (0.88 - 1.26) | 1.02 (0.73 - 1.42) | Reference |
| Age adjusted HR (95% CI) | 3.18 (0.44 - 23.03) | 0.62 (0.47 - 0.83) | 0.87 (0.67 - 1.14) | 1.00 (0.70 - 1.44) | Reference | 1.76 (0.92 - 3.38) | 1.01 (0.82 - 1.24) | 1.38 (1.10 - 1.72) | 1.35 (0.94 - 1.94) | Reference | 0.69 (0.52 - 0.90) | 1.13 (0.96 - 1.32) | 1.31 (1.08 - 1.59) | 1.78 (1.31 - 2.40) | Reference | 0.85 (0.71 - 1.01) | 1.21 (1.05 - 1.40) | 1.18 (0.98 - 1.42) | 1.20 (0.86 - 1.67) | Reference |
| Age and gender adjusted HR (95% CI) | ( - ) | 0.60 (0.45 - 0.79) | 0.86 (0.66 - 1.13) | 0.99 (0.69 - 1.43) | Reference | 1.73 (0.90 - 3.31) | 0.98 (0.79 - 1.20) | 1.35 (1.08 - 1.69) | 1.34 (0.93 - 1.94) | Reference | 0.67 (0.51 - 0.88) | 1.10 (0.94 - 1.29) | 1.30 (1.07 - 1.58) | 1.77 (1.31 - 2.39) | Reference | 0.81 (0.68 - 0.97) | 1.18 (1.02 - 1.37) | 1.17 (0.98 - 1.41) | 1.20 (0.86 - 1.67) | Reference |
| Age, gender and nursing home adjusted HR (95% CI) | 3.45 (0.47 - 25.11) | 0.58 (0.43 - 0.77) | 0.87 (0.66 - 1.13) | 0.99 (0.69 - 1.42) | Reference | 1.70 (0.87 - 3.31) | 0.95 (0.77 - 1.17) | 1.33 (1.06 - 1.66) | 1.34 (0.93 - 1.94) | Reference | 0.64 (0.48 - 0.84) | 1.09 (0.93 - 1.28) | 1.31 (1.08 - 1.58) | 1.77 (1.31 - 2.39) | Reference | 0.78 (0.66 - 0.94) | 1.18 (1.02 - 1.36) | 1.17 (0.97 - 1.41) | 1.19 (0.86 - 1.66) | Reference |

| **Table S39 continued:** | | | | | | | | | | | | | | | | | | | | |
| --- | --- | --- | --- | --- | --- | --- | --- | --- | --- | --- | --- | --- | --- | --- | --- | --- | --- | --- | --- | --- |
|  | 2022 | | | | | | | | | | | | | | | | | | | |
|  | first quarter | | | | | second quarter | | | | | third quarter | | | | | fourth quarter | | | | |
|  | Four or more vaccine doses | Three vaccine doses | Two vaccine doses | One vaccine dose | Unvacc | Four or more vaccine doses | Three vaccine doses | Two vaccine doses | One vaccine dose | Unvacc | Four or more vaccine doses | Three vaccine doses | Two vaccine doses | One vaccine dose | Unvacc | Four or more vaccine doses | Three vaccine doses | Two vaccine doses | One vaccine dose | Unvacc |
|  | Transport Accidents | | | | | Transport Accidents | | | | | Transport Accidents | | | | | Transport Accidents | | | | |
| Deaths (n) |  | 2* |  | 1* | 2 |  | 4 | 5 | 2* | 7 | 1 | 13 | 5 |  | 17 | 3* | 11 | 11 |  | 4 |
| Events per 100,000 person days |  | 0.01 |  | 0.01 | 0.01 |  | 0.01 | 0.01 | 0.02 | 0.02 | 0.02 | 0.01 | 0.01 |  | 0.03 | 0.01 | 0.01 | 0.02 |  | 0.01 |
| Crude HR (95%CI) |  |  |  |  | Reference |  | 0.42 (0.12 - 1.46) | 0.79 (0.25 - 2.49) |  | Reference | 0.81 (0.11 - 6.14) | 0.41 (0.20 - 0.85) | 0.34 (0.12 - 0.92) |  | Reference |  | 1.45 (0.46 - 4.57) | 3.30 (1.05 - 10.36) |  | Reference |
| Age adjusted HR (95% CI) |  |  |  |  | Reference |  | 0.34 (0.09 - 1.21) | 0.78 (0.25 - 2.46) |  | Reference | 0.62 (0.07 - 5.55) | 0.35 (0.17 - 0.73) | 0.34 (0.12 - 0.92) |  | Reference |  | 1.40 (0.44 - 4.44) | 3.26 (1.03 - 10.26) |  | Reference |
| Age and gender adjusted HR (95% CI) |  |  |  |  | Reference |  | 0.33 (0.09 - 1.18) | 0.77 (0.24 - 2.42) |  | Reference | ( - ) | 0.34 (0.16 - 0.72) | ( - ) |  | Reference |  | 1.40 (0.44 - 4.45) | 3.21 (1.02 - 10.11) |  | Reference |
| Age, gender and nursing home adjusted HR (95% CI) |  |  |  |  | Reference |  | 0.33 (0.09 - 1.19) | 0.77 (0.24 - 2.43) |  | Reference | 0.70 (0.08 - 6.09) | 0.35 (0.17 - 0.72) | 0.34 (0.13 - 0.92) |  | Reference |  | 1.39 (0.44 - 4.41) | 3.08 (0.98 - 9.71) |  | Reference |
|  | 2023 | | | | | | | | | | | | | | | | | | | |
|  | first quarter | | | | | second quarter | | | | | third quarter | | | | | fourth quarter | | | | |
|  | Four or more vaccine doses | Three vaccine doses | Two vaccine doses | One vaccine dose | Unvaccinated | Four or more vaccine doses | Three vaccine doses | Two vaccine doses | One vaccine dose | Unvaccinated | Four or more vaccine doses | Three vaccine doses | Two vaccine doses | One vaccine dose | Unvaccinated | Four or more vaccine doses | Three vaccine doses | Two vaccine doses | One vaccine dose | Unvaccinated |
|  | Cancer | | | | | Cancer | | | | | Cancer | | | | | Cancer | | | | |
| Deaths (n) | 607 | 828 | 198 | 25 | 258 | 776 | 745 | 197 | 34 | 298 | 864 | 820 | 203 | 30 | 266 | 856 | 830 | 199 | 39 | 309 |
| Events per 100,000 person days | 1.27 | 0.64 | 0.35 | 0.23 | 0.38 | 1.42 | 0.54 | 0.34 | 0.3 | 0.43 | 1.5 | 0.57 | 0.34 | 0.26 | 0.37 | 1.49 | 0.58 | 0.33 | 0.34 | 0.43 |
| Crude HR (95%CI) | 3.30 (2.85 - 3.81) | 1.67 (1.45 - 1.92) | 0.93 (0.77 - 1.11) | 0.60 (0.40 - 0.91) | Reference | 3.33 (2.91 - 3.81) | 1.26 (1.10 - 1.44) | 0.79 (0.66 - 0.95) | 0.71 (0.50 - 1.02) | Reference | 4.00 (3.49 - 4.59) | 1.53 (1.34 - 1.76) | 0.91 (0.76 - 1.09) | 0.70 (0.48 - 1.03) | Reference | 3.43 (3.01 - 3.91) | 1.34 (1.18 - 1.53) | 0.77 (0.64 - 0.92) | 0.79 (0.56 - 1.10) | Reference |
| Age adjusted HR (95% CI) | 0.91 (0.78 - 1.07) | 1.16 (1.01 - 1.33) | 1.09 (0.91 - 1.31) | 0.73 (0.48 - 1.10) | Reference | 0.95 (0.82 - 1.09) | 0.88 (0.77 - 1.01) | 0.95 (0.79 - 1.14) | 0.88 (0.62 - 1.26) | Reference | 1.19 (1.03 - 1.38) | 1.08 (0.94 - 1.24) | 1.09 (0.91 - 1.31) | 0.87 (0.60 - 1.28) | Reference | 0.96 (0.83 - 1.10) | 0.94 (0.83 - 1.07) | 0.92 (0.77 - 1.10) | 0.98 (0.70 - 1.37) | Reference |
| Age and gender adjusted HR (95% CI) | 0.87 (0.75 - 1.02) | 1.13 (0.99 - 1.31) | 1.08 (0.90 - 1.30) | 0.73 (0.48 - 1.10) | Reference | 0.89 (0.78 - 1.03) | 0.87 (0.76 - 1.00) | 0.94 (0.79 - 1.13) | 0.88 (0.61 - 1.25) | Reference | 1.15 (0.99 - 1.33) | 1.06 (0.92 - 1.22) | 1.09 (0.91 - 1.31) | 0.87 (0.60 - 1.27) | Reference | 0.92 (0.80 - 1.06) | 0.92 (0.81 - 1.05) | 0.91 (0.76 - 1.09) | 0.98 (0.70 - 1.36) | Reference |
| Age, gender and nursing home adjusted HR (95% CI) | 0.87 (0.74 - 1.01) | 1.13 (0.98 - 1.30) | 1.08 (0.90 - 1.30) | 0.72 (0.48 - 1.09) | Reference | 0.89 (0.77 - 1.03) | 0.87 (0.76 - 1.00) | 0.94 (0.79 - 1.13) | 0.88 (0.61 - 1.25) | Reference | 1.15 (0.99 - 1.33) | 1.06 (0.92 - 1.22) | 1.09 (0.91 - 1.31) | 0.87 (0.60 - 1.28) | Reference | 0.92 (0.80 - 1.05) | 0.92 (0.81 - 1.05) | 0.91 (0.76 - 1.09) | 0.97 (0.70 - 1.36) | Reference |
|  | Transport Accidents | | | | | Transport Accidents | | | | | Transport Accidents | | | | | Transport Accidents | | | | |
| Deaths (n) | 7 | 13 | 6 | 4 | 11 | 7 | 19 | 12 | 1 | 15 | 11 | 28 | 15 | 2 | 11 | 6 | 14 | 6 | 2 | 9 |
| Events per 100,000 person days | 0.01 | 0.01 | 0.01 | 0.04 | 0.02 | 0.01 | 0.01 | 0.02 | 0.01 | 0.02 | 0.02 | 0.02 | 0.03 | 0.02 | 0.02 | 0.01 | 0.01 | 0.01 | 0.02 | 0.01 |
| Crude HR (95%CI) | 0.89 (0.34 - 2.28) | 0.61 (0.28 - 1.37) | 0.66 (0.24 - 1.78) | 2.25 (0.72 - 7.08) | Reference | 0.60 (0.24 - 1.47) | 0.64 (0.32 - 1.25) | 0.96 (0.45 - 2.05) | 0.42 (0.05 - 3.15) | Reference | 1.23 (0.53 - 2.84) | 1.27 (0.63 - 2.54) | 1.63 (0.75 - 3.55) | 1.14 (0.25 - 5.13) | Reference | 0.83 (0.29 - 2.32) | 0.78 (0.34 - 1.79) | 0.79 (0.28 - 2.23) | 1.39 (0.30 - 6.42) | Reference |
| Age adjusted HR (95% CI) | 0.57 (0.20 - 1.66) | 0.54 (0.24 - 1.21) | 0.67 (0.25 - 1.81) | 2.33 (0.74 - 7.36) | Reference | 0.56 (0.21 - 1.52) | 0.60 (0.31 - 1.19) | 0.95 (0.44 - 2.03) | 0.42 (0.05 - 3.16) | Reference | 1.13 (0.44 - 2.89) | 1.21 (0.60 - 2.45) | 1.56 (0.71 - 3.40) | 1.06 (0.23 - 4.79) | Reference | 1.09 (0.35 - 3.40) | 0.69 (0.30 - 1.60) | 0.69 (0.24 - 1.95) | 1.27 (0.27 - 5.92) | Reference |
| Age and gender adjusted HR (95% CI) | 0.55 (0.19 - 1.62) | 0.54 (0.24 - 1.21) | 0.66 (0.24 - 1.79) | ( - ) | Reference | 0.55 (0.20 - 1.51) | 0.60 (0.30 - 1.19) | 0.93 (0.44 - 2.00) | ( - ) | Reference | 1.12 (0.43 - 2.88) | 1.21 (0.60 - 2.45) | 1.52 (0.70 - 3.33) | ( - ) | Reference | 1.10 (0.35 - 3.44) | 0.68 (0.29 - 1.58) | 0.68 (0.24 - 1.92) | ( - ) | Reference |
| Age, gender and nursing home adjusted HR (95% CI) | 0.56 (0.19 - 1.62) | 0.54 (0.24 - 1.21) | 0.66 (0.24 - 1.79) | 2.34 (0.74 - 7.38) | Reference | 0.56 (0.20 - 1.51) | 0.60 (0.30 - 1.19) | 0.93 (0.44 - 2.00) | 0.42 (0.05 - 3.17) | Reference | 1.12 (0.43 - 2.87) | 1.22 (0.60 - 2.45) | 1.49 (0.68 - 3.26) | 1.06 (0.23 - 4.80) | Reference | 1.10 (0.35 - 3.44) | 0.68 (0.29 - 1.58) | 0.68 (0.24 - 1.92) | 1.27 (0.27 - 5.93) | Reference |
| * HRs not reported because there were less than 10 events total (including reference group) | | | | | | | | | | | | | | | | | | | | |

| **Table S40:** Hazard ratios (HR) with 95% confidence intervals (95% CI) for cancer and transport accidents according to number of SARS-CoV-2 vaccine doses during different time periods of high and low COVID-19 disease burden. | | | | | | | | | | |
| --- | --- | --- | --- | --- | --- | --- | --- | --- | --- | --- |
|  | June and July 2021 (low COVID-19 disease burden) | | | | | October and November 2021 (high COVID-19 disease burden) | | | | |
|  | Four or more vaccine doses | Three vaccine doses | Two vaccine doses | One vaccine dose | Unvaccinated | Four or more vaccine doses | Three vaccine doses | Two vaccine doses | One vaccine dose | Unvaccinated |
|  | Cancer | | | | | Cancer | | | | |
| Deaths (n) | 0 | 0 | 54 | 29 | 100 | 0 | 12 | 84 | 50 | 72 |
| Events per 100,000 person days |  |  | 0.95 | 0.4 | 0.87 |  | 1.02 | 0.68 | 0.51 | 0.72 |
| Crude HR (95%CI) |  |  | 1.06 (0.76 - 1.48) | 0.45 (0.30 - 0.68) | Reference |  | 1.43 (0.75 - 2.73) | 0.93 (0.68 - 1.27) | 0.72 (0.50 - 1.04) | Reference |
| Age adjusted HR (95% CI) |  |  | 0.42 (0.29 - 0.58) | 0.38 (0.25 - 0.57) | Reference |  | 0.23 (0.12 - 0.44) | 0.59 (0.43 - 0.80) | 0.62 (0.43 - 0.88) | Reference |
| Age and gender adjusted HR (95% CI) |  |  | 0.41 (0.29 - 0.58) | 0.37 (0.24 - 0.56) | Reference |  | 0.23 (0.12 - 0.44) | 0.58 (0.42 - 0.79) | 0.61 (0.42 - 0.87) | Reference |
| Age, gender and nursing home adjusted HR (95% CI) |  |  | 0.40 (0.28 - 0.57) | 0.36 (0.24 - 0.55) | Reference |  | 0.25 (0.13 - 0.48) | 0.58 (0.42 - 0.80) | 0.62 (0.43 - 0.89) | Reference |
| Age, gender, nursing home and year of last infection adjusted HR (95% CI) |  |  | 0.40 (0.28 - 0.56) | 0.37 (0.24 - 0.55) | Reference |  | 0.25 (0.13 - 0.48) | 0.57 (0.42 - 0.79) | 0.62 (0.43 - 0.89) | Reference |
|  | Transport accidents | | | | | Transport accidents | | | | |
| Deaths (n) | 0 | 0 | 4* | 1* | 2 | 0 | 0 | 1* | 0 | 0 |
| Events per 100,000 person days |  |  | 0.07 | 0.01 | 0.02 |  |  | 0.01 |  |  |
| Crude HR (95%CI) |  |  |  |  | Reference |  |  |  |  | Reference |
| Age adjusted HR (95% CI) |  |  |  |  | Reference |  |  |  |  | Reference |
| Age and gender adjusted HR (95% CI) |  |  |  |  | Reference |  |  |  |  | Reference |
| Age, gender and nursing home adjusted HR (95% CI) |  |  |  |  | Reference |  |  |  |  | Reference |
| Age, gender, nursing home and year of last infection adjusted HR (95% CI) | | |  |  | Reference |  |  |  |  | Reference |
|  | May and June 2022 (low COVID-19 disease burden) | | | | | February and March 2022 (high COVID-19 disease burden) | | | | |
|  | Four or more vaccine doses | Three vaccine doses | Two vaccine doses | One vaccine dose | Unvaccinated | Four or more vaccine doses | Three vaccine doses | Two vaccine doses | One vaccine dose | Unvaccinated |
|  | Cancer | | | | | Cancer | | | | |
| Deaths (n) | 8 | 155 | 98 | 18 | 69 | 1 | 78 | 85 | 23 | 39 |
| Events per 100,000 person days | 6.89 | 0.65 | 0.44 | 0.32 | 0.30 | 10.13 | 0.66 | 0.65 | 0.44 | 0.49 |
| Crude HR (95%CI) | 22.41 (10.68 - 47.01) | 2.19 (1.65 - 2.91) | 1.49 (1.10 - 2.03) | 1.07 (0.64 - 1.80) | Reference | 20.28 (2.78 - 147.78) | 1.36 (0.92 - 2.00) | 1.33 (0.91 - 1.95) | 0.89 (0.53 - 1.49) | Reference |
| Age adjusted HR (95% CI) | 1.74 (0.80 - 3.80) | 0.82 (0.61 - 1.10) | 1.00 (0.74 - 1.37) | 1.02 (0.61 - 1.71) | Reference | 4.22 (0.57 - 31.16) | 0.59 (0.40 - 0.87) | 0.87 (0.59 - 1.27) | 0.97 (0.58 - 1.62) | Reference |
| Age and gender adjusted HR (95% CI) | 1.72 (0.79 - 3.74) | 0.80 (0.60 - 1.07) | 0.99 (0.72 - 1.35) | 1.02 (0.60 - 1.71) | Reference | 4.04 (0.55 - 29.79) | 0.55 (0.37 - 0.82) | 0.85 (0.58 - 1.25) | 0.95 (0.57 - 1.59) | Reference |
| Age, gender and nursing home adjusted HR (95% CI) | 1.60 (0.70 - 3.67) | 0.77 (0.57 - 1.03) | 0.99 (0.72 - 1.35) | 1.01 (0.60 - 1.70) | Reference | 4.05 (0.54 - 30.32) | 0.53 (0.36 - 0.79) | 0.85 (0.58 - 1.25) | 0.95 (0.57 - 1.60) | Reference |
| Age, gender, nursing home and year of last infection adjusted HR (95% CI) | 1.53 (0.66 - 3.51) | 0.76 (0.56 - 1.02) | 0.94 (0.69 - 1.30) | 0.99 (0.58 - 1.68) | Reference | 4.27 (0.57 - 32.26) | 0.54 (0.36 - 0.81) | 0.86 (0.59 - 1.26) | 0.96 (0.57 - 1.62) | Reference |
|  | Transport accidents | | | | | Transport accidents | | | | |
| Deaths (n) | 0 | 2* | 1* | 1* | 2 | 0 | 2* | 0 | 1* | 0 |
| Events per 100,000 person days |  | 0.01 | 0 | 0.02 | 0.01 |  | 0.02 |  | 0.02 |  |
| Crude HR (95%CI) |  |  |  |  | Reference |  |  |  |  | Reference |
| Age adjusted HR (95% CI) |  |  |  |  | Reference |  |  |  |  | Reference |
| Age and gender adjusted HR (95% CI) |  |  |  |  | Reference |  |  |  |  | Reference |
| Age, gender and nursing home adjusted HR (95% CI) |  |  |  |  | Reference |  |  |  |  | Reference |
| Age, gender, nursing home and year of last infection adjusted HR (95% CI) | | |  |  | Reference |  |  |  |  | Reference |

| **Table S40 continued:** | | | | | | | | | | |
| --- | --- | --- | --- | --- | --- | --- | --- | --- | --- | --- |
|  | July and August 2023 (low COVID-19 disease burden) | | | | | February and March 2023 (high COVID-19 disease burden) | | | | |
|  | Four or more vaccine doses | Three vaccine doses | Two vaccine doses | One vaccine dose | Unvaccinated | Four or more vaccine doses | Three vaccine doses | Two vaccine doses | One vaccine dose | Unvaccinated |
|  | Cancer | | | | | Cancer | | | | |
| Deaths (n) | 561 | 553 | 134 | 18 | 155 | 388 | 479 | 114 | 20 | 152 |
| Events per 100,000 person days | 1.28 | 0.51 | 0.3 | 0.21 | 0.28 | 1.11 | 0.52 | 0.28 | 0.25 | 0.31 |
| Crude HR (95%CI) | 4.51 (3.77 - 5.39) | 1.78 (1.49 - 2.13) | 1.04 (0.82 - 1.31) | 0.73 (0.45 - 1.18) | Reference | 3.59 (2.98 - 4.33) | 1.67 (1.40 - 2.01) | 0.91 (0.72 - 1.17) | 0.82 (0.52 - 1.31) | Reference |
| Age adjusted HR (95% CI) | 1.34 (1.10 - 1.61) | 1.25 (1.04 - 1.49) | 1.24 (0.98 - 1.56) | 0.90 (0.55 - 1.47) | Reference | 0.98 (0.80 - 1.20) | 1.16 (0.96 - 1.39) | 1.07 (0.84 - 1.37) | 1.00 (0.63 - 1.59) | Reference |
| Age and gender adjusted HR (95% CI) | 1.29 (1.07 - 1.56) | 1.23 (1.03 - 1.47) | 1.24 (0.98 - 1.56) | 0.90 (0.56 - 1.47) | Reference | 0.94 (0.77 - 1.14) | 1.13 (0.94 - 1.36) | 1.06 (0.83 - 1.36) | 0.99 (0.62 - 1.59) | Reference |
| Age, gender and nursing home adjusted HR (95% CI) | 1.29 (1.07 - 1.56) | 1.23 (1.03 - 1.47) | 1.24 (0.99 - 1.57) | 0.91 (0.56 - 1.48) | Reference | 0.93 (0.76 - 1.14) | 1.13 (0.94 - 1.36) | 1.06 (0.83 - 1.35) | 0.99 (0.62 - 1.58) | Reference |
| Age, gender, nursing home and year of last infection adjusted HR (95% CI) | 1.23 (1.01 - 1.48) | 1.18 (0.98 - 1.41) | 1.32 (1.05 - 1.67) | 1.07 (0.65 - 1.75) | Reference | 0.93 (0.76 - 1.13) | 1.12 (0.94 - 1.35) | 1.11 (0.86 - 1.42) | 1.15 (0.71 - 1.84) | Reference |
|  | Transport accidents | | | | | Transport accidents | | | | |
| Deaths (n) | 6 | 15 | 10 | 1 | 9 | 4 | 6 | 4 | 2 | 8 |
| Events per 100,000 person days | 0.01 | 0.01 | 0.02 | 0.01 | 0.02 | 0.01 | 0.01 | 0.01 | 0.03 | 0.02 |
| Crude HR (95%CI) | 0.83 (0.30 - 2.33) | 0.83 (0.36 - 1.90) | 1.33 (0.54 - 3.28) | 0.70 (0.09 - 5.49) | Reference | 0.70 (0.21 - 2.33) | 0.40 (0.14 - 1.15) | 0.61 (0.18 - 2.02) | 1.56 (0.33 - 7.36) | Reference |
| Age adjusted HR (95% CI) | 0.91 (0.29 - 2.86) | 0.80 (0.35 - 1.84) | 1.28 (0.52 - 3.15) | 0.64 (0.08 - 5.10) | Reference | 0.46 (0.12 - 1.76) | 0.34 (0.12 - 1.00) | 0.63 (0.19 - 2.11) | 1.64 (0.35 - 7.78) | Reference |
| Age and gender adjusted HR (95% CI) | 0.90 (0.28 - 2.88) | 0.80 (0.35 - 1.84) | 1.24 (0.50 - 3.05) | 0.62 (0.08 - 4.90) | Reference | 0.46 (0.12 - 1.76) | 0.34 (0.12 - 1.00) | 0.63 (0.19 - 2.10) | 1.63 (0.34 - 7.70) | Reference |
| Age, gender and nursing home adjusted HR (95% CI) | 0.90 (0.28 - 2.88) | 0.80 (0.35 - 1.84) | 1.24 (0.50 - 3.05) | 0.62 (0.08 - 4.90) | Reference | 0.47 (0.12 - 1.77) | 0.35 (0.12 - 1.00) | 0.63 (0.19 - 2.10) | 1.63 (0.35 - 7.72) | Reference |
| Age, gender, nursing home and year of last infection adjusted HR (95% CI) | 0.94 (0.29 - 3.05) | 0.86 (0.37 - 1.99) | 1.24 (0.50 - 3.06) | 0.59 (0.07 - 4.83) | Reference | 0.50 (0.13 - 1.88) | 0.34 (0.12 - 1.00) | 0.64 (0.19 - 2.14) | 1.67 (0.34 - 8.25) | Reference |
| * HRs not reported because there were less than 10 events total (including reference group) | | | | | | | | | | |

### **COVID-19 Mortality**

| **Table S41:** Hazard ratios (HR) with 95% confidence intervals (95% CI) for COVID 19 mortality according to number of SARS-CoV-2 vaccine doses over the entire period. | | | | | |
| --- | --- | --- | --- | --- | --- |
|  | COVID-19 | | | | |
|  | Four or more vaccine doses | Three vaccine doses | Two vaccine doses | One vaccine dose | Unvaccinated |
| Deaths (n) | 333 | 447 | 226 | 49 | 291 |
| Events per 100,000 person days | 0.25 | 0.11 | 0.12 | 0.09 | 0.14 |
| Crude HR (95%CI) | 5.30 (4.23 - 6.64) | 1.54 (1.30 - 1.84) | 1.14 (0.95 - 1.37) | 0.54 (0.40 - 0.74) | Reference |
| Age adjusted HR (95% CI) | 0.91 (0.73 - 1.14) | 0.73 (0.61 - 0.86) | 0.69 (0.58 - 0.83) | 0.48 (0.35 - 0.65) | Reference |
| Age and gender adjusted HR (95% CI) | 0.87 (0.70 - 1.09) | 0.70 (0.59 - 0.83) | 0.69 (0.57 - 0.82) | 0.47 (0.34 - 0.64) | Reference |
| Age, gender and nursing home adjusted HR (95% CI) | 0.74 (0.59 - 0.93) | 0.64 (0.54 - 0.75) | 0.62 (0.52 - 0.74) | 0.47 (0.35 - 0.65) | Reference |

| **Table S42:** Hazard ratios (HR) with 95% confidence intervals (95% CI) for **COVID-19** mortality according to number of SARS-CoV-2 vaccine doses for the period from 2021 to 2023 split into 3-month intervals. | | | | | | | | | | | | | | | | | | | | |
| --- | --- | --- | --- | --- | --- | --- | --- | --- | --- | --- | --- | --- | --- | --- | --- | --- | --- | --- | --- | --- |
|  | 2021 | | | | | | | | | | | | | | | | | | | |
|  | first quarter | | | | | second quarter | | | | | third quarter | | | | | fourth quarter | | | | |
|  | Four or more vaccine doses | Three vaccine doses | Two vaccine doses | One vaccine dose | Unvaccinated | Four or more vaccine doses | Three vaccine doses | Two vaccine doses | One vaccine dose | Unvaccinated | Four or more vaccine doses | Three vaccine doses | Two vaccine doses | One vaccine dose | Unvaccinated | Four or more vaccine doses | Three vaccine doses | Two vaccine doses | One vaccine dose | Unvaccinated |
| Deaths (n) |  |  | 7 | 7 | 32 |  |  | 29 | 5 | 35 |  |  | 11 | 6 | 29 |  | 5 | 14 | 7 | 28 |
| Events per 100,000 person days | |  | 0.96 | 0.92 | 0.23 |  |  | 0.71 | 0.07 | 0.17 |  |  | 0.08 | 0.05 | 0.16 |  | 0.12 | 0.08 | 0.05 | 0.21 |
| Crude HR (95%CI) |  |  | 4.46 (1.92 - 10.37) | 4.21 (1.82 - 9.77) | Reference |  |  | 5.34 (3.24 - 8.81) | 0.58 (0.22 - 1.51) | Reference |  |  | 0.56 (0.28 - 1.12) | 0.34 (0.14 - 0.82) | Reference |  | 0.44 (0.16 - 1.19) | 0.36 (0.19 - 0.69) | 0.23 (0.10 - 0.54) | Reference |
| Age adjusted HR (95% CI) |  |  | 0.52 (0.22 - 1.27) | 1.16 (0.50 - 2.71) | Reference |  |  | 0.97 (0.57 - 1.65) | 0.34 (0.13 - 0.87) | Reference |  |  | 0.24 (0.12 - 0.48) | 0.26 (0.11 - 0.62) | Reference |  | 0.10 (0.04 - 0.27) | 0.23 (0.12 - 0.44) | 0.23 (0.10 - 0.53) | Reference |
| Age and gender adjusted HR (95% CI) | |  | ( - ) | 1.17 (0.50 - 2.73) | Reference |  |  | 0.98 (0.58 - 1.67) | 0.33 (0.13 - 0.86) | Reference |  |  | 0.23 (0.12 - 0.47) | 0.25 (0.10 - 0.60) | Reference |  | 0.10 (0.04 - 0.26) | 0.22 (0.12 - 0.43) | 0.22 (0.10 - 0.50) | Reference |
| Age, gender and nursing home adjusted HR (95% CI) | | | 0.24 (0.10 - 0.59) | 0.85 (0.36 - 2.02) | Reference |  |  | 0.87 (0.50 - 1.52) | 0.34 (0.13 - 0.86) | Reference |  |  | 0.24 (0.12 - 0.49) | 0.24 (0.10 - 0.59) | Reference |  | 0.09 (0.03 - 0.24) | 0.22 (0.11 - 0.41) | 0.23 (0.10 - 0.52) | Reference |
|  | 2022 | | | | | | | | | | | | | | | | | | | |
|  | first quarter | | | | | second quarter | | | | | third quarter | | | | | fourth quarter | | | | |
|  | Four or more vaccine doses | Three vaccine doses | Two vaccine doses | One vaccine dose | Unvaccinated | Four or more vaccine doses | Three vaccine doses | Two vaccine doses | One vaccine dose | Unvaccinated | Four or more vaccine doses | Three vaccine doses | Two vaccine doses | One vaccine dose | Unvaccinated | Four or more vaccine doses | Three vaccine doses | Two vaccine doses | One vaccine dose | Unvaccinated |
| Deaths (n) | 2 | 39 | 37 | 4 | 39 | 1 | 40 | 32 | 5 | 16 | 10 | 71 | 26 | 6 | 19 | 50 | 81 | 16 | 2 | 22 |
| Events per 100,000 person days | 15.68 | 0.24 | 0.16 | 0.04 | 0.21 | 0.48 | 0.07 | 0.08 | 0.05 | 0.04 | 0.18 | 0.06 | 0.05 | 0.06 | 0.03 | 0.16 | 0.06 | 0.03 | 0.02 | 0.03 |
| Crude HR (95%CI) | 73.99 (17.84 - 306.79) | 1.14 (0.73 - 1.78) | 0.78 (0.49 - 1.22) | 0.22 (0.08 - 0.61) | Reference | 18.68 (2.41 - 144.71) | 2.32 (1.29 - 4.16) | 2.14 (1.17 - 3.90) | 1.22 (0.45 - 3.36) | Reference | 6.28 (2.85 - 13.87) | 2.02 (1.22 - 3.35) | 1.58 (0.87 - 2.85) | 1.83 (0.73 - 4.57) | Reference | 5.27 (3.17 - 8.76) | 1.97 (1.23 - 3.15) | 0.87 (0.46 - 1.66) | 0.55 (0.13 - 2.33) | Reference |
| Age adjusted HR (95% CI) | 8.76 (2.09 - 36.71) | 0.32 (0.20 - 0.50) | 0.41 (0.26 - 0.65) | 0.22 (0.08 - 0.61) | Reference | 1.00 (0.13 - 8.01) | 0.70 (0.39 - 1.26) | 1.37 (0.75 - 2.50) | 1.19 (0.43 - 3.26) | Reference | 0.54 (0.24 - 1.23) | 0.86 (0.52 - 1.43) | 1.45 (0.80 - 2.62) | 1.91 (0.76 - 4.80) | Reference | 0.77 (0.46 - 1.29) | 1.17 (0.73 - 1.87) | 0.96 (0.51 - 1.84) | 0.63 (0.15 - 2.68) | Reference |
| Age and gender adjusted HR (95% CI) | ( - ) | 0.32 (0.20 - 0.50) | 0.40 (0.25 - 0.63) | 0.21 (0.08 - 0.60) | Reference | ( - ) | 0.67 (0.37 - 1.20) | 1.30 (0.71 - 2.38) | ( - ) | Reference | 0.49 (0.22 - 1.10) | 0.83 (0.50 - 1.38) | 1.44 (0.80 - 2.61) | 1.90 (0.76 - 4.75) | Reference | 0.77 (0.46 - 1.29) | 1.13 (0.70 - 1.81) | 0.95 (0.50 - 1.81) | 0.63 (0.15 - 2.66) | Reference |
| Age, gender and nursing home adjusted HR (95% CI) | 5.92 (1.38 - 25.30) | 0.24 (0.15 - 0.38) | 0.40 (0.26 - 0.64) | 0.23 (0.08 - 0.65) | Reference | 0.85 (0.10 - 7.05) | 0.54 (0.29 - 0.98) | 1.24 (0.68 - 2.28) | 1.19 (0.43 - 3.27) | Reference | 0.34 (0.14 - 0.79) | 0.73 (0.43 - 1.22) | 1.33 (0.73 - 2.41) | 1.82 (0.73 - 4.58) | Reference | 0.57 (0.33 - 0.98) | 1.01 (0.63 - 1.62) | 0.89 (0.47 - 1.70) | 0.60 (0.14 - 2.57) | Reference |
|  | 2023 | | | | | | | | | | | | | | | | | | | |
|  | first quarter | | | | | second quarter | | | | | third quarter | | | | | fourth quarter | | | | |
|  | Four or more vaccine doses | Three vaccine doses | Two vaccine doses | One vaccine dose | Unvaccinated | Four or more vaccine doses | Three vaccine doses | Two vaccine doses | One vaccine dose | Unvaccinated | Four or more vaccine doses | Three vaccine doses | Two vaccine doses | One vaccine dose | Unvaccinated | Four or more vaccine doses | Three vaccine doses | Two vaccine doses | One vaccine dose | Unvaccinated |
| Deaths (n) | 98 | 65 | 19 |  | 28 | 33 | 25 | 9 | 2 | 10 | 11 | 18 | 3* | 2* | 6 | 128 | 103 | 23 | 3 | 27 |
| Events per 100,000 person days | 0.2 | 0.05 | 0.03 |  | 0.04 | 0.06 | 0.02 | 0.02 | 0.02 | 0.01 | 0.02 | 0.01 | 0.01 | 0.02 | 0.01 | 0.22 | 0.07 | 0.04 | 0.03 | 0.04 |
| Crude HR (95%CI) | 4.93 (3.24 - 7.50) | 1.21 (0.78 - 1.88) | 0.82 (0.46 - 1.46) |  | Reference | 4.29 (2.12 - 8.71) | 1.26 (0.61 - 2.63) | 1.08 (0.44 - 2.66) | 1.25 (0.27 - 5.69) | Reference | 2.26 (0.84 - 6.11) | 1.49 (0.59 - 3.76) |  |  | Reference | 5.88 (3.88 - 8.90) | 1.90 (1.25 - 2.91) | 1.01 (0.58 - 1.77) | 0.69 (0.21 - 2.29) | Reference |
| Age adjusted HR (95% CI) | 0.86 (0.56 - 1.31) | 0.78 (0.50 - 1.21) | 0.98 (0.55 - 1.76) |  | Reference | 0.78 (0.38 - 1.60) | 0.82 (0.40 - 1.71) | 1.32 (0.54 - 3.25) | 1.55 (0.34 - 7.06) | Reference | 0.41 (0.15 - 1.12) | 0.99 (0.39 - 2.49) |  |  | Reference | 1.13 (0.75 - 1.73) | 1.24 (0.81 - 1.89) | 1.27 (0.73 - 2.22) | 0.88 (0.27 - 2.90) | Reference |
| Age and gender adjusted HR (95% CI) | 0.86 (0.56 - 1.31) | 0.75 (0.48 - 1.17) | 0.97 (0.54 - 1.74) |  | Reference | 0.76 (0.37 - 1.56) | 0.80 (0.39 - 1.68) | 1.32 (0.53 - 3.24) | 1.53 (0.34 - 6.99) | Reference | 0.37 (0.14 - 1.01) | 0.94 (0.37 - 2.38) |  |  | Reference | 1.08 (0.71 - 1.65) | 1.18 (0.77 - 1.81) | 1.26 (0.72 - 2.19) | 0.87 (0.26 - 2.87) | Reference |
| Age, gender and nursing home adjusted HR (95% CI) | 0.63 (0.41 - 0.97) | 0.71 (0.45 - 1.10) | 0.92 (0.51 - 1.65) |  | Reference | 0.63 (0.30 - 1.31) | 0.76 (0.37 - 1.59) | 1.31 (0.53 - 3.23) | 1.49 (0.33 - 6.82) | Reference | 0.33 (0.12 - 0.91) | 0.93 (0.37 - 2.35) |  |  | Reference | 0.97 (0.63 - 1.50) | 1.17 (0.76 - 1.79) | 1.24 (0.71 - 2.17) | 0.87 (0.26 - 2.86) | Reference |
| * HRs not reported because there were less than 10 events total (including reference group) | | | | | | | | | | | | | | | | | | | | |

| **Table S43:** Hazard ratios (HR) with 95% confidence intervals (95% CI) for COVID 19 mortality according to number of SARS-CoV-2 vaccine doses during different time periods. | | | | | | | | | | |
| --- | --- | --- | --- | --- | --- | --- | --- | --- | --- | --- |
|  | 2021 | | | | | | | | | |
|  | June and July 2021 (low COVID-19 disease burden) | | | | | October and November 2021 (high COVID-19 disease burden) | | | | |
|  | Four or more vaccine doses | Three vaccine doses | Two vaccine doses | One vaccine dose | Unvaccinated | Four or more vaccine doses | Three vaccine doses | Two vaccine doses | One vaccine dose | Unvaccinated |
| Deaths (n) |  |  | 10 | 2 | 9 |  | 2 | 10 | 4 | 13 |
| Events per 100,000 person days |  |  | 0.18 | 0.03 | 0.08 |  | 0.17 | 0.08 | 0.04 | 0.13 |
| Crude HR (95%CI) |  |  | 2.24 (0.89 - 5.64) | 0.33 (0.07 - 1.54) | Reference |  | 0.96 (0.21 - 4.39) | 0.59 (0.26 - 1.34) | 0.27 (0.09 - 0.84) | Reference |
| Age adjusted HR (95% CI) |  |  | 0.75 (0.30 - 1.89) | 0.29 (0.06 - 1.35) | Reference |  | 0.14 (0.03 - 0.65) | 0.37 (0.16 - 0.85) | 0.28 (0.09 - 0.85) | Reference |
| Age and gender adjusted HR (95% CI) |  |  | 0.75 (0.30 - 1.90) | 0.30 (0.07 - 1.41) | Reference |  | 0.05 (0.01 - 0.20) | 0.35 (0.15 - 0.79) | 0.24 (0.10 - 0.61) | Reference |
| Age, gender and nursing home adjusted HR (95% CI) |  |  | 0.77 (0.30 - 1.98) | 0.30 (0.06 - 1.37) | Reference |  | 0.04 (0.01 - 0.19) | 0.34 (0.15 - 0.78) | 0.26 (0.10 - 0.64) | Reference |
| Age, gender, nursing home and year of last infection adjusted HR (95% CI) |  |  | 0.79 (0.30 - 2.07) | 0.31 (0.07 - 1.45) | Reference |  | 0.04 (0.01 - 0.20) | 0.37 (0.16 - 0.85) | 0.25 (0.10 - 0.63) | Reference |
|  | May and June 2022 (low COVID-19 disease burden) | | | | | February and March 2022 (high COVID-19 disease burden) | | | | |
|  | Four or more vaccine doses | Three vaccine doses | Two vaccine doses | One vaccine dose | Unvaccinated | Four or more vaccine doses | Three vaccine doses | Two vaccine doses | One vaccine dose | Unvaccinated |
| Deaths (n) |  | 10 | 5 | 1* | 5 | 2 | 27 | 17 | 2 | 15 |
| Events per 100,000 person days |  | 0.04 | 0.02 | 0.02 | 0.02 | 20.26 | 0.23 | 0.13 | 0.04 | 0.19 |
| Crude HR (95%CI) |  | 1.95 (0.67 - 5.71) | 1.05 (0.30 - 3.64) |  | Reference | 99.91 (22.80 - 437.88) | 1.18 (0.63 - 2.23) | 0.69 (0.35 - 1.39) | 0.21 (0.05 - 0.90) | Reference |
| Age adjusted HR (95% CI) |  | 0.57 (0.19 - 1.66) | 0.63 (0.18 - 2.16) |  | Reference | 15.37 (3.46 - 68.27) | 0.42 (0.22 - 0.80) | 0.43 (0.21 - 0.86) | 0.24 (0.05 - 1.03) | Reference |
| Age and gender adjusted HR (95% CI) |  | 0.52 (0.18 - 1.53) | 0.60 (0.17 - 2.08) |  | Reference | 15.00 (3.39 - 66.44) | 0.42 (0.22 - 0.79) | 0.42 (0.21 - 0.84) | 0.23 (0.05 - 1.01) | Reference |
| Age, gender and nursing home adjusted HR (95% CI) |  | 0.47 (0.16 - 1.41) | 0.60 (0.17 - 2.07) |  | Reference | 12.56 (2.72 - 58.04) | 0.33 (0.17 - 0.64) | 0.42 (0.21 - 0.85) | 0.24 (0.05 - 1.05) | Reference |
| Age, gender, nursing home and year of last infection adjusted HR (95% CI) |  | 0.43 (0.14 - 1.32) | 0.54 (0.15 - 1.93) |  | Reference | 12.83 (2.69 - 61.14) | 0.34 (0.17 - 0.65) | 0.42 (0.21 - 0.85) | 0.24 (0.06 - 1.06) | Reference |
|  | July and August 2023 (low COVID-19 disease burden) | | | | | February and March 2023 (high COVID-19 disease burden) | | | | |
|  | Four or more vaccine doses | Three vaccine doses | Two vaccine doses | One vaccine dose | Unvaccinated | Four or more vaccine doses | Three vaccine doses | Two vaccine doses | One vaccine dose | Unvaccinated |
| Deaths (n) | 3* | 7 |  | 1* | 4 | 56 | 31 | 8 |  | 16 |
| Events per 100,000 person days | 0.01 | 0.01 |  | 0.01 | 0.01 | 0.16 | 0.03 | 0.02 |  | 0.03 |
| Crude HR (95%CI) |  | 0.88 (0.26 - 2.99) |  |  | Reference | 4.91 (2.82 - 8.57) | 1.03 (0.56 - 1.88) | 0.61 (0.26 - 1.42) |  | Reference |
| Age adjusted HR (95% CI) |  | 0.58 (0.17 - 1.98) |  |  | Reference | 0.86 (0.49 - 1.50) | 0.66 (0.36 - 1.20) | 0.73 (0.31 - 1.71) |  | Reference |
| Age and gender adjusted HR (95% CI) |  | 0.54 (0.16 - 1.84) |  |  | Reference | 0.84 (0.48 - 1.47) | 0.62 (0.34 - 1.13) | 0.72 (0.31 - 1.68) |  | Reference |
| Age, gender and nursing home adjusted HR (95% CI) |  | 0.52 (0.15 - 1.79) |  |  | Reference | 0.60 (0.34 - 1.07) | 0.58 (0.32 - 1.06) | 0.68 (0.29 - 1.59) |  | Reference |
| Age, gender, nursing home and year of last infection adjusted HR (95% CI) |  | 0.51 (0.15 - 1.77) |  |  | Reference | 0.61 (0.34 - 1.08) | 0.58 (0.32 - 1.06) | 0.65 (0.28 - 1.54) |  | Reference |
| * HRs not reported because there were less than 10 events total (including reference group) | | | | | | | | | | |

### **Matched two weeks post vaccination analyses**

| **Table S44:** Hazard ratios (HR) with 95% confidence intervals (95% CI) for non-COVID-19 and all-cause mortality according to number of SARS-CoV-2 vaccine doses in the two weeks after vaccination with controls matched based on age group, gender and nursing home residency. | | | | |
| --- | --- | --- | --- | --- |
|  | All-cause mortality | | | |
|  | Four or more vaccine doses | Unvaccinated | Three vaccine doses | Unvaccinated |
| Deaths (n) | 223 | 505 | 126 | 419 |
| Events per 100,000 person days | 2.87 | 6.5 | 2.32 | 7.71 |
| Crude HR (95%CI) | 0.44 (0.38 - 0.52) | Reference | 0.30 (0.25 - 0.37) | Reference |
| Age adjusted HR (95% CI) | 0.44 (0.38 - 0.52) | Reference | 0.30 (0.25 - 0.37) | Reference |
|  | Two vaccine doses | Unvaccinated | One vaccine dose | Unvaccinated |
| Deaths (n) | 136 | 568 | 150 | 456 |
| Events per 100,000 person days | 2.33 | 9.75 | 2.32 | 7.06 |
| Crude HR (95%CI) | 0.24 (0.20 - 0.29) | Reference | 0.33 (0.27 - 0.40) | Reference |
| Age adjusted HR (95% CI) | 0.24 (0.20 - 0.29) | Reference | 0.33 (0.27 - 0.40) | Reference |
|  | Non-COVID-19 | | | |
|  | Four or more vaccine doses | Unvaccinated | Three vaccine doses | Unvaccinated |
| Deaths (n) | 219 | 503 | 122 | 405 |
| Events per 100,000 person days | 2.82 | 6.48 | 2.25 | 7.46 |
| Crude HR (95%CI) | 0.44 (0.37 - 0.51) | Reference | 0.30 (0.25 - 0.37) | Reference |
| Age adjusted HR (95% CI) | 0.44 (0.37 - 0.51) | Reference | 0.30 (0.25 - 0.37) | Reference |
|  | Two vaccine doses | Unvaccinated | One vaccine dose | Unvaccinated |
| Deaths (n) | 117 | 523 | 129 | 410 |
| Events per 100,000 person days | 2.01 | 8.98 | 2 | 6.35 |
| Crude HR (95%CI) | 0.22 (0.18 - 0.27) | Reference | 0.31 (0.26 - 0.38) | Reference |
| Age adjusted HR (95% CI) | 0.22 (0.18 - 0.27) | Reference | 0.32 (0.26 - 0.38) | Reference |

| **Table S45:** Hazard ratios (HR) with 95% confidence intervals (95% CI) for cancer and COVID-19 mortality according to number of SARS-CoV-2 vaccine doses in the two weeks after vaccination with controls matched based on age group, gender and nursing home residency. | | | | |
| --- | --- | --- | --- | --- |
|  | Cancer | | | |
|  | Four or more vaccine doses | Unvaccinated | Three vaccine doses | Unvaccinated |
| Deaths (n) | 30 | 96 | 21 | 90 |
| Events per 100,000 person days | 0.39 | 1.24 | 0.39 | 1.66 |
| Crude HR (95%CI) | 0.31 (0.21 - 0.47) | Reference | 0.23 (0.15 - 0.38) | Reference |
| Age adjusted HR (95% CI) | 0.31 (0.21 - 0.47) | Reference | 0.23 (0.14 - 0.37) | Reference |
|  | Two vaccine doses | Unvaccinated | One vaccine dose | Unvaccinated |
| Deaths (n) | 17 | 102 | 18 | 88 |
| Events per 100,000 person days | 0.29 | 1.75 | 0.28 | 1.36 |
| Crude HR (95%CI) | 0.17 (0.10 - 0.28) | Reference | 0.20 (0.12 - 0.34) | Reference |
| Age adjusted HR (95% CI) | 0.17 (0.10 - 0.28) | Reference | 0.20 (0.12 - 0.34) | Reference |
|  | COVID-19 | | | |
|  | Four or more vaccine doses | Unvaccinated | Three vaccine doses | Unvaccinated |
| Deaths (n) | 4 | 2 | 4 | 14 |
| Events per 100,000 person days | 0.05 | 0.03 | 0.07 | 0.26 |
| Crude HR (95%CI) | * | Reference | 0.29 (0.09 - 0.87) | Reference |
| Age adjusted HR (95% CI) | * | Reference | 0.29 (0.09 - 0.87) | Reference |
|  | Two vaccine doses | Unvaccinated | One vaccine dose | Unvaccinated |
| Deaths (n) | 19 | 45 | 21 | 46 |
| Events per 100,000 person days | 0.33 | 0.77 | 0.33 | 0.71 |
| Crude HR (95%CI) | 0.42 (0.25 - 0.72) | Reference | 0.46 (0.27 - 0.76) | Reference |
| Age adjusted HR (95% CI) | 0.42 (0.25 - 0.73) | Reference | 0.46 (0.27 - 0.77) | Reference |
| Transport accidents are not displayed due to the lack of events. | | |  |  |

### **Vaccinations**

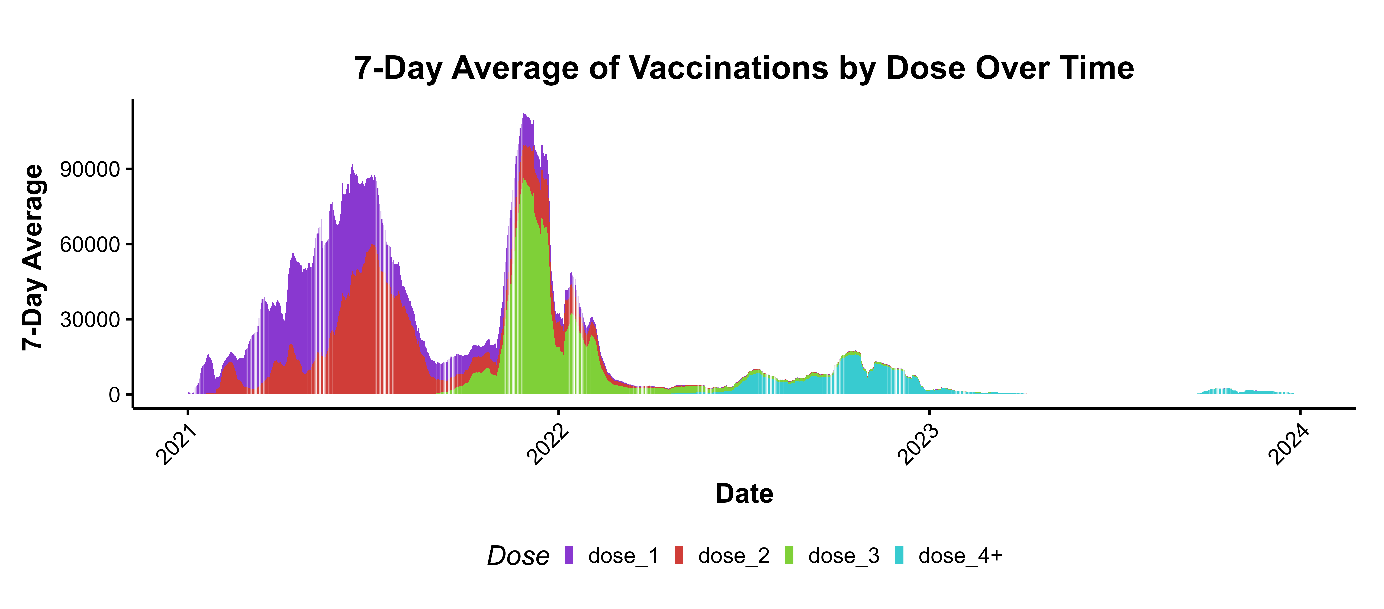
**Figure S1:** Seven-day average of daily vaccinations, by dose.
